# Decreased expression of mitochondrial aminoacyl-tRNA synthetases causes downregulation of mitochondrial OXPHOS subunits in type 2 diabetic skeletal muscle

**DOI:** 10.1101/2022.12.30.22283970

**Authors:** Iliana López-Soldado, Adrian Gabriel Torres, Raúl Ventura, Inma Martínez-Ruiz, Angels Díaz-Ramos, Evarist Planet, Diane Cooper, Agnieszka Pazderska, Krzysztof Wanic, Declan O’Hanlon, Donal J. O’Gorman, Teresa Carbonell, Lluís Ribas de Pouplana, John J. Nolan, María Isabel Hernández-Alvarez

**Author notes:** Corresponding author. María Isabel Hernández-Alvarez.

## Abstract

Type 2 diabetes mellitus (T2D) affects millions of people worldwide and is one of the leading causes of morbidity and mortality. The skeletal muscle (SKM) is the most important tissue involved in maintaining glucose homeostasis and substrate oxidation, and it undergoes insulin resistance in T2D. In this study, we identify the existence of alterations in the expression of mitochondrial aminoacyl-tRNA synthetases (mt-aaRSs) in skeletal muscle from two different forms of T2D: early-onset type 2 diabetes (YT2) (onset of the disease before 30 years of age) and the classical form of the disease (OT2). GSEA analysis from microarray studies revealed the repression of mitochondrial mt-aaRSs independently of age, which was validated by real-time PCR assays. In agreement with this, a reduced expression of several encoding mt-aaRSs was also detected in skeletal muscle from diabetic (db/db) mice but not in obese ob/ob mice. In addition, the expression of the mt-aaRSs proteins most relevant in the synthesis of mitochondrial proteins, threonyl-tRNA, and leucyl-tRNA synthetases (LARS2 and TARS2) were also repressed in muscle from db/db mice. It is likely that these alterations participate in the reduced expression of proteins synthesized in the mitochondria detected in db/db mice. Because it is known that, nitrosative stress inhibits aminoacylation of TARS2 and LARS2 activities, we noticed an increased protein expression of iNOS in isolated muscle mitochondria in diabetic mice.

Our results indicate a reduced expression of mitochondrial mt-aaRSs in skeletal muscle from T2D patients, which may participate in the reduced expression of proteins synthesized in mitochondria. This may be due to an enhanced NO production secondary to enhanced iNOS expression in muscle under diabetic conditions.

**Highlights:**

- Mt-aaRSs are downregulated in the skeletal muscle of type 2 diabetic patients and diabetic mice.
- The downregulation of mt-aaRSs in the skeletal muscle of diabetic mice is affecting the synthesis of ND2 which is a *mitochondrially* encoded *subunit* of complex I.
- Mitochondrial iNOS could be a target for reduced expression of mt-aaRSs in the skeletal muscle of diabetic mice.

## Introduction

The prevalence of type 2 diabetes (T2D) has reached epidemic proportions worldwide. Furthermore, T2D is related to many chronic comorbidities that can undermine the quality of life and life span (1). T2D is characterized by insulin resistance (IR) in skeletal muscle (2), which can result from combining a genetic predisposition with obesity, a sedentary lifestyle, and an unhealthy diet (3).

Mitochondria are the major functional components of cellular fuel oxidation and ATP production (4). The preliminary evidence linking mitochondrial dysfunction to insulin resistance in skeletal muscle comes from studies performed in non-insulin-dependent diabetic individuals who exhibited an increased glycolytic to oxidative ratio, which suggests that a dysregulation between mitochondrial oxidative capacity and capacity for glycolysis is an important component of the expression of insulin resistance (5). Later on, it was described that the expression of many genes of oxidative metabolism is reduced in T2D (6, 7). Among them, PPARγ coactivator 1-α and -β (PGC1-α and PGC1-β) is decreased in both diabetic subjects and family history-positive nondiabetic subjects (6). Also, the total activity of mitochondrial electron transport in the skeletal muscle is reduced in T2DM subjects (8). Moreover, the skeletal muscle mitochondrial proteins are downregulated in the transition from prediabetes into type 2 diabetes (9). Additionally, insulin resistance in the skeletal muscle of insulin-resistant offspring of patients with type 2 diabetes is associated with dysregulation of intramyocellular fatty acid metabolism, possibly because of an inherited defect in mitochondrial oxidative phosphorylation (10). Despite the well-documented association between mitochondrial dysfunction and insulin resistance, whether impaired mitochondrial oxidative capacity is causal to or is a consequence of insulin resistance remains a matter of debate (11).

Mitochondrial protein synthesis provides key components of the oxidative phosphorylation complexes. For mitochondrial protein synthesis, a collaboration of two genomes is required: mitochondrial tRNAs and ribosomal RNAs are encoded by the mitochondrial genome, whereas all protein factors involved in the translation of mitochondrial proteins are encoded by nuclear genes, translated in the cytoplasm and imported into mitochondria.

The mitochondrial aminoacyl-tRNA synthetase proteins (mt-aaRSs) are a group of nuclear-encoded enzymes that facilitate the conjugation of each of the 20 amino acids to its cognate tRNA molecule, through a two-step reaction, where they first activate the amino acid with ATP, forming an intermediate aminoacyl-adenylate, and then transfer the aminoacyl group to the 3□-end of its own tRNA (12). These mt-aaRSs are sometimes encoded by the same genes as the cytosolic ones (dualy-localized mt-aaRS), yet in many cases, are encoded by a different gene (exclusively localized) (13). In humans, only two aaRs are dualy-localized (Glycil- and lysil-tRNA synthetase). Each of the 19 human genes encoding mt-aaRS have been associated with a wide spectrum of human disorders (14), highlighting their importance for human health (15, 16). The main organs affected are the central nervous system, and the musculoskeletal, cardiovascular, and urinary systems (17).

Recently, it has been described that nitrosative stress inhibits aminoacylation of mitochondrial threonyl-tRNA and leucyl-tRNA synthetases (TARS2 and LARS2) by S-nitrosation (18). These data clearly showed that mt-aaRSs constitute a family of enzymes sensitive to posttranslational S-nitrosation modification mediated by reactive nitrogen species (RNS) (18). Protein nitrosation is a recently highlighted protein modification. Nitrosation is a general term that indicates nitrogen-associated gaseous modification and includes the ligation of nitric oxide (NO) or nitrogen dioxide on proteins, peptides, and metal ions (19). NO is synthesized by NO synthase (NOS) during the catalysis of L-arginine into L-citrulline. Three NOS isoforms have been reported: neuronal NOS (nNOS and NOS1), inducible NOS (iNOS and NOS2), and endothelial NOS (eNOS and NOS3) (20). Although NO is usually beneficial in the basal context, cumulative stress from chronic inflammation or oxidative insult produces a large amount of NO, which induces atypical protein nitrosation (20).

In this study, we wanted to determine if patients or animal models with type 2 diabetes or obesity presented alterations in the muscle that could be associated with mitochondrial dysfunction and insulin resistance and explore whether mitochondrial nitrosative stress could be involved in these alterations.

## Methods

### Study subjects

As previously reported (21), early-onset type 2 diabetes was described as the onset of disease before the age of thirty, while classical onset type 2 diabetes refers to the onset of disease in patients over fifty. Participants of the study who presented with comorbidities or a secondary form of diabetes were not included. All subjects had normal or high fasting c-peptide concentrations at diagnosis (≥2.5ng/ml), were clinically obese (BMI > 30), and were weight-stable and negative for glutamic acid decarboxylase (GAD) antibodies. The exclusion criteria included active cancer, a history of ketoacidosis or ketosis, the use of corticosteroids, and pregnancy. Controls who were healthy, obese, had no family history of diabetes and had normal glucose tolerance were recruited from endocrine clinics at St James’ Hospital and through advertisements published by Dublin City University. All patients gave written informed consent, approved by the local Research Clinics Committee. They were sedentary at baseline (as determined by the International Physical Activity Questionnaire-Long Format), alongside a low VO2 max. 46% of patients with early-onset, and 39% with classical-onset type 2 diabetes, had a family history of this disease via a first-degree relative.

Prior to study enrolment, the duration of diabetes was 2.1 years for early-onset disease and 4.8 years for classical-onset disease. Study participants presented with stable blood glucose concentrations, with 88% of the early-onset, and 78% of the classical-onset type 2 diabetes group, being treated with oral hypoglycaemic medications. Insulin was administered to 23% of early-onset and 17% of classical-onset type 2 diabetes patients. All subjects underwent screening, during which a physical examination was carried out, and a medical history was taken. Blood, urine, and its components were analysed. Anthropometric data were obtained by measuring the participant&s BMI, weight (upon fasting and taken using a calibrated medical scale), waist-to-hip ratio (measured around the abdomen at the narrowest point between the iliac crests and the lowest costal margin above the umbilicus, as well as around the greater trochanters of the femur and gluteal mass). The patient&s height was also taken using a stadiometer. This study forms part of a larger study in which the participants were subjected to various lifestyle changes (22, 23). All methods explained above adhered to the relevant regulations and guidance.

Patients who accessed the diabetes or endocrinology services at St James’ Hospital in Dublin and who had either obesity or diabetes were invited to participate in the study. The local Research Ethics Committee approved the study protocol and consent was provided by all subjects. The project was funded with a grant to JJN at Trinity College Dublin from EFSD/Novo Nordisk Clinical Research Program in Adolescents with Type 2 Diabetes (Cellular Mechanisms of Insulin Resistance in Early Onset Type 2 Diabtetes, grant reference T04001) with the project being approved by the ‘Joint St James’ Hospital and Adelaide and Meath Hospital Ethics Committee, with a reference number of 2008/06/03.

### Muscle Biopsies

As previously described (21), all patients were in a fasting state before the muscle biopsy was taken. The biopsies were obtained from the vastus lateralis muscle (100mg) under local anaesthesia. Specifically, an area of skin was anaesthetised with 1% lidocaine, and a small incision of 0.5 cm was made. To obtain the tissue, a Bergstrom biopsy needle was inserted into the muscle, and approximately 100mg of tissue was extracted using suction. The samples were snap-frozen in liquid nitrogen and stored at a temperature of −80ºC before their RNA was extracted.

### Microarray assays

As previously reported (21), the IRB Functional Genomics Core Faculty gave access to their microarray services, which included quality control testing of total RNA using Nanodrop spectrophotometry, and the Agilent Bioanalyser. cDNA libraries were constructed and amplified from 25 ng of total RNA with 17 cycles of amplification and using WTA2 (Sigma-Aldrich). 8 μg of the cDNA was cut into fragments by the enzyme DNAseI and biotinylated by a terminal transferase, which was obtained from the GeneChip Mapping 250K Nsp Assay Kit (Affymetrix). Following the Affymetrix protocol, a hybridization mixture was prepared with each sample being hybridized to a PrimeView Human array (Affymetrix). As detailed in the Fluidics protocol FS450_002, the arrays were washed and stained in Fluidics station 450 the samples were then scanned using the GeneChip scanner 3000 (both Affymetrix), following the recommendations of the manufacturer. The GCOS software permitted the generation of CEL files using DAT files (Affymetrix). The RMA algorithm 29 was used to determine the normalized expression intensities from Affymetrix CEL files, and microarray pre-processing was carried out using the R package (R core team (2014). Results in supplementary table 1 as previously reported (21).

### Mouse models

All procedures were approved by the Barcelona Science Park’s Animal Experimentation Committee and carried out in accordance with the European Community Council Directive and the National Institute of Health guidelines for the care and use of laboratory animals. Male db/db mice and db/+ littermates (n□=□8 per group) were purchased from Charles River Laboratories (Sulzfeld, Germany). Male ob/ob mice and C57BL6/J wild-type littermates (n□=□8 per group) were purchased from Charles River Laboratories (Sulzfeld, Germany), and were obtained at the age of 5 weeks. Mice were fed a standard chow diet and water ad libitum and were kept in 12-hour dark-light periods. Animals were killed in a non-fasted state at the age of 8 weeks. Mice were anesthetized using isofluorane and sacrificed by cervical dislocation.

### RNA extraction and Real-time PCR assays

RNA from muscle in humans and mice was extracted by using a protocol combining Trizol reagent (Invitrogen) and RNeasy Mini Kit columns (Qiagen) following the manufacturer’s instructions. RNA was reverse-transcribed with the SuperScriptIII reverse transcriptase kit (Invitrogen). Quantitative polymerase chain reaction (PCR) was performed using SYBER green (Applied Biosystems) or Taqman Universal PCR Mastermix (Applied Biosystems) on the QuantStudio 6 Flex. As endogenous control to correct for potential variation in RNA loading and quantification, Human PPIA (cyclophilin A) (#431883E) and mouse PPIA (Mm02342429_g1) from Applied Biosystems were used. mRNA expression was calculated using the ΔCT method. Briefly, the ΔCT was calculated by subtracting the CT for cyclophilin A, from the CT for the gene of interest. The relative expression of the gene of interest is calculated using the expression 2−ΔCT and reported as arbitrary units. The TaqMan probes used for humans were CARS2 (Hs00226049_m1), MARS2 (Hs00536599_s1), SARS2 (Hs00215168_m1), TARS2 (Hs00372819_m1), NARS2 (Hs00372778_m1), DARS2 (Hs00216620_m1), WARS2 (Hs00210571_m1), HARS2 (Hs00247230_m1), PARS2 (Hs00384448_m1), IARS2 (Hs01058371_m1), LARS2 (Hs01118920_m1) and VARS2 (Hs00383681_m1). For mouse: CARS2 (Mm01209058_m1), LARS2 (Mm00467701_m1), PARS2 (Mm01954197_s1), NARS2 (Mm00841042_m1), VARS2 (Mm00841195_m1), IARS2 (Mm01318288_m1), DARS2 (Mm00554081_m1), MARS2 (Mm00615287_s1), SARS2 (Mm00458332_m1), WARS2 (Mm04208966_m1), TARS2 (Mm00510878_m1) and HARS2 (Mm00475675_m1).

The primer sequence for the iNOS mouse forward was GTTCTCAGCCCAACAATACAAGA and for iNOS mouse reverse was GTGGACGGGTCGATGTCAC. For eNOS mouse forward CTCCCAGCTGTGTCCAACAT. For eNOS mouse reverse CACACAGCCACATCCTCAAG. For NDUFB8 mouse forward TGTTGCCGGGGTCATATCCTA. For NDUFB8 mouse reverse AGCATCGGGTAGTCGCCATA. For MT-ND2 mouse forward TTGCGTGAGATTCGCGTTCA. For MT-ND2 mouse reverse ATTCGCGGATCAGAATGGGC. For SDHB mouse forward AATTTGCCATTTACCGATGGGA. For SDHB mouse reverse AGCATCCAACACCATAGGTCC. For UQCRC2 mouse forward AAAGTTGCCCCGAAGGTTAAA. For UQCRC2 mouse reverse GAGCATAGTTTTCCAGAGAAGCA. For MT-CO1 mouse forward ATTGGCAAGAGAGCCATTTCTAC. For MT-CO1 mouse reverse CACGCCGATCAGCGTAAGT. For ATP5A mouse forward TCTCCATGCCTCTAACACTCG. For ATP5A mouse reverse TCTCCATGCCTCTAACACTCG. For MT-ATP6 mouse forward AGGATTCCCAATCGTTGTAGCC. For MT-ATP6 mouse reverse CCTTTTGGTGTGTGGATTAGCA.

### Mitochondrial isolation

Mitochondria were isolated from mouse skeletal muscle by homogenization using a polytron followed by a Douncer homogenizer with a Teflon pestle in homogenization buffer I (0.1M KCl, 5mM MgCl2, 5 mM EGTA, 5 mM sodium pyrophosphate, pH 6.8, and protease inhibitor tablet, Roche). Homogenates were centrifuged at 1,300x g for 10 min at 4°C. The supernatant (SN1) was kept, and the pellet was resuspended in buffer II (0.25 M sucrose, 50 mM KCL, 5 mM EDTA, 1 mM sodium pyrophosphate, pH 7.4, and protease inhibitor tablet, Roche) and centrifuged at 1,300xg for 10 min at 4°C. The supernatant (SN2), together with SN1 were centrifuged at 9,000xg for 15 min at 4°C. The pellet (isolated mitochondria) was resuspended in buffer II.

### Western blot

Homogenates for Western blot analyses were obtained from muscle. Tissues samples were homogenized in 10 vol of lysis buffer [10 mM Tris (pH 7.2), 150 mM NaCl, 1% Triton X-100, 0.1% SDS, 5 mM EDTA, 2 mM sodium orthovanadate, 50 mM NaF, 20 mM sodium pyrophosphate, and protease inhibitors mixture tablet (Roche)] with a mini-beadbeater (Biospec) twice for 30 sec. Homogenates were rotated for 1 h at 4 °C in an orbital shaker and centrifuged at 16,000 × g for 15 min at 4°C. Supernatants were aliquoted and kept at -20°C.

Proteins from total homogenates and the mitochondria isolation fraction were resolved in 10%, 12.5% or 15% acrylamide gels for SDS/PAGE and transferred to Immobilon membranes (Millipore). The following antibodies were used: Oxphos (abcam ab110413), ND2 (Proteintech 16879184), ATP6 (Millipore MABS1995), VDAC/Porin (Santa cruz 73614), Vinculin (Santa cruz 73614), LARS2 (Proteintech 17097-1-AP), TARS2 (Proteintech 15067-1-AP), NOS2 (Santa cruz sc-7271), NOS3 (BD Transduction 610296).

### Statistical analysis

Microarray data was analyzed as previously described (21), adjusting the data for batch and gender. Group comparisons in data other than microarray samples were performed using the Graphpad software. Human data were analysed as unpaired nonparametric distribution and the Mann-Whitney test was used. We assume a normal distribution of the data for animal studies using an unpaired parametric t-test. Data were presented as mean± standard error unless otherwise stated. Significance was established at p< 0.05.

## Results

### Identification of the downregulated aminoacyl-tRNA synthetases gene expression in early-onset and classical forms of type 2 diabetes

Microarray analysis was accomplished to identify the global expression profile of muscles biopsies from control [young obese control (YC), and old obese control (OC)] and type 2 diabetic patients [early-onset type 2 diabetes (YT2), and classical T2D (OT2)] as previously reported (21). The Gene Set Enrichment Analysis (GSEA) revealed the existence of genes dysregulated in both classical and early-onset type 2 diabetic patients that were enriched in metabolic pathways such as the metabolism of aminoacyl tRNA biosynthesis (Fig. 1A). As observed in figure 1B, the mitochondrial aminoacyl-tRNA synthetases (identified with number 2) were downregulated in type 2 diabetes; however, the cytosolic forms of the aminoacyl-tRNA synthetases were similar in type 2 diabetes and control subjects, showing that the defect in aminoacyl-tRNA synthetases is presumable mitochondrial.

**Figure 1.**
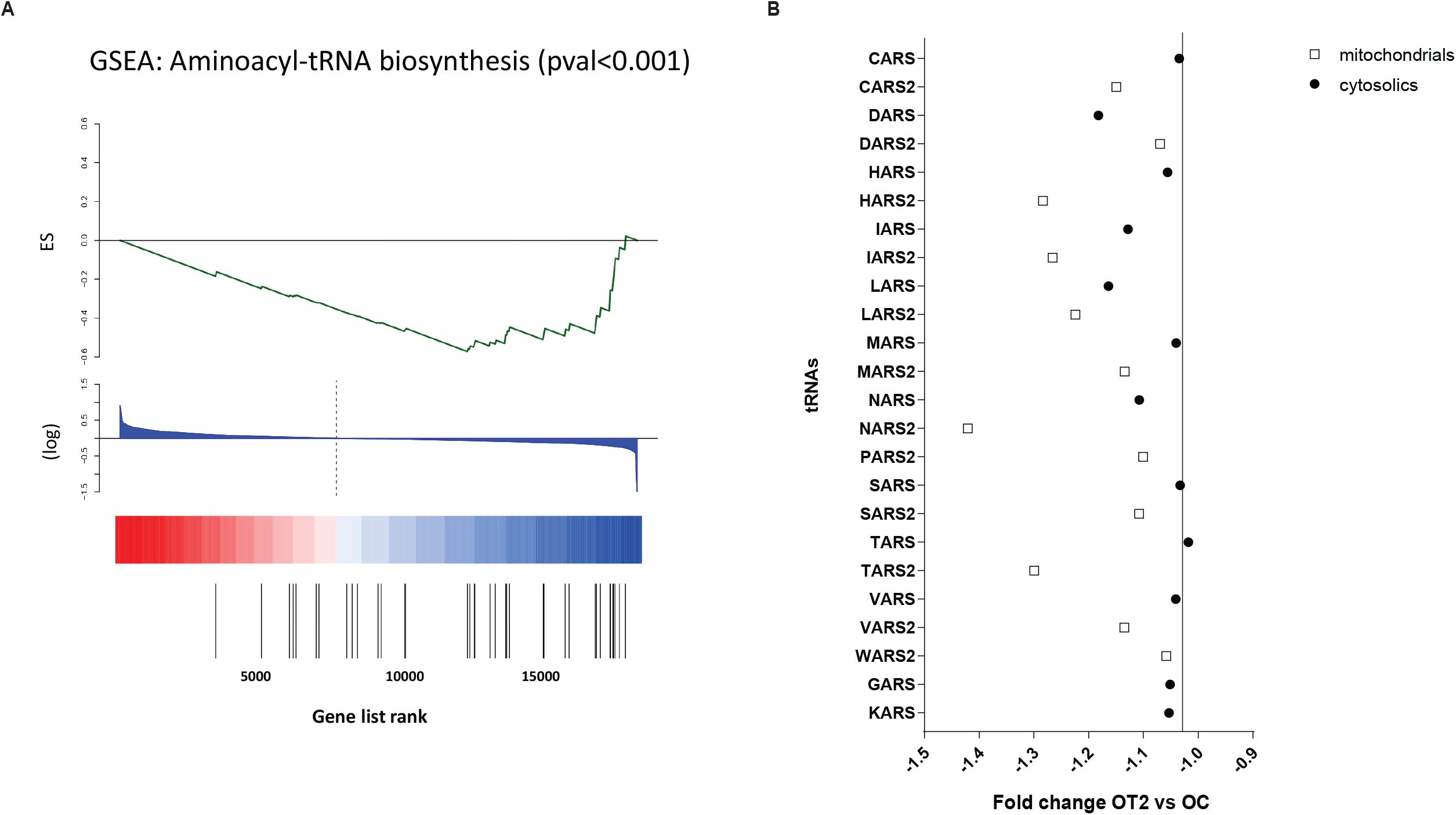
Identification of the downregulated aminoacyl-tRNA synthetases gene expression in early-onset and classical forms of type 2 diabetes. **(A)** The enrichment gene set expression of the aminocyl-tRNA synthetases. (**B)** Fold change OT2 vs OC of the main mitochondrial (identified with number 2) and cytosolic aminoacyl-tRNA synthetases.

### Type 2 diabetes is characterized by reduced muscle expression of genes encoding mt-aaRSs

To validate the result obtained by microarrays, we next performed real-time PCR assays to analyze the expression of the mt-aaRSs in muscle from the control and type 2 diabetic groups. YT2 was characterized only by reduced expression of the mt-aaRSs CARS2, HARS2 and, LARS2 (Fig. 2A, C and, E). However, in type 2 diabetes of older subjects, reduced expression of CARS2, DARS2, HARS2, IARS2, LARS2, NARS2, PARS2, TARS2, and VARS2 was detected (Fig 2. A-E, G-H and J-K). No change in any of the conditions was observed in the case of MARS2, SARS2, and WARS2 (Fig 2. F, I and L).

**Figure 2.**
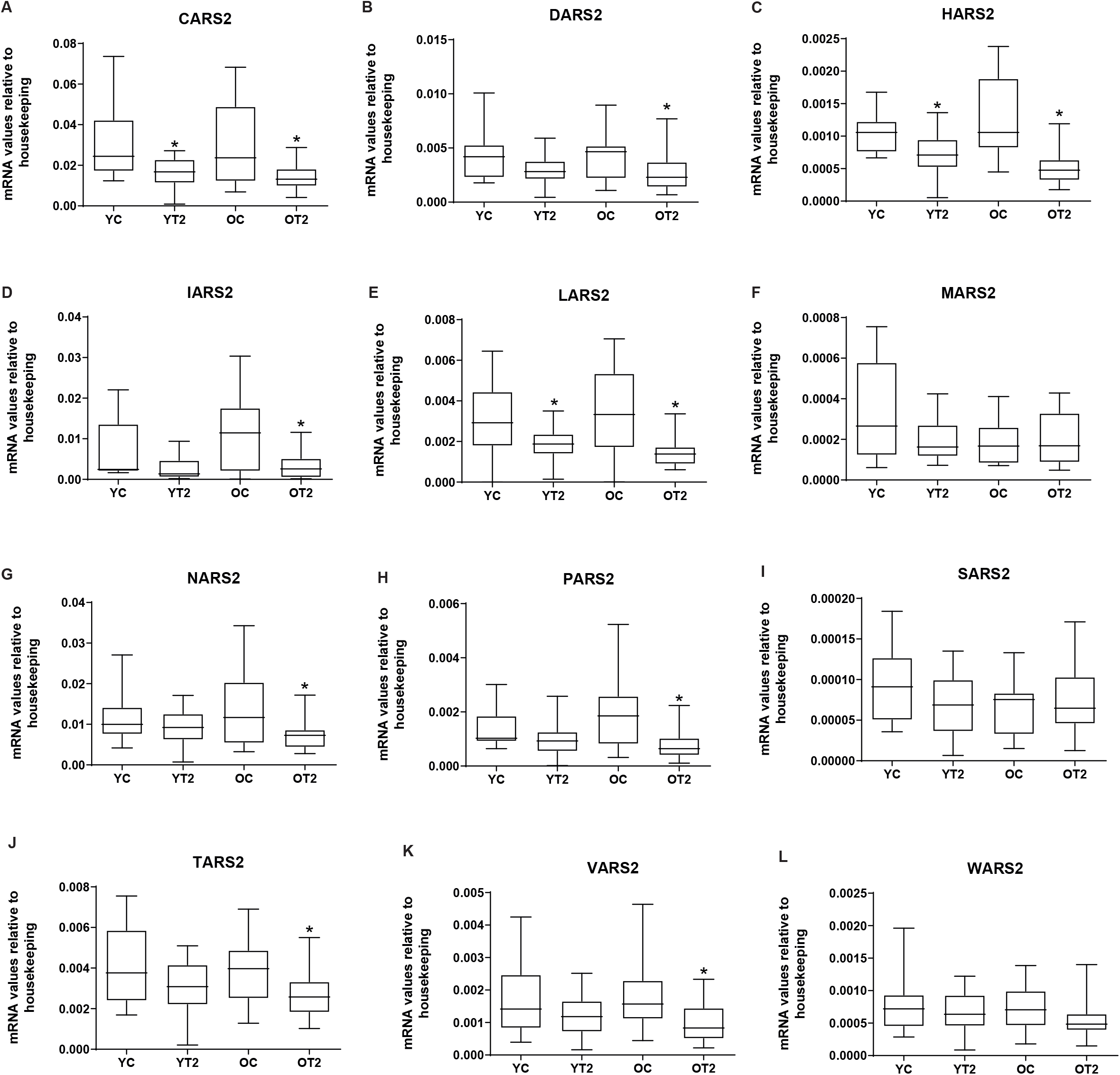
Skeletal muscle gene expression of mitochondrial tRNA synthetases in early-onset type 2 diabetic subjects (YT2) and late-onset type 2 diabetic subjects (OT2) and their respective matched control groups (YC and OC). Real-time PCR was performed in skeletal muscle biopsies. YT2 are early-onset type 2 diabetic subjects; OT2 are late-onset type 2 diabetic subjects, and their respective controls are YC and OC. **(A)** CARS2, **(B)** DARS2, **(C)** HARS2, **(D)** IARS2, **(E)** LARS2, **(F)** MARS2, **(G)** NARS2, **(H)** PARS2, **(I)** SARS2. Data are presented in boxplots; on each box, the central mark indicates the median, and the bottom and top edges of the box indicate the 25^th^ and 74^th^ percentiles, respectively. Unpaired non-parametric distribution and Mann-Whitney test was used *p<0.05. YC (n=12), YT2 (n=21), OC (n=17), OT2 (n=24).

To be sure that the changes in type 2 diabetes excluded changes in mt-aaRSs associated with methionine, serine and tryptophan, we decided to unify the data by obese phenotype and T2D (independently of age). In this context, general type 2 diabetes (YT2+ OT2) was characterized by reduced expression of genes encoding for the mt-aaRSs: CARS2, DARS2, HARS2, IARS2, LARS2, NARS2, PARS2, TARS2, and VARS2 (Fig 3. A-E, G-H and J-K). However, no changes were observed in the mRNA expression of MARS2, SARS2, or WARS2 in the diabetic groups (Fig 3. F, I, and L) as observed in separate groups of diabetic patients.

**Figure 3.**
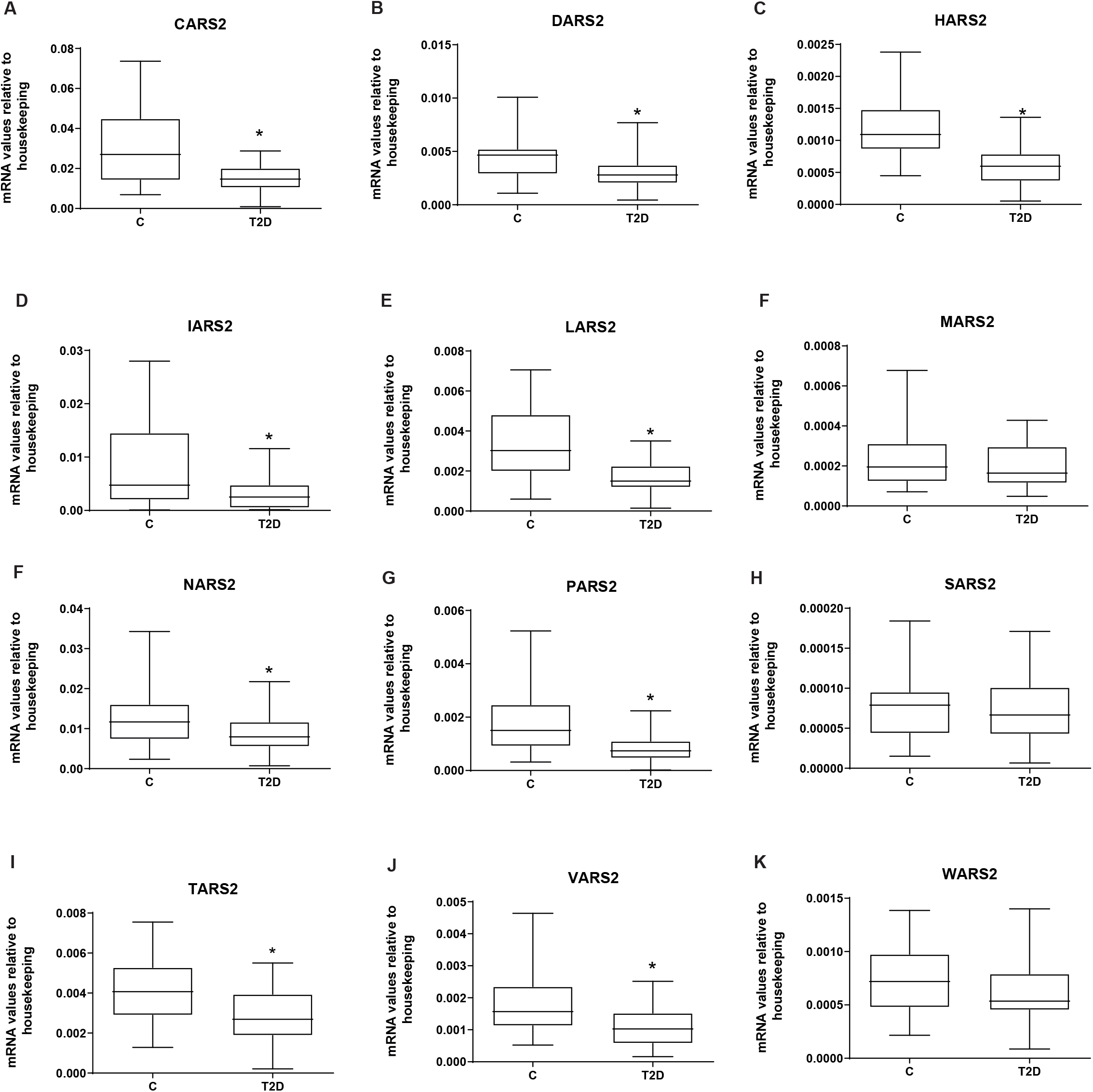
Skeletal muscle gene expression of mitochondrial tRNA synthetases in type 2 diabetic subjects (T2D) and their respective matched control groups. Real-time PCR was performed in skeletal muscle biopsies. T2D are type 2 diabetic subjects and their respective matched control groups. **(A)** CARS2, **(B)** DARS2, **(C)** HARS2, **(D)** IARS2, **(E)** LARS2, **(F)** MARS2, **(G)** NARS2, **(H)** PARS2, **(I)** SARS2. Data are presented in boxplots; on each box, the central mark indicates the median, and the bottom and top edges of the box indicate the 25^th^ and 74^th^ percentiles, respectively. Statistical analyses comparing type 2 diabetic subjects vs respective controls were performed by unpaired *t*-test *p < 0.05. C (n=29), T2D (n=45).

### mt-aaRSs are dysregulated in diabetic obese mice

To further explore the link between mt-aaRSs expression and T2D, we employed a series of mouse models. Given that all human subjects (both diabetic and non-diabetic) in this study are obese, we wanted to i) verify that diabetic obese mice recapitulate the mt-aaRSs expression patterns observed in obese T2D patients, and ii) evaluate the contribution of obesity towards mt-aaRSs expression in diabetic and non-diabetic contexts.

We employed the well-established mouse models ob/ob (obese, insulin-resistant, metabolically non-diabetic) and db/db (obese, insulin-resistant, metabolically diabetic) (24). Note that these two mouse models have different genetic backgrounds (ob/ob: C57BL/6 carrying a mutation in the leptin gene; db/db: C57BLKS/J carrying a mutation in the leptin receptor gene). Therefore, we performed comparative analyses of mt-aaRSs expression in ob/ob mice relative to non-diabetic lean wild-type mice (C57BL/6); and an equivalent analysis in db/db mice against their diabetic lean siblings (db+). We did not observe a statistically significant differential expression of mt-aaRSs in ob/ob mice compared to C57BL/6 mice (Fig. 4A), indicating that downregulation of mt-aaRSs is not associated with obesity in a non-diabetic context. However, we found a statistically significant downregulation of CARS2, DARS2, LARS2, TARS2, and VARS2 in db/db mice with respect to db+ mice (Fig. 4B). Together, this data showed that downregulation of mt-aaRSs in diabetic obese individuals is a conserved signature in mice and human patients.

**Figure 4.**
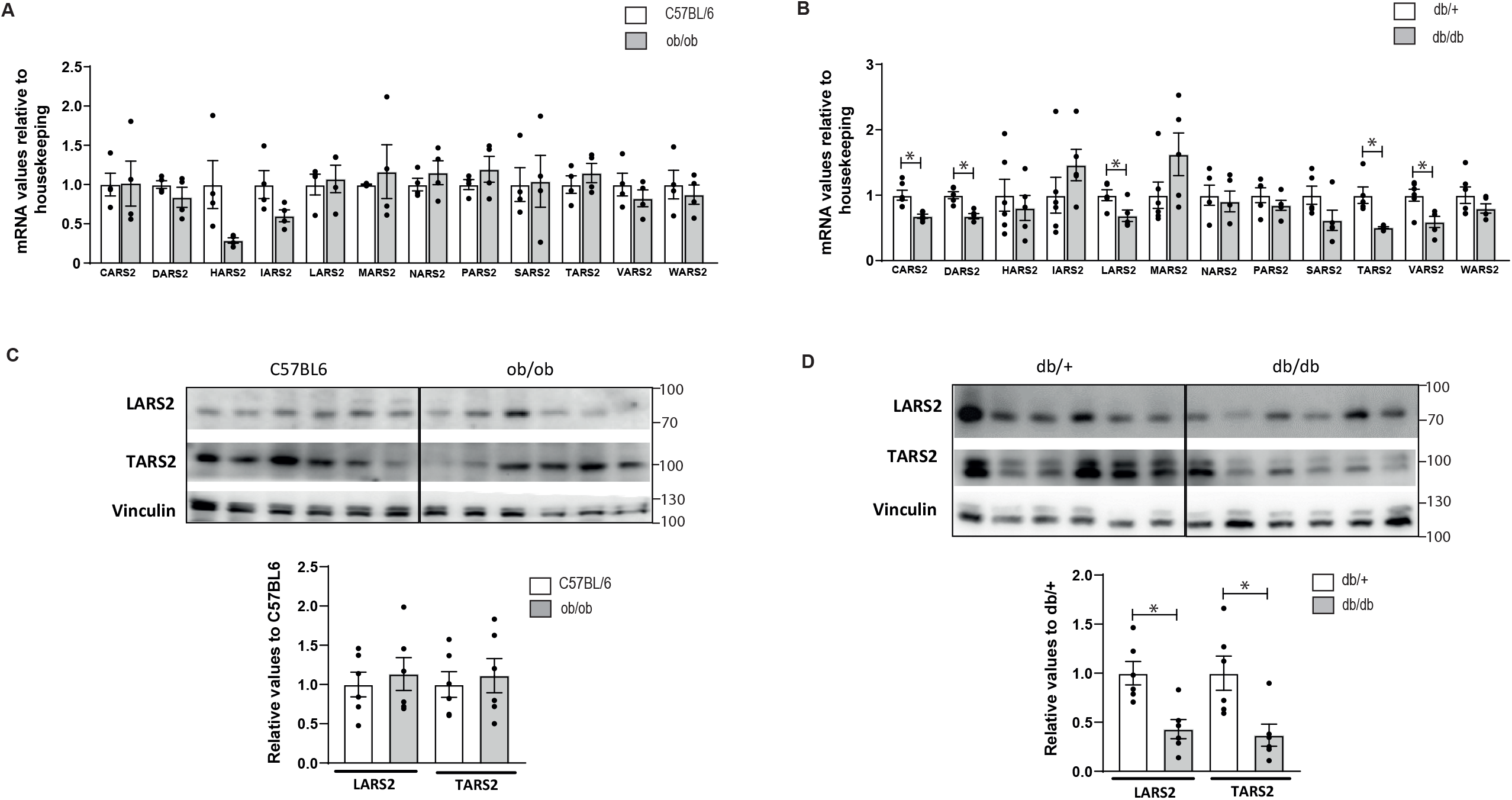
Skeletal muscle gene and protein expression of mitochondrial tRNA synthetases in obese and diabetic mice and their respective matched control group. Real-time PCR and western blot were performed in skeletal muscle biopsies. Ob/ob are obese mice, db/db are diabetic mice, and their respective controls are C57BL/6 and db/+. **(A)** Skeletal muscle gene expression of mt-aaRSs in obese mice. **(B)** Skeletal muscle gene expression of mt-aaRSs in diabetic mice. **(C)** Skeletal muscle protein expression of LARS2 and TARS2 in obese mice. **(D)** Skeletal muscle protein expression of LARS2 and TARS2 in diabetic mice. Data are mean ± SEM. Statistical analyses comparing C57BL6 vs ob/ob or db/+ vs db/db mice were performed by unpaired *t*-test *p<0.05. n=4-6/group.

### Mitochondrial mtRNA synthetases expresion is impaired in diabetic mice and leads to an imbalanced in mitochondrial OXPHOS complex formation

Reduced expression of mt-aaRSs should impact mitochondrial translation. We analyzed the amino acid composition of the 13 well-validated proteins encoded and translated in animal mitochondria (25). These proteins are highly conserved in humans (Hs) and mice (Mm). We found them to be primarily enriched in amino acids Leu (17 % Hs/ 15.5 % Mm), Ile (8.42 % Hs/9.79 % Mm), and Thr (9.26 % Hs/8.12 % Mm) (Suppl. Table 2). Notably, these amino acids are charged into mt-tRNAs by three of the most downregulated mt-aaRS that we detected in obese T2D patients (i.e. LARS2, IARS2, and TARS2, respectively) (Fig. 3E, D, J). In the case of the diabetic mice (db/db) only the gene expression of LARS2 and TARS2 was downregulated (Fig. 4B). Moreover, the protein expression of LARS2 and TARS2 was decreased in the skeletal muscle of diabetic mice (Fig. 4D) while no differences were found in ob/ob mice (Fig. 4C).

Mitochondrial-encoded proteins are involved in the formation of the mitochondrial electron transport chain (OXPHOS) complex (26). Therefore, aberrant mitochondrial protein synthesis due to defects in mitochondrial aminoacyl-tRNAs could impact OXPHOS complex formation. To assess the effects of lower expression of mt-aaRSs in the skeletal muscle of db/db mice on the OXPHOS biogenesis, we carried out western blotting analysis to examine the levels in 7 subunits of OXPHOS complexes in the skeletal muscle of db/db, ob/ob, and their respective control mice. These subunits included three mtDNA-encoding polypeptides (NADH:Ubiquinone Oxidoreductase Core Subunit 2 (ND2), cytochrome c oxidase subunit 1 (CO1) and ATP synthase subunit 6 (ATP6)), and four nucleus-encoding proteins (NADH:Ubiquinone Oxidoreductase Subunit B8 (NDUFB8), succinate dehydrogenase iron-sulfur subunit (SDHB), Ubiquinol-Cytochrome C Reductase Core Protein 2 (UQCRC2) and ATP synthase F1 subunit alpha (ATP5A)).

In obese (ob/ob) and diabetic (db/db) mice, the skeletal muscle gene expression of NDUFB8, MT-ND2, SDHB, UQCRC2, MT-CO1, ATP5 and, MT-ATP6 was similar to the corresponding control mice (Fig. 5A and B). Moreover, in obese mice, an upregulation of the protein expression of mitochondrial subunits MT-CO1 (Complex IV) and MT-ATP6 (Complex V) was observed (Fig. 5C). No changes in the protein expression of the subunits NDUFB8 and mitochondrial MT-ND2 (Complex I), SDHB (Complex II), UQCRC2 (Complex III) and ATP5A (Complex V) were detected (Fig. 5C). By contrast, in db/db mice, alterations in the protein expression of three subunits of the OXPHOS complexes were detected (Fig. 5D). These included downregulation of the subunits mitochondrial MT-ND2 (Complex I) and SDHB (Complex II); and upregulation of mitochondrial MT-CO1 (Complex IV) (Fig. 5D). Note that SDHB is a nuclear-encoded mitochondrial protein and therefore, their protein levels should not be directly dependent on mt-aaRS expression. However, MT-ND2 and MT-CO1 are encoded and translated in the mitochondria. We found a ∼ 55 % reduction in MT-ND2 protein levels in db/db mice compared to db/+ mice, consistent with impaired mitochondrial translation in obese diabetic mice. No changes were observed in the subunits NDUFB8 (Complex I), UQCRC2 (Complex III), ATP5A, and MT-ATP6 (Complex V) between db/db and db/+ mice. However, obesity caused a considerable increase in MT-ATP6 that is no longer observed in db/db mice.

**Figure 5.**
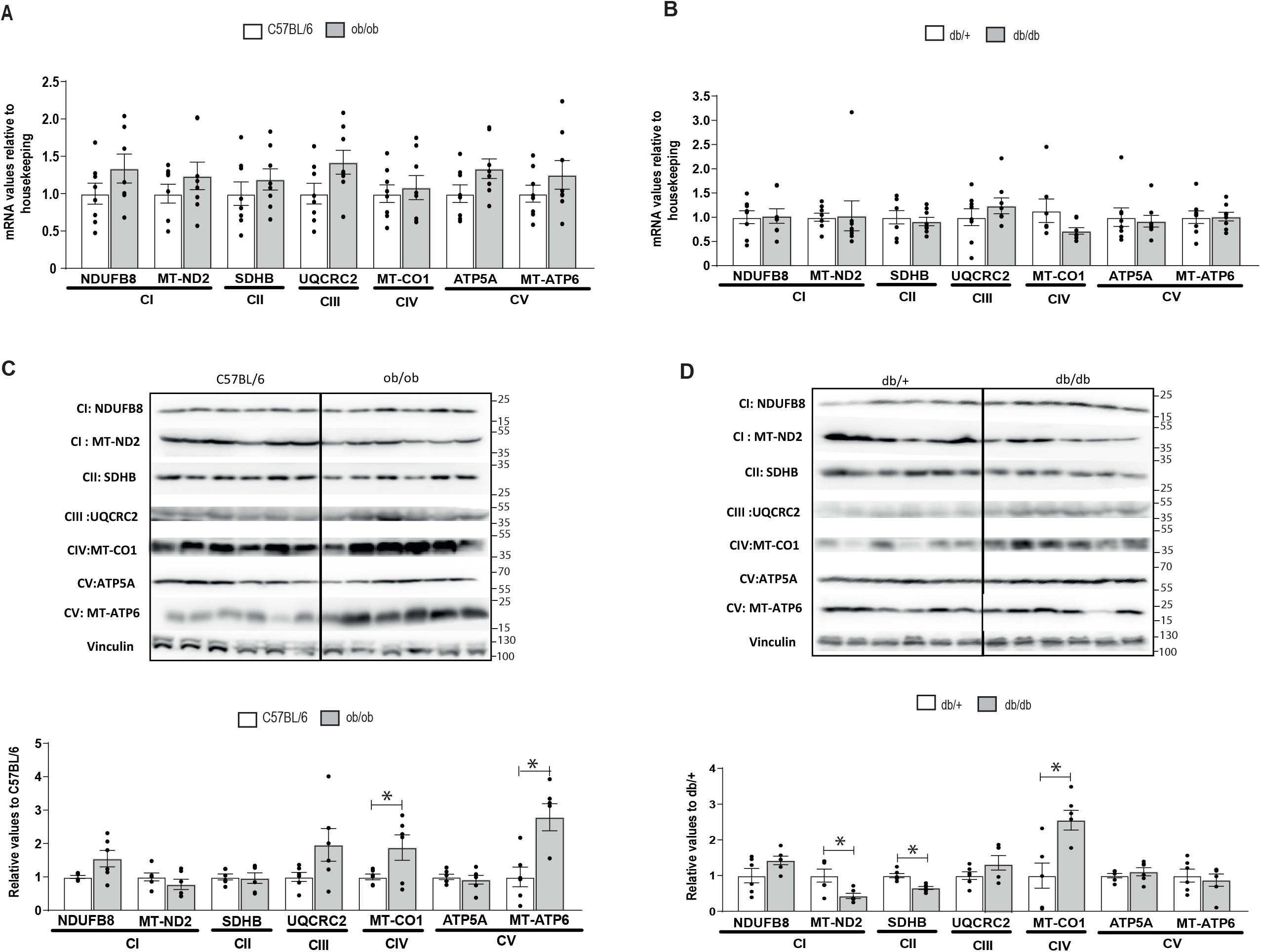
Skeletal muscle gene and protein expression of mitochondrial complexes in obese and diabetic mice and their respective matched control group. Real-time PCCR and western blot were performed in skeletal muscle biopsies. Ob/ob are obese mice, db/db are diabetic mice, and their respective controls are C57BL/6 and db/+. **(A)** Skeletal muscle gene expression of mitochondrial complexes in obese mice. Skeletal muscle gene expression of mitochondrial complexes in diabetic mice. **(C)** Skeletal muscle protein expression of mitochondrial complexes in obese mice. **(D)** Skeletal muscle protein expression of mitochondrial complexes in diabetic mice. Data are mean ± SEM. Statistical analyses comparing C57BL6 vs ob/ob or db/+ vs db/db mice were performed by unpaired *t*-test *p<0.05. n=6-8/group.

### iNOS expression is increased in enriched mitochondrial fraction from skeletal muscle in diabetic mice

It has been reported that nitrosative stress inhibits aminoacylation of mitochondrial threonyl-tRNA and leucyl-tRNA synthetases (TARS2 and LARS2) by S-nitrosation (18). Nitric oxide is synthesized enzymatically from L-arginine through the actions of the nitric oxide synthases (NOS). Therefore we measured if NOS expression was dysregulated in diabetes or obesity. iNOS and eNOS gene expression was higher in the skeletal muscle of diabetic mice (Fig. 6B), and not in obese mice (Fig. 6A). Moreover, iNOS protein expression was induced in enriched mitochondrial fraction from diabetic mice (Fig. 6D) and no differences were observed in obese mice (Fig. 6C).

**Figure 6.**
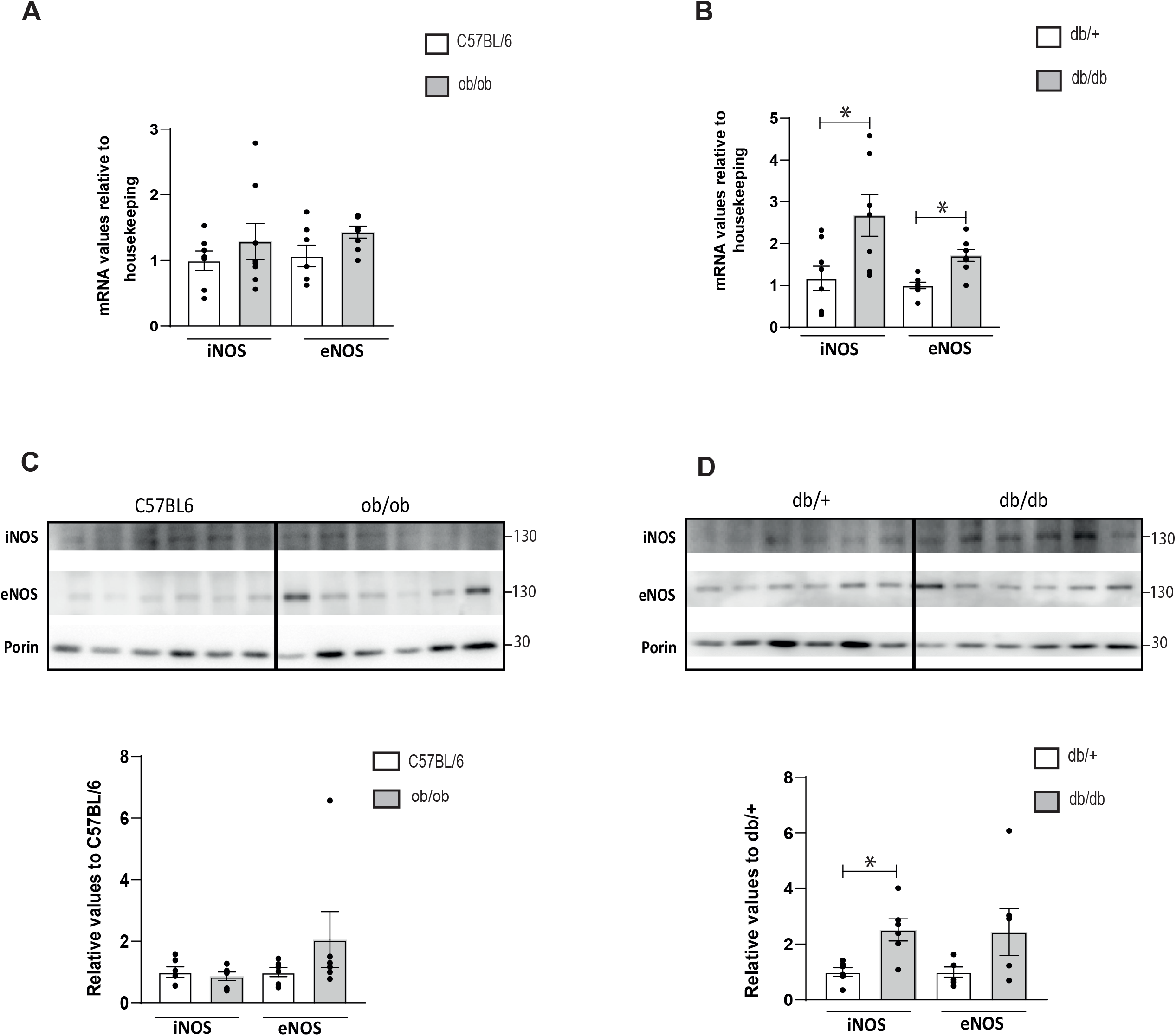
Skeletal muscle gene expression of iNOS and eNOS in obese and diabetic mice and their respective matched control group. iNOS and eNOS protein expression in mitochondria isolated from skeletal muscle of obese and diabetic mice and their respective matched control group. Real-time PCR or western blot was performed in skeletal muscle biopsies. Ob/ob are obese mice, db/db are diabetic mice, and their respective controls are C57BL/6 and db/+. **(A)** Skeletal muscle gene expression of iNOS and eNOS in obese mice. **(B)** Skeletal muscle gene expression of iNOS and eNOS in diabetic mice. **(C)** iNOS and eNOS protein expression in enriched mitochondrial fractions from skeletal muscle of obese mice. **(D)** iNOS and eNOS protein expression in enriched mitochondrial fractions from skeletal muscle of diabetic mice. Data are mean ± SEM. Statistical analyses comparing C57BL6 vs ob/ob or db/+ vs db/db mice were performed by unpaired *t*-test *p<0.05. n=6-8/group.

Altogether our data suggest that an impaired translation of mitochondrial-encoded proteins leading to an imbalance in the formation of mitochondrial OXPHOS complexes and mitochondrial dysfunction, which is one of the most important defects found in skeletal muscle from type 2 diabetic patients.

## Discussion

In this study, we focused our attention on mt-aaRSs, which constitute a class of enzymes that control cellular protein homeostasis in the mitochondria. Our results establish a diabetic signature: <u>reduced expression of mt-aaRSs in type 2 diabetic skeletal muscle</u> that is applicable independently of the age of diabetes onset and in a diabetic mice model.

Mt-aaRSs participate in the first step of protein biosynthesis charging mitochondrial tRNAs with their cognate amino acids. Mitochondrial protein biosynthesis is a critical process because the human mitochondrial genome encodes 13 proteins, which are all essential members of the OXPHOS complex. A downregulation of mt-aaRSs could be affecting the synthesis of the mitochondrial subunits of OXPHOS complexes in the skeletal muscle of diabetic patients. Recently, data from the largest proteomic analysis of human skeletal muscle reveal the downregulation of proteins of OXPHOS complexes in prediabetes and T2D (9). Among these proteins that were downregulated in T2D samples, seven were mitochondrial genome encode proteins (MT-ATP6, MT-CO1, MT-CO2, MT-ND2, MT-ND4, MT-ND5 and MT-CYB), and 49 were nuclear genome encode proteins, suggesting that reduced expression of mt-aaRSs could impact mitochondrial translation in the skeletal muscle of T2D patients.

Strikingly, in our results, a nuclear genome-encoded protein SDHB (Complex II) was downregulated in the skeletal muscle of diabetic mice. These results agree with a DARS2 (mitochondrial aspartyl-tRNA synthetase) knockout mice specifically in the skeletal muscle (27). These mice showed a substantial decrease in COX1 and COX4-1 (Complex IV), both of them mitochondrial-encoded proteins, and moderate reductions in the levels of NDUFA9 (Complex 1) and UQCRC2 (Complex 3), both of them nuclear-encoded proteins in the skeletal muscle. However, they showed a strong upregulation of MT-ND2 a mitochondrially encoded subunit that completely lacks aspartate residues (27). Also, YARS2 (mitochondrial tyrosyl-tRNA synthetase) knockout (KO) HeLa cells presented aberrant tRNA^Tyr^ aminoacylation and reductions in the levels of mitochondrion- and nucleus-encoding subunits of OXPHOS. Furthermore, YARS2 ablation caused defects in the stability and activities of OXPHOS complexes (28).

Our results also showed that in diabetic mice, a downregulation of mt-aaRSs was affecting the synthesis of the mitochondrially encoded MT-ND2 (Complex I). This subunit in mice is primarily enriched in the amino acid leucine (18,44%) and threonine (12,39%) (Suppl. Table 3). Then, the reduction of MT-ND2 correlates with the decrease in LARS2 and TARS2 protein expression that was also found in the muscle of diabetic mice. In addition, the MT-ATP6 subunit also has an important enrichment of leucine (19,47%) and threonine (11,50%) (Suppl. Table 3) and in diabetic mice did not show that considerable increased observed in ob/ob mice. This implies that MT-ATP6 is also affected by the decreased protein expression of LARS2 and TARS2 observed in db/db mice. By contrast, the contribution of threonine (6,6%) (Suppl. Table 3) is lower in the subunit MT-CO1. This observation suggests an attempt to compensate for OXPHOS defects associated with impaired mitochondrial translation by stimulating mitochondrial biogenesis as previously reported in tissues of mitochondrial disease patients (29). However, MT-CO1 is the main subunit of the cytochrome c oxidase complex and is the last enzyme in the mitochondrial electron transport chain, which drives oxidative phosphorylation where O2 is converted to H2O. In obesity condition, the upregulation of both MT-CO1 and MT-ATP6 subunits suggets more activity of the respiratory chain. More oxigen converted to H2O and more ATP production. However in diabetic condition, there is a lack of protons due to defects in Complex I and II, the overactivation of Complex IV and non enough activity of Complex V to generate ATP, will induce a release of oxygen that can be used to produce reactive oxygen species (ROS). An enhanced level of ROS could induce an overproduction of NO, which in turn, reacting with ROS, produces reactive peroxynitrite (ONOO^-^), causing mitochondrial DNA breakage and mitochondrial damage (30) as a vicious cycle.

Nitric oxide (NO) has been studied in several areas of health sciences, and it has been recognized as an important signalling molecule in several cellular pathways (31). There is evidence that mitochondria produce NO (32) and that there is a mitochondrial NOS. However, the identity of this mitochondrial NOS is still debated (33, 34). We have detected iNOS and eNOS protein expression in enriched fractions of mitochondria from skeletal muscle from mice. These results agree with several studies that demonstrate mitochondrial co-localization of the immunoreactivity using antibodies against iNOS (35) and eNOS (36). eNOS is constitutive and is activated by interactions with calcium and calmodulin, while iNOS is calcium-independent and is usually activated in defence responses such as infection and inflammation (37). eNOS produces lower concentrations of NO for short periods, while iNOS can produce high amounts of NO during long periods (31). NO can promote nitrosative modifications, which will act in physiological responses through signaling or promoting mitochondrial damage (31). It has been reported that nitrosative stress inhibits the aminoacylation of mitochondrial threonyl-tRNA and leucyl-tRNA synthetases (TARS2 and LARS2) by S-nitrosation (18). We have shown higher nitrosative stress indicators in enriched fractions of mitochondria from diabetic mice that could come from the vicinity that mitochondria have with the endoplasmic reticulum. The NO produced in this vicinity could be involved in the downregulation of TARS2 and LARS2 protein levels and with the decreased expression of mt-aaRSs due to NO action in DNA breakage. We conclude that the downregulation of mt-aaRSs plays a role in the pathogenesis of T2D and nitrosative stress is involved in this effect.

## Data Availability

All data produced in the present work are contained in the manuscript

## Abbreviations

ATP5A: ATP synthase F1 subunit Alpha
BCAAs: branched chain aminoacids
CARS2: cysteynil-tRNA synthetase 2
DARS2: aspartyl-tRNA synthetase 2
eNOS: endothelial nitric oxide synthase
GARS2: glycine-tRNA synthetase
GSEA: gene Set Enrichment Analysis
HARS2: histidyl-tRNA synthetase 2
IARS2: isoleucyl-tRNA synthetase 2
iNOS: inducible nitric oxide synthase
KARS2: lycine-tRNA synthetase
LARS2: leucyl-tRNA synthetase 2
MARS2: methionyl-tRNA synthetase 2
MT-ND2: NADH-dehydrogenase subunit 2
mt-aaRSs: mitochondrial aminoacyl-tRNA synthetases
MT-CO1: mitochondrial cytochrome c oxidase subunit 1
MT-ATP6: mitochondrial ATP synthase F0 subunit 6
MT-ND2: mitocondrial NADH:Ubiquinone Oxidoreductase Core Subunit 2
NARS2: asparaginyl-tRNA synthetase 2
NDUFB8: NADH:Ubiquinone Oxidoreductase Subunit B8
NO: nitric oxide
PARS2: prolyl-tRNA synthetase 2
RNS: reactive nitrogen species
VARS2: valyl-tRNA synthetase 2
OT2: classical type 2 diabetes
OXPHOS: oxidative phosphorylation system
PCA: principal component analysis
ROS: reactive oxygen species
SARS2: serine-tRNA synthetase 2
SDHB: succinate dehydrogenase iron-sulfur subunit
TARS2: trheonyl-tRNA synthetase 2
T2DM: type 2 diabetes mellitus
UQCRC2: Ubiquinol-Cytochrome C Reductase Core Protein 2
YT2: early-onset type 2 diabetes
YARS2: tyrosyl-tRNA synthetase 2
WARS2: tryptophanyl-tRNA synthetase 2

## Acknowledgments

We thank the Biostatistics/Bioinformatics Unit (IRB Barcelona), the Functional Genomics Facility (IRB Barcelona), and Jorge Manuel Seco (IRB Barcelona) for technological assistance. We thank all study participants who generously shared their time for the purposes of this project.

## Contribution statement

IL-S, AD-R, and MIHA conceived the study. IL-S, AD-R, RV, IM-R, AD-R, EP, DC, AP, KW, DOH, DO, TC, LR, JJN and MIH-A performed the experiments. IL-S, AD-R, RV, IM-R, EP, TC, LR and MIH-A analysed the data. IL-S and MIH-A wrote the manuscript and all authors contributed to editing. MIH-A is the guarantor of this work.

## References

1. Rocha M, Apostolova N, Diaz-Rua R, Muntane J, and Victor VM. Mitochondria and T2D: Role of Autophagy, ER Stress, and Inflammasome. Trends Endocrinol Metab. 2020;31(10):725–41.

2. DeFronzo RA, and Tripathy D. Skeletal muscle insulin resistance is the primary defect in type 2 diabetes. Diabetes Care. 2009;32 Suppl 2(S157–63.

3. Galicia-Garcia U, Benito-Vicente A, Jebari S, Larrea-Sebal A, Siddiqi H, Uribe KB, Ostolaza H, and Martin C. Pathophysiology of Type 2 Diabetes Mellitus. Int J Mol Sci. 2020;21(17).

4. Sherratt HS, and Turnbull DM. Mitochondrial oxidations and ATP synthesis in muscle. Baillieres Clin Endocrinol Metab. 1990;4(3):523–60.

5. Simoneau JA, and Kelley DE. Altered glycolytic and oxidative capacities of skeletal muscle contribute to insulin resistance in NIDDM. J Appl Physiol (1985). 1997;83(1):166–71.

6. Patti ME, Butte AJ, Crunkhorn S, Cusi K, Berria R, Kashyap S, Miyazaki Y, Kohane I, Costello M, Saccone R, et al. Coordinated reduction of genes of oxidative metabolism in humans with insulin resistance and diabetes: Potential role of PGC1 and NRF1. Proc Natl Acad Sci U S A. 2003;100(14):8466–71.

7. Mootha VK, Lindgren CM, Eriksson KF, Subramanian A, Sihag S, Lehar J, Puigserver P, Carlsson E, Ridderstrale M, Laurila E, et al. PGC-1alpha-responsive genes involved in oxidative phosphorylation are coordinately downregulated in human diabetes. Nat Genet. 2003;34(3):267–73.

8. Kelley DE, He J, Menshikova EV, and Ritov VB. Dysfunction of mitochondria in human skeletal muscle in type 2 diabetes. Diabetes. 2002;51(10):2944–50.

9. Ohman T, Teppo J, Datta N, Makinen S, Varjosalo M, and Koistinen HA. Skeletal muscle proteomes reveal downregulation of mitochondrial proteins in transition from prediabetes into type 2 diabetes. iScience. 2021;24(7):102712.

10. Petersen KF, Dufour S, Befroy D, Garcia R, and Shulman GI. Impaired mitochondrial activity in the insulin-resistant offspring of patients with type 2 diabetes. N Engl J Med. 2004;350(7):664–71.

11. Sergi D, Naumovski N, Heilbronn LK, Abeywardena M, O’Callaghan N, Lionetti L, and Luscombe-Marsh N. Mitochondrial (Dys)function and Insulin Resistance: From Pathophysiological Molecular Mechanisms to the Impact of Diet. Front Physiol. 2019;10(532.

12. Delarue M. Aminoacyl-tRNA synthetases. Curr Opin Struct Biol. 1995;5(1):48–55.

13. Garin S, Levi O, Cohen B, Golani-Armon A, and Arava YS. Localization and RNA Binding of Mitochondrial Aminoacyl tRNA Synthetases. Genes (Basel). 2020;11(10).

14. Moulinier L, Ripp R, Castillo G, Poch O, and Sissler M. MiSynPat: An integrated knowledge base linking clinical, genetic, and structural data for disease-causing mutations in human mitochondrial aminoacyl-tRNA synthetases. Hum Mutat. 2017;38(10):1316–24.

15. Diodato D, Ghezzi D, and Tiranti V. The Mitochondrial Aminoacyl tRNA Synthetases: Genes and Syndromes. Int J Cell Biol. 2014;2014(787956.

16. Oprescu SN, Griffin LB, Beg AA, and Antonellis A. Predicting the pathogenicity of aminoacyl-tRNA synthetase mutations. Methods. 2017;113(139–51.

17. Sissler M, Gonzalez-Serrano LE, and Westhof E. Recent Advances in Mitochondrial Aminoacyl-tRNA Synthetases and Disease. Trends Mol Med. 2017;23(8):693–708.

18. Zheng WQ, Zhang Y, Yao Q, Chen Y, Qiao X, Wang ED, Chen C, and Zhou XL. Nitrosative stress inhibits aminoacylation and editing activities of mitochondrial threonyl-tRNA synthetase by S-nitrosation. Nucleic Acids Res. 2020;48(12):6799–810.

19. Wang PG, Xian M, Tang X, Wu X, Wen Z, Cai T, and Janczuk AJ. Nitric oxide donors: chemical activities and biological applications. Chem Rev. 2002;102(4):1091–134.

20. Yoon S, Eom GH, and Kang G. Nitrosative Stress and Human Disease: Therapeutic Potential of Denitrosylation. Int J Mol Sci. 2021;22(18).

21. Hernandez-Alvarez MI, Diaz-Ramos A, Berdasco M, Cobb J, Planet E, Cooper D, Pazderska A, Wanic K, O’Hanlon D, Gomez A, et al. Early-onset and classical forms of type 2 diabetes show impaired expression of genes involved in muscle branched-chain amino acids metabolism. Sci Rep. 2017;7(1):13850.

22. Burns N, Finucane FM, Hatunic M, Gilman M, Murphy M, Gasparro D, Mari A, Gastaldelli A, and Nolan JJ. Early-onset type 2 diabetes in obese white subjects is characterised by a marked defect in beta cell insulin secretion, severe insulin resistance and a lack of response to aerobic exercise training. Diabetologia. 2007;50(7):1500–8.

23. Hernandez-Alvarez MI, Thabit H, Burns N, Shah S, Brema I, Hatunic M, Finucane F, Liesa M, Chiellini C, Naon D, et al. Subjects with early-onset type 2 diabetes show defective activation of the skeletal muscle PGC-1alpha/Mitofusin-2 regulatory pathway in response to physical activity. Diabetes Care. 2010;33(3):645–51.

24. Kennedy AJ, Ellacott KL, King VL, and Hasty AH. Mouse models of the metabolic syndrome. Dis Model Mech. 2010;3(3-4):156–66.

25. Capt C, Passamonti M, and Breton S. The human mitochondrial genome may code for more than 13 proteins. Mitochondrial DNA Part A, DNA mapping, sequencing, and analysis. 2016;27(5):3098–101.

26. Scheffler IE. A century of mitochondrial research: achievements and perspectives. Mitochondrion. 2001;1(1):3–31.

27. Dogan SA, Pujol C, Maiti P, Kukat A, Wang S, Hermans S, Senft K, Wibom R, Rugarli EI, and Trifunovic A. Tissue-specific loss of DARS2 activates stress responses independently of respiratory chain deficiency in the heart. Cell Metab. 2014;19(3):458–69.

28. Jin X, Zhang Z, Nie Z, Wang C, Meng F, Yi Q, Chen M, Sun J, Zou J, Jiang P, et al. An animal model for mitochondrial tyrosyl-tRNA synthetase deficiency reveals links between oxidative phosphorylation and retinal function. J Biol Chem. 2021;296(100437.

29. Heddi A, Stepien G, Benke PJ, and Wallace DC. Coordinate induction of energy gene expression in tissues of mitochondrial disease patients. J Biol Chem. 1999;274(33):22968–76.

30. Bhatti JS, Bhatti GK, and Reddy PH. Mitochondrial dysfunction and oxidative stress in metabolic disorders - A step towards mitochondria based therapeutic strategies. Biochim Biophys Acta Mol Basis Dis. 2017;1863(5):1066–77.

31. Tengan CH, and Moraes CT. NO control of mitochondrial function in normal and transformed cells. Biochim Biophys Acta Bioenerg. 2017;1858(8):573–81.

32. Giulivi C, Poderoso JJ, and Boveris A. Production of nitric oxide by mitochondria. J Biol Chem. 1998;273(18):11038–43.

33. Tengan CH, Rodrigues GS, and Godinho RO. Nitric oxide in skeletal muscle: role on mitochondrial biogenesis and function. Int J Mol Sci. 2012;13(12):17160–84.

34. Figueira TR, Barros MH, Camargo AA, Castilho RF, Ferreira JC, Kowaltowski AJ, Sluse FE, Souza-Pinto NC, and Vercesi AE. Mitochondria as a source of reactive oxygen and nitrogen species: from molecular mechanisms to human health. Antioxid Redox Signal. 2013;18(16):2029–74.

35. Tatoyan A, and Giulivi C. Purification and characterization of a nitric-oxide synthase from rat liver mitochondria. J Biol Chem. 1998;273(18):11044–8.

36. Bates TE, Loesch A, Burnstock G, and Clark JB. Mitochondrial nitric oxide synthase: a ubiquitous regulator of oxidative phosphorylation? Biochem Biophys Res Commun. 1996;218(1):40–4.

37. Nathan C, and Xie QW. Nitric oxide synthases: roles, tolls, and controls. Cell. 1994;78(6):915–8.

