## Supplementary material for "Decreased expression of mitochondrial aminoacyl-tRNA synthetases causes downregulation of mitochondrial OXPHOS subunits in type 2 diabetic skeletal muscle": Suplemental Table1

| probeset | entrezid | symbol |
|---|---|---|
| 11715255_s_at | 259293 | TAS2R30 |
| 11715293_s_at | 83707 | TRPT1 |
| 11715315_s_at | NA | NA |
| 11715497_a_at | 5018 | OXA1L |
| 11715514_a_at | 9709 | HERPUD1 |
| 11715618_a_at | 51150 | SDF4 |
| 11715733_a_at | 8508 | NIPSNAP1 |
| 11715947_a_at | 115207 | KCTD12 |
| 11715966_s_at | NA | NA |
| 11715995_at | 54948 | MRPL16 |
| 11716033_at | 6590 | SLPI |
| 11716047_a_at | 8209 | C21orf33 |
| 11716081_a_at | 79084 | WDR77 |
| 11716180_at | 84313 | VPS25 |
| 11716210_at | 10370 | CITED2 |
| 11716243_a_at | 84988 | PPP1R16A |
| 11716272_a_at | 5434 | POLR2E |
| 11716273_x_at | 5434 | POLR2E |
| 11716276_a_at | 23468 | CBX5 |
| 11716277_a_at | 23468 | CBX5 |
| 11716278_s_at | 23468 | CBX5 |
| 11716280_a_at | 23468 | CBX5 |
| 11716301_a_at | 3685 | ITGAV |
| 11716338_a_at | 3638 | INSIG1 |
| 11716550_at | 3280 | HES1 |
| 11716642_s_at | 1841 | DTYMK |
| 11716737_at | 81572 | PDRG1 |
| 11716747_a_at | 4299 | AFF1 |
| 11716782_a_at | 79718 | TBL1XR1 |
| 11716783_a_at | 79718 | TBL1XR1 |
| 11716891_at | 90871 | C9orf123 |
| 11716897_x_at | 10450 | PPIE |
| 11716980_a_at | 113829 | SLC35A4 |
| 11717031_a_at | 9812 | KIAA0141 |
| 11717032_at | 9812 | KIAA0141 |
| 11717033_a_at | 9812 | KIAA0141 |
| 11717054_a_at | 6745 | SSR1 |
| 11717205_at | 55316 | RSAD1 |
| 11717244_at | 25804 | LSM4 |
| 11717262_a_at | 84310 | C7orf50 |
| 11717269_a_at | 2356 | FPGS |
| 11717338_a_at | 1523 | CUX1 |
| 11717339_s_at | 6652 | SORD |
| 11717354_at | 5092 | PCBD1 |
| 11717364_a_at | 83596 | BCL2L12 |
| 11717489_a_at | 5413 | SEPT5 |
| 11717620_a_at | 4047 | LSS |
| 11717654_a_at | NA | NA |
| 11717715_at | 64778 | FNDC3B |
| 11717716_at | 64778 | FNDC3B |
| 11717717_at | 64778 | FNDC3B |
| 11717788_at | 25980 | C20orf4 |

| | | |
|---|---|---|
| 11717795_a_at | 51304 | ZDHHC3 |
| 11717868_a_at | 10127 | ZNF263 |
| 11717963_a_at | 3845 | KRAS |
| 11717987_a_at | 64976 | MRPL40 |
| 11718011_a_at | 51073 | MRPL4 |
| 11718117_a_at | 9054 | NFS1 |
| 11718152_a_at | 8495 | PPFIBP2 |
| 11718162_a_at | 10445 | MCRS1 |
| 11718227_a_at | 617 | BCS1L |
| 11718285_s_at | 84311 | MRPL45 |
| 11718323_at | 23527 | ACAP2 |
| 11718379_x_at | 51422 | PRKAG2 |
| 11718436_a_at | 91647 | ATPAF2 |
| 11718437_at | 91647 | ATPAF2 |
| 11718537_s_at | NA | NA |
| 11718553_at | 54726 | OTUD4 |
| 11718580_a_at | 57017 | COQ9 |
| 11718631_at | 29950 | SERTAD1 |
| 11718639_a_at | 9632 | SEC24C |
| 11718651_at | 83857 | TMTC1 |
| 11718685_a_at | 10587 | TXNRD2 |
| 11718701_a_at | 27236 | ARFIP1 |
| 11718727_x_at | 79027 | ZNF655 |
| 11718782_s_at | 11018 | TMED1 |
| 11718927_a_at | 84159 | ARID5B |
| 11718987_at | 23395 | LARS2 |
| 11719013_at | 8322 | FZD4 |
| 11719082_a_at | 90480 | GADD45GIP1 |
| 11719083_x_at | 26995 | TRUB2 |
| 11719161_a_at | 84706 | GPT2 |
| 11719195_a_at | 9533 | POLR1C |
| 11719224_s_at | 93058 | COQ10A |
| 11719234_at | 51285 | RASL12 |
| 11719246_a_at | 2952 | GSTT1 |
| 11719256_a_at | 4040 | LRP6 |
| 11719286_s_at | 8799 | PEX11B |
| 11719310_a_at | 1408 | CRY2 |
| 11719457_s_at | 10640 | EXOC5 |
| 11719558_at | 90196 | SYS1 |
| 11719606_a_at | 54700 | RRN3 |
| 11719607_x_at | 54700 | RRN3 |
| 11719637_a_at | 56940 | DUSP22 |
| 11719859_a_at | 56731 | SLC2A4RG |
| 11719877_a_at | 83707 | TRPT1 |
| 11719918_s_at | 53918 | PELO |
| 11720009_at | 7727 | ZNF174 |
| 11720013_a_at | 472 | ATM |
| 11720045_a_at | 54431 | DNAJC10 |
| 11720056_a_at | 23173 | METAP1 |
| 11720084_x_at | 84304 | NUDT22 |
| 11720133_a_at | 5627 | PROS1 |
| 11720180_at | 55794 | DDX28 |
| 11720188_a_at | 4701 | NDUFA7 |

| | | |
|---|---|---|
| 11720221_at | 2673 | GFPT1 |
| 11720329_at | 55178 | RNMTL1 |
| 11720366_at | 112812 | FDX1L |
| 11720406_a_at | 9701 | PPP6R2 |
| 11720447_s_at | 6662 | SOX9 |
| 11720469_a_at | 100131187 | TSTD1 |
| 11720526_s_at | 5037 | PEBP1 |
| 11720547_a_at | 23590 | PDSS1 |
| 11720619_a_at | 148223 | C19orf25 |
| 11720639_a_at | 23107 | MRPS27 |
| 11720640_x_at | 23107 | MRPS27 |
| 11720642_a_at | 3712 | IVD |
| 11720643_a_at | 3712 | IVD |
| 11720662_at | 115939 | C16orf42 |
| 11720695_at | 79630 | C1orf54 |
| 11720709_x_at | 23095 | KIF1B |
| 11720743_x_at | 54677 | CROT |
| 11720771_x_at | 10105 | PPIF |
| 11720809_x_at | 80097 | MZT2B |
| 11720900_x_at | 54762 | GRAMD1C |
| 11720921_s_at | 30 | ACAA1 |
| 11721012_at | 64093 | SMOC1 |
| 11721033_a_at | 11170 | FAM107A |
| 11721044_a_at | 90459 | ERI1 |
| 11721096_a_at | 81542 | TMX1 |
| 11721157_at | 4913 | NTHL1 |
| 11721176_at | 9858 | PPP1R26 |
| 11721216_s_at | 54664 | TMEM106B |
| 11721251_at | 4236 | MFAP1 |
| 11721287_a_at | 51741 | WWOX |
| 11721306_at | 22822 | PHLDA1 |
| 11721311_s_at | 10772 | SRSF10 |
| 11721325_a_at | 23464 | GCAT |
| 11721519_at | 23390 | ZDHHC17 |
| 11721727_x_at | 6878 | TAF6 |
| 11721735_at | 55526 | DHTKD1 |
| 11721840_at | 11010 | GLIPR1 |
| 11721904_a_at | 79140 | CCDC28B |
| 11721914_a_at | 4194 | MDM4 |
| 11721915_a_at | 4194 | MDM4 |
| 11721920_at | 205564 | SENP5 |
| 11721941_x_at | 1577 | CYP3A5 |
| 11721961_at | 54542 | RC3H2 |
| 11722029_a_at | 9690 | UBE3C |
| 11722042_s_at | 57107 | PDSS2 |
| 11722062_at | 221035 | REEP3 |
| 11722087_a_at | 9975 | NR1D2 |
| 11722110_x_at | 22990 | PCNX |
| 11722154_a_at | 166929 | SGMS2 |
| 11722173_x_at | 55245 | UQCC |
| 11722174_a_at | 55245 | UQCC |
| 11722196_x_at | 4836 | NMT1 |
| 11722279_a_at | 80012 | PHC3 |

| | | |
|---|---|---|
| 11722281_a_at | 80012 | PHC3 |
| 11722372_a_at | 23062 | GGA2 |
| 11722426_a_at | 10875 | FGL2 |
| 11722446_a_at | 23410 | SIRT3 |
| 11722447_a_at | 23410 | SIRT3 |
| 11722572_at | 890 | CCNA2 |
| 11722579_a_at | 29082 | CHMP4A |
| 11722602_at | 79156 | PLEKHF1 |
| 11722656_s_at | 9263 | STK17A |
| 11722690_at | 6711 | SPTBN1 |
| 11722699_a_at | 84162 | KIAA1109 |
| 11722747_at | 84376 | HOOK3 |
| 11722748_at | 84376 | HOOK3 |
| 11722764_at | 284403 | WDR62 |
| 11722797_a_at | 90488 | C12orf23 |
| 11722808_at | 6667 | SP1 |
| 11722814_s_at | NA | NA |
| 11722850_a_at | 330 | BIRC3 |
| 11722902_a_at | 54482 | CCDC76 |
| 11722985_a_at | 22888 | UBOX5 |
| 11722998_a_at | 4329 | ALDH6A1 |
| 11723083_a_at | 51409 | HEMK1 |
| 11723089_s_at | NA | NA |
| 11723090_x_at | 91966 | CXorf40A |
| 11723143_at | 259232 | NALCN |
| 11723151_s_at | 79783 | C7orf10 |
| 11723231_s_at | NA | NA |
| 11723238_at | 2788 | GNG7 |
| 11723239_a_at | 2788 | GNG7 |
| 11723313_s_at | 5827 | PXMP2 |
| 11723365_a_at | 50813 | COPS7A |
| 11723505_at | 23060 | ZNF609 |
| 11723507_s_at | 23060 | ZNF609 |
| 11723521_x_at | 25946 | ZNF385A |
| 11723601_a_at | 23130 | ATG2A |
| 11723631_a_at | 28511 | NKIRAS2 |
| 11723681_a_at | 9736 | USP34 |
| 11723684_a_at | 27079 | RPUSD2 |
| 11723749_a_at | NA | NA |
| 11723790_a_at | 6620 | SNCB |
| 11723793_s_at | 114885 | OSBPL11 |
| 11724042_a_at | 57595 | PDZD4 |
| 11724051_at | 55128 | TRIM68 |
| 11724058_x_at | 80772 | GLTPD1 |
| 11724064_a_at | 60386 | SLC25A19 |
| 11724084_a_at | 126823 | KLHDC9 |
| 11724123_a_at | 54753 | ZNF853 |
| 11724128_a_at | 140876 | FAM65C |
| 11724214_a_at | 283377 | SPRYD4 |
| 11724241_at | 6925 | TCF4 |
| 11724261_a_at | 7204 | TRIO |
| 11724264_a_at | 64792 | RABL5 |
| 11724283_a_at | 306 | ANXA3 |

| | | |
|---|---|---|
| 11724338_a_at | 222553 | SLC35F1 |
| 11724339_a_at | 222553 | SLC35F1 |
| 11724356_a_at | 80775 | TMEM177 |
| 11724366_a_at | 9677 | PPIP5K1 |
| 11724526_at | 81889 | FAHD1 |
| 11724527_x_at | 81889 | FAHD1 |
| 11724562_x_at | 54509 | RHOF |
| 11724574_a_at | 9718 | ECE2 |
| 11724588_a_at | 148741 | ANKRD35 |
| 11724622_a_at | 3249 | HPN |
| 11724639_a_at | 5051 | PAFAH2 |
| 11724641_a_at | 9912 | ARHGAP44 |
| 11724651_a_at | 64327 | LMBR1 |
| 11724720_a_at | 79695 | GALNT12 |
| 11724721_a_at | 79695 | GALNT12 |
| 11724723_a_at | 3306 | HSPA2 |
| 11724907_a_at | 11183 | MAP4K5 |
| 11724936_a_at | 79591 | C10orf76 |
| 11724950_a_at | 132864 | CPEB2 |
| 11724958_at | 56953 | NT5M |
| 11725008_a_at | 23157 | SEPT6 |
| 11725117_a_at | 57486 | NLN |
| 11725144_a_at | 147808 | ZNF784 |
| 11725163_x_at | 51302 | CYP39A1 |
| 11725216_s_at | 8547 | FCN3 |
| 11725237_a_at | 348093 | RBPMS2 |
| 11725358_a_at | 176 | ACAN |
| 11725522_at | 55897 | MESP1 |
| 11725539_at | 8912 | CACNA1H |
| 11725643_a_at | 64949 | MRPS26 |
| 11725721_a_at | 79922 | MRM1 |
| 11725791_a_at | 4361 | MRE11A |
| 11725841_at | 80345 | ZSCAN16 |
| 11725924_a_at | 83593 | RASSF5 |
| 11725994_a_at | 65003 | MRPL11 |
| 11726019_at | 80820 | EEPD1 |
| 11726074_a_at | 8874 | ARHGEF7 |
| 11726086_x_at | 91574 | C12orf65 |
| 11726114_at | 115572 | FAM46B |
| 11726131_a_at | 5318 | PKP2 |
| 11726133_a_at | 6853 | SYN1 |
| 11726266_a_at | 3421 | IDH3G |
| 11726322_at | 51 | ACOX1 |
| 11726327_at | 54165 | DCUN1D1 |
| 11726340_a_at | 256281 | NUDT14 |
| 11726361_a_at | 4287 | ATXN3 |
| 11726431_a_at | 26031 | OSBPL3 |
| 11726433_a_at | 26031 | OSBPL3 |
| 11726587_a_at | 80222 | TARS2 |
| 11726633_s_at | 81603 | TRIM8 |
| 11726638_at | 57534 | MIB1 |
| 11726751_at | 79929 | MAP6D1 |
| 11726756_a_at | 993 | CDC25A |

| | | |
|---|---|---|
| 11726884_a_at | 197257 | LDHD |
| 11726968_a_at | 55653 | BCAS4 |
| 11726969_x_at | 55653 | BCAS4 |
| 11726996_at | 65018 | PINK1 |
| 11727125_a_at | 25945 | PVRL3 |
| 11727246_at | 1140 | CHRNB1 |
| 11727247_x_at | 1140 | CHRNB1 |
| 11727294_s_at | NA | NA |
| 11727400_a_at | 2954 | GSTZ1 |
| 11727416_a_at | 51084 | CRYL1 |
| 11727417_a_at | 55101 | ATP5SL |
| 11727418_s_at | 55101 | ATP5SL |
| 11727461_x_at | 79364 | ZXDC |
| 11727544_a_at | 587 | BCAT2 |
| 11727572_s_at | 89857 | KLHL6 |
| 11727593_a_at | 158358 | KIAA2026 |
| 11727729_x_at | 79673 | ZNF329 |
| 11727745_at | 196541 | METTL21C |
| 11727776_at | 2773 | GNAI3 |
| 11727813_a_at | 6426 | SRSF1 |
| 11727836_a_at | NA | NA |
| 11727943_a_at | 2359 | FPR3 |
| 11728004_s_at | 55103 | RALGPS2 |
| 11728016_at | 9866 | TRIM66 |
| 11728033_at | 375035 | SFT2D2 |
| 11728059_s_at | 29946 | SERTAD3 |
| 11728120_at | 80759 | KHDC1 |
| 11728130_a_at | 54919 | HEATR2 |
| 11728178_at | 23409 | SIRT4 |
| 11728248_a_at | 10102 | TSFM |
| 11728290_a_at | 4068 | SH2D1A |
| 11728312_at | 7707 | ZNF148 |
| 11728327_at | 4958 | OMD |
| 11728354_a_at | 84269 | CHCHD5 |
| 11728429_a_at | 84458 | LCOR |
| 11728437_at | 54014 | BRWD1 |
| 11728470_a_at | 121551 | BTBD11 |
| 11728537_a_at | 4163 | MCC |
| 11728538_a_at | 4163 | MCC |
| 11728554_at | 90423 | ATP6V1E2 |
| 11728634_a_at | 23431 | AP4E1 |
| 11728673_s_at | 3835 | KIF22 |
| 11728691_a_at | 23608 | MKRN1 |
| 11728728_s_at | 154 | ADRB2 |
| 11728826_s_at | 388610 | TRNP1 |
| 11728879_at | 5534 | PPP3R1 |
| 11728908_at | 145173 | B3GALTL |
| 11728951_at | 56169 | GSDMC |
| 11728988_at | 151313 | FAHD2B |
| 11729023_at | 3741 | KCNA5 |
| 11729034_s_at | 162073 | ITPRIPL2 |
| 11729190_a_at | 4149 | MAX |
| 11729316_a_at | 79763 | ISOC2 |

| | | |
|---|---|---|
| 11729353_a_at | 4012 | LNPEP |
| 11729371_a_at | 7704 | ZBTB16 |
| 11729394_a_at | 143279 | HECTD2 |
| 11729401_x_at | 26974 | ZNF285 |
| 11729402_at | 26974 | ZNF285 |
| 11729446_a_at | 1638 | DCT |
| 11729505_a_at | 374659 | HDDC3 |
| 11729654_a_at | 4087 | SMAD2 |
| 11729667_a_at | 8996 | NOL3 |
| 11729670_a_at | 622 | BDH1 |
| 11729682_a_at | 79583 | TMEM231 |
| 11729704_a_at | 93134 | ZNF561 |
| 11729725_at | 10773 | ZBTB6 |
| 11729828_at | 90362 | FAM110B |
| 11729911_a_at | 200205 | IBA57 |
| 11729912_at | 200205 | IBA57 |
| 11729923_a_at | 80194 | TMEM134 |
| 11729932_a_at | 7694 | ZNF135 |
| 11729933_x_at | 7694 | ZNF135 |
| 11729952_a_at | 84614 | ZBTB37 |
| 11729982_a_at | 57097 | PARP11 |
| 11730015_s_at | 4761 | NEUROD2 |
| 11730164_a_at | 4591 | TRIM37 |
| 11730213_s_at | 51081 | MRPS7 |
| 11730249_at | 28957 | MRPS28 |
| 11730433_a_at | 84905 | ZNF341 |
| 11730465_at | 3759 | KCNJ2 |
| 11730466_a_at | 3759 | KCNJ2 |
| 11730519_a_at | 10350 | ABCA9 |
| 11730542_x_at | 30 | ACAA1 |
| 11730594_at | 55096 | EBLN2 |
| 11730620_a_at | 117283 | IP6K3 |
| 11730646_a_at | 9427 | ECEL1 |
| 11730732_at | 1777 | DNASE2 |
| 11730733_x_at | 1777 | DNASE2 |
| 11730795_at | 51073 | MRPL4 |
| 11730838_a_at | 131870 | NUDT16 |
| 11730890_at | 221078 | NSUN6 |
| 11730932_at | 79611 | ACSS3 |
| 11731021_a_at | 118430 | MUCL1 |
| 11731023_a_at | 6314 | ATXN7 |
| 11731024_s_at | 6314 | ATXN7 |
| 11731062_s_at | 9658 | ZNF516 |
| 11731089_a_at | 1652 | DDT |
| 11731230_a_at | 285367 | RPUSD3 |
| 11731356_a_at | 117283 | IP6K3 |
| 11731465_a_at | 1075 | CTSC |
| 11731466_a_at | 1075 | CTSC |
| 11731474_at | 6424 | SFRP4 |
| 11731565_x_at | 4193 | MDM2 |
| 11731566_a_at | 4193 | MDM2 |
| 11731570_a_at | 29903 | CCDC106 |
| 11731615_s_at | 1268 | CNR1 |

| | | |
|---|---|---|
| 11731683_a_at | 55183 | RIF1 |
| 11731698_s_at | NA | NA |
| 11731702_a_at | 29121 | CLEC2D |
| 11731751_at | 57674 | RNF213 |
| 11731752_at | 57674 | RNF213 |
| 11731907_s_at | 80204 | FBXO11 |
| 11731924_a_at | 283518 | KCNRG |
| 11731952_s_at | 284451 | ODF3L2 |
| 11731997_at | 25893 | TRIM58 |
| 11732080_a_at | 83856 | FSD1L |
| 11732138_at | 26517 | TIMM13 |
| 11732149_a_at | 80344 | DCAF11 |
| 11732150_x_at | 80344 | DCAF11 |
| 11732151_a_at | 80344 | DCAF11 |
| 11732158_a_at | 9510 | ADAMTS1 |
| 11732188_at | 55361 | PI4K2A |
| 11732227_a_at | 5192 | PEX10 |
| 11732297_at | 80856 | KIAA1715 |
| 11732298_a_at | 80856 | KIAA1715 |
| 11732300_at | 80856 | KIAA1715 |
| 11732313_at | 81573 | ANKRD13C |
| 11732504_a_at | 3249 | HPN |
| 11732515_x_at | 578 | BAK1 |
| 11732632_at | 3204 | HOXA7 |
| 11732844_a_at | 57553 | MICAL3 |
| 11733014_a_at | 57674 | RNF213 |
| 11733058_a_at | 9991 | PTBP3 |
| 11733059_a_at | 9991 | PTBP3 |
| 11733093_a_at | 80020 | FOXRED2 |
| 11733096_a_at | 84993 | UBL7 |
| 11733270_at | 339983 | NAT8L |
| 11733462_at | 26032 | SUSD5 |
| 11733490_at | 619279 | ZNF704 |
| 11733495_a_at | 170622 | COMMD6 |
| 11733496_x_at | 170622 | COMMD6 |
| 11733593_a_at | 5858 | PZP |
| 11733615_at | 80031 | SEMA6D |
| 11733731_s_at | 593 | BCKDHA |
| 11733835_a_at | 252839 | TMEM9 |
| 11733875_a_at | 10051 | SMC4 |
| 11733893_a_at | 55666 | NPLOC4 |
| 11733981_a_at | 5163 | PDK1 |
| 11733993_at | 441027 | TMEM150C |
| 11734022_at | 9953 | HS3ST3B1 |
| 11734023_a_at | NA | NA |
| 11734024_x_at | NA | NA |
| 11734038_a_at | 84364 | ARFGAP2 |
| 11734103_a_at | 55277 | FGGY |
| 11734126_a_at | 10392 | NOD1 |
| 11734162_a_at | 22797 | TFEC |
| 11734199_a_at | 284391 | ZNF844 |
| 11734244_a_at | 83734 | ATG10 |
| 11734273_s_at | 2841 | GPR18 |

| | | |
|---|---|---|
| 11734325_a_at | 253559 | CADM2 |
| 11734643_a_at | 255189 | PLA2G4F |
| 11734737_a_at | 84188 | FAR1 |
| 11734829_a_at | 10216 | PRG4 |
| 11734953_at | 55796 | MBNL3 |
| 11735036_a_at | 114815 | SORCS1 |
| 11735038_a_at | 114815 | SORCS1 |
| 11735119_x_at | 4077 | NBR1 |
| 11735134_a_at | 7093 | TLL2 |
| 11735167_x_at | 5982 | RFC2 |
| 11735219_at | 1300 | COL10A1 |
| 11735279_a_at | 80059 | LRRTM4 |
| 11735504_a_at | 58484 | NLRC4 |
| 11735578_a_at | 157680 | VPS13B |
| 11735643_a_at | 5918 | RARRES1 |
| 11735676_a_at | 29948 | OSGIN1 |
| 11735695_a_at | 381 | ARF5 |
| 11735728_x_at | 25946 | ZNF385A |
| 11735844_s_at | NA | NA |
| 11735845_x_at | 9720 | CCDC144A |
| 11735858_a_at | 284485 | RIIAD1 |
| 11735878_s_at | NA | NA |
| 11735891_a_at | 1896 | EDA |
| 11735934_at | 7483 | WNT9A |
| 11736057_s_at | 119032 | C10orf32 |
| 11736131_a_at | 131566 | DCBLD2 |
| 11736401_a_at | 196403 | DTX3 |
| 11736405_a_at | 1786 | DNMT1 |
| 11736473_a_at | 6932 | TCF7 |
| 11736475_x_at | 6932 | TCF7 |
| 11736494_a_at | 23112 | TNRC6B |
| 11736495_a_at | 23112 | TNRC6B |
| 11736551_x_at | 5966 | REL |
| 11736565_at | 5784 | PTPN14 |
| 11736581_a_at | 2650 | GCNT1 |
| 11736732_a_at | 147991 | DPY19L3 |
| 11736814_at | 54810 | GIPC2 |
| 11736838_a_at | 9734 | HDAC9 |
| 11736849_a_at | 83856 | FSD1L |
| 11736977_x_at | 170959 | ZNF431 |
| 11737134_x_at | 25946 | ZNF385A |
| 11737271_a_at | 390980 | ZNF805 |
| 11737320_a_at | 6239 | RREB1 |
| 11737321_a_at | 6239 | RREB1 |
| 11737454_x_at | 284390 | ZNF763 |
| 11737497_at | 151 | ADRA2B |
| 11737589_a_at | 57157 | PHTF2 |
| 11737591_a_at | 114803 | MYSM1 |
| 11737611_a_at | 340481 | ZDHHC21 |
| 11737643_at | 121275 | OR10AD1 |
| 11737747_a_at | 2764 | GMFB |
| 11737831_a_at | 54874 | FNBP1L |
| 11737866_s_at | 3702 | ITK |

| | | |
|---|---|---|
| 11737893_a_at | 51086 | TNNI3K |
| 11737915_at | 57623 | ZFAT |
| 11738095_x_at | 10922 | FASTK |
| 11738260_at | 26040 | SETBP1 |
| 11738338_at | 140687 | GCNT7 |
| 11738690_s_at | NA | NA |
| 11738741_at | 93408 | MYL10 |
| 11738867_a_at | 54438 | GFOD1 |
| 11739071_a_at | 54749 | EPDR1 |
| 11739122_a_at | 2035 | EPB41 |
| 11739144_a_at | 6502 | SKP2 |
| 11739190_a_at | 10000 | AKT3 |
| 11739222_a_at | 4763 | NF1 |
| 11739300_a_at | 284106 | CISD3 |
| 11739325_a_at | 5771 | PTPN2 |
| 11739340_at | 54855 | FAM46C |
| 11739354_x_at | 55349 | CHDH |
| 11739371_a_at | 1371 | CPOX |
| 11739385_x_at | 9475 | ROCK2 |
| 11739386_at | 79577 | CDC73 |
| 11739405_a_at | 2356 | FPGS |
| 11739414_a_at | 7750 | ZMYM2 |
| 11739434_a_at | 23200 | ATP11B |
| 11739469_at | 84910 | TMEM87B |
| 11739484_a_at | 55704 | CCDC88A |
| 11739492_a_at | 11320 | MGAT4A |
| 11739533_a_at | 10450 | PPIE |
| 11739690_a_at | 80344 | DCAF11 |
| 11739773_a_at | 124460 | SNX20 |
| 11739785_a_at | 6620 | SNCB |
| 11739810_x_at | 23410 | SIRT3 |
| 11739811_a_at | 23410 | SIRT3 |
| 11739904_a_at | 23460 | ABCA6 |
| 11739913_a_at | 8738 | CRADD |
| 11739924_a_at | 7587 | ZNF37A |
| 11739925_x_at | 7587 | ZNF37A |
| 11739949_at | 5208 | PFKFB2 |
| 11739952_a_at | 54438 | GFOD1 |
| 11739964_a_at | 91893 | FDXACB1 |
| 11740125_x_at | 8336 | HIST1H2AM |
| 11740126_a_at | 10114 | HIPK3 |
| 11740131_at | 151050 | KANSL1L |
| 11740199_s_at | 398 | ARHGDIG |
| 11740267_at | 343099 | CCDC18 |
| 11740319_s_at | 3760 | KCNJ3 |
| 11740430_x_at | 23272 | FAM208A |
| 11740499_a_at | 23468 | CBX5 |
| 11740626_a_at | 56995 | TULP4 |
| 11740747_a_at | 1788 | DNMT3A |
| 11740751_a_at | 146542 | ZNF688 |
| 11740828_at | 8340 | HIST1H2BL |
| 11740969_x_at | 23742 | C15orf2 |
| 11741034_x_at | 93611 | FBXO44 |

| | | |
|---|---|---|
| 11741114_x_at | 27250 | PDCD4 |
| 11741192_a_at | 64080 | RBKS |
| 11741295_s_at | NA | NA |
| 11741349_at | 100506736 | SLFN12L |
| 11741365_x_at | 64792 | RABL5 |
| 11741401_a_at | 842 | CASP9 |
| 11741402_a_at | 8209 | C21orf33 |
| 11741407_at | 57713 | SFMBT2 |
| 11741416_s_at | 1977 | EIF4E |
| 11741482_a_at | 1184 | CLCN5 |
| 11741493_a_at | 5830 | PEX5 |
| 11741510_a_at | 9709 | HERPUD1 |
| 11741552_x_at | 653643 | GOLGA6D |
| 11741642_s_at | NA | NA |
| 11741697_a_at | 3704 | ITPA |
| 11741748_a_at | 1739 | DLG1 |
| 11741766_a_at | 116988 | AGAP3 |
| 11741781_a_at | 154790 | CLEC2L |
| 11741784_x_at | 55653 | BCAS4 |
| 11741932_a_at | 283518 | KCNRG |
| 11741948_a_at | 4763 | NF1 |
| 11741960_a_at | 8754 | ADAM9 |
| 11741966_a_at | 84899 | TMTC4 |
| 11742099_x_at | 8763 | CD164 |
| 11742188_a_at | 8671 | SLC4A4 |
| 11742379_x_at | 441282 | AKR1B15 |
| 11742480_a_at | 2639 | GCDH |
| 11742491_at | 390594 | KBTBD13 |
| 11742769_a_at | 23767 | FLRT3 |
| 11742889_at | 26270 | FBXO6 |
| 11742893_at | 10542 | HBXIP |
| 11742925_a_at | 55004 | LAMTOR1 |
| 11743077_s_at | 1051 | CEBPB |
| 11743121_a_at | 5575 | PRKAR1B |
| 11743145_a_at | NA | NA |
| 11743200_a_at | 84071 | ARMC2 |
| 11743281_a_at | 65125 | WNK1 |
| 11743302_a_at | 26053 | AUTS2 |
| 11743462_a_at | 29988 | SLC2A8 |
| 11743474_a_at | 440957 | C3orf78 |
| 11743493_x_at | 3895 | KTN1 |
| 11743505_a_at | 79731 | NARS2 |
| 11743524_a_at | 5927 | KDM5A |
| 11743525_a_at | 5927 | KDM5A |
| 11743629_at | 3667 | IRS1 |
| 11743636_at | 7130 | TNFAIP6 |
| 11743672_at | 23036 | ZNF292 |
| 11743738_a_at | 2908 | NR3C1 |
| 11743762_at | 79753 | SNIP1 |
| 11743764_a_at | 6238 | RRBP1 |
| 11743863_a_at | 490 | ATP2B1 |
| 11743864_a_at | 490 | ATP2B1 |
| 11743923_at | 10732 | TCFL5 |

| | | |
|---|---|---|
| 11744040_a_at | 28958 | CCDC56 |
| 11744044_s_at | 5833 | PCYT2 |
| 11744045_a_at | 29948 | OSGIN1 |
| 11744112_a_at | 84274 | COQ5 |
| 11744113_s_at | 84274 | COQ5 |
| 11744136_at | 199731 | CADM4 |
| 11744171_a_at | 3338 | DNAJC4 |
| 11744172_s_at | 3338 | DNAJC4 |
| 11744173_x_at | 3338 | DNAJC4 |
| 11744205_x_at | 56654 | NPDC1 |
| 11744221_a_at | 55726 | ASUN |
| 11744280_a_at | 10469 | TIMM44 |
| 11744283_a_at | 51117 | COQ4 |
| 11744284_a_at | 6183 | MRPS12 |
| 11744308_a_at | 22996 | TTC39A |
| 11744385_a_at | 283951 | C16orf91 |
| 11744389_a_at | 10139 | ARFRP1 |
| 11744495_a_at | 56912 | IFT46 |
| 11744590_a_at | 7204 | TRIO |
| 11744608_a_at | 10039 | PARP3 |
| 11744615_s_at | 126823 | KLHDC9 |
| 11744625_x_at | 79035 | OBFC2B |
| 11744630_a_at | 91775 | FAM55C |
| 11744667_a_at | 160518 | DENND5B |
| 11744730_s_at | NA | NA |
| 11744872_x_at | 381 | ARF5 |
| 11744930_a_at | 8672 | EIF4G3 |
| 11744940_s_at | NA | NA |
| 11744999_a_at | 10587 | TXNRD2 |
| 11745000_x_at | 10587 | TXNRD2 |
| 11745124_a_at | 622 | BDH1 |
| 11745175_a_at | 23608 | MKRN1 |
| 11745179_x_at | 80344 | DCAF11 |
| 11745210_s_at | 5101 | PCDH9 |
| 11745212_s_at | NA | NA |
| 11745294_a_at | 79676 | OGFOD2 |
| 11745410_a_at | 206358 | SLC36A1 |
| 11745447_x_at | 29901 | SAC3D1 |
| 11745546_a_at | 472 | ATM |
| 11745561_s_at | NA | NA |
| 11745564_x_at | 3338 | DNAJC4 |
| 11745601_a_at | 64167 | ERAP2 |
| 11745652_s_at | 4072 | EPCAM |
| 11745703_a_at | 11123 | RCAN3 |
| 11745706_s_at | 442245 | GSTM2P1 |
| 11745779_a_at | 55904 | MLL5 |
| 11745824_x_at | 30 | ACAA1 |
| 11745858_a_at | 55101 | ATP5SL |
| 11745859_x_at | 55101 | ATP5SL |
| 11745860_x_at | 55101 | ATP5SL |
| 11745863_a_at | 730098 | LOC730098 |
| 11745872_a_at | 10914 | PAPOLA |
| 11746029_a_at | 79970 | ZNF767 |

| | | |
|---|---|---|
| 11746060_a_at | 7127 | TNFAIP2 |
| 11746090_a_at | 9748 | SLK |
| 11746095_a_at | 9360 | PPIG |
| 11746157_a_at | 84964 | ALKBH6 |
| 11746208_a_at | 578 | BAK1 |
| 11746244_a_at | 84818 | IL17RC |
| 11746245_x_at | 84818 | IL17RC |
| 11746295_a_at | 55245 | UQCC |
| 11746417_x_at | 10922 | FASTK |
| 11746422_a_at | 84909 | C9orf3 |
| 11746434_a_at | 5019 | OXCT1 |
| 11746438_a_at | 6472 | SHMT2 |
| 11746441_a_at | 7046 | TGFBR1 |
| 11746488_a_at | 27077 | B9D1 |
| 11746499_a_at | 119391 | GSTO2 |
| 11746500_x_at | 119391 | GSTO2 |
| 11746655_a_at | 30 | ACAA1 |
| 11746668_a_at | 223 | ALDH9A1 |
| 11746717_a_at | 23081 | KDM4C |
| 11746746_a_at | 10142 | AKAP9 |
| 11746751_a_at | NA | NA |
| 11746827_a_at | 54919 | HEATR2 |
| 11746896_a_at | 217 | ALDH2 |
| 11746914_a_at | 27034 | ACAD8 |
| 11746915_x_at | 27034 | ACAD8 |
| 11746945_a_at | 8266 | UBL4A |
| 11746947_a_at | 5786 | PTPRA |
| 11746971_s_at | 4888 | NPY6R |
| 11747004_a_at | 6821 | SUOX |
| 11747011_a_at | 23142 | DCUN1D4 |
| 11747031_a_at | 1760 | DMPK |
| 11747048_a_at | 253832 | ZDHHC20 |
| 11747080_a_at | 80222 | TARS2 |
| 11747093_a_at | 5534 | PPP3R1 |
| 11747142_a_at | 80012 | PHC3 |
| 11747170_x_at | 55798 | METTL2B |
| 11747219_a_at | 51742 | ARID4B |
| 11747253_a_at | 7374 | UNG |
| 11747288_s_at | 84942 | WDR73 |
| 11747313_a_at | 5018 | OXA1L |
| 11747322_s_at | 5833 | PCYT2 |
| 11747332_a_at | 23640 | HSPBP1 |
| 11747375_a_at | 80194 | TMEM134 |
| 11747382_a_at | 26297 | SERGEF |
| 11747385_a_at | 10494 | STK25 |
| 11747408_x_at | 54751 | FBLIM1 |
| 11747456_a_at | 2875 | GPT |
| 11747518_a_at | 7099 | TLR4 |
| 11747535_a_at | 55245 | UQCC |
| 11747536_a_at | 1757 | SARDH |
| 11747537_x_at | 6472 | SHMT2 |
| 11747540_a_at | 8754 | ADAM9 |
| 11747583_s_at | 139728 | PNCK |

| | | |
|---|---|---|
| 11747592_x_at | 79033 | ERI3 |
| 11747609_x_at | 5027 | P2RX7 |
| 11747645_a_at | 6868 | ADAM17 |
| 11747654_a_at | 10110 | SGK2 |
| 11747671_x_at | 1397 | CRIP2 |
| 11747721_s_at | NA | NA |
| 11747870_a_at | 4300 | MLLT3 |
| 11747892_a_at | 84134 | TOMM40L |
| 11747933_a_at | 93100 | NAPRT1 |
| 11747939_a_at | 670 | BPHL |
| 11748011_a_at | 252839 | TMEM9 |
| 11748012_x_at | 252839 | TMEM9 |
| 11748175_x_at | 284325 | C19orf54 |
| 11748203_x_at | 9701 | PPP6R2 |
| 11748288_a_at | 51285 | RASL12 |
| 11748343_a_at | 64792 | RABL5 |
| 11748344_x_at | 64792 | RABL5 |
| 11748370_a_at | 10916 | MAGED2 |
| 11748408_a_at | 51430 | C1orf9 |
| 11748446_a_at | 84818 | IL17RC |
| 11748553_x_at | 55245 | UQCC |
| 11748587_a_at | 4299 | AFF1 |
| 11748595_a_at | 2570 | GABRR2 |
| 11748649_a_at | 54832 | VPS13C |
| 11748726_s_at | 11170 | FAM107A |
| 11748729_s_at | NA | NA |
| 11748770_a_at | 23203 | PMPCA |
| 11748771_a_at | 253943 | YTHDF3 |
| 11748778_a_at | 54953 | C1orf27 |
| 11748782_a_at | 842 | CASP9 |
| 11748784_a_at | 585 | BBS4 |
| 11748821_a_at | 8470 | SORBS2 |
| 11748858_a_at | 29086 | BABAM1 |
| 11748871_a_at | 54988 | ACSM5 |
| 11748904_x_at | 80174 | DBF4B |
| 11748972_a_at | 994 | CDC25B |
| 11748999_a_at | 57097 | PARP11 |
| 11749031_a_at | 9991 | PTBP3 |
| 11749032_x_at | 9991 | PTBP3 |
| 11749046_a_at | 51741 | WWOX |
| 11749130_a_at | 6878 | TAF6 |
| 11749133_a_at | 79671 | NLRX1 |
| 11749183_a_at | 8659 | ALDH4A1 |
| 11749226_x_at | 10385 | BTN2A2 |
| 11749228_a_at | 55754 | TMEM30A |
| 11749251_a_at | 57109 | REXO4 |
| 11749257_a_at | 9709 | HERPUD1 |
| 11749275_a_at | 8935 | SKAP2 |
| 11749285_a_at | 3595 | IL12RB2 |
| 11749299_a_at | 79084 | WDR77 |
| 11749300_x_at | 79084 | WDR77 |
| 11749310_x_at | 58497 | PRUNE |
| 11749311_a_at | 80273 | GRPEL1 |

| | | |
|---|---|---|
| 11749319_a_at | 80055 | PGAP1 |
| 11749334_a_at | 54464 | XRN1 |
| 11749350_x_at | 670 | BPHL |
| 11749366_a_at | 374868 | ATP9B |
| 11749375_a_at | 6843 | VAMP1 |
| 11749390_x_at | 55004 | LAMTOR1 |
| 11749407_a_at | 100128252 | LOC100128252 |
| 11749450_a_at | 1777 | DNASE2 |
| 11749496_a_at | 1523 | CUX1 |
| 11749553_a_at | 3037 | HAS2 |
| 11749634_a_at | 50937 | CDON |
| 11749649_x_at | 3091 | HIF1A |
| 11749681_a_at | 10640 | EXOC5 |
| 11749682_s_at | 10640 | EXOC5 |
| 11749744_a_at | 79718 | TBL1XR1 |
| 11749777_s_at | NA | NA |
| 11749810_s_at | 51477 | ISYNA1 |
| 11749820_a_at | 1140 | CHRNB1 |
| 11749833_s_at | 183 | AGT |
| 11749879_a_at | 27315 | PGAP2 |
| 11749885_a_at | 35 | ACADS |
| 11749925_a_at | 23567 | ZNF346 |
| 11750008_a_at | 5434 | POLR2E |
| 11750113_a_at | 57634 | EP400 |
| 11750132_a_at | 255967 | PAN3 |
| 11750222_a_at | 84159 | ARID5B |
| 11750227_a_at | 55316 | RSAD1 |
| 11750228_x_at | 55316 | RSAD1 |
| 11750239_a_at | 157680 | VPS13B |
| 11750408_a_at | 976 | CD97 |
| 11750441_a_at | 4001 | LMNB1 |
| 11750484_a_at | 4361 | MRE11A |
| 11750561_a_at | 1130 | LYST |
| 11750580_x_at | 23365 | ARHGEF12 |
| 11750666_a_at | 23339 | VPS39 |
| 11750789_a_at | 2060 | EPS15 |
| 11750838_a_at | 93134 | ZNF561 |
| 11750839_x_at | 93134 | ZNF561 |
| 11750932_a_at | 2053 | EPHX2 |
| 11751041_x_at | 55251 | PCMTD2 |
| 11751069_a_at | 112770 | C1orf85 |
| 11751264_a_at | 4122 | MAN2A2 |
| 11751347_a_at | 2332 | FMR1 |
| 11751384_a_at | 23253 | ANKRD12 |
| 11751554_s_at | NA | NA |
| 11751573_a_at | 3930 | LBR |
| 11751625_a_at | 54165 | DCUN1D1 |
| 11751630_a_at | 79885 | HDAC11 |
| 11751667_a_at | 84274 | COQ5 |
| 11751679_a_at | 51285 | RASL12 |
| 11751686_a_at | 79084 | WDR77 |
| 11751787_a_at | 80174 | DBF4B |
| 11751788_x_at | 80174 | DBF4B |

| | | |
|---|---|---|
| 11751828_x_at | 9054 | NFS1 |
| 11751858_a_at | 9616 | RNF7 |
| 11751874_a_at | 472 | ATM |
| 11751902_a_at | 50937 | CDON |
| 11751922_a_at | 4040 | LRP6 |
| 11751967_a_at | NA | NA |
| 11752013_s_at | 80205 | CHD9 |
| 11752023_x_at | 11320 | MGAT4A |
| 11752034_a_at | 10351 | ABCA8 |
| 11752108_a_at | 55291 | PPP6R3 |
| 11752179_s_at | 4077 | NBR1 |
| 11752183_s_at | 9093 | DNAJA3 |
| 11752277_a_at | 85439 | STON2 |
| 11752291_x_at | 55676 | SLC30A6 |
| 11752407_x_at | NA | NA |
| 11752413_a_at | 578 | BAK1 |
| 11752444_a_at | 4763 | NF1 |
| 11752495_a_at | 3275 | PRMT2 |
| 11752512_a_at | NA | NA |
| 11752534_x_at | 50649 | ARHGEF4 |
| 11752612_at | 23555 | TSPAN15 |
| 11752632_a_at | 8243 | SMC1A |
| 11752657_a_at | 1789 | DNMT3B |
| 11752678_a_at | 1620 | DBC1 |
| 11752688_a_at | 6239 | RREB1 |
| 11752738_a_at | 55183 | RIF1 |
| 11752788_a_at | 9686 | VGLL4 |
| 11752790_a_at | 23112 | TNRC6B |
| 11752835_a_at | 84335 | AKT1S1 |
| 11752868_a_at | 23130 | ATG2A |
| 11752873_a_at | 8631 | SKAP1 |
| 11753004_a_at | 1152 | CKB |
| 11753012_s_at | 51031 | GLOD4 |
| 11753061_a_at | 162394 | SLFN5 |
| 11753163_a_at | 79577 | CDC73 |
| 11753301_a_at | 873 | CBR1 |
| 11753335_a_at | 80775 | TMEM177 |
| 11753361_a_at | 132320 | SCLT1 |
| 11753421_a_at | 10370 | CITED2 |
| 11753531_s_at | 126299 | ZNF428 |
| 11753579_a_at | 3572 | IL6ST |
| 11753623_s_at | 5908 | RAP1B |
| 11753625_a_at | 252839 | TMEM9 |
| 11753652_x_at | 2356 | FPGS |
| 11753689_x_at | 4193 | MDM2 |
| 11753820_x_at | 4193 | MDM2 |
| 11753841_x_at | 170622 | COMMD6 |
| 11753871_a_at | 2788 | GNG7 |
| 11753903_a_at | 11257 | TP53TG1 |
| 11753926_x_at | 93436 | ARMC6 |
| 11754024_a_at | 8402 | SLC25A11 |
| 11754055_x_at | 10573 | MRPL28 |
| 11754088_a_at | 79156 | PLEKHF1 |

| | | |
|---|---|---|
| 11754093_x_at | 23640 | HSPBP1 |
| 11754130_a_at | 8815 | BANF1 |
| 11754156_x_at | 381 | ARF5 |
| 11754211_a_at | 9360 | PPIG |
| 11754247_s_at | 546 | ATRX |
| 11754288_a_at | 81562 | LMAN2L |
| 11754289_a_at | 81562 | LMAN2L |
| 11754290_x_at | 81562 | LMAN2L |
| 11754292_a_at | 7260 | TSSC1 |
| 11754306_a_at | 56922 | MCCC1 |
| 11754338_a_at | 55684 | C9orf86 |
| 11754343_a_at | 84895 | FAM73B |
| 11754346_a_at | 79007 | DBNDD1 |
| 11754347_x_at | 79007 | DBNDD1 |
| 11754449_a_at | 201514 | ZNF584 |
| 11754505_a_at | 55630 | SLC39A4 |
| 11754506_x_at | 10922 | FASTK |
| 11754557_s_at | 5578 | PRKCA |
| 11754632_a_at | 199745 | THAP8 |
| 11754640_a_at | 2053 | EPHX2 |
| 11754663_s_at | 50813 | COPS7A |
| 11754673_s_at | NA | NA |
| 11754778_a_at | 128308 | MRPL55 |
| 11754873_a_at | 121274 | ZNF641 |
| 11754890_x_at | 54973 | CPSF3L |
| 11755074_x_at | 255189 | PLA2G4F |
| 11755102_a_at | 83642 | SELO |
| 11755132_s_at | NA | NA |
| 11755200_a_at | 131566 | DCBLD2 |
| 11755220_a_at | 400566 | C17orf97 |
| 11755222_a_at | 10672 | GNA13 |
| 11755240_s_at | 2908 | NR3C1 |
| 11755338_a_at | 9552 | SPAG7 |
| 11755397_a_at | 9734 | HDAC9 |
| 11755406_x_at | 55666 | NPLOC4 |
| 11755481_a_at | 92797 | HELB |
| 11755561_x_at | 5518 | PPP2R1A |
| 11755567_x_at | 1760 | DMPK |
| 11755693_a_at | 143458 | LDLRAD3 |
| 11755762_a_at | 23670 | TMEM2 |
| 11755863_a_at | 3595 | IL12RB2 |
| 11755885_a_at | 26043 | UBXN7 |
| 11755942_x_at | 284325 | C19orf54 |
| 11755946_x_at | 9862 | MED24 |
| 11755953_a_at | 63951 | DMRTA1 |
| 11755956_x_at | 54107 | POLE3 |
| 11755972_a_at | 10724 | MGEA5 |
| 11755987_a_at | 91526 | ANKRD44 |
| 11756025_a_at | 6284 | S100A13 |
| 11756046_x_at | 4193 | MDM2 |
| 11756183_x_at | 8402 | SLC25A11 |
| 11756193_a_at | 131616 | TMEM42 |
| 11756250_s_at | 3155 | HMGCL |

| | | |
|---|---|---|
| 11756299_x_at | 6626 | SNRPA |
| 11756334_x_at | 306 | ANXA3 |
| 11756435_a_at | 112770 | C1orf85 |
| 11756439_a_at | 137872 | ADHFE1 |
| 11756489_a_at | 254552 | NUDT8 |
| 11756506_a_at | 10752 | CHL1 |
| 11756593_a_at | 7389 | UROD |
| 11756634_a_at | 2936 | GSR |
| 11756642_a_at | 26063 | DECR2 |
| 11756665_a_at | 8763 | CD164 |
| 11756669_x_at | 3249 | HPN |
| 11756686_a_at | 84680 | ACCS |
| 11756759_a_at | 26160 | IFT172 |
| 11756796_a_at | 26043 | UBXN7 |
| 11756875_x_at | 170622 | COMMD6 |
| 11756937_s_at | NA | NA |
| 11757095_s_at | NA | NA |
| 11757141_s_at | NA | NA |
| 11757325_a_at | 10519 | CIB1 |
| 11757365_s_at | 84311 | MRPL45 |
| 11757371_x_at | 51477 | ISYNA1 |
| 11757415_s_at | 6526 | SLC5A3 |
| 11757488_a_at | 10248 | POP7 |
| 11757508_x_at | 55101 | ATP5SL |
| 11757542_s_at | 6745 | SSR1 |
| 11757547_a_at | NA | NA |
| 11757577_x_at | 80194 | TMEM134 |
| 11757586_a_at | 10998 | SLC27A5 |
| 11757606_s_at | 1892 | ECHS1 |
| 11757612_a_at | 80150 | ASRGL1 |
| 11757684_a_at | 7165 | TPD52L2 |
| 11757786_s_at | 10640 | EXOC5 |
| 11757822_x_at | 280636 | C11orf31 |
| 11757823_s_at | 23112 | TNRC6B |
| 11757825_a_at | NA | NA |
| 11757876_s_at | 201229 | C17orf108 |
| 11757888_s_at | 54919 | HEATR2 |
| 11757895_s_at | 56995 | TULP4 |
| 11757992_s_at | 57468 | SLC12A5 |
| 11758060_s_at | 84333 | PCGF5 |
| 11758114_x_at | 8935 | SKAP2 |
| 11758175_s_at | 206358 | SLC36A1 |
| 11758270_s_at | 79007 | DBNDD1 |
| 11758366_s_at | 29777 | ABT1 |
| 11758404_s_at | 9774 | BCLAF1 |
| 11758435_s_at | 55239 | OGFOD1 |
| 11758436_s_at | 7095 | SEC62 |
| 11758444_s_at | 55526 | DHTKD1 |
| 11758446_s_at | 8848 | TSC22D1 |
| 11758450_s_at | 55693 | KDM4D |
| 11758517_s_at | 10782 | ZNF274 |
| 11758538_s_at | 4097 | MAFG |
| 11758553_s_at | 1638 | DCT |

| | | |
|---|---|---|
| 11758555_s_at | 1880 | GPR183 |
| 11758559_s_at | NA | NA |
| 11758591_s_at | 55086 | CXorf57 |
| 11758671_s_at | 10320 | IKZF1 |
| 11758775_at | 58477 | SRPRB |
| 11758911_at | 8445 | DYRK2 |
| 11758920_at | 51373 | MRPS17 |
| 11758921_x_at | 51373 | MRPS17 |
| 11758925_at | 55269 | PSPC1 |
| 11758942_s_at | 55233 | MOB1A |
| 11759011_at | 9128 | PRPF4 |
| 11759032_at | 135154 | C6orf57 |
| 11759058_at | 10001 | MED6 |
| 11759080_at | 2936 | GSR |
| 11759149_at | 26273 | FBXO3 |
| 11759252_at | 8329 | HIST1H2AI |
| 11759305_at | 7750 | ZMYM2 |
| 11759339_at | 4915 | NTRK2 |
| 11759404_at | 10098 | TSPAN5 |
| 11759429_a_at | 8609 | KLF7 |
| 11759433_at | 11128 | POLR3A |
| 11759449_x_at | NA | NA |
| 11759450_x_at | NA | NA |
| 11759499_at | 81553 | FAM49A |
| 11759522_a_at | 84266 | ALKBH7 |
| 11759542_s_at | 11252 | PACSIN2 |
| 11759554_at | 10914 | PAPOLA |
| 11759584_a_at | 1327 | COX4I1 |
| 11759585_at | 22931 | RAB18 |
| 11759602_a_at | 57035 | C1orf63 |
| 11759615_x_at | 55119 | PRPF38B |
| 11759619_at | 7109 | TRAPPC10 |
| 11759631_at | 7204 | TRIO |
| 11759673_s_at | 80174 | DBF4B |
| 11759711_a_at | 58517 | RBM25 |
| 11759723_at | 10426 | TUBGCP3 |
| 11759776_at | 79712 | GTDC1 |
| 11759855_s_at | 23042 | PDXDC1 |
| 11759858_at | 4253 | CTAGE5 |
| 11759872_at | 23042 | PDXDC1 |
| 11759895_at | 5101 | PCDH9 |
| 11760012_at | 1794 | DOCK2 |
| 11760056_at | 1462 | VCAN |
| 11760057_at | 57553 | MICAL3 |
| 11760065_at | NA | NA |
| 11760120_s_at | NA | NA |
| 11760125_a_at | 1316 | KLF6 |
| 11760212_at | 9043 | SPAG9 |
| 11760246_at | 8473 | OGT |
| 11760300_at | 25957 | PNISR |
| 11760312_at | 10135 | NAMPT |
| 11760359_at | 5286 | PIK3C2A |
| 11760380_x_at | 23157 | SEPT6 |

| | | |
|---|---|---|
| 11760392_s_at | 10147 | SUGP2 |
| 11760499_at | 26036 | ZNF451 |
| 11760500_x_at | 26036 | ZNF451 |
| 11760508_at | 7444 | VRK2 |
| 11760717_at | 26750 | RPS6KC1 |
| 11760719_at | 23034 | SAMD4A |
| 11760753_a_at | 5156 | PDGFRA |
| 11760853_a_at | 84186 | ZCCHC7 |
| 11760854_x_at | 84186 | ZCCHC7 |
| 11760914_x_at | 9015 | TAF1A |
| 11761106_at | 83605 | CCM2 |
| 11761135_at | 55304 | SPTLC3 |
| 11761251_at | 5373 | PMM2 |
| 11761360_x_at | 6944 | VPS72 |
| 11761451_a_at | 80144 | FRAS1 |
| 11761464_at | 80267 | EDEM3 |
| 11761510_at | 22944 | KIN |
| 11761528_a_at | 151011 | SEPT10 |
| 11761560_x_at | 54482 | CCDC76 |
| 11761596_x_at | 9709 | HERPUD1 |
| 11761675_at | 7404 | UTY |
| 11761715_at | 113201 | CASC4 |
| 11761745_at | 10261 | IGSF6 |
| 11761753_x_at | 25949 | SYF2 |
| 11761781_a_at | 9295 | SRSF11 |
| 11761823_at | 751816 | DSERG1 |
| 11761874_at | 7871 | SLMAP |
| 11761929_at | 84750 | FUT10 |
| 11761956_at | 79963 | ABCA11P |
| 11761958_s_at | 10730 | YME1L1 |
| 11762038_at | 284391 | ZNF844 |
| 11762039_at | 6868 | ADAM17 |
| 11762056_at | 100133299 | LOC100133299 |
| 11762069_a_at | 23157 | SEPT6 |
| 11762131_a_at | 10299 | 03/06/16 |
| 11762151_a_at | 4773 | NFATC2 |
| 11762320_a_at | 51490 | C9orf114 |
| 11762347_at | 9616 | RNF7 |
| 11762358_at | 1106 | CHD2 |
| 11762552_at | NA | NA |
| 11762598_x_at | 79800 | ALS2CR8 |
| 11762609_at | NA | NA |
| 11762761_a_at | NA | NA |
| 11762810_x_at | 841 | CASP8 |
| 11762885_at | NA | NA |
| 11763307_s_at | 4773 | NFATC2 |
| 11763328_a_at | NA | NA |
| 11763335_a_at | 4191 | MDH2 |
| 11763403_a_at | 10658 | CELF1 |
| 11763419_x_at | 55317 | AP5S1 |
| 11763503_a_at | 4763 | NF1 |
| 11763508_s_at | 55778 | ZNF839 |
| 11763560_a_at | 4763 | NF1 |

| | | |
|---|---|---|
| 11763617_a_at | 1303 | COL12A1 |
| 11763627_a_at | 6182 | MRPL12 |
| 11763683_a_at | 51329 | ARL6IP4 |
| 11763730_x_at | 51329 | ARL6IP4 |
| 11763812_at | 7404 | UTY |
| 11763858_a_at | 7404 | UTY |
| 11763859_x_at | 7404 | UTY |
| 11763875_at | 100506797 | LOC100506797 |
| 11763892_at | 692086 | SNORD17 |
| 11763903_at | 677830 | SNORA50 |
| 11763911_at | 692157 | SNORA16B |
| 11763926_x_at | 100124538 | SNORA70C |
| 11763975_a_at | 64963 | MRPS11 |
| 11763990_a_at | 84266 | ALKBH7 |
| 11764001_x_at | 64963 | MRPS11 |
| 11764016_a_at | 51693 | TRAPPC2L |
| 11764145_at | NA | NA |
| 11715113_x_at | 55199 | FAM86C1 |
| 11715475_a_at | 10521 | DDX17 |
| 11715619_x_at | 51150 | SDF4 |
| 11715747_a_at | 4681 | NBL1 |
| 11715826_x_at | 10362 | HMG20B |
| 11715847_x_at | 5327 | PLAT |
| 11715905_x_at | 1654 | DDX3X |
| 11716522_x_at | 11067 | C10orf10 |
| 11716622_x_at | 10263 | CDK2AP2 |
| 11716640_a_at | 5699 | PSMB10 |
| 11716705_at | 54765 | TRIM44 |
| 11716748_at | 4299 | AFF1 |
| 11716781_s_at | 79718 | TBL1XR1 |
| 11717240_at | 5928 | RBBP4 |
| 11717488_s_at | NA | NA |
| 11718574_at | 26135 | SERBP1 |
| 11718671_a_at | 79096 | C11orf49 |
| 11718702_a_at | 27236 | ARFIP1 |
| 11719454_a_at | NA | NA |
| 11719513_a_at | 8751 | ADAM15 |
| 11719734_x_at | 126321 | MFSD12 |
| 11719738_at | 167227 | DCP2 |
| 11719939_a_at | 55832 | CAND1 |
| 11720282_at | 112939 | NACC1 |
| 11720758_s_at | 10625 | IVNS1ABP |
| 11720822_a_at | 435 | ASL |
| 11721088_a_at | 56252 | YLPM1 |
| 11721312_a_at | 475 | ATOX1 |
| 11721321_x_at | 57128 | LYRM4 |
| 11721878_x_at | 4494 | MT1F |
| 11721901_a_at | 57407 | NMRAL1 |
| 11721916_at | 4194 | MDM4 |
| 11722491_a_at | 26470 | SEZ6L2 |
| 11722492_x_at | 26470 | SEZ6L2 |
| 11722583_a_at | 3485 | IGFBP2 |
| 11722710_at | 84733 | CBX2 |

| | | |
|---|---|---|
| 11722717_a_at | 2551 | GABPA |
| 11722914_s_at | 657 | BMPR1A |
| 11723483_at | 54433 | GAR1 |
| 11723557_a_at | 54876 | DCAF16 |
| 11723603_a_at | 122616 | C14orf79 |
| 11724153_at | 60677 | CELF6 |
| 11724173_at | 113675 | SDSL |
| 11724844_s_at | NA | NA |
| 11724967_at | 57478 | USP31 |
| 11725025_s_at | 51477 | ISYNA1 |
| 11726357_a_at | 55904 | MLL5 |
| 11726359_a_at | 55904 | MLL5 |
| 11726829_at | 441250 | TYW1B |
| 11727008_at | 6272 | SORT1 |
| 11727281_a_at | 79148 | MMP28 |
| 11727584_a_at | 169981 | SPIN3 |
| 11727594_a_at | 158358 | KIAA2026 |
| 11729471_x_at | 220382 | FAM181B |
| 11730287_a_at | 394 | ARHGAP5 |
| 11730771_at | 8774 | NAPG |
| 11732668_a_at | NA | NA |
| 11732871_a_at | 158062 | LCN6 |
| 11733125_a_at | 5824 | PEX19 |
| 11733754_a_at | 57664 | PLEKHA4 |
| 11733929_a_at | 25852 | ARMC8 |
| 11733967_at | 7432 | VIP |
| 11734399_a_at | 54832 | VPS13C |
| 11734767_at | 57187 | THOC2 |
| 11735450_at | 348013 | FAM70B |
| 11735535_at | 285349 | ZNF660 |
| 11735706_a_at | 682 | BSG |
| 11736763_x_at | 84671 | ZNF347 |
| 11737663_a_at | 161436 | EML5 |
| 11737990_s_at | NA | NA |
| 11739260_a_at | 286451 | YIPF6 |
| 11739409_s_at | 58508 | MLL3 |
| 11739802_at | 80124 | VCPIP1 |
| 11739875_a_at | 682 | BSG |
| 11740007_at | 10622 | POLR3G |
| 11740260_a_at | 9578 | CDC42BPB |
| 11740679_at | 4985 | OPRD1 |
| 11741033_a_at | 93611 | FBXO44 |
| 11741194_x_at | 53 | ACP2 |
| 11741294_a_at | 84301 | DDI2 |
| 11741395_a_at | 65982 | ZSCAN18 |
| 11741551_x_at | NA | NA |
| 11741657_a_at | 85360 | SYDE1 |
| 11742113_a_at | 27 | ABL2 |
| 11742806_at | 113419 | TEX261 |
| 11743180_at | 4863 | NPAT |
| 11743276_s_at | 10274 | STAG1 |
| 11743867_at | 25983 | NGDN |
| 11743886_at | 342371 | ATXN1L |

| | | |
|---|---|---|
| 11744022_a_at | 146542 | ZNF688 |
| 11744167_a_at | 10072 | DPP3 |
| 11744375_x_at | 54976 | C20orf27 |
| 11744477_x_at | 282974 | STK32C |
| 11744703_x_at | 85360 | SYDE1 |
| 11744924_a_at | 26036 | ZNF451 |
| 11745503_x_at | 706 | TSPO |
| 11746036_s_at | 873 | CBR1 |
| 11746241_s_at | NA | NA |
| 11746473_x_at | 10423 | CDIPT |
| 11746851_a_at | 80011 | FAM192A |
| 11747074_x_at | 81 | ACTN4 |
| 11747099_a_at | 26033 | ATRNL1 |
| 11747359_a_at | 79960 | PHF17 |
| 11747542_a_at | 9526 | MPDU1 |
| 11747543_x_at | 9526 | MPDU1 |
| 11748141_a_at | 1397 | CRIP2 |
| 11748554_a_at | 199745 | THAP8 |
| 11748849_a_at | 578 | BAK1 |
| 11748895_a_at | 134266 | GRPEL2 |
| 11749809_a_at | 5327 | PLAT |
| 11749985_a_at | 4681 | NBL1 |
| 11750237_a_at | 54869 | EPS8L1 |
| 11750280_x_at | 30008 | EFEMP2 |
| 11750506_x_at | 5778 | PTPN7 |
| 11751386_x_at | 6120 | RPE |
| 11751538_a_at | 55109 | AGGF1 |
| 11751615_a_at | 56889 | TM9SF3 |
| 11751900_a_at | 23041 | 04/02/16 |
| 11751901_x_at | 23041 | 04/02/16 |
| 11751972_a_at | 27332 | ZNF638 |
| 11751974_a_at | 4548 | MTR |
| 11752144_a_at | 51111 | SUV420H1 |
| 11752216_a_at | 7450 | VWF |
| 11752350_x_at | 285203 | C3orf64 |
| 11752469_a_at | 54414 | SIAE |
| 11752751_s_at | 284889 | LOC284889 |
| 11752936_a_at | 55131 | RBM28 |
| 11753302_s_at | 873 | CBR1 |
| 11753461_x_at | 93611 | FBXO44 |
| 11753688_a_at | 4193 | MDM2 |
| 11753829_a_at | 5327 | PLAT |
| 11754083_a_at | 682 | BSG |
| 11754283_s_at | 5903 | RANBP2 |
| 11754332_x_at | 85360 | SYDE1 |
| 11754672_a_at | 55743 | CHFR |
| 11754739_a_at | 394 | ARHGAP5 |
| 11755218_a_at | 7694 | ZNF135 |
| 11755339_a_at | 26033 | ATRNL1 |
| 11755456_a_at | 1175 | AP2S1 |
| 11755886_a_at | 65250 | C5orf42 |
| 11755939_a_at | 4059 | BCAM |
| 11756172_x_at | 25771 | TBC1D22A |

| | | | |
|---|---|---|---|
| 11756482_x_at | | 201595 | STT3B |
| 11756644_x_at | | 92714 | ARRDC1 |
| 11756789_a_at | | 26033 | ATRNL1 |
| 11757035_a_at | | 2532 | DARC |
| 11757148_at | | 619505 | SNORA21 |
| 11758471_s_at | | 9825 | SPATA2 |
| 11758726_at | | 509 | ATP5C1 |
| 11759036_at | | 201595 | STT3B |
| 11759411_x_at | NA | | NA |
| 11759883_x_at | NA | | NA |
| 11760639_at | | 26130 | GAPVD1 |
| 11760682_at | | 3300 | DNAJB2 |
| 11761190_x_at | NA | | NA |
| 11762845_a_at | | 400943 | LOC400943 |
| 11763190_x_at | | 59307 | SIGIRR |
| 11763359_at | NA | | NA |
| 11763850_at | | 5901 | RAN |
| 11764223_at | NA | | NA |

genename
taste receptor, type 2, member 30
tRNA phosphotransferase 1
NA
oxidase (cytochrome c) assembly 1-like
homocysteine-inducible, endoplasmic reticulum stress-inducible, ubiquitin-like domain member 1
stromal cell derived factor 4
nipsnap homolog 1 (C. elegans)
potassium channel tetramerisation domain containing 12
NA
mitochondrial ribosomal protein L16
secretory leukocyte peptidase inhibitor
chromosome 21 open reading frame 33
WD repeat domain 77
vacuolar protein sorting 25 homolog (S. cerevisiae)
Cbp/p300-interacting transactivator, with Glu/Asp-rich carboxy-terminal domain, 2
protein phosphatase 1, regulatory subunit 16A
polymerase (RNA) II (DNA directed) polypeptide E, 25kDa
polymerase (RNA) II (DNA directed) polypeptide E, 25kDa
chromobox homolog 5
chromobox homolog 5
chromobox homolog 5
chromobox homolog 5
integrin, alpha V (vitronectin receptor, alpha polypeptide, antigen CD51)
insulin induced gene 1
hairy and enhancer of split 1, (Drosophila)
deoxythymidylate kinase (thymidylate kinase)
p53 and DNA-damage regulated 1
AF4/FMR2 family, member 1
transducin (beta)-like 1 X-linked receptor 1
transducin (beta)-like 1 X-linked receptor 1
chromosome 9 open reading frame 123
peptidylprolyl isomerase E (cyclophilin E)
solute carrier family 35, member A4
KIAA0141
KIAA0141
KIAA0141
signal sequence receptor, alpha
radical S-adenosyl methionine domain containing 1
LSM4 homolog, U6 small nuclear RNA associated (S. cerevisiae)
chromosome 7 open reading frame 50
folylpolyglutamate synthase
cut-like homeobox 1
sorbitol dehydrogenase
pterin-4 alpha-carbinolamine dehydratase/dimerization cofactor of hepatocyte nuclear factor 1 alpha
BCL2-like 12 (proline rich)
septin 5
lanosterol synthase (2,3-oxidosqualene-lanosterol cyclase)
NA
fibronectin type III domain containing 3B
fibronectin type III domain containing 3B
fibronectin type III domain containing 3B
chromosome 20 open reading frame 4

zinc finger, DHHC-type containing 3
zinc finger protein 263
v-Ki-ras2 Kirsten rat sarcoma viral oncogene homolog
mitochondrial ribosomal protein L40
mitochondrial ribosomal protein L4
NFS1 nitrogen fixation 1 homolog (S. cerevisiae)
PTPRF interacting protein, binding protein 2 (liprin beta 2)
microspherule protein 1
BCS1-like (S. cerevisiae)
mitochondrial ribosomal protein L45
ArfGAP with coiled-coil, ankyrin repeat and PH domains 2
protein kinase, AMP-activated, gamma 2 non-catalytic subunit
ATP synthase mitochondrial F1 complex assembly factor 2
ATP synthase mitochondrial F1 complex assembly factor 2
NA
OTU domain containing 4
coenzyme Q9 homolog (S. cerevisiae)
SERTA domain containing 1
SEC24 family, member C (S. cerevisiae)
transmembrane and tetratricopeptide repeat containing 1
thioredoxin reductase 2
ADP-ribosylation factor interacting protein 1
zinc finger protein 655
transmembrane emp24 protein transport domain containing 1
AT rich interactive domain 5B (MRF1-like)
leucyl-tRNA synthetase 2, mitochondrial
frizzled family receptor 4
growth arrest and DNA-damage-inducible, gamma interacting protein 1
TruB pseudouridine (psi) synthase homolog 2 (E. coli)
glutamic pyruvate transaminase (alanine aminotransferase) 2
polymerase (RNA) I polypeptide C, 30kDa
coenzyme Q10 homolog A (S. cerevisiae)
RAS-like, family 12
glutathione S-transferase theta 1
low density lipoprotein receptor-related protein 6
peroxisomal biogenesis factor 11 beta
cryptochrome 2 (photolyase-like)
exocyst complex component 5
SYS1 Golgi-localized integral membrane protein homolog (S. cerevisiae)
RRN3 RNA polymerase I transcription factor homolog (S. cerevisiae)
RRN3 RNA polymerase I transcription factor homolog (S. cerevisiae)
dual specificity phosphatase 22
SLC2A4 regulator
tRNA phosphotransferase 1
pelota homolog (Drosophila)
zinc finger protein 174
ataxia telangiectasia mutated
DnaJ (Hsp40) homolog, subfamily C, member 10
methionyl aminopeptidase 1
nudix (nucleoside diphosphate linked moiety X)-type motif 22
protein S (alpha)
DEAD (Asp-Glu-Ala-Asp) box polypeptide 28
NADH dehydrogenase (ubiquinone) 1 alpha subcomplex, 7, 14.5kDa

glutamine--fructose-6-phosphate transaminase 1
RNA methyltransferase like 1
ferredoxin 1-like
protein phosphatase 6, regulatory subunit 2
SRY (sex determining region Y)-box 9
thiosulfate sulfurtransferase (rhodanese)-like domain containing 1
phosphatidylethanolamine binding protein 1
prenyl (decaprenyl) diphosphate synthase, subunit 1
chromosome 19 open reading frame 25
mitochondrial ribosomal protein S27
mitochondrial ribosomal protein S27
isovaleryl-CoA dehydrogenase
isovaleryl-CoA dehydrogenase
chromosome 16 open reading frame 42
chromosome 1 open reading frame 54
kinesin family member 1B
carnitine O-octanoyltransferase
peptidylprolyl isomerase F
mitotic spindle organizing protein 2B
GRAM domain containing 1C
acetyl-CoA acyltransferase 1
SPARC related modular calcium binding 1
family with sequence similarity 107, member A
exoribonuclease 1
thioredoxin-related transmembrane protein 1
nth endonuclease III-like 1 (E. coli)
protein phosphatase 1, regulatory subunit 26
transmembrane protein 106B
microfibrillar-associated protein 1
WW domain containing oxidoreductase
pleckstrin homology-like domain, family A, member 1
serine/arginine-rich splicing factor 10
glycine C-acetyltransferase
zinc finger, DHHC-type containing 17
TAF6 RNA polymerase II, TATA box binding protein (TBP)-associated factor, 80kDa
dehydrogenase E1 and transketolase domain containing 1
GLI pathogenesis-related 1
coiled-coil domain containing 28B
Mdm4 p53 binding protein homolog (mouse)
Mdm4 p53 binding protein homolog (mouse)
SUMO1/sentrin specific peptidase 5
cytochrome P450, family 3, subfamily A, polypeptide 5
ring finger and CCCH-type domains 2
ubiquitin protein ligase E3C
prenyl (decaprenyl) diphosphate synthase, subunit 2
receptor accessory protein 3
nuclear receptor subfamily 1, group D, member 2
pecanex homolog (Drosophila)
sphingomyelin synthase 2
ubiquinol-cytochrome c reductase complex chaperone
ubiquinol-cytochrome c reductase complex chaperone
N-myristoyltransferase 1
polyhomeotic homolog 3 (Drosophila)

polyhomeotic homolog 3 (Drosophila)
golgi-associated, gamma adaptin ear containing, ARF binding protein 2
fibrinogen-like 2
sirtuin 3
sirtuin 3
cyclin A2
charged multivesicular body protein 4A
pleckstrin homology domain containing, family F (with FYVE domain) member 1
serine/threonine kinase 17a
spectrin, beta, non-erythrocytic 1
KIAA1109
hook homolog 3 (Drosophila)
hook homolog 3 (Drosophila)
WD repeat domain 62
chromosome 12 open reading frame 23
Sp1 transcription factor
NA
baculoviral IAP repeat containing 3
coiled-coil domain containing 76
U-box domain containing 5
aldehyde dehydrogenase 6 family, member A1
HemK methyltransferase family member 1
NA
chromosome X open reading frame 40A
sodium leak channel, non-selective
chromosome 7 open reading frame 10
NA
guanine nucleotide binding protein (G protein), gamma 7
guanine nucleotide binding protein (G protein), gamma 7
peroxisomal membrane protein 2, 22kDa
COP9 constitutive photomorphogenic homolog subunit 7A (Arabidopsis)
zinc finger protein 609
zinc finger protein 609
zinc finger protein 385A
ATG2 autophagy related 2 homolog A (S. cerevisiae)
NFKB inhibitor interacting Ras-like 2
ubiquitin specific peptidase 34
RNA pseudouridylate synthase domain containing 2
NA
synuclein, beta
oxysterol binding protein-like 11
PDZ domain containing 4
tripartite motif containing 68
glycolipid transfer protein domain containing 1
solute carrier family 25 (mitochondrial thiamine pyrophosphate carrier), member 19
kelch domain containing 9
zinc finger protein 853
family with sequence similarity 65, member C
SPRY domain containing 4
transcription factor 4
triple functional domain (PTPRF interacting)
RAB, member RAS oncogene family-like 5
annexin A3

solute carrier family 35, member F1
solute carrier family 35, member F1
transmembrane protein 177
diphosphoinositol pentakisphosphate kinase 1
fumarylacetoacetate hydrolase domain containing 1
fumarylacetoacetate hydrolase domain containing 1
ras homolog family member F (in filopodia)
endothelin converting enzyme 2
ankyrin repeat domain 35
hepsin
platelet-activating factor acetylhydrolase 2, 40kDa
Rho GTPase activating protein 44
limb region 1 homolog (mouse)
UDP-N-acetyl-alpha-D-galactosamine:polypeptide N-acetylgalactosaminyltransferase 12 (GalNAc-T12)
UDP-N-acetyl-alpha-D-galactosamine:polypeptide N-acetylgalactosaminyltransferase 12 (GalNAc-T12)
heat shock 70kDa protein 2
mitogen-activated protein kinase kinase kinase kinase 5
chromosome 10 open reading frame 76
cytoplasmic polyadenylation element binding protein 2
5',3'-nucleotidase, mitochondrial
septin 6
neurolysin (metallopeptidase M3 family)
zinc finger protein 784
cytochrome P450, family 39, subfamily A, polypeptide 1
ficolin (collagen/fibrinogen domain containing) 3 (Hakata antigen)
RNA binding protein with multiple splicing 2
aggrecan
mesoderm posterior 1 homolog (mouse)
calcium channel, voltage-dependent, T type, alpha 1H subunit
mitochondrial ribosomal protein S26
mitochondrial rRNA methyltransferase 1 homolog (S. cerevisiae)
MRE11 meiotic recombination 11 homolog A (S. cerevisiae)
zinc finger and SCAN domain containing 16
Ras association (RalGDS/AF-6) domain family member 5
mitochondrial ribosomal protein L11
endonuclease/exonuclease/phosphatase family domain containing 1
Rho guanine nucleotide exchange factor (GEF) 7
chromosome 12 open reading frame 65
family with sequence similarity 46, member B
plakophilin 2
synapsin I
isocitrate dehydrogenase 3 (NAD+) gamma
acyl-CoA oxidase 1, palmitoyl
DCN1, defective in cullin neddylation 1, domain containing 1 (S. cerevisiae)
nudix (nucleoside diphosphate linked moiety X)-type motif 14
ataxin 3
oxysterol binding protein-like 3
oxysterol binding protein-like 3
threonyl-tRNA synthetase 2, mitochondrial (putative)
tripartite motif containing 8
mindbomb E3 ubiquitin protein ligase 1
MAP6 domain containing 1
cell division cycle 25 homolog A (S. pombe)

lactate dehydrogenase D
breast carcinoma amplified sequence 4
breast carcinoma amplified sequence 4
PTEN induced putative kinase 1
poliovirus receptor-related 3
cholinergic receptor, nicotinic, beta 1 (muscle)
cholinergic receptor, nicotinic, beta 1 (muscle)
NA
glutathione transferase zeta 1
crystallin, lambda 1
ATP5S-like
ATP5S-like
ZXD family zinc finger C
branched chain amino-acid transaminase 2, mitochondrial
kelch-like 6 (Drosophila)
KIAA2026
zinc finger protein 329
methyltransferase like 21C
guanine nucleotide binding protein (G protein), alpha inhibiting activity polypeptide 3
serine/arginine-rich splicing factor 1
NA
formyl peptide receptor 3
Ral GEF with PH domain and SH3 binding motif 2
tripartite motif containing 66
SFT2 domain containing 2
SERTA domain containing 3
KH homology domain containing 1
HEAT repeat containing 2
sirtuin 4
Ts translation elongation factor, mitochondrial
SH2 domain containing 1A
zinc finger protein 148
osteomodulin
coiled-coil-helix-coiled-coil-helix domain containing 5
ligand dependent nuclear receptor corepressor
bromodomain and WD repeat domain containing 1
BTB (POZ) domain containing 11
mutated in colorectal cancers
mutated in colorectal cancers
ATPase, H+ transporting, lysosomal 31kDa, V1 subunit E2
adaptor-related protein complex 4, epsilon 1 subunit
kinesin family member 22
makorin ring finger protein 1
adrenergic, beta-2-, receptor, surface
TMF1-regulated nuclear protein 1
protein phosphatase 3, regulatory subunit B, alpha
beta 1,3-galactosyltransferase-like
gasdermin C
fumarylacetoacetate hydrolase domain containing 2B
potassium voltage-gated channel, shaker-related subfamily, member 5
inositol 1,4,5-trisphosphate receptor interacting protein-like 2
MYC associated factor X
isochorismatase domain containing 2

leucyl/cystinyl aminopeptidase
zinc finger and BTB domain containing 16
HECT domain containing E3 ubiquitin protein ligase 2
zinc finger protein 285
zinc finger protein 285
dopachrome tautomerase (dopachrome delta-isomerase, tyrosine-related protein 2)
HD domain containing 3
SMAD family member 2
nucleolar protein 3 (apoptosis repressor with CARD domain)
3-hydroxybutyrate dehydrogenase, type 1
transmembrane protein 231
zinc finger protein 561
zinc finger and BTB domain containing 6
family with sequence similarity 110, member B
IBA57, iron-sulfur cluster assembly homolog (S. cerevisiae)
IBA57, iron-sulfur cluster assembly homolog (S. cerevisiae)
transmembrane protein 134
zinc finger protein 135
zinc finger protein 135
zinc finger and BTB domain containing 37
poly (ADP-ribose) polymerase family, member 11
neuronal differentiation 2
tripartite motif containing 37
mitochondrial ribosomal protein S7
mitochondrial ribosomal protein S28
zinc finger protein 341
potassium inwardly-rectifying channel, subfamily J, member 2
potassium inwardly-rectifying channel, subfamily J, member 2
ATP-binding cassette, sub-family A (ABC1), member 9
acetyl-CoA acyltransferase 1
endogenous Bornavirus-like nucleoprotein 2
inositol hexakisphosphate kinase 3
endothelin converting enzyme-like 1
deoxyribonuclease II, lysosomal
deoxyribonuclease II, lysosomal
mitochondrial ribosomal protein L4
nudix (nucleoside diphosphate linked moiety X)-type motif 16
NOP2/Sun domain family, member 6
acyl-CoA synthetase short-chain family member 3
mucin-like 1
ataxin 7
ataxin 7
zinc finger protein 516
D-dopachrome tautomerase
RNA pseudouridylate synthase domain containing 3
inositol hexakisphosphate kinase 3
cathepsin C
cathepsin C
secreted frizzled-related protein 4
Mdm2, p53 E3 ubiquitin protein ligase homolog (mouse)
Mdm2, p53 E3 ubiquitin protein ligase homolog (mouse)
coiled-coil domain containing 106
cannabinoid receptor 1 (brain)

RAP1 interacting factor homolog (yeast)
NA
C-type lectin domain family 2, member D
ring finger protein 213
ring finger protein 213
F-box protein 11
potassium channel regulator
outer dense fiber of sperm tails 3-like 2
tripartite motif containing 58
fibronectin type III and SPRY domain containing 1-like
translocase of inner mitochondrial membrane 13 homolog (yeast)
DDB1 and CUL4 associated factor 11
DDB1 and CUL4 associated factor 11
DDB1 and CUL4 associated factor 11
ADAM metallopeptidase with thrombospondin type 1 motif, 1
phosphatidylinositol 4-kinase type 2 alpha
peroxisomal biogenesis factor 10
KIAA1715
KIAA1715
KIAA1715
ankyrin repeat domain 13C
hepsin
BCL2-antagonist/killer 1
homeobox A7
microtubule associated monoxygenase, calponin and LIM domain containing 3
ring finger protein 213
polypyrimidine tract binding protein 3
polypyrimidine tract binding protein 3
FAD-dependent oxidoreductase domain containing 2
ubiquitin-like 7 (bone marrow stromal cell-derived)
N-acetyltransferase 8-like (GCN5-related, putative)
sushi domain containing 5
zinc finger protein 704
COMM domain containing 6
COMM domain containing 6
pregnancy-zone protein
sema domain, transmembrane domain (TM), and cytoplasmic domain, (semaphorin) 6D
branched chain keto acid dehydrogenase E1, alpha polypeptide
transmembrane protein 9
structural maintenance of chromosomes 4
nuclear protein localization 4 homolog (S. cerevisiae)
pyruvate dehydrogenase kinase, isozyme 1
transmembrane protein 150C
heparan sulfate (glucosamine) 3-O-sulfotransferase 3B1
NA
NA
ADP-ribosylation factor GTPase activating protein 2
FGGY carbohydrate kinase domain containing
nucleotide-binding oligomerization domain containing 1
transcription factor EC
zinc finger protein 844
ATG10 autophagy related 10 homolog (S. cerevisiae)
G protein-coupled receptor 18

cell adhesion molecule 2
phospholipase A2, group IVF
fatty acyl CoA reductase 1
proteoglycan 4
muscleblind-like splicing regulator 3
sortilin-related VPS10 domain containing receptor 1
sortilin-related VPS10 domain containing receptor 1
neighbor of BRCA1 gene 1
tolloid-like 2
replication factor C (activator 1) 2, 40kDa
collagen, type X, alpha 1
leucine rich repeat transmembrane neuronal 4
NLR family, CARD domain containing 4
vacuolar protein sorting 13 homolog B (yeast)
retinoic acid receptor responder (tazarotene induced) 1
oxidative stress induced growth inhibitor 1
ADP-ribosylation factor 5
zinc finger protein 385A
NA
coiled-coil domain containing 144A
regulatory subunit of type II PKA R-subunit (RIIa) domain containing 1
NA
ectodysplasin A
wingless-type MMTV integration site family, member 9A
chromosome 10 open reading frame 32
discoidin, CUB and LCCL domain containing 2
deltex homolog 3 (Drosophila)
DNA (cytosine-5-)-methyltransferase 1
transcription factor 7 (T-cell specific, HMG-box)
transcription factor 7 (T-cell specific, HMG-box)
trinucleotide repeat containing 6B
trinucleotide repeat containing 6B
v-rel reticuloendotheliosis viral oncogene homolog (avian)
protein tyrosine phosphatase, non-receptor type 14
glucosaminyl (N-acetyl) transferase 1, core 2
dpy-19-like 3 (C. elegans)
GIPC PDZ domain containing family, member 2
histone deacetylase 9
fibronectin type III and SPRY domain containing 1-like
zinc finger protein 431
zinc finger protein 385A
zinc finger protein 805
ras responsive element binding protein 1
ras responsive element binding protein 1
zinc finger protein 763
adrenergic, alpha-2B-, receptor
putative homeodomain transcription factor 2
Myb-like, SWIRM and MPN domains 1
zinc finger, DHHC-type containing 21
olfactory receptor, family 10, subfamily AD, member 1
glia maturation factor, beta
formin binding protein 1-like
IL2-inducible T-cell kinase

TNNI3 interacting kinase
zinc finger and AT hook domain containing
Fas-activated serine/threonine kinase
SET binding protein 1
glucosaminyl (N-acetyl) transferase family member 7
NA
myosin, light chain 10, regulatory
glucose-fructose oxidoreductase domain containing 1
ependymin related protein 1 (zebrafish)
erythrocyte membrane protein band 4.1 (elliptocytosis 1, RH-linked)
S-phase kinase-associated protein 2, E3 ubiquitin protein ligase
v-akt murine thymoma viral oncogene homolog 3 (protein kinase B, gamma)
neurofibromin 1
CDGSH iron sulfur domain 3
protein tyrosine phosphatase, non-receptor type 2
family with sequence similarity 46, member C
choline dehydrogenase
coproporphyrinogen oxidase
Rho-associated, coiled-coil containing protein kinase 2
cell division cycle 73, Paf1/RNA polymerase II complex component, homolog (S. cerevisiae)
folylpolyglutamate synthase
zinc finger, MYM-type 2
ATPase, class VI, type 11B
transmembrane protein 87B
coiled-coil domain containing 88A
mannosyl (alpha-1,3-)-glycoprotein beta-1,4-N-acetylglucosaminyltransferase, isozyme A
peptidylprolyl isomerase E (cyclophilin E)
DDB1 and CUL4 associated factor 11
sorting nexin 20
synuclein, beta
sirtuin 3
sirtuin 3
ATP-binding cassette, sub-family A (ABC1), member 6
CASP2 and RIPK1 domain containing adaptor with death domain
zinc finger protein 37A
zinc finger protein 37A
6-phosphofructo-2-kinase/fructose-2,6-biphosphatase 2
glucose-fructose oxidoreductase domain containing 1
ferredoxin-fold anticodon binding domain containing 1
histone cluster 1, H2am
homeodomain interacting protein kinase 3
KAT8 regulatory NSL complex subunit 1-like
Rho GDP dissociation inhibitor (GDI) gamma
coiled-coil domain containing 18
potassium inwardly-rectifying channel, subfamily J, member 3
family with sequence similarity 208, member A
chromobox homolog 5
tubby like protein 4
DNA (cytosine-5-)-methyltransferase 3 alpha
zinc finger protein 688
histone cluster 1, H2bl
chromosome 15 open reading frame 2
F-box protein 44

programmed cell death 4 (neoplastic transformation inhibitor)
ribokinase
NA
schlafen family member 12-like
RAB, member RAS oncogene family-like 5
caspase 9, apoptosis-related cysteine peptidase
chromosome 21 open reading frame 33
Scm-like with four mbt domains 2
eukaryotic translation initiation factor 4E
chloride channel, voltage-sensitive 5
peroxisomal biogenesis factor 5
homocysteine-inducible, endoplasmic reticulum stress-inducible, ubiquitin-like domain member 1
golgin A6 family, member D
NA
inosine triphosphatase (nucleoside triphosphate pyrophosphatase)
discs, large homolog 1 (Drosophila)
ArfGAP with GTPase domain, ankyrin repeat and PH domain 3
C-type lectin domain family 2, member L
breast carcinoma amplified sequence 4
potassium channel regulator
neurofibromin 1
ADAM metallopeptidase domain 9
transmembrane and tetratricopeptide repeat containing 4
CD164 molecule, sialomucin
solute carrier family 4, sodium bicarbonate cotransporter, member 4
aldo-keto reductase family 1, member B15
glutaryl-CoA dehydrogenase
kelch repeat and BTB (POZ) domain containing 13
fibronectin leucine rich transmembrane protein 3
F-box protein 6
hepatitis B virus x interacting protein
late endosomal/lysosomal adaptor, MAPK and MTOR activator 1
CCAAT/enhancer binding protein (C/EBP), beta
protein kinase, cAMP-dependent, regulatory, type I, beta
NA
armadillo repeat containing 2
WNK lysine deficient protein kinase 1
autism susceptibility candidate 2
solute carrier family 2 (facilitated glucose transporter), member 8
chromosome 3 open reading frame 78
kinectin 1 (kinesin receptor)
asparaginyl-tRNA synthetase 2, mitochondrial (putative)
lysine (K)-specific demethylase 5A
lysine (K)-specific demethylase 5A
insulin receptor substrate 1
tumor necrosis factor, alpha-induced protein 6
zinc finger protein 292
nuclear receptor subfamily 3, group C, member 1 (glucocorticoid receptor)
Smad nuclear interacting protein 1
ribosome binding protein 1 homolog 180kDa (dog)
ATPase, Ca++ transporting, plasma membrane 1
ATPase, Ca++ transporting, plasma membrane 1
transcription factor-like 5 (basic helix-loop-helix)

coiled-coil domain containing 56
phosphate cytidylyltransferase 2, ethanolamine
oxidative stress induced growth inhibitor 1
coenzyme Q5 homolog, methyltransferase (S. cerevisiae)
coenzyme Q5 homolog, methyltransferase (S. cerevisiae)
cell adhesion molecule 4
DnaJ (Hsp40) homolog, subfamily C, member 4
DnaJ (Hsp40) homolog, subfamily C, member 4
DnaJ (Hsp40) homolog, subfamily C, member 4
neural proliferation, differentiation and control, 1
asunder, spermatogenesis regulator homolog (Drosphila)
translocase of inner mitochondrial membrane 44 homolog (yeast)
coenzyme Q4 homolog (S. cerevisiae)
mitochondrial ribosomal protein S12
tetratricopeptide repeat domain 39A
chromosome 16 open reading frame 91
ADP-ribosylation factor related protein 1
intraflagellar transport 46 homolog (Chlamydomonas)
triple functional domain (PTPRF interacting)
poly (ADP-ribose) polymerase family, member 3
kelch domain containing 9
oligonucleotide/oligosaccharide-binding fold containing 2B
family with sequence similarity 55, member C
DENN/MADD domain containing 5B
NA
ADP-ribosylation factor 5
eukaryotic translation initiation factor 4 gamma, 3
NA
thioredoxin reductase 2
thioredoxin reductase 2
3-hydroxybutyrate dehydrogenase, type 1
makorin ring finger protein 1
DDB1 and CUL4 associated factor 11
protocadherin 9
NA
2-oxoglutarate and iron-dependent oxygenase domain containing 2
solute carrier family 36 (proton/amino acid symporter), member 1
SAC3 domain containing 1
ataxia telangiectasia mutated
NA
DnaJ (Hsp40) homolog, subfamily C, member 4
endoplasmic reticulum aminopeptidase 2
epithelial cell adhesion molecule
RCAN family member 3
glutathione S-transferase mu 2 (muscle) pseudogene 1
myeloid/lymphoid or mixed-lineage leukemia 5 (trithorax homolog, Drosophila)
acetyl-CoA acyltransferase 1
ATP5S-like
ATP5S-like
ATP5S-like
uncharacterized LOC730098
poly(A) polymerase alpha
zinc finger family member 767

tumor necrosis factor, alpha-induced protein 2
STE20-like kinase
peptidylprolyl isomerase G (cyclophilin G)
alkB, alkylation repair homolog 6 (E. coli)
BCL2-antagonist/killer 1
interleukin 17 receptor C
interleukin 17 receptor C
ubiquinol-cytochrome c reductase complex chaperone
Fas-activated serine/threonine kinase
chromosome 9 open reading frame 3
3-oxoacid CoA transferase 1
serine hydroxymethyltransferase 2 (mitochondrial)
transforming growth factor, beta receptor 1
B9 protein domain 1
glutathione S-transferase omega 2
glutathione S-transferase omega 2
acetyl-CoA acyltransferase 1
aldehyde dehydrogenase 9 family, member A1
lysine (K)-specific demethylase 4C
A kinase (PRKA) anchor protein (yotiao) 9
NA
HEAT repeat containing 2
aldehyde dehydrogenase 2 family (mitochondrial)
acyl-CoA dehydrogenase family, member 8
acyl-CoA dehydrogenase family, member 8
ubiquitin-like 4A
protein tyrosine phosphatase, receptor type, A
neuropeptide Y receptor Y6 (pseudogene)
sulfite oxidase
DCN1, defective in cullin neddylation 1, domain containing 4 (S. cerevisiae)
dystrophia myotonica-protein kinase
zinc finger, DHHC-type containing 20
threonyl-tRNA synthetase 2, mitochondrial (putative)
protein phosphatase 3, regulatory subunit B, alpha
polyhomeotic homolog 3 (Drosophila)
methyltransferase like 2B
AT rich interactive domain 4B (RBP1-like)
uracil-DNA glycosylase
WD repeat domain 73
oxidase (cytochrome c) assembly 1-like
phosphate cytidylyltransferase 2, ethanolamine
HSPA (heat shock 70kDa) binding protein, cytoplasmic cochaperone 1
transmembrane protein 134
secretion regulating guanine nucleotide exchange factor
serine/threonine kinase 25
filamin binding LIM protein 1
glutamic-pyruvate transaminase (alanine aminotransferase)
toll-like receptor 4
ubiquinol-cytochrome c reductase complex chaperone
sarcosine dehydrogenase
serine hydroxymethyltransferase 2 (mitochondrial)
ADAM metallopeptidase domain 9
pregnancy up-regulated non-ubiquitously expressed CaM kinase

ERI1 exoribonuclease family member 3
purinergic receptor P2X, ligand-gated ion channel, 7
ADAM metallopeptidase domain 17
serum/glucocorticoid regulated kinase 2
cysteine-rich protein 2
NA
myeloid/lymphoid or mixed-lineage leukemia (trithorax homolog, Drosophila); translocated to, 3
translocase of outer mitochondrial membrane 40 homolog (yeast)-like
nicotinate phosphoribosyltransferase domain containing 1
biphenyl hydrolase-like (serine hydrolase)
transmembrane protein 9
transmembrane protein 9
chromosome 19 open reading frame 54
protein phosphatase 6, regulatory subunit 2
RAS-like, family 12
RAB, member RAS oncogene family-like 5
RAB, member RAS oncogene family-like 5
melanoma antigen family D, 2
chromosome 1 open reading frame 9
interleukin 17 receptor C
ubiquinol-cytochrome c reductase complex chaperone
AF4/FMR2 family, member 1
gamma-aminobutyric acid (GABA) A receptor, rho 2
vacuolar protein sorting 13 homolog C (S. cerevisiae)
family with sequence similarity 107, member A
NA
peptidase (mitochondrial processing) alpha
YTH domain family, member 3
chromosome 1 open reading frame 27
caspase 9, apoptosis-related cysteine peptidase
Bardet-Biedl syndrome 4
sorbin and SH3 domain containing 2
BRISC and BRCA1 A complex member 1
acyl-CoA synthetase medium-chain family member 5
DBF4 homolog B (S. cerevisiae)
cell division cycle 25 homolog B (S. pombe)
poly (ADP-ribose) polymerase family, member 11
polypyrimidine tract binding protein 3
polypyrimidine tract binding protein 3
WW domain containing oxidoreductase
TAF6 RNA polymerase II, TATA box binding protein (TBP)-associated factor, 80kDa
NLR family member X1
aldehyde dehydrogenase 4 family, member A1
butyrophilin, subfamily 2, member A2
transmembrane protein 30A
REX4, RNA exonuclease 4 homolog (S. cerevisiae)
homocysteine-inducible, endoplasmic reticulum stress-inducible, ubiquitin-like domain member 1
src kinase associated phosphoprotein 2
interleukin 12 receptor, beta 2
WD repeat domain 77
WD repeat domain 77
prune homolog (Drosophila)
GrpE-like 1, mitochondrial (E. coli)

post-GPI attachment to proteins 1
5'-3' exoribonuclease 1
biphenyl hydrolase-like (serine hydrolase)
ATPase, class II, type 9B
vesicle-associated membrane protein 1 (synaptobrevin 1)
late endosomal/lysosomal adaptor, MAPK and MTOR activator 1
uncharacterized LOC100128252
deoxyribonuclease II, lysosomal
cut-like homeobox 1
hyaluronan synthase 2
Cdon homolog (mouse)
hypoxia inducible factor 1, alpha subunit (basic helix-loop-helix transcription factor)
exocyst complex component 5
exocyst complex component 5
transducin (beta)-like 1 X-linked receptor 1
NA
inositol-3-phosphate synthase 1
cholinergic receptor, nicotinic, beta 1 (muscle)
angiotensinogen (serpin peptidase inhibitor, clade A, member 8)
post-GPI attachment to proteins 2
acyl-CoA dehydrogenase, C-2 to C-3 short chain
zinc finger protein 346
polymerase (RNA) II (DNA directed) polypeptide E, 25kDa
E1A binding protein p400
PAN3 poly(A) specific ribonuclease subunit homolog (S. cerevisiae)
AT rich interactive domain 5B (MRF1-like)
radical S-adenosyl methionine domain containing 1
radical S-adenosyl methionine domain containing 1
vacuolar protein sorting 13 homolog B (yeast)
CD97 molecule
lamin B1
MRE11 meiotic recombination 11 homolog A (S. cerevisiae)
lysosomal trafficking regulator
Rho guanine nucleotide exchange factor (GEF) 12
vacuolar protein sorting 39 homolog (S. cerevisiae)
epidermal growth factor receptor pathway substrate 15
zinc finger protein 561
zinc finger protein 561
epoxide hydrolase 2, cytoplasmic
protein-L-isoaspartate (D-aspartate) O-methyltransferase domain containing 2
chromosome 1 open reading frame 85
mannosidase, alpha, class 2A, member 2
fragile X mental retardation 1
ankyrin repeat domain 12
NA
lamin B receptor
DCN1, defective in cullin neddylation 1, domain containing 1 (S. cerevisiae)
histone deacetylase 11
coenzyme Q5 homolog, methyltransferase (S. cerevisiae)
RAS-like, family 12
WD repeat domain 77
DBF4 homolog B (S. cerevisiae)
DBF4 homolog B (S. cerevisiae)

NFS1 nitrogen fixation 1 homolog (S. cerevisiae)
ring finger protein 7
ataxia telangiectasia mutated
Cdon homolog (mouse)
low density lipoprotein receptor-related protein 6
NA
chromodomain helicase DNA binding protein 9
mannosyl (alpha-1,3-)-glycoprotein beta-1,4-N-acetylglucosaminyltransferase, isozyme A
ATP-binding cassette, sub-family A (ABC1), member 8
protein phosphatase 6, regulatory subunit 3
neighbor of BRCA1 gene 1
DnaJ (Hsp40) homolog, subfamily A, member 3
stonin 2
solute carrier family 30 (zinc transporter), member 6
NA
BCL2-antagonist/killer 1
neurofibromin 1
protein arginine methyltransferase 2
NA
Rho guanine nucleotide exchange factor (GEF) 4
tetraspanin 15
structural maintenance of chromosomes 1A
DNA (cytosine-5-)-methyltransferase 3 beta
deleted in bladder cancer 1
ras responsive element binding protein 1
RAP1 interacting factor homolog (yeast)
vestigial like 4 (Drosophila)
trinucleotide repeat containing 6B
AKT1 substrate 1 (proline-rich)
ATG2 autophagy related 2 homolog A (S. cerevisiae)
src kinase associated phosphoprotein 1
creatine kinase, brain
glyoxalase domain containing 4
schlafen family member 5
cell division cycle 73, Paf1/RNA polymerase II complex component, homolog (S. cerevisiae)
carbonyl reductase 1
transmembrane protein 177
sodium channel and clathrin linker 1
Cbp/p300-interacting transactivator, with Glu/Asp-rich carboxy-terminal domain, 2
zinc finger protein 428
interleukin 6 signal transducer (gp130, oncostatin M receptor)
RAP1B, member of RAS oncogene family
transmembrane protein 9
folylpolyglutamate synthase
Mdm2, p53 E3 ubiquitin protein ligase homolog (mouse)
Mdm2, p53 E3 ubiquitin protein ligase homolog (mouse)
COMM domain containing 6
guanine nucleotide binding protein (G protein), gamma 7
TP53 target 1 (non-protein coding)
armadillo repeat containing 6
solute carrier family 25 (mitochondrial carrier; oxoglutarate carrier), member 11
mitochondrial ribosomal protein L28
pleckstrin homology domain containing, family F (with FYVE domain) member 1

HSPA (heat shock 70kDa) binding protein, cytoplasmic cochaperone 1
barrier to autointegration factor 1
ADP-ribosylation factor 5
peptidylprolyl isomerase G (cyclophilin G)
alpha thalassemia/mental retardation syndrome X-linked
lectin, mannose-binding 2-like
lectin, mannose-binding 2-like
lectin, mannose-binding 2-like
tumor suppressing subtransferable candidate 1
methylcrotonoyl-CoA carboxylase 1 (alpha)
chromosome 9 open reading frame 86
family with sequence similarity 73, member B
dysbindin (dystrobrevin binding protein 1) domain containing 1
dysbindin (dystrobrevin binding protein 1) domain containing 1
zinc finger protein 584
solute carrier family 39 (zinc transporter), member 4
Fas-activated serine/threonine kinase
protein kinase C, alpha
THAP domain containing 8
epoxide hydrolase 2, cytoplasmic
COP9 constitutive photomorphogenic homolog subunit 7A (Arabidopsis)
NA
mitochondrial ribosomal protein L55
zinc finger protein 641
cleavage and polyadenylation specific factor 3-like
phospholipase A2, group IVF
selenoprotein O
NA
discoidin, CUB and LCCL domain containing 2
chromosome 17 open reading frame 97
guanine nucleotide binding protein (G protein), alpha 13
nuclear receptor subfamily 3, group C, member 1 (glucocorticoid receptor)
sperm associated antigen 7
histone deacetylase 9
nuclear protein localization 4 homolog (S. cerevisiae)
helicase (DNA) B
protein phosphatase 2, regulatory subunit A, alpha
dystrophia myotonica-protein kinase
low density lipoprotein receptor class A domain containing 3
transmembrane protein 2
interleukin 12 receptor, beta 2
UBX domain protein 7
chromosome 19 open reading frame 54
mediator complex subunit 24
DMRT-like family A1
polymerase (DNA directed), epsilon 3 (p17 subunit)
meningioma expressed antigen 5 (hyaluronidase)
ankyrin repeat domain 44
S100 calcium binding protein A13
Mdm2, p53 E3 ubiquitin protein ligase homolog (mouse)
solute carrier family 25 (mitochondrial carrier; oxoglutarate carrier), member 11
transmembrane protein 42
3-hydroxymethyl-3-methylglutaryl-CoA lyase

small nuclear ribonucleoprotein polypeptide A
annexin A3
chromosome 1 open reading frame 85
alcohol dehydrogenase, iron containing, 1
nudix (nucleoside diphosphate linked moiety X)-type motif 8
cell adhesion molecule with homology to L1CAM (close homolog of L1)
uroporphyrinogen decarboxylase
glutathione reductase
2,4-dienoyl CoA reductase 2, peroxisomal
CD164 molecule, sialomucin
hepsin
1-aminocyclopropane-1-carboxylate synthase homolog (Arabidopsis)(non-functional)
intraflagellar transport 172 homolog (Chlamydomonas)
UBX domain protein 7
COMM domain containing 6
NA
NA
NA
calcium and integrin binding 1 (calmyrin)
mitochondrial ribosomal protein L45
inositol-3-phosphate synthase 1
solute carrier family 5 (sodium/myo-inositol cotransporter), member 3
processing of precursor 7, ribonuclease P/MRP subunit (S. cerevisiae)
ATP5S-like
signal sequence receptor, alpha
NA
transmembrane protein 134
solute carrier family 27 (fatty acid transporter), member 5
enoyl CoA hydratase, short chain, 1, mitochondrial
asparaginase like 1
tumor protein D52-like 2
exocyst complex component 5
chromosome 11 open reading frame 31
trinucleotide repeat containing 6B
NA
chromosome 17 open reading frame 108
HEAT repeat containing 2
tubby like protein 4
solute carrier family 12 (potassium/chloride transporter), member 5
polycomb group ring finger 5
src kinase associated phosphoprotein 2
solute carrier family 36 (proton/amino acid symporter), member 1
dysbindin (dystrobrevin binding protein 1) domain containing 1
activator of basal transcription 1
BCL2-associated transcription factor 1
2-oxoglutarate and iron-dependent oxygenase domain containing 1
SEC62 homolog (S. cerevisiae)
dehydrogenase E1 and transketolase domain containing 1
TSC22 domain family, member 1
lysine (K)-specific demethylase 4D
zinc finger protein 274
v-maf musculoaponeurotic fibrosarcoma oncogene homolog G (avian)
dopachrome tautomerase (dopachrome delta-isomerase, tyrosine-related protein 2)

G protein-coupled receptor 183
NA
chromosome X open reading frame 57
IKAROS family zinc finger 1 (Ikaros)
signal recognition particle receptor, B subunit
dual-specificity tyrosine-(Y)-phosphorylation regulated kinase 2
mitochondrial ribosomal protein S17
mitochondrial ribosomal protein S17
paraspeckle component 1
MOB kinase activator 1A
PRP4 pre-mRNA processing factor 4 homolog (yeast)
chromosome 6 open reading frame 57
mediator complex subunit 6
glutathione reductase
F-box protein 3
histone cluster 1, H2ai
zinc finger, MYM-type 2
neurotrophic tyrosine kinase, receptor, type 2
tetraspanin 5
Kruppel-like factor 7 (ubiquitous)
polymerase (RNA) III (DNA directed) polypeptide A, 155kDa
NA
NA
family with sequence similarity 49, member A
alkB, alkylation repair homolog 7 (E. coli)
protein kinase C and casein kinase substrate in neurons 2
poly(A) polymerase alpha
cytochrome c oxidase subunit IV isoform 1
RAB18, member RAS oncogene family
chromosome 1 open reading frame 63
PRP38 pre-mRNA processing factor 38 (yeast) domain containing B
trafficking protein particle complex 10
triple functional domain (PTPRF interacting)
DBF4 homolog B (S. cerevisiae)
RNA binding motif protein 25
tubulin, gamma complex associated protein 3
glycosyltransferase-like domain containing 1
pyridoxal-dependent decarboxylase domain containing 1
CTAGE family, member 5
pyridoxal-dependent decarboxylase domain containing 1
protocadherin 9
dedicator of cytokinesis 2
versican
microtubule associated monoxygenase, calponin and LIM domain containing 3
NA
NA
Kruppel-like factor 6
sperm associated antigen 9
O-linked N-acetylglucosamine (GlcNAc) transferase (UDP-N-acetylglucosamine:polypeptide-N-acetylglucos
PNN-interacting serine/arginine-rich protein
nicotinamide phosphoribosyltransferase
phosphoinositide-3-kinase, class 2, alpha polypeptide
septin 6

SURP and G patch domain containing 2
zinc finger protein 451
zinc finger protein 451
vaccinia related kinase 2
ribosomal protein S6 kinase, 52kDa, polypeptide 1
sterile alpha motif domain containing 4A
platelet-derived growth factor receptor, alpha polypeptide
zinc finger, CCHC domain containing 7
zinc finger, CCHC domain containing 7
TATA box binding protein (TBP)-associated factor, RNA polymerase I, A, 48kDa
cerebral cavernous malformation 2
serine palmitoyltransferase, long chain base subunit 3
phosphomannomutase 2
vacuolar protein sorting 72 homolog (S. cerevisiae)
Fraser syndrome 1
ER degradation enhancer, mannosidase alpha-like 3
KIN, antigenic determinant of recA protein homolog (mouse)
septin 10
coiled-coil domain containing 76
homocysteine-inducible, endoplasmic reticulum stress-inducible, ubiquitin-like domain member 1
ubiquitously transcribed tetratricopeptide repeat gene, Y-linked
cancer susceptibility candidate 4
immunoglobulin superfamily, member 6
SYF2 homolog, RNA splicing factor (S. cerevisiae)
serine/arginine-rich splicing factor 11
Down syndrome encephalopathy related protein 1
sarcolemma associated protein
fucosyltransferase 10 (alpha (1,3) fucosyltransferase)
ATP-binding cassette, sub-family A (ABC1), member 11, pseudogene
YME1-like 1 (S. cerevisiae)
zinc finger protein 844
ADAM metallopeptidase domain 17
GALI1870
septin 6
membrane-associated ring finger (C3HC4) 6, E3 ubiquitin protein ligase
nuclear factor of activated T-cells, cytoplasmic, calcineurin-dependent 2
chromosome 9 open reading frame 114
ring finger protein 7
chromodomain helicase DNA binding protein 2
NA
amyotrophic lateral sclerosis 2 (juvenile) chromosome region, candidate 8
NA
NA
caspase 8, apoptosis-related cysteine peptidase
NA
nuclear factor of activated T-cells, cytoplasmic, calcineurin-dependent 2
NA
malate dehydrogenase 2, NAD (mitochondrial)
CUGBP, Elav-like family member 1
adaptor-related protein complex 5, sigma 1 subunit
neurofibromin 1
zinc finger protein 839
neurofibromin 1

collagen, type XII, alpha 1
mitochondrial ribosomal protein L12
ADP-ribosylation-like factor 6 interacting protein 4
ADP-ribosylation-like factor 6 interacting protein 4
ubiquitously transcribed tetratricopeptide repeat gene, Y-linked
ubiquitously transcribed tetratricopeptide repeat gene, Y-linked
ubiquitously transcribed tetratricopeptide repeat gene, Y-linked
uncharacterized LOC100506797
small nucleolar RNA, C/D box 17
small nucleolar RNA, H/ACA box 50
small nucleolar RNA, H/ACA box 16B
small nucleolar RNA, H/ACA box 70C (retrotransposed)
mitochondrial ribosomal protein S11
alkB, alkylation repair homolog 7 (E. coli)
mitochondrial ribosomal protein S11
trafficking protein particle complex 2-like
NA
family with sequence similarity 86, member C1
DEAD (Asp-Glu-Ala-Asp) box helicase 17
stromal cell derived factor 4
neuroblastoma, suppression of tumorigenicity 1
high mobility group 20B
plasminogen activator, tissue
DEAD (Asp-Glu-Ala-Asp) box polypeptide 3, X-linked
chromosome 10 open reading frame 10
cyclin-dependent kinase 2 associated protein 2
proteasome (prosome, macropain) subunit, beta type, 10
tripartite motif containing 44
AF4/FMR2 family, member 1
transducin (beta)-like 1 X-linked receptor 1
retinoblastoma binding protein 4
NA
SERPINE1 mRNA binding protein 1
chromosome 11 open reading frame 49
ADP-ribosylation factor interacting protein 1
NA
ADAM metallopeptidase domain 15
major facilitator superfamily domain containing 12
DCP2 decapping enzyme homolog (S. cerevisiae)
cullin-associated and neddylation-dissociated 1
nucleus accumbens associated 1, BEN and BTB (POZ) domain containing
influenza virus NS1A binding protein
argininosuccinate lyase
YLP motif containing 1
ATX1 antioxidant protein 1 homolog (yeast)
LYR motif containing 4
metallothionein 1F
NmrA-like family domain containing 1
Mdm4 p53 binding protein homolog (mouse)
seizure related 6 homolog (mouse)-like 2
seizure related 6 homolog (mouse)-like 2
insulin-like growth factor binding protein 2, 36kDa
chromobox homolog 2

GA binding protein transcription factor, alpha subunit 60kDa
bone morphogenetic protein receptor, type IA
GAR1 ribonucleoprotein homolog (yeast)
DDB1 and CUL4 associated factor 16
chromosome 14 open reading frame 79
CUGBP, Elav-like family member 6
serine dehydratase-like
NA
ubiquitin specific peptidase 31
inositol-3-phosphate synthase 1
myeloid/lymphoid or mixed-lineage leukemia 5 (trithorax homolog, Drosophila)
myeloid/lymphoid or mixed-lineage leukemia 5 (trithorax homolog, Drosophila)
tRNA-yW synthesizing protein 1 homolog B (S. cerevisiae)
sortilin 1
matrix metallopeptidase 28
spindlin family, member 3
KIAA2026
family with sequence similarity 181, member B
Rho GTPase activating protein 5
N-ethylmaleimide-sensitive factor attachment protein, gamma
NA
lipocalin 6
peroxisomal biogenesis factor 19
pleckstrin homology domain containing, family A (phosphoinositide binding specific) member 4
armadillo repeat containing 8
vasoactive intestinal peptide
vacuolar protein sorting 13 homolog C (S. cerevisiae)
THO complex 2
family with sequence similarity 70, member B
zinc finger protein 660
basigin (Ok blood group)
zinc finger protein 347
echinoderm microtubule associated protein like 5
NA
Yip1 domain family, member 6
myeloid/lymphoid or mixed-lineage leukemia 3
valosin containing protein (p97)/p47 complex interacting protein 1
basigin (Ok blood group)
polymerase (RNA) III (DNA directed) polypeptide G (32kD)
CDC42 binding protein kinase beta (DMPK-like)
opioid receptor, delta 1
F-box protein 44
acid phosphatase 2, lysosomal
DNA-damage inducible 1 homolog 2 (S. cerevisiae)
zinc finger and SCAN domain containing 18
NA
synapse defective 1, Rho GTPase, homolog 1 (C. elegans)
v-abl Abelson murine leukemia viral oncogene homolog 2
testis expressed 261
nuclear protein, ataxia-telangiectasia locus
stromal antigen 1
neuroguidin, EIF4E binding protein
ataxin 1-like

zinc finger protein 688
dipeptidyl-peptidase 3
chromosome 20 open reading frame 27
serine/threonine kinase 32C
synapse defective 1, Rho GTPase, homolog 1 (C. elegans)
zinc finger protein 451
translocator protein (18kDa)
carbonyl reductase 1
NA
CDP-diacylglycerol--inositol 3-phosphatidyltransferase
family with sequence similarity 192, member A
actinin, alpha 4
attractin-like 1
PHD finger protein 17
mannose-P-dolichol utilization defect 1
mannose-P-dolichol utilization defect 1
cysteine-rich protein 2
THAP domain containing 8
BCL2-antagonist/killer 1
GrpE-like 2, mitochondrial (E. coli)
plasminogen activator, tissue
neuroblastoma, suppression of tumorigenicity 1
EPS8-like 1
EGF containing fibulin-like extracellular matrix protein 2
protein tyrosine phosphatase, non-receptor type 7
ribulose-5-phosphate-3-epimerase
angiogenic factor with G patch and FHA domains 1
transmembrane 9 superfamily member 3
MON2 homolog (S. cerevisiae)
MON2 homolog (S. cerevisiae)
zinc finger protein 638
5-methyltetrahydrofolate-homocysteine methyltransferase
suppressor of variegation 4-20 homolog 1 (Drosophila)
von Willebrand factor
chromosome 3 open reading frame 64
sialic acid acetylesterase
uncharacterized LOC284889
RNA binding motif protein 28
carbonyl reductase 1
F-box protein 44
Mdm2, p53 E3 ubiquitin protein ligase homolog (mouse)
plasminogen activator, tissue
basigin (Ok blood group)
RAN binding protein 2
synapse defective 1, Rho GTPase, homolog 1 (C. elegans)
checkpoint with forkhead and ring finger domains, E3 ubiquitin protein ligase
Rho GTPase activating protein 5
zinc finger protein 135
attractin-like 1
adaptor-related protein complex 2, sigma 1 subunit
chromosome 5 open reading frame 42
basal cell adhesion molecule (Lutheran blood group)
TBC1 domain family, member 22A

STT3, subunit of the oligosaccharyltransferase complex, homolog B (S. cerevisiae)
arrestin domain containing 1
attractin-like 1
Duffy blood group, chemokine receptor
small nucleolar RNA, H/ACA box 21
spermatogenesis associated 2
ATP synthase, H+ transporting, mitochondrial F1 complex, gamma polypeptide 1
STT3, subunit of the oligosaccharyltransferase complex, homolog B (S. cerevisiae)
NA
NA
GTPase activating protein and VPS9 domains 1
DnaJ (Hsp40) homolog, subfamily B, member 2
NA
uncharacterized LOC400943
single immunoglobulin and toll-interleukin 1 receptor (TIR) domain
NA
RAN, member RAS oncogene family
NA

| global.mean | global.sd | OC.mean | OT2.mean | YC.mean | YT2.mean | T2.vs.C.FC |
|---|---|---|---|---|---|---|
| 15,1136781 | 0,472894167 | 14,828 | 15,247747 | 14,82864 | 15,366647 | 1,39365947 |
| 15,8073348 | 0,581391254 | 16,15773 | 15,629093 | 16,25663 | 15,453808 | -1,5863729 |
| 5,05853568 | 0,238688141 | 4,91317 | 5,1898249 | 4,890048 | 5,1256417 | 1,1942658 |
| 15,6390815 | 0,225198741 | 15,82885 | 15,489924 | 15,73279 | 15,600545 | -1,1773832 |
| 13,3293 | 0,266863722 | 13,54784 | 13,167753 | 13,40888 | 13,289649 | -1,1889264 |
| 7,23242007 | 0,24661079 | 7,402634 | 7,1240838 | 7,440282 | 7,0930244 | -1,2422057 |
| 8,39617773 | 0,348031545 | 8,549835 | 8,243706 | 8,782129 | 8,2169645 | -1,3525169 |
| 8,92051402 | 0,383063047 | 8,615772 | 9,2587632 | 8,729987 | 8,8879621 | 1,31994994 |
| 9,85338849 | 0,29073629 | 10,02765 | 9,7489022 | 10,08334 | 9,6926819 | -1,2611158 |
| 18,5941483 | 0,201700626 | 18,77925 | 18,435039 | 18,66001 | 18,588227 | -1,1550829 |
| 7,05473913 | 0,630173557 | 7,735615 | 7,0587549 | 6,767006 | 6,6438159 | -1,3195308 |
| 16,6653461 | 0,190087742 | 16,82823 | 16,53439 | 16,73955 | 16,639519 | -1,1462617 |
| 12,9419345 | 0,236631957 | 13,0907 | 12,741193 | 13,11525 | 12,952381 | -1,1943209 |
| 9,89940783 | 0,221829778 | 10,06566 | 9,7767023 | 10,04697 | 9,8168066 | -1,1971118 |
| 8,12851027 | 0,347684408 | 7,919014 | 8,330276 | 7,901462 | 8,2006922 | 1,27920368 |
| 10,1234292 | 0,388580806 | 10,43208 | 9,887318 | 10,34152 | 10,013556 | -1,3531874 |
| 9,0431122 | 0,222676588 | 9,221765 | 8,9347118 | 9,133471 | 8,9671224 | -1,1701561 |
| 9,8185661 | 0,213116729 | 9,990478 | 9,7123085 | 9,928124 | 9,7342153 | -1,1777547 |
| 7,98427288 | 0,459318001 | 7,60563 | 8,217775 | 7,682389 | 8,2070469 | 1,48287938 |
| 8,90512144 | 0,345968588 | 8,660855 | 9,1330035 | 8,688708 | 8,9691372 | 1,29799838 |
| 7,3937369 | 0,300710849 | 7,176078 | 7,5920383 | 7,225604 | 7,4416645 | 1,24488289 |
| 9,24017499 | 0,426063869 | 8,855332 | 9,517821 | 8,937324 | 9,4158271 | 1,48503425 |
| 12,0384267 | 0,299110053 | 11,81376 | 12,165191 | 11,93279 | 12,140659 | 1,21390043 |
| 8,85019772 | 0,323103245 | 8,559222 | 8,9663458 | 8,928203 | 8,9113465 | 1,14483036 |
| 4,88703001 | 0,441406306 | 5,212679 | 4,8077869 | 5,047044 | 4,6093118 | -1,3391449 |
| 10,2431203 | 0,212211648 | 10,3959 | 10,119691 | 10,35394 | 10,194878 | -1,1628277 |
| 19,7529146 | 0,310289308 | 20,07591 | 19,517425 | 19,80826 | 19,727749 | -1,2478962 |
| 9,00262496 | 0,226785304 | 8,81415 | 9,1310551 | 8,902262 | 9,0689305 | 1,1824563 |
| 12,472958 | 0,32307971 | 12,21636 | 12,555486 | 12,36886 | 12,65449 | 1,24175275 |
| 13,6775596 | 0,347547235 | 13,38061 | 13,799636 | 13,53921 | 13,866491 | 1,29518413 |
| 14,316177 | 0,326415943 | 14,6239 | 14,124149 | 14,29377 | 14,298494 | -1,1871567 |
| 12,4871799 | 0,202804868 | 12,70882 | 12,353863 | 12,53111 | 12,432405 | -1,1702636 |
| 6,54112415 | 0,188021631 | 6,717975 | 6,4496347 | 6,555786 | 6,4917912 | -1,1220737 |
| 12,9158025 | 0,222283896 | 13,05269 | 12,772486 | 13,08603 | 12,869292 | -1,1879478 |
| 13,5276086 | 0,240396034 | 13,66979 | 13,371053 | 13,70518 | 13,488076 | -1,1957541 |
| 13,5837676 | 0,269848066 | 13,78909 | 13,377036 | 13,73268 | 13,567978 | -1,2212642 |
| 11,3972948 | 0,36225423 | 11,12678 | 11,559266 | 11,23805 | 11,528417 | 1,28469752 |
| 16,9094826 | 0,346544387 | 17,2202 | 16,621529 | 16,99259 | 16,941048 | -1,2527589 |
| 9,72225432 | 0,215023732 | 9,906356 | 9,592871 | 9,810285 | 9,6682101 | -1,1710312 |
| 12,9490521 | 0,266732518 | 13,17997 | 12,852796 | 13,01843 | 12,826653 | -1,1970438 |
| 2,90521726 | 0,249941544 | 3,064369 | 2,7893659 | 3,068291 | 2,8111159 | -1,2025434 |
| 9,6287126 | 0,278101303 | 9,801028 | 9,5031681 | 9,817729 | 9,5194881 | -1,2294818 |
| 11,2439812 | 0,269426631 | 11,38076 | 11,050763 | 11,51953 | 11,19425 | -1,2549586 |
| 15,4593124 | 0,222973289 | 15,58883 | 15,268437 | 15,59413 | 15,49738 | -1,1555446 |
| 13,9402474 | 0,244438178 | 14,16998 | 13,764205 | 14,0257 | 13,904949 | -1,2001928 |
| 6,53848319 | 0,434323318 | 6,968342 | 6,4914313 | 6,710436 | 6,126394 | -1,4444059 |
| 5,39815999 | 0,247304038 | 5,608451 | 5,28707 | 5,466542 | 5,3116916 | -1,1794511 |
| 12,0951056 | 0,275503393 | 12,29081 | 11,968443 | 12,24512 | 11,990741 | -1,2212642 |
| 14,7144301 | 0,351773118 | 14,47994 | 14,856945 | 14,4829 | 14,881648 | 1,3084657 |
| 13,2203067 | 0,276925997 | 12,98056 | 13,335608 | 13,15698 | 13,323725 | 1,1982235 |
| 9,26056727 | 0,251030187 | 9,030995 | 9,4163055 | 9,173943 | 9,3207922 | 1,20253565 |
| 7,73842313 | 0,272168698 | 7,948359 | 7,5794745 | 7,911189 | 7,6470568 | -1,2453127 |

| | | | | | | |
|---|---|---|---|---|---|---|
| 12,5607722 | 0,301918134 | 12,83352 | 12,407129 | 12,59401 | 12,493364 | -1,2004029 |
| 7,41434751 | 0,226907361 | 7,623952 | 7,3074939 | 7,487564 | 7,3204783 | -1,1824439 |
| 16,1027216 | 0,339814679 | 15,93459 | 16,205663 | 15,84277 | 16,278079 | 1,27738592 |
| 14,1275968 | 0,197536603 | 14,34925 | 14,041528 | 14,12032 | 14,046835 | -1,1412436 |
| 8,10347532 | 0,280629342 | 8,276996 | 7,9751074 | 8,267476 | 8,0116241 | -1,2132443 |
| 13,4077075 | 0,314252705 | 13,70279 | 13,151846 | 13,52418 | 13,394039 | -1,2662325 |
| 3,13318834 | 0,197266582 | 2,979739 | 3,2733889 | 3,056995 | 3,1410961 | 1,13987515 |
| 6,37977168 | 0,197766199 | 6,545026 | 6,2793221 | 6,449584 | 6,3179576 | -1,1476361 |
| 10,0962195 | 0,195265099 | 10,25802 | 9,9861297 | 10,19721 | 10,0302 | -1,1642912 |
| 15,0787026 | 0,208065042 | 15,27113 | 14,941343 | 15,13089 | 15,048658 | -1,1534928 |
| 9,79240622 | 0,252657121 | 9,560524 | 9,9687095 | 9,684632 | 9,8426067 | 1,21678998 |
| 8,80952837 | 0,327198378 | 9,018545 | 8,6662628 | 8,966273 | 8,7097363 | -1,234913 |
| 7,59359739 | 0,21359394 | 7,765447 | 7,4947361 | 7,704378 | 7,4996907 | -1,1791105 |
| 8,58458166 | 0,24300568 | 8,748368 | 8,4388259 | 8,762682 | 8,5134098 | -1,2136961 |
| 13,5071368 | 0,240424156 | 13,67672 | 13,369829 | 13,62382 | 13,457748 | -1,1781181 |
| 13,2612761 | 0,300779525 | 13,05427 | 13,340423 | 13,06484 | 13,460114 | 1,26638137 |
| 14,6712983 | 0,281432218 | 14,88271 | 14,491821 | 14,79335 | 14,633746 | -1,2101973 |
| 8,46182644 | 0,317235778 | 8,746693 | 8,2974382 | 8,543488 | 8,3679584 | -1,2417653 |
| 4,38075952 | 0,217839035 | 4,579607 | 4,2807518 | 4,415712 | 4,3107773 | -1,1502079 |
| 19,8997608 | 0,336313059 | 20,24192 | 19,826403 | 19,67689 | 19,830676 | -1,0949506 |
| 10,1289431 | 0,306212988 | 10,36172 | 9,9079328 | 10,26139 | 10,116826 | -1,2304422 |
| 10,9585274 | 0,223396143 | 10,76503 | 11,056433 | 10,90355 | 11,038502 | 1,1592391 |
| 18,107336 | 0,30886986 | 17,85832 | 18,177958 | 17,99797 | 18,299875 | 1,24037149 |
| 11,2245322 | 0,29042316 | 11,44961 | 11,044986 | 11,34603 | 11,175772 | -1,2204733 |
| 14,7996981 | 0,377350758 | 14,58471 | 15,014455 | 14,56436 | 14,865931 | 1,28846816 |
| 9,20885769 | 0,216561884 | 9,380382 | 9,0886493 | 9,304201 | 9,1501058 | -1,1670887 |
| 9,23104748 | 0,322337917 | 8,880534 | 9,3894481 | 9,278245 | 9,3105851 | 1,20633228 |
| 6,3587116 | 0,252841546 | 6,524209 | 6,2171081 | 6,538372 | 6,280167 | -1,2164296 |
| 13,1650038 | 0,256067419 | 13,35458 | 13,063741 | 13,27218 | 13,06107 | -1,1900129 |
| 11,701783 | 0,433136306 | 12,12418 | 11,513768 | 11,70118 | 11,568729 | -1,2936334 |
| 11,2906457 | 0,174739367 | 11,44523 | 11,17476 | 11,34944 | 11,263035 | -1,1316571 |
| 16,348155 | 0,328954582 | 16,53154 | 16,090915 | 16,53076 | 16,391399 | -1,2226376 |
| 12,4888958 | 0,30161774 | 12,67648 | 12,313937 | 12,65683 | 12,438639 | -1,2229515 |
| 8,91056033 | 1,755704249 | 10,27235 | 8,222213 | 9,279898 | 8,3574536 | -2,801677 |
| 4,38121305 | 0,241697714 | 4,209603 | 4,4838547 | 4,261922 | 4,4754867 | 1,18419647 |
| 12,3815294 | 0,202160509 | 12,55597 | 12,213008 | 12,49753 | 12,365873 | -1,1787955 |
| 7,78148139 | 0,242191245 | 8,012408 | 7,6657252 | 7,677165 | 7,786691 | -1,0856645 |
| 15,4892649 | 0,330651406 | 15,29556 | 15,590031 | 15,26346 | 15,668481 | 1,27433877 |
| 9,13007536 | 0,304969503 | 9,347672 | 8,9644382 | 9,257829 | 9,067231 | -1,2200293 |
| 14,4234529 | 0,308668842 | 14,73812 | 14,320009 | 14,36977 | 14,31233 | -1,1791721 |
| 14,1696895 | 0,295341295 | 14,48759 | 14,013817 | 14,12073 | 14,115899 | -1,180421 |
| 16,5744996 | 0,21459647 | 16,74758 | 16,435163 | 16,68078 | 16,530816 | -1,1738032 |
| 9,56908645 | 0,34766946 | 9,909428 | 9,4675721 | 9,604215 | 9,3805363 | -1,2594268 |
| 14,8618976 | 0,369695309 | 15,09504 | 14,684007 | 15,07585 | 14,748826 | -1,2914817 |
| 10,8839548 | 0,207019665 | 11,04797 | 10,732495 | 10,94963 | 10,886889 | -1,1400589 |
| 5,14935832 | 0,213878512 | 5,319571 | 5,0415856 | 5,265473 | 5,0643364 | -1,1806332 |
| 12,3119198 | 0,443316775 | 11,98187 | 12,476237 | 12,08171 | 12,533404 | 1,38801939 |
| 18,4119899 | 0,28870469 | 18,13186 | 18,468456 | 18,34104 | 18,624915 | 1,2399113 |
| 12,1908853 | 0,180703205 | 12,36591 | 12,065526 | 12,21704 | 12,176849 | -1,1252844 |
| 4,65603606 | 0,209204351 | 4,777227 | 4,5139815 | 4,797809 | 4,638426 | -1,1577421 |
| 11,9918817 | 0,286951984 | 11,76555 | 12,139582 | 11,89561 | 12,064785 | 1,2071486 |
| 7,83862888 | 0,280198347 | 8,079753 | 7,6547601 | 7,903461 | 7,8154164 | -1,1945928 |
| 12,2552308 | 0,22063927 | 12,451 | 12,153162 | 12,30894 | 12,179081 | -1,1597797 |

| | | | | | | |
|---|---|---|---|---|---|---|
| 10,9020654 | 0,24105553 | 10,72873 | 11,005134 | 10,76384 | 11,008657 | 1,19798775 |
| 12,2277936 | 0,232855911 | 12,45057 | 12,093029 | 12,21951 | 12,205123 | -1,1375767 |
| 12,5861347 | 0,248336787 | 12,76612 | 12,414569 | 12,70475 | 12,567853 | -1,1844568 |
| 4,96468721 | 0,240861945 | 5,159335 | 4,8493779 | 5,110481 | 4,8501319 | -1,2185394 |
| 1,82675662 | 0,50058647 | 1,605759 | 2,1356148 | 1,353294 | 1,9280517 | 1,46642811 |
| 19,7656514 | 0,482686395 | 20,23456 | 19,531748 | 19,94952 | 19,537446 | -1,4716563 |
| 18,8152112 | 0,24237164 | 19,08643 | 18,685519 | 18,84761 | 18,720866 | -1,2006598 |
| 11,4308957 | 0,296180718 | 11,58196 | 11,264583 | 11,67525 | 11,355458 | -1,2471036 |
| 8,59708785 | 0,215206911 | 8,814632 | 8,4482946 | 8,74628 | 8,5012116 | -1,2360209 |
| 13,672019 | 0,336477754 | 13,93704 | 13,430005 | 13,84173 | 13,635343 | -1,2805018 |
| 13,0910105 | 0,301570456 | 13,31186 | 12,879094 | 13,26394 | 13,053831 | -1,2495748 |
| 10,845505 | 0,248071076 | 11,06983 | 10,6439 | 10,98594 | 10,812494 | -1,2308781 |
| 15,5416822 | 0,40875805 | 15,88928 | 15,284503 | 15,7463 | 15,432073 | -1,3750638 |
| 11,4297983 | 0,207453776 | 11,59729 | 11,318104 | 11,55349 | 11,347242 | -1,1832216 |
| 18,0586697 | 0,424684829 | 18,32017 | 17,787619 | 18,44362 | 17,930683 | -1,4366871 |
| 4,71460585 | 0,457609297 | 4,295482 | 4,7961081 | 4,597652 | 5,043231 | 1,38809152 |
| 18,5636213 | 0,266061771 | 18,83645 | 18,426202 | 18,47736 | 18,548378 | -1,1247575 |
| 8,58098972 | 0,349964279 | 8,762359 | 8,3830571 | 8,803688 | 8,5307258 | -1,2536479 |
| 14,9893648 | 0,225452931 | 15,22259 | 14,918113 | 14,97229 | 14,886866 | -1,1446877 |
| 6,0569476 | 0,307816724 | 6,352132 | 6,0346276 | 5,941976 | 5,9018073 | -1,1319709 |
| 13,2829074 | 0,223395198 | 13,47872 | 13,160777 | 13,40038 | 13,192538 | -1,1998833 |
| 10,0415791 | 0,389074552 | 10,40049 | 9,9590586 | 9,893163 | 9,9245768 | -1,1526944 |
| 12,1657291 | 0,382528756 | 12,55952 | 12,09063 | 12,10554 | 11,957234 | -1,238507 |
| 12,0683028 | 0,275760815 | 11,89323 | 12,183453 | 11,86406 | 12,20148 | 1,24299568 |
| 15,4595816 | 0,272982535 | 15,22748 | 15,59737 | 15,30242 | 15,585817 | 1,25409115 |
| 7,95447658 | 0,225755904 | 8,131139 | 7,8494089 | 8,109562 | 7,8373435 | -1,2116512 |
| 12,2571484 | 0,317867402 | 12,38577 | 12,02448 | 12,51824 | 12,27037 | -1,2350578 |
| 10,6356387 | 0,355900889 | 10,37282 | 10,750857 | 10,46562 | 10,822781 | 1,29020117 |
| 14,6824321 | 0,266615966 | 14,9488 | 14,55306 | 14,65218 | 14,629416 | -1,1560899 |
| 7,33458826 | 0,220909427 | 7,101253 | 7,4053194 | 7,29196 | 7,4736232 | 1,18334039 |
| 8,80117338 | 0,436572689 | 8,478119 | 8,9843091 | 8,659702 | 8,9408896 | 1,31374833 |
| 15,4560185 | 0,294337235 | 15,25154 | 15,556526 | 15,296 | 15,605225 | 1,23722171 |
| 5,87140585 | 0,367878559 | 6,148319 | 5,6157296 | 6,120979 | 5,7930978 | -1,347453 |
| 20,3108866 | 0,360303146 | 20,09807 | 20,45609 | 20,03653 | 20,482149 | 1,32117185 |
| 4,81575931 | 0,215475362 | 5,001873 | 4,7164758 | 4,873756 | 4,7419047 | -1,1555857 |
| 10,5793307 | 0,340143237 | 10,89179 | 10,343353 | 10,59361 | 10,588348 | -1,2115456 |
| 6,04407659 | 0,272223201 | 5,816349 | 6,1823989 | 5,996688 | 6,1000912 | 1,1766836 |
| 11,1627861 | 0,366543265 | 11,48047 | 10,949343 | 11,26867 | 11,085358 | -1,2809531 |
| 10,423134 | 0,284427945 | 10,1949 | 10,570171 | 10,19824 | 10,575626 | 1,29803443 |
| 8,78864213 | 0,313292692 | 8,607857 | 8,9370578 | 8,576254 | 8,8916441 | 1,25031867 |
| 11,4300485 | 0,294947616 | 11,18396 | 11,502658 | 11,32597 | 11,61454 | 1,23425025 |
| 10,6479069 | 0,465358345 | 10,37037 | 10,864136 | 10,34671 | 10,805055 | 1,3909336 |
| 10,9300623 | 0,298516454 | 10,72055 | 11,005596 | 10,78564 | 11,104158 | 1,23266402 |
| 14,4983113 | 0,256038769 | 14,60242 | 14,289168 | 14,6341 | 14,579316 | -1,1360439 |
| 15,9958642 | 0,273099835 | 16,19626 | 15,82562 | 16,18916 | 15,913844 | -1,2509101 |
| 14,9203014 | 0,273762024 | 14,74506 | 15,062929 | 14,73523 | 15,009142 | 1,22763986 |
| 14,1484561 | 0,295891538 | 13,96912 | 14,285701 | 13,8852 | 14,294153 | 1,28589054 |
| 13,5689666 | 0,376485432 | 13,30103 | 13,616262 | 13,39537 | 13,844115 | 1,30313488 |
| 12,956522 | 0,324959306 | 12,74758 | 13,100744 | 12,77223 | 13,071632 | 1,25377947 |
| 13,2426295 | 0,3431526 | 13,59407 | 12,989523 | 13,35655 | 13,17928 | -1,3112193 |
| 13,1989888 | 0,315738687 | 13,46719 | 12,966941 | 13,35784 | 13,154164 | -1,276296 |
| 8,78663512 | 0,265808248 | 8,995272 | 8,7140304 | 8,874665 | 8,6436014 | -1,1942896 |
| 9,7934411 | 0,272985993 | 9,615712 | 9,9276289 | 9,552172 | 9,9282465 | 1,26926678 |

| | | | | | | |
|---|---|---|---|---|---|---|
| 4,65510507 | 0,225885155 | 4,498594 | 4,7833835 | 4,547423 | 4,6988145 | 1,16319298 |
| 6,38288862 | 0,227564184 | 6,155678 | 6,4867985 | 6,320571 | 6,4887166 | 1,18890478 |
| 15,0115479 | 0,486366784 | 14,52848 | 15,15288 | 15,08436 | 15,20887 | 1,29635046 |
| 9,96855997 | 0,266805711 | 10,24768 | 9,8380266 | 10,04161 | 9,8441139 | -1,234201 |
| 13,9610092 | 0,290928466 | 14,25277 | 13,774867 | 14,02942 | 13,895337 | -1,2362693 |
| 7,16249404 | 0,27677758 | 6,904267 | 7,278905 | 7,171254 | 7,2370381 | 1,16490411 |
| 12,4533293 | 0,288724254 | 12,65618 | 12,355537 | 12,63669 | 12,288237 | -1,2522744 |
| 3,04177977 | 0,323534626 | 3,40801 | 2,9850422 | 2,88556 | 2,8923008 | -1,1551767 |
| 15,2475858 | 0,42494995 | 15,0158 | 15,388004 | 14,83915 | 15,521159 | 1,44103284 |
| 13,9675338 | 0,305925075 | 13,66927 | 14,019064 | 13,95144 | 14,168883 | 1,21724649 |
| 14,0182941 | 0,368806042 | 13,75642 | 14,135624 | 13,76078 | 14,254598 | 1,35332638 |
| 13,8748892 | 0,399369963 | 13,60996 | 14,028424 | 13,6639 | 14,042428 | 1,31813112 |
| 10,6880209 | 0,363826003 | 10,39062 | 10,822495 | 10,52771 | 10,875627 | 1,31029719 |
| 6,54652879 | 0,482838627 | 6,845265 | 6,3943006 | 6,817391 | 6,3127594 | -1,3926166 |
| 13,7684486 | 0,296668446 | 13,5322 | 13,840341 | 13,66079 | 13,947583 | 1,2289839 |
| 10,4784353 | 0,208730552 | 10,32983 | 10,59508 | 10,32275 | 10,558182 | 1,18948525 |
| 18,7776858 | 0,331954396 | 18,6119 | 18,886557 | 18,51332 | 18,946577 | 1,27806079 |
| 5,83879724 | 0,280541074 | 5,614564 | 5,9795164 | 5,769254 | 5,9022577 | 1,18836468 |
| 12,7296751 | 0,394019756 | 12,50326 | 12,881842 | 12,40274 | 12,935687 | 1,37150792 |
| 0,98026118 | 0,25597853 | 1,157673 | 0,8578218 | 1,09171 | 0,9095196 | -1,1818283 |
| 14,3728038 | 0,462498049 | 14,82446 | 14,196891 | 14,33717 | 14,22137 | -1,2938634 |
| 11,1844502 | 0,27645803 | 11,37634 | 10,99959 | 11,32241 | 11,160396 | -1,2052939 |
| 15,9712047 | 0,281624955 | 16,16813 | 15,770795 | 16,18348 | 15,916946 | -1,2586994 |
| 13,5350369 | 0,253547755 | 13,68422 | 13,365046 | 13,70335 | 13,511233 | -1,1938699 |
| 1,169034 | 0,189688197 | 1,360643 | 1,0607593 | 1,23498 | 1,0965286 | -1,1640616 |
| 12,8740044 | 0,332519733 | 13,16437 | 12,656256 | 12,92957 | 12,85515 | -1,2237157 |
| 11,6964024 | 0,331072177 | 11,45197 | 11,872494 | 11,40978 | 11,864833 | 1,35452662 |
| 5,5848837 | 0,255640726 | 5,748581 | 5,45001 | 5,724016 | 5,5241102 | -1,1885794 |
| 7,94832125 | 0,385186507 | 8,269164 | 7,7498302 | 8,14142 | 7,7979343 | -1,348551 |
| 15,1402443 | 0,315661544 | 15,48377 | 14,932291 | 15,24615 | 15,03425 | -1,3028652 |
| 14,1372777 | 0,262089311 | 14,37468 | 13,983236 | 14,29725 | 14,024353 | -1,2589057 |
| 5,22835876 | 0,209529521 | 5,031049 | 5,3219843 | 5,298756 | 5,2414829 | 1,08435037 |
| 5,45896228 | 0,260671651 | 5,18007 | 5,5913275 | 5,482219 | 5,5232282 | 1,16969571 |
| 1,25309803 | 0,430054484 | 1,138469 | 1,6275712 | 1,092972 | 0,9972403 | 1,14606193 |
| 1,14767206 | 0,243949682 | 1,334445 | 1,053857 | 1,248425 | 1,0410408 | -1,1842605 |
| 7,97911078 | 0,24235348 | 8,111237 | 7,8281752 | 8,170382 | 7,9331635 | -1,197595 |
| 16,5542282 | 0,312670366 | 16,34521 | 16,697996 | 16,33851 | 16,6888 | 1,27591849 |
| 3,21174517 | 0,262296056 | 3,387772 | 3,0488976 | 3,394627 | 3,14781 | -1,2250546 |
| 8,26789068 | 0,304265312 | 8,440892 | 8,0877311 | 8,477985 | 8,2109746 | -1,2397813 |
| 2,58073029 | 0,661137027 | 3,050786 | 2,2442616 | 3,095221 | 2,2762509 | -1,7565531 |
| 20,2153158 | 0,385515291 | 20,62462 | 19,999956 | 20,19297 | 20,139249 | -1,2650479 |
| 7,8888946 | 0,305159578 | 8,124518 | 7,723318 | 8,03816 | 7,7977477 | -1,2490281 |
| 10,1612088 | 0,266300796 | 10,38182 | 9,976367 | 10,3198 | 10,100347 | -1,2418155 |
| 3,26954702 | 0,340823108 | 3,470388 | 3,0898723 | 3,475617 | 3,1907992 | -1,2593395 |
| 8,11784717 | 0,205686052 | 8,258145 | 7,9936858 | 8,249925 | 8,0683404 | -1,1671761 |
| 7,7298018 | 0,329705912 | 8,052719 | 7,4551512 | 7,844598 | 7,716025 | -1,2861603 |
| 16,6145298 | 0,489422475 | 17,06758 | 16,381733 | 16,63663 | 16,495533 | -1,3318865 |
| 10,9683963 | 0,260162699 | 11,15231 | 10,825717 | 11,07904 | 10,9169 | -1,1845724 |
| 13,0684073 | 0,224350735 | 13,28905 | 12,935262 | 13,09323 | 13,025743 | -1,157199 |
| 14,8104311 | 0,307804138 | 14,59251 | 14,973421 | 14,66605 | 14,886704 | 1,23181208 |
| 4,65683999 | 0,245103877 | 4,439123 | 4,7908157 | 4,576262 | 4,7294754 | 1,19123093 |
| 13,6864854 | 0,31982743 | 13,95679 | 13,494176 | 13,87897 | 13,572007 | -1,3056683 |
| 19,4493593 | 0,478303879 | 20,00197 | 19,222702 | 19,44642 | 19,25339 | -1,4007021 |

| | | | | | | |
|---|---|---|---|---|---|---|
| 5,5301717 | 0,487761813 | 5,187478 | 5,720441 | 5,314115 | 5,7227722 | 1,38588743 |
| 9,49692514 | 0,74763459 | 9,090645 | 9,7985793 | 9,046533 | 9,7505132 | 1,63122637 |
| 10,6794147 | 0,219144442 | 10,78813 | 10,503336 | 10,81948 | 10,71426 | -1,1447307 |
| 4,97425261 | 0,2953431 | 5,220224 | 4,8402023 | 5,07455 | 4,8658586 | -1,226338 |
| 16,6565932 | 0,324131419 | 16,86399 | 16,443838 | 16,88724 | 16,597223 | -1,2790613 |
| 14,4840688 | 0,306035515 | 14,66986 | 14,288327 | 14,74929 | 14,401901 | -1,2874016 |
| -1,06232938 | 0,286733895 | -1,256884 | -0,8764607 | -1,19634 | -1,039597 | 1,20462193 |
| 1,9671219 | 0,260656642 | 2,156086 | 1,8238646 | 2,068832 | 1,9173854 | -1,1824948 |
| 13,4402125 | 0,326670299 | 13,75539 | 13,299552 | 13,34691 | 13,397088 | -1,1509534 |
| 5,85364495 | 0,482327667 | 6,344296 | 5,7149628 | 5,891476 | 5,5803111 | -1,3853489 |
| 10,2296983 | 0,274928217 | 10,47253 | 10,063702 | 10,45635 | 10,086494 | -1,309796 |
| 7,86773954 | 0,304804299 | 8,138496 | 7,7609482 | 7,888312 | 7,7534029 | -1,1943523 |
| 14,3999475 | 0,289871715 | 14,21372 | 14,505687 | 14,22068 | 14,53892 | 1,23551081 |
| 7,19515993 | 0,331863076 | 6,963671 | 7,3958229 | 7,033998 | 7,2478266 | 1,25092035 |
| 5,54142446 | 0,357194155 | 5,251435 | 5,7739358 | 5,384069 | 5,6033151 | 1,29313549 |
| 10,5876167 | 0,545939962 | 11,18321 | 10,468016 | 10,28124 | 10,408711 | -1,225916 |
| 11,7269034 | 0,244561928 | 11,47813 | 11,829818 | 11,62758 | 11,87446 | 1,23053545 |
| 11,0083091 | 0,271825819 | 11,19262 | 10,882245 | 11,20091 | 10,887366 | -1,2413897 |
| 10,4254621 | 0,292945109 | 10,24081 | 10,581311 | 10,25174 | 10,499634 | 1,22620533 |
| 8,50951284 | 0,348943324 | 8,778291 | 8,2781609 | 8,733074 | 8,4245365 | -1,323478 |
| 6,12957363 | 0,233813927 | 5,911549 | 6,2324838 | 6,262518 | 6,1116353 | 1,06070662 |
| 14,5211544 | 0,253455927 | 14,78235 | 14,406047 | 14,47242 | 14,466503 | -1,1416447 |
| 7,40757604 | 0,258514374 | 7,644593 | 7,2814196 | 7,402251 | 7,3606938 | -1,1505835 |
| 8,57592942 | 0,313215292 | 8,790164 | 8,418727 | 8,712522 | 8,5005174 | -1,2240994 |
| 5,64476415 | 0,321582551 | 5,861346 | 5,5766929 | 5,80006 | 5,4491777 | -1,2464005 |
| 6,11451965 | 0,475512461 | 6,436142 | 6,0891686 | 6,306872 | 5,7561509 | -1,3649489 |
| 0,88524213 | 0,238291403 | 1,038763 | 0,7719144 | 1,02639 | 0,8060536 | -1,1839374 |
| 12,679121 | 0,323806935 | 12,98867 | 12,459047 | 12,78361 | 12,617406 | -1,2727148 |
| 0,39354716 | 0,26679727 | 0,642655 | 0,3005973 | 0,514528 | 0,2207567 | -1,2465273 |
| 11,656894 | 0,173589087 | 11,81704 | 11,552815 | 11,73502 | 11,598785 | -1,1488835 |
| 5,53114841 | 0,304564865 | 5,700872 | 5,3120542 | 5,727476 | 5,5320001 | -1,2244609 |
| 8,90105992 | 0,242479266 | 8,675563 | 9,0289966 | 8,8873 | 8,9474639 | 1,15412427 |
| 14,4265921 | 0,315397667 | 14,26708 | 14,559065 | 14,17046 | 14,556889 | 1,2650604 |
| 5,32385921 | 0,435585089 | 4,913469 | 5,4954415 | 5,4631 | 5,3832479 | 1,19008152 |
| 9,95977716 | 0,303850321 | 10,20485 | 9,793657 | 10,1206 | 9,854317 | -1,2646487 |
| 5,98626596 | 0,273768047 | 6,160136 | 5,8750114 | 6,118875 | 5,8924166 | -1,1939905 |
| 10,9577911 | 0,342869296 | 10,77297 | 11,091345 | 10,72485 | 11,094383 | 1,26922648 |
| 15,4121902 | 0,297045805 | 15,72306 | 15,202499 | 15,50505 | 15,343863 | -1,2665246 |
| 0,54055395 | 0,286300208 | 0,75896 | 0,401635 | 0,65816 | 0,4510479 | -1,2160635 |
| 4,43612637 | 0,220794444 | 4,632975 | 4,3653645 | 4,524495 | 4,3006978 | -1,1856712 |
| 2,37539976 | 0,397256667 | 2,686843 | 2,2368682 | 2,61129 | 2,135377 | -1,3783513 |
| 8,71293225 | 0,22524406 | 8,8863 | 8,607942 | 8,791782 | 8,6442487 | -1,1590519 |
| 10,6635664 | 0,354983965 | 10,39351 | 10,70937 | 10,52237 | 10,922866 | 1,28180569 |
| 13,3752211 | 0,346722808 | 13,20773 | 13,498914 | 13,06732 | 13,553894 | 1,30937382 |
| 6,51860112 | 0,284017788 | 6,760116 | 6,4584626 | 6,599398 | 6,3370014 | -1,2159004 |
| 14,8516255 | 0,304016488 | 14,6234 | 14,991169 | 14,65755 | 14,994608 | 1,27669292 |
| 4,76020706 | 0,314545649 | 4,476482 | 4,7555002 | 4,916778 | 4,913079 | 1,100119 |
| 11,7146449 | 0,531228776 | 11,21013 | 11,784844 | 11,964 | 11,909632 | 1,19762256 |
| 12,9628449 | 0,260149922 | 13,21721 | 12,814067 | 13,05494 | 12,869913 | -1,2261051 |
| 6,50973208 | 0,212328262 | 6,30845 | 6,5943267 | 6,52207 | 6,5719052 | 1,12338751 |
| 11,4701039 | 0,328048631 | 11,31469 | 11,630616 | 11,20138 | 11,570825 | 1,26811399 |
| 10,56254 | 0,447438509 | 10,81252 | 10,279896 | 10,83814 | 10,523873 | -1,3411245 |
| 10,105761 | 0,676281997 | 9,860604 | 10,419753 | 9,536139 | 10,279128 | 1,57033102 |

| | | | | | | |
|---|---|---|---|---|---|---|
| 10,3886112 | 0,394673679 | 10,79506 | 10,140892 | 10,54294 | 10,2478 | -1,389581 |
| 7,00189943 | 0,275237492 | 7,157018 | 6,8231058 | 7,206653 | 6,9617485 | -1,2221392 |
| 6,30374389 | 0,247455541 | 6,467852 | 6,1314032 | 6,48271 | 6,2636809 | -1,2122935 |
| 12,6256958 | 0,283385916 | 12,82349 | 12,45273 | 12,75061 | 12,59018 | -1,2021313 |
| 14,615307 | 0,359347813 | 14,33071 | 14,794447 | 14,46417 | 14,73293 | 1,2889964 |
| 14,118537 | 0,395980018 | 14,40522 | 13,769041 | 14,32957 | 14,167635 | -1,3186432 |
| 14,197301 | 0,419462349 | 14,50731 | 13,83334 | 14,39939 | 14,249292 | -1,3305607 |
| 8,41541863 | 0,365183114 | 8,14344 | 8,5478515 | 8,187397 | 8,6244944 | 1,33862763 |
| 9,07312293 | 0,28640128 | 9,276302 | 8,9254103 | 9,22997 | 8,9835673 | -1,2299907 |
| 8,72458476 | 0,247701239 | 8,930006 | 8,6526364 | 8,778267 | 8,6041052 | -1,1693978 |
| 9,19623098 | 0,298460874 | 9,45707 | 8,9619835 | 9,449951 | 9,1033827 | -1,3386952 |
| 13,554781 | 0,237093171 | 13,76456 | 13,357084 | 13,71179 | 13,519498 | -1,2310464 |
| 10,6723371 | 0,25197575 | 10,52876 | 10,829015 | 10,46044 | 10,733504 | 1,21981368 |
| 13,0409464 | 0,301184837 | 13,35425 | 12,851656 | 13,12159 | 12,953403 | -1,2617178 |
| 3,18895685 | 0,225157611 | 2,968393 | 3,3287407 | 3,17552 | 3,2167578 | 1,14932981 |
| 16,5464951 | 0,311538651 | 16,37679 | 16,674509 | 16,31856 | 16,673888 | 1,25398749 |
| 13,3499727 | 0,32602728 | 13,49486 | 13,219781 | 13,619 | 13,221632 | -1,2624468 |
| 9,1217411 | 0,895335917 | 9,8905 | 9,0507518 | 8,740331 | 8,7823291 | -1,3184796 |
| 8,55396558 | 0,249310963 | 8,400536 | 8,6696387 | 8,349839 | 8,6680485 | 1,2257424 |
| 16,3129754 | 0,370953292 | 16,14859 | 16,419529 | 15,99379 | 16,516344 | 1,31653203 |
| 0,41129448 | 0,343275075 | 0,59506 | 0,2850073 | 0,769714 | 0,1915868 | -1,3604557 |
| 6,21328005 | 0,651268247 | 6,027322 | 6,68307 | 5,984283 | 5,9449948 | 1,23818763 |
| 7,92420791 | 0,30010409 | 7,76945 | 8,041763 | 7,71461 | 8,0404446 | 1,23035415 |
| 11,1484114 | 0,355125223 | 10,90052 | 11,304375 | 10,92475 | 11,306162 | 1,31278985 |
| 9,55593554 | 0,29442693 | 9,36788 | 9,7317895 | 9,413465 | 9,5902404 | 1,20609406 |
| 6,373706 | 0,348491483 | 6,631116 | 6,2222397 | 6,495366 | 6,2636709 | -1,2485778 |
| 2,25909295 | 0,254295349 | 2,535144 | 2,1151656 | 2,307648 | 2,1680294 | -1,2140253 |
| 8,23771042 | 0,314203643 | 8,428044 | 8,0665176 | 8,474231 | 8,1394463 | -1,272932 |
| 8,94866963 | 0,321491751 | 9,213261 | 8,7798418 | 9,050438 | 8,8652995 | -1,2390881 |
| 17,3034886 | 0,263914446 | 17,56608 | 17,119163 | 17,25361 | 17,331403 | -1,1364726 |
| 6,35255222 | 0,487495172 | 5,905215 | 6,6526126 | 6,271876 | 6,421122 | 1,36445208 |
| 16,1079302 | 0,357708523 | 15,90231 | 16,22598 | 15,83765 | 16,303215 | 1,31459304 |
| 10,2875847 | 0,426554155 | 10,04099 | 10,546305 | 10,00975 | 10,353424 | 1,34210026 |
| 10,1425152 | 0,23989633 | 10,27442 | 10,003216 | 10,35698 | 10,068878 | -1,2139016 |
| 17,9389109 | 0,501520782 | 17,58935 | 18,145878 | 17,58513 | 18,199941 | 1,50073166 |
| 15,8815083 | 0,416617071 | 15,60812 | 16,041973 | 15,62352 | 16,076124 | 1,35964447 |
| 3,65864483 | 0,215821194 | 3,806956 | 3,5399888 | 3,751329 | 3,6193573 | -1,1482758 |
| 9,8840298 | 0,332510583 | 9,652268 | 9,946873 | 9,729043 | 10,098608 | 1,25883162 |
| 13,437766 | 0,409324708 | 13,16636 | 13,538036 | 13,22524 | 13,675655 | 1,32965013 |
| 15,7370568 | 0,503996723 | 16,11118 | 15,435221 | 15,91959 | 15,671732 | -1,3773648 |
| 9,08100416 | 0,344930091 | 8,844813 | 9,1708021 | 8,874656 | 9,2978177 | 1,29645781 |
| 16,0674696 | 0,44859234 | 16,39196 | 15,806242 | 16,28701 | 15,973399 | -1,3657246 |
| 15,4297395 | 0,304375513 | 15,19089 | 15,57392 | 15,24802 | 15,568775 | 1,27623301 |
| 7,65256232 | 0,33897102 | 7,41058 | 7,8906261 | 7,516726 | 7,654073 | 1,23858838 |
| 6,21731177 | 0,304490568 | 6,492858 | 6,0645398 | 6,430723 | 6,0383774 | -1,3289914 |
| 5,1045473 | 0,26321105 | 4,843669 | 5,1663985 | 5,175877 | 5,2092749 | 1,13136452 |
| 1,90849837 | 0,692783362 | 1,430153 | 2,3462721 | 1,554534 | 2,0021423 | 1,604211 |
| 11,4520543 | 0,874593512 | 12,35311 | 11,209668 | 11,4272 | 10,991933 | -1,7283016 |
| 12,2831709 | 0,348138397 | 12,57743 | 12,117122 | 12,39555 | 12,164887 | -1,2705757 |
| 7,88678399 | 0,476402361 | 8,197084 | 7,5010586 | 8,091525 | 7,9630546 | -1,3307577 |
| 2,94503533 | 0,277836165 | 2,705087 | 3,0897398 | 2,883576 | 3,0122214 | 1,19470043 |
| 3,78595679 | 0,247522358 | 3,531595 | 3,9041815 | 3,838063 | 3,8290314 | 1,13428072 |
| 8,79838561 | 0,300046349 | 8,980766 | 8,596134 | 9,090888 | 8,7105631 | -1,3035791 |

| | | | | | | |
|---|---|---|---|---|---|---|
| 11,0337918 | 0,328757811 | 10,81166 | 11,160034 | 10,84902 | 11,181977 | 1,26634146 |
| 10,4697423 | 0,332132407 | 10,8088 | 10,435135 | 10,30034 | 10,324719 | -1,1286826 |
| 8,73600077 | 0,416786355 | 8,546737 | 8,9065406 | 8,433423 | 8,8737739 | 1,31957868 |
| 8,38722739 | 0,27325845 | 8,657556 | 8,2493231 | 8,369514 | 8,3335613 | -1,1664244 |
| 20,2724258 | 0,433984714 | 20,72426 | 20,055968 | 20,25009 | 20,161521 | -1,2999255 |
| 5,49702253 | 0,363423219 | 5,896517 | 5,298393 | 5,3532 | 5,4821011 | -1,1765899 |
| 12,387907 | 0,270589871 | 12,5481 | 12,228726 | 12,56153 | 12,338588 | -1,2067754 |
| 13,7694365 | 0,370967538 | 13,49319 | 13,922752 | 13,56817 | 13,941026 | 1,32061364 |
| 4,93734167 | 0,200300313 | 5,087733 | 4,8216276 | 5,032311 | 4,8913847 | -1,151501 |
| 7,10801473 | 0,443097795 | 7,359791 | 6,8995675 | 7,427512 | 6,9524433 | -1,3828514 |
| 4,66409862 | 0,208812503 | 4,837413 | 4,549271 | 4,76978 | 4,5911657 | -1,1755845 |
| 11,6156732 | 0,280495975 | 11,8695 | 11,462611 | 11,70128 | 11,532231 | -1,2209208 |
| 9,55638149 | 0,271447446 | 9,311262 | 9,6177607 | 9,464198 | 9,746388 | 1,22632751 |
| 8,83614015 | 0,303140449 | 8,866917 | 8,6018008 | 8,985376 | 9,0016454 | -1,090072 |
| 10,6380202 | 0,382940106 | 10,92857 | 10,346727 | 10,88147 | 10,594537 | -1,351337 |
| 11,5090866 | 0,259102248 | 11,64172 | 11,285292 | 11,6279 | 11,593613 | -1,1450081 |
| 15,4674323 | 0,291924462 | 15,7531 | 15,318365 | 15,52446 | 15,369275 | -1,2268525 |
| 7,76327772 | 0,43294186 | 8,038388 | 7,6388393 | 8,220293 | 7,4045505 | -1,5237707 |
| 9,70008466 | 0,415931496 | 9,958723 | 9,5681101 | 10,10339 | 9,3966293 | -1,4627529 |
| 9,54405093 | 0,243934173 | 9,350964 | 9,6426005 | 9,470548 | 9,6340174 | 1,17084745 |
| 11,3845113 | 0,278656638 | 11,08448 | 11,536757 | 11,25866 | 11,532353 | 1,28608297 |
| -0,8781057 | 0,212647002 | -0,715324 | -0,9891072 | -0,72059 | -0,977777 | -1,2020397 |
| 13,3160415 | 0,27262348 | 13,58685 | 13,133975 | 13,31969 | 13,302144 | -1,1770787 |
| 14,5271058 | 0,25128171 | 14,73314 | 14,374088 | 14,60509 | 14,488809 | -1,1790831 |
| 17,9698627 | 0,246760219 | 18,17329 | 17,772839 | 18,07045 | 17,973024 | -1,1883339 |
| 3,04061155 | 0,302300924 | 3,343823 | 2,9154845 | 2,976103 | 2,9717396 | -1,1617912 |
| 11,0278705 | 0,498352482 | 10,81506 | 11,260947 | 10,6405 | 11,161494 | 1,3980763 |
| 9,02395981 | 0,448298947 | 8,815207 | 9,2454793 | 8,637693 | 9,1673365 | 1,394703 |
| 8,2632979 | 0,273984195 | 8,159249 | 8,4794832 | 8,044722 | 8,2234632 | 1,1887852 |
| 12,3027254 | 0,2085997 | 12,48617 | 12,190947 | 12,40652 | 12,21866 | -1,1822517 |
| 12,7789966 | 0,336905169 | 12,57526 | 12,941099 | 12,57132 | 12,882249 | 1,26433585 |
| 8,32817566 | 0,467263867 | 8,791326 | 8,184619 | 8,196901 | 8,1855301 | -1,2388825 |
| 3,0873887 | 0,43735379 | 3,20746 | 2,8320969 | 3,527773 | 3,0274478 | -1,3545785 |
| 12,2133744 | 0,391174722 | 12,57823 | 12,007251 | 12,38779 | 12,045948 | -1,3721227 |
| 10,6693999 | 0,35804445 | 11,00564 | 10,484795 | 10,84687 | 10,498634 | -1,3514833 |
| 8,37123192 | 0,2155226 | 8,505984 | 8,2248822 | 8,501365 | 8,3542321 | -1,1599941 |
| 16,0023546 | 0,215693067 | 16,18874 | 15,855719 | 16,07362 | 15,977136 | -1,1605028 |
| 13,0560595 | 0,327644097 | 12,85402 | 13,230735 | 12,86757 | 13,131279 | 1,2485161 |
| 17,3796357 | 0,351656407 | 17,08981 | 17,555034 | 17,16209 | 17,546036 | 1,34218578 |
| 3,97341227 | 0,471309694 | 4,132214 | 3,736475 | 4,423615 | 3,8526336 | -1,3979962 |
| 16,2123333 | 0,370775215 | 15,95374 | 16,299482 | 15,98464 | 16,464171 | 1,33111359 |
| 16,4927985 | 0,365402793 | 16,16233 | 16,627556 | 16,26663 | 16,747692 | 1,38813364 |
| 9,68736097 | 0,23522914 | 9,445092 | 9,739718 | 9,642359 | 9,8574622 | 1,19322361 |
| 11,7481805 | 0,235098728 | 11,96498 | 11,602097 | 11,78457 | 11,717365 | -1,16074 |
| 6,55426512 | 0,270205639 | 6,76791 | 6,4411249 | 6,659994 | 6,4449981 | -1,2065522 |
| 5,3837814 | 0,455199576 | 5,816515 | 5,2657328 | 5,273943 | 5,2235189 | -1,2316594 |
| 5,69792777 | 0,307540555 | 5,389241 | 5,8401827 | 5,699433 | 5,7887027 | 1,20589624 |
| 3,8028202 | 0,452452336 | 3,360328 | 4,0864763 | 3,875076 | 3,7951972 | 1,25104564 |
| 3,40668021 | 0,266377071 | 3,170913 | 3,5219293 | 3,404715 | 3,4699626 | 1,15519151 |
| 16,3366777 | 0,401543603 | 16,12118 | 16,487558 | 15,99844 | 16,541733 | 1,37062394 |
| 14,0990568 | 0,349435495 | 13,95269 | 14,222001 | 13,78972 | 14,261544 | 1,29286563 |
| 4,21384802 | 0,204205514 | 4,412773 | 4,0811932 | 4,327553 | 4,1357247 | -1,1988939 |
| 3,86894898 | 0,282889126 | 4,089611 | 3,7894353 | 3,941116 | 3,7335023 | -1,1924218 |

| | | | | | | |
|---|---|---|---|---|---|---|
| 15,3472972 | 0,333404105 | 15,18693 | 15,546911 | 15,01814 | 15,441569 | 1,31194365 |
| 8,52781066 | 0,355115862 | 8,268368 | 8,669448 | 8,299962 | 8,7150809 | 1,32693636 |
| 10,2178431 | 0,34719895 | 9,884919 | 10,415124 | 10,15436 | 10,302182 | 1,26489085 |
| 3,92070648 | 0,238366289 | 3,699019 | 4,0096337 | 3,936723 | 3,9928186 | 1,13552177 |
| 5,7730126 | 0,323242342 | 5,510007 | 5,8878632 | 5,68002 | 5,9145426 | 1,23643782 |
| 6,4565308 | 0,261028496 | 6,269674 | 6,5932295 | 6,284706 | 6,5544154 | 1,22827383 |
| 17,8833338 | 0,545830294 | 17,59387 | 18,198833 | 17,43677 | 18,018717 | 1,50885519 |
| 3,53563446 | 0,45695479 | 3,799491 | 3,4129801 | 3,825279 | 3,2847548 | -1,3788998 |
| 3,22616793 | 0,803242016 | 2,532885 | 3,6227083 | 3,156466 | 3,3814313 | 1,57723141 |
| 12,6559349 | 0,402147331 | 12,33436 | 12,746493 | 12,53709 | 12,891913 | 1,30448459 |
| 5,56428344 | 0,241622049 | 5,78599 | 5,4483199 | 5,59672 | 5,4955267 | -1,1642752 |
| 10,5601402 | 0,320906458 | 10,85103 | 10,37919 | 10,66762 | 10,465537 | -1,2630939 |
| 9,25849657 | 0,280973855 | 9,489204 | 9,1064487 | 9,349757 | 9,1900963 | -1,2068179 |
| 11,9618236 | 0,260075187 | 12,19666 | 11,797629 | 12,07892 | 11,888989 | -1,226444 |
| 10,5763705 | 0,499390774 | 10,25829 | 10,742266 | 10,29418 | 10,816979 | 1,41753796 |
| 4,96896142 | 0,294993054 | 5,214596 | 4,8659289 | 5,022122 | 4,8519151 | -1,1970113 |
| 7,46986379 | 0,226656123 | 7,652954 | 7,3543691 | 7,53101 | 7,4161433 | -1,1540659 |
| 11,927771 | 0,394524919 | 11,62676 | 12,056956 | 11,77496 | 12,12029 | 1,30835994 |
| 12,2853966 | 0,380438801 | 11,99745 | 12,441434 | 12,04353 | 12,488021 | 1,36059135 |
| 14,0616543 | 0,465124131 | 13,72449 | 14,242653 | 13,77765 | 14,301454 | 1,43493776 |
| 8,96185524 | 0,252660453 | 8,764167 | 9,0293396 | 8,870548 | 9,103694 | 1,18851467 |
| 4,15314531 | 0,425805208 | 4,598932 | 4,0065168 | 4,254083 | 3,8896187 | -1,3932357 |
| 6,41841832 | 0,364473726 | 6,634395 | 6,2336306 | 6,718838 | 6,2763313 | -1,3394453 |
| 4,05972636 | 0,249464486 | 4,290275 | 3,9989099 | 4,085196 | 3,9214579 | -1,1708462 |
| 6,78427466 | 0,293638731 | 6,536879 | 6,8546863 | 6,678731 | 6,9733934 | 1,23647671 |
| 5,09096626 | 0,21818888 | 4,866098 | 5,1879652 | 5,175046 | 5,1152574 | 1,09508216 |
| 15,7034269 | 0,300295558 | 15,47174 | 15,913822 | 15,56231 | 15,732551 | 1,23640952 |
| 12,7583666 | 0,333217971 | 12,46422 | 13,003818 | 12,66532 | 12,769681 | 1,25004618 |
| 6,61695035 | 0,3556476 | 6,903642 | 6,5245308 | 6,71408 | 6,4258881 | -1,2601989 |
| 5,09344925 | 0,274289909 | 5,343786 | 5,0073018 | 5,187827 | 4,927413 | -1,2298218 |
| 4,97032004 | 0,312569509 | 5,167157 | 4,8433172 | 5,246671 | 4,7896012 | -1,3108066 |
| 3,40958149 | 0,337577069 | 3,636431 | 3,2396897 | 3,598552 | 3,3072472 | -1,2692913 |
| 10,0568784 | 0,276067495 | 9,91155 | 10,193989 | 9,859138 | 10,134519 | 1,21327796 |
| 19,7168983 | 0,292372834 | 19,87576 | 19,541885 | 19,96468 | 19,643218 | -1,2549797 |
| 17,708212 | 0,292099321 | 17,84687 | 17,538624 | 17,99197 | 17,623604 | -1,2642695 |
| 9,72399453 | 0,290094005 | 9,492708 | 9,8424824 | 9,619279 | 9,8412319 | 1,21913986 |
| 11,2705427 | 0,491512282 | 10,96306 | 11,464895 | 11,03326 | 11,441044 | 1,37059882 |
| 9,11260083 | 0,308525598 | 9,425191 | 8,9860114 | 9,109434 | 9,0007066 | -1,2091166 |
| 16,2770753 | 0,458250953 | 16,32386 | 15,899784 | 16,66643 | 16,456441 | -1,2457687 |
| 17,0884414 | 0,465367221 | 16,81378 | 17,291735 | 16,74666 | 17,283025 | 1,42125343 |
| 5,67645761 | 0,246826076 | 5,862654 | 5,5691428 | 5,783268 | 5,5828821 | -1,1866945 |
| 15,8308669 | 0,313492484 | 16,105 | 15,582133 | 15,96867 | 15,81365 | -1,2648323 |
| 14,5314039 | 0,246305737 | 14,66364 | 14,38859 | 14,73235 | 14,469813 | -1,2047998 |
| 1,01730953 | 0,210232881 | 0,810026 | 1,0761106 | 1,103032 | 1,0715051 | 1,08468671 |
| 19,4448214 | 0,616872651 | 19,2296 | 19,749284 | 18,73468 | 19,688486 | 1,66640925 |
| 19,1092187 | 0,631348155 | 18,88182 | 19,418347 | 18,40838 | 19,352055 | 1,67029045 |
| 11,1787887 | 0,197236309 | 11,40291 | 11,109931 | 11,10823 | 11,113254 | -1,1049454 |
| 19,4159608 | 0,411581258 | 19,83719 | 19,386978 | 19,11969 | 19,270457 | -1,1093561 |
| 11,8635167 | 0,297369452 | 12,07551 | 11,748283 | 11,99664 | 11,741731 | -1,2235445 |
| 2,75089273 | 0,18899391 | 2,587884 | 2,8611715 | 2,710163 | 2,7815537 | 1,12688422 |
| 9,22673533 | 0,422040405 | 9,494596 | 9,0874066 | 9,43485 | 9,0413789 | -1,3198103 |
| 10,7681626 | 0,31548905 | 11,04827 | 10,665943 | 10,86563 | 10,59425 | -1,2542781 |
| 16,3395549 | 0,447657658 | 16,01476 | 16,56146 | 16,10451 | 16,490374 | 1,38154628 |

| | | | | | | |
|---|---|---|---|---|---|---|
| 8,77171293 | 0,369529139 | 8,776472 | 8,45131 | 9,107335 | 8,9507782 | -1,1816963 |
| 7,94367375 | 0,502513822 | 8,37729 | 7,771519 | 7,99189 | 7,7527563 | -1,3402035 |
| 13,7126806 | 0,361940237 | 13,50817 | 13,848505 | 13,394 | 13,91473 | 1,34772939 |
| 2,93815551 | 0,194144475 | 2,769429 | 3,0512044 | 2,909612 | 2,9630401 | 1,1231896 |
| 10,4330642 | 0,767009192 | 9,69782 | 10,765517 | 10,36477 | 10,700056 | 1,62618641 |
| 3,96600478 | 0,379594939 | 4,346323 | 3,8819053 | 3,999701 | 3,7234356 | -1,292659 |
| 5,96917615 | 0,37966487 | 6,311948 | 5,8939881 | 5,919963 | 5,7975739 | -1,2059535 |
| 15,7005267 | 0,217447809 | 15,92193 | 15,604466 | 15,59838 | 15,688892 | -1,0818328 |
| 5,83944184 | 0,607552356 | 6,28765 | 5,5640679 | 6,069734 | 5,6507382 | -1,4858506 |
| 8,6579039 | 0,245277281 | 8,869292 | 8,5550256 | 8,773145 | 8,5325334 | -1,2120413 |
| 7,48255069 | 0,341438432 | 7,293408 | 7,6562746 | 7,229811 | 7,586497 | 1,28322692 |
| 7,5727598 | 0,360150783 | 7,363264 | 7,6571412 | 7,317942 | 7,8024641 | 1,30966651 |
| 7,35452118 | 0,251235362 | 7,178833 | 7,4804484 | 7,192673 | 7,4498523 | 1,21368776 |
| 14,514983 | 0,360603702 | 14,30917 | 14,712949 | 14,28027 | 14,59319 | 1,28195714 |
| 9,69990844 | 0,377728478 | 9,473959 | 9,8472988 | 9,490316 | 9,8408521 | 1,28515082 |
| 6,32016925 | 0,339578865 | 6,635463 | 6,1080759 | 6,456191 | 6,2250688 | -1,3006696 |
| 14,6451912 | 0,253352787 | 14,87971 | 14,527206 | 14,73393 | 14,53419 | -1,2109353 |
| 0,98500856 | 0,294317407 | 0,883813 | 1,2523199 | 0,85718 | 0,8269489 | 1,12438673 |
| 9,92047013 | 0,635021735 | 9,611159 | 10,173518 | 9,466457 | 10,152135 | 1,54116158 |
| 9,02738721 | 0,591720995 | 8,775035 | 9,2622087 | 8,581182 | 9,2278236 | 1,48134467 |
| 7,03183025 | 0,409182596 | 7,22946 | 6,7946228 | 7,358884 | 6,9522617 | -1,3386044 |
| 1,81747033 | 0,480620542 | 2,167599 | 1,5374862 | 1,98994 | 1,7523598 | -1,3508305 |
| 5,92882486 | 0,259816492 | 5,685636 | 6,0563666 | 5,85989 | 6,0238458 | 1,20358909 |
| 2,70242767 | 0,194502687 | 2,904488 | 2,6395122 | 2,691035 | 2,6130105 | -1,126229 |
| 24,9593743 | 0,385983549 | 25,37884 | 24,659341 | 24,92682 | 24,982405 | -1,2587175 |
| 10,4315372 | 0,268342268 | 10,20259 | 10,533622 | 10,37956 | 10,534828 | 1,18357526 |
| 8,08406854 | 0,306347369 | 8,277456 | 7,9590772 | 8,257775 | 7,9654548 | -1,2357181 |
| 8,01582354 | 0,288005828 | 7,742263 | 8,0854078 | 8,207835 | 8,0496421 | 1,06619852 |
| 2,75515347 | 0,401132355 | 2,44345 | 3,0165765 | 2,607224 | 2,7951519 | 1,30181785 |
| 2,74977645 | 0,384548954 | 2,426995 | 2,9853247 | 2,61541 | 2,8221028 | 1,30360902 |
| 13,1522155 | 0,259420611 | 12,99191 | 13,269363 | 12,99282 | 13,243532 | 1,20087047 |
| 11,659771 | 0,331058841 | 11,42098 | 11,781876 | 11,50348 | 11,80999 | 1,26024286 |
| 6,67021071 | 0,223897957 | 6,460705 | 6,8019866 | 6,579959 | 6,7443107 | 1,19153118 |
| 5,34585275 | 0,264786322 | 5,195633 | 5,467922 | 5,159603 | 5,4388059 | 1,21061972 |
| 5,17810569 | 0,317214477 | 4,957381 | 5,3515816 | 4,987927 | 5,271658 | 1,26484968 |
| 5,57268358 | 0,245706201 | 5,390131 | 5,7158832 | 5,48391 | 5,6092779 | 1,16923097 |
| 10,0585968 | 0,33207809 | 9,816009 | 10,275376 | 9,901877 | 10,098693 | 1,25535145 |
| 5,52797096 | 0,370846119 | 5,187196 | 5,6163639 | 5,561937 | 5,6911781 | 1,21352527 |
| 14,9816557 | 0,444500014 | 14,67238 | 15,07164 | 14,73347 | 15,285472 | 1,39052758 |
| 9,66409917 | 0,310108372 | 9,416926 | 9,8443712 | 9,504575 | 9,7535847 | 1,2642025 |
| 0,97633863 | 0,474506052 | 0,776892 | 1,4094249 | 0,671719 | 0,8089362 | 1,30574655 |
| 4,28431581 | 0,308237286 | 4,010595 | 4,4249572 | 4,13471 | 4,4379722 | 1,28236962 |
| 2,89255989 | 0,280895092 | 2,607396 | 2,9906 | 2,894533 | 3,0161171 | 1,19118199 |
| 4,07212788 | 0,249938777 | 3,84192 | 4,1458962 | 4,10138 | 4,1617317 | 1,13458456 |
| 8,29047764 | 0,279216088 | 8,057857 | 8,3987572 | 8,202787 | 8,4108839 | 1,20957348 |
| 3,39113592 | 0,268454112 | 3,633758 | 3,2126072 | 3,544378 | 3,3071957 | -1,2562878 |
| 17,5041352 | 0,653643894 | 17,04071 | 17,619517 | 17,2381 | 17,91921 | 1,54752085 |
| 16,5578025 | 0,420890599 | 16,26105 | 16,724862 | 16,27138 | 16,781422 | 1,40145449 |
| 13,3630131 | 0,340874127 | 13,12868 | 13,518731 | 13,10371 | 13,530921 | 1,32742763 |
| 6,59655545 | 0,436021282 | 6,449892 | 6,7772008 | 6,220814 | 6,7298896 | 1,33625185 |
| 16,8425262 | 0,380505295 | 16,55188 | 16,960849 | 16,67271 | 17,049478 | 1,31300238 |
| 11,4371029 | 0,376178728 | 11,70221 | 11,265027 | 11,58023 | 11,332376 | -1,267969 |
| 3,03757544 | 0,246247308 | 2,810755 | 3,1702522 | 2,995032 | 3,0966872 | 1,17330365 |

| | | | | | | |
|---|---|---|---|---|---|---|
| 9,85183611 | 0,345164545 | 10,07591 | 9,6579681 | 10,02485 | 9,7902024 | -1,2537918 |
| 4,02573858 | 0,293408128 | 3,84779 | 4,1631245 | 3,841307 | 4,1227907 | 1,22978754 |
| 9,42030545 | 0,249690828 | 9,576051 | 9,2853846 | 9,553297 | 9,3700314 | -1,1785119 |
| 5,56498609 | 0,42727365 | 5,196259 | 5,7106692 | 5,455534 | 5,7692557 | 1,33243574 |
| 3,35630228 | 0,334036549 | 3,047084 | 3,584748 | 3,211579 | 3,4318375 | 1,30040562 |
| 2,67848019 | 0,206623214 | 2,856605 | 2,5878768 | 2,723072 | 2,6090429 | -1,1418545 |
| 0,49182682 | 0,256534078 | 0,688594 | 0,362455 | 0,639245 | 0,3913696 | -1,2201069 |
| 6,46858851 | 0,352208347 | 6,173194 | 6,6048786 | 6,387512 | 6,6047715 | 1,25220609 |
| 16,3942729 | 0,395992655 | 16,75906 | 16,120003 | 16,52337 | 16,335871 | -1,3317075 |
| 3,55609108 | 0,398153262 | 3,209404 | 3,697039 | 3,578504 | 3,6681899 | 1,2215056 |
| 8,65384437 | 0,241708952 | 8,42431 | 8,6875399 | 8,713623 | 8,7726461 | 1,11815957 |
| 18,2241649 | 0,39150318 | 17,95651 | 18,406453 | 17,97241 | 18,383978 | 1,34793866 |
| 9,00960678 | 0,260398346 | 8,752374 | 9,1072087 | 9,049031 | 9,0874781 | 1,146027 |
| 11,9401923 | 0,245037149 | 12,10548 | 11,802286 | 12,12731 | 11,852912 | -1,2216216 |
| 11,514777 | 0,40087396 | 11,22931 | 11,692565 | 11,36248 | 11,635452 | 1,29066126 |
| 6,77241051 | 0,417065842 | 6,478186 | 6,9932184 | 6,61454 | 6,8522542 | 1,29807459 |
| 3,80863716 | 0,200631783 | 4,00369 | 3,7090208 | 3,894805 | 3,7106811 | -1,1804989 |
| 8,36787068 | 0,234833933 | 8,565634 | 8,2150065 | 8,476366 | 8,3181115 | -1,1928735 |
| 19,876794 | 0,440117413 | 19,74417 | 20,025627 | 19,37336 | 20,112985 | 1,42458479 |
| 10,8078243 | 0,3528504 | 10,60242 | 10,889878 | 10,5748 | 11,023764 | 1,29074781 |
| 4,16514516 | 0,296506484 | 4,362377 | 4,0444088 | 4,29052 | 4,0671569 | -1,2063643 |
| 11,9974504 | 0,337542195 | 11,69918 | 12,115303 | 11,73837 | 12,265009 | 1,38643798 |
| 15,565044 | 0,313303761 | 15,34756 | 15,737995 | 15,35809 | 15,666538 | 1,27406799 |
| 6,12735194 | 0,236401172 | 5,953474 | 6,2555839 | 6,018965 | 6,1863018 | 1,17668094 |
| 17,4221094 | 0,344612465 | 17,22958 | 17,549596 | 17,14455 | 17,599308 | 1,30802103 |
| 9,72435784 | 0,345931028 | 9,353604 | 9,9604117 | 9,567213 | 9,850521 | 1,36136879 |
| 13,3938738 | 0,188933423 | 13,58589 | 13,286304 | 13,44842 | 13,327019 | -1,1570826 |
| 9,11454557 | 0,244965464 | 9,33489 | 8,94882 | 9,247506 | 9,0463474 | -1,225707 |
| 8,59907382 | 0,528426738 | 9,066888 | 8,4233362 | 8,690754 | 8,3573089 | -1,4029839 |
| 2,58073029 | 0,661137027 | 3,050786 | 2,2442616 | 3,095221 | 2,2762509 | -1,7565531 |
| 9,51807382 | 0,221089601 | 9,753199 | 9,3628811 | 9,619928 | 9,4433359 | -1,2171063 |
| 11,311599 | 0,317857781 | 11,6364 | 11,112984 | 11,4077 | 11,216201 | -1,2811621 |
| 14,3085507 | 0,36631302 | 14,09286 | 14,583687 | 14,00034 | 14,346651 | 1,33660157 |
| 10,2066812 | 0,23154351 | 10,40054 | 10,051088 | 10,31683 | 10,162519 | -1,1907616 |
| 9,67892894 | 0,274461277 | 9,525653 | 9,7938183 | 9,420568 | 9,8263624 | 1,26310931 |
| 16,5194257 | 0,45738821 | 16,3409 | 16,652603 | 16,02097 | 16,810435 | 1,46468058 |
| 8,15259978 | 0,551234853 | 8,685048 | 8,0939951 | 7,902013 | 7,9206962 | -1,2194115 |
| 8,83538982 | 0,272800904 | 8,589223 | 8,8904544 | 8,821005 | 8,9871852 | 1,17585148 |
| 2,15726226 | 0,19758501 | 2,328958 | 2,0234617 | 2,29888 | 2,0869112 | -1,196427 |
| -1,34140547 | 0,239304169 | -1,565509 | -1,2779771 | -1,16398 | -1,333488 | 1,04175088 |
| 12,0803183 | 0,342421331 | 11,91436 | 12,177836 | 11,82822 | 12,255616 | 1,27053259 |
| 17,5407217 | 0,334122685 | 17,40001 | 17,683977 | 17,19379 | 17,696583 | 1,31346925 |
| 4,13434695 | 0,405522503 | 4,338921 | 4,0416193 | 4,400767 | 3,9118801 | -1,3132069 |
| 13,1774156 | 0,390864935 | 12,95196 | 13,286958 | 12,92265 | 13,390466 | 1,32079855 |
| 0,97833491 | 0,2136121 | 1,19387 | 0,873237 | 1,0964 | 0,850408 | -1,2169862 |
| 18,8370377 | 0,409390117 | 18,60828 | 19,05875 | 18,45417 | 18,995147 | 1,41002769 |
| 12,2765156 | 0,23719806 | 12,12359 | 12,400543 | 12,13447 | 12,342893 | 1,18319281 |
| 13,0924673 | 0,422354772 | 12,73569 | 13,163485 | 12,8923 | 13,430608 | 1,39769775 |
| 10,0133137 | 0,239656034 | 9,78346 | 10,061439 | 9,996997 | 10,160729 | 1,1654246 |
| 8,80842632 | 0,270302202 | 8,959214 | 8,6821646 | 9,026398 | 8,7009876 | -1,2321946 |
| -0,40800432 | 0,309217486 | -0,722388 | -0,3137079 | -0,328 | -0,301936 | 1,16261367 |
| 1,86188843 | 0,224123586 | 1,667873 | 2,0011414 | 1,735971 | 1,9352483 | 1,20269664 |
| 4,72448375 | 0,214149438 | 4,86926 | 4,5921304 | 4,881654 | 4,6659458 | -1,1862589 |

| | | | | | | |
|---|---|---|---|---|---|---|
| 17,5375367 | 0,288931752 | 17,30823 | 17,687603 | 17,40882 | 17,6296 | 1,23121146 |
| 16,845552 | 0,347448456 | 17,10346 | 16,554215 | 17,10011 | 16,823197 | -1,3315265 |
| 7,46216892 | 0,283427267 | 7,251186 | 7,5745386 | 7,336985 | 7,5817713 | 1,21762461 |
| 5,76879172 | 0,350391396 | 5,43139 | 5,972552 | 5,760841 | 5,815841 | 1,22950769 |
| 14,2793049 | 0,316165257 | 14,52743 | 14,091062 | 14,45307 | 14,19003 | -1,2742995 |
| 7,17785185 | 0,188503801 | 7,288946 | 7,0200569 | 7,355247 | 7,1663392 | -1,1719395 |
| 13,4919801 | 0,309636042 | 13,75646 | 13,261034 | 13,65999 | 13,443506 | -1,2798305 |
| 3,63019979 | 0,340205302 | 3,372396 | 3,8537938 | 3,355336 | 3,7459383 | 1,3528482 |
| 14,899729 | 0,386598693 | 14,65161 | 14,916231 | 14,68763 | 15,218086 | 1,31725784 |
| 4,97690469 | 0,329685688 | 4,703754 | 5,0460335 | 4,892548 | 5,1767427 | 1,24249275 |
| 7,72959033 | 0,258519379 | 7,978607 | 7,5800242 | 7,756686 | 7,681148 | -1,1785888 |
| 12,8733943 | 0,267323322 | 13,10444 | 12,711377 | 12,95206 | 12,824221 | -1,1978554 |
| 6,36857458 | 0,435143668 | 6,546694 | 6,1784422 | 6,689398 | 6,2528382 | -1,3217099 |
| 13,4373855 | 0,346470081 | 13,21566 | 13,54023 | 13,21942 | 13,633219 | 1,29162251 |
| 9,64092752 | 0,180525726 | 9,756601 | 9,4926383 | 9,782138 | 9,6358257 | -1,1527963 |
| 16,1604101 | 0,298988001 | 15,98776 | 16,300502 | 15,85096 | 16,324722 | 1,31335063 |
| 6,21921724 | 0,272381528 | 6,402698 | 6,0727941 | 6,400268 | 6,130336 | -1,2310743 |
| 4,85724878 | 0,411794068 | 5,134634 | 4,5685106 | 5,287508 | 4,7098015 | -1,4864954 |
| 5,5813624 | 0,261945722 | 5,762693 | 5,4035945 | 5,76983 | 5,5274719 | -1,2317663 |
| 17,0425433 | 0,595945198 | 16,70095 | 17,412107 | 16,57528 | 17,169773 | 1,57223985 |
| 8,30259245 | 0,355857356 | 8,148896 | 8,4527142 | 8,035885 | 8,4131131 | 1,26621583 |
| 15,7354285 | 0,341292854 | 15,54792 | 15,839648 | 15,38587 | 15,979481 | 1,35911706 |
| 11,4450085 | 0,328180545 | 11,53567 | 11,1932 | 11,66929 | 11,535553 | -1,1794382 |
| 14,5168974 | 0,270092027 | 14,29622 | 14,655283 | 14,36352 | 14,63044 | 1,2422821 |
| 1,80563802 | 0,460786864 | 1,415948 | 1,900892 | 1,743664 | 2,0597538 | 1,31998048 |
| 6,41066022 | 0,387807215 | 6,6144 | 6,1191648 | 6,552119 | 6,5024007 | -1,2078797 |
| 11,015342 | 0,346152277 | 11,33475 | 10,811218 | 11,10389 | 10,935661 | -1,2709288 |
| 1,00510781 | 0,22021676 | 1,163474 | 0,899425 | 1,158791 | 0,9051055 | -1,196539 |
| 11,834722 | 0,592686122 | 11,52285 | 12,028383 | 11,36985 | 12,146349 | 1,55943137 |
| 9,06603161 | 0,263304058 | 9,245453 | 8,9407844 | 9,221422 | 8,970586 | -1,2123046 |
| 6,16127586 | 0,172764689 | 6,318125 | 6,0519903 | 6,218007 | 6,1250578 | -1,1325245 |
| 11,1965661 | 0,261868575 | 11,36925 | 11,013231 | 11,40029 | 11,147549 | -1,2348894 |
| 2,8652108 | 0,37837728 | 3,219569 | 2,812774 | 2,799311 | 2,6664702 | -1,2056557 |
| 4,18185699 | 0,281092227 | 4,456736 | 4,1304945 | 4,241161 | 3,9742623 | -1,228221 |
| 6,2197494 | 0,284236599 | 6,452809 | 6,0851027 | 6,361964 | 6,0978959 | -1,2447769 |
| 6,21254564 | 0,230708041 | 6,028412 | 6,3336726 | 6,066116 | 6,3115645 | 1,21029137 |
| 8,0843426 | 0,227093105 | 7,869429 | 8,1996524 | 8,043864 | 8,1529348 | 1,16444881 |
| -0,40973083 | 0,373941209 | -0,784504 | -0,2631826 | -0,32966 | -0,315074 | 1,20409929 |
| 7,29247005 | 0,311893665 | 7,565647 | 7,144732 | 7,369562 | 7,1913 | -1,2307933 |
| 15,0510254 | 0,293062884 | 15,33948 | 14,852323 | 15,19149 | 14,959998 | -1,2828283 |
| 15,3690433 | 0,333405386 | 15,17684 | 15,464141 | 15,15126 | 15,54897 | 1,26795787 |
| 19,4916767 | 0,287294634 | 19,7985 | 19,291876 | 19,40227 | 19,524281 | -1,1425891 |
| 11,4680162 | 0,283599305 | 11,28146 | 11,586952 | 11,30853 | 11,57956 | 1,22116844 |
| 14,288816 | 0,327898297 | 14,122 | 14,429446 | 14,0355 | 14,413843 | 1,26829826 |
| 7,36402896 | 0,408302731 | 6,977471 | 7,4589356 | 7,450471 | 7,5268501 | 1,21328799 |
| 7,96014058 | 0,388170875 | 7,586927 | 8,1272301 | 7,999144 | 8,0534633 | 1,22885232 |
| 14,5463145 | 0,368533133 | 14,40494 | 14,688942 | 14,22147 | 14,690238 | 1,29808599 |
| 9,26708458 | 0,229543453 | 9,113696 | 9,3926612 | 9,106293 | 9,3432478 | 1,19578666 |
| 9,35882082 | 0,269822206 | 9,600827 | 9,2372272 | 9,393873 | 9,2779966 | -1,1807782 |
| 7,48151986 | 0,310285884 | 7,190508 | 7,6528286 | 7,493478 | 7,5161343 | 1,18303134 |
| 11,4928722 | 0,316540207 | 11,28362 | 11,598875 | 11,30484 | 11,656354 | 1,25996578 |
| 16,8863414 | 0,391948889 | 16,70594 | 16,997177 | 16,56866 | 17,097287 | 1,32862315 |
| 9,9671446 | 0,26484871 | 10,18091 | 9,8545972 | 10,18475 | 9,7899403 | -1,2839238 |

| | | | | | | |
|---|---|---|---|---|---|---|
| 12,3992316 | 0,183132072 | 12,53604 | 12,261786 | 12,54557 | 12,360078 | -1,1727298 |
| 5,89394618 | 0,33490814 | 6,135045 | 5,7258428 | 6,059948 | 5,7911355 | -1,2648857 |
| 4,51998627 | 0,324958728 | 4,815962 | 4,3526266 | 4,596465 | 4,4233514 | -1,2467952 |
| 20,913855 | 0,421678482 | 21,27565 | 20,611039 | 21,03209 | 20,89877 | -1,3185608 |
| 21,6720507 | 0,437108807 | 22,06665 | 21,342536 | 21,83541 | 21,634042 | -1,3781593 |
| 2,0482103 | 0,204852717 | 2,208079 | 1,9199484 | 2,222375 | 1,9617372 | -1,2094779 |
| 10,480161 | 0,317182821 | 10,70959 | 10,273446 | 10,68361 | 10,411136 | -1,2783725 |
| 10,5735149 | 0,207449479 | 10,75188 | 10,448085 | 10,66135 | 10,519721 | -1,166924 |
| 9,95812001 | 0,261900085 | 10,15488 | 9,7840772 | 10,14255 | 9,8890699 | -1,2415482 |
| 8,4696652 | 0,268606609 | 8,643357 | 8,3643951 | 8,675872 | 8,3246272 | -1,2441008 |
| 12,0944409 | 0,213046613 | 12,21909 | 11,928312 | 12,26324 | 12,086564 | -1,1758691 |
| 10,1719755 | 0,185745354 | 10,32345 | 10,048473 | 10,25342 | 10,142558 | -1,1430743 |
| 8,41779796 | 0,223135779 | 8,56964 | 8,3017673 | 8,568497 | 8,3375499 | -1,1887206 |
| 9,47123119 | 0,300737762 | 9,685193 | 9,2911712 | 9,684422 | 9,3775211 | -1,2749683 |
| 4,87294313 | 0,264680713 | 5,10993 | 4,7554147 | 4,895932 | 4,7987452 | -1,1694668 |
| 13,5881026 | 0,224176008 | 13,75964 | 13,404524 | 13,71325 | 13,587499 | -1,1813487 |
| 9,33398238 | 0,198424452 | 9,522243 | 9,1946958 | 9,470475 | 9,2592091 | -1,2053119 |
| 10,4358276 | 0,267325812 | 10,61584 | 10,26256 | 10,65059 | 10,361881 | -1,2491905 |
| 3,87736561 | 0,265873828 | 3,620734 | 4,0098552 | 3,846431 | 3,9550752 | 1,18828612 |
| 12,1337329 | 0,309465389 | 12,3156 | 11,953201 | 12,35278 | 12,064358 | -1,25302 |
| 18,007343 | 0,520407641 | 18,52224 | 17,564961 | 18,17538 | 17,999717 | -1,4808973 |
| 9,83769903 | 0,272463915 | 10,05611 | 9,697461 | 9,966151 | 9,7432656 | -1,2232899 |
| 19,4170461 | 0,376522917 | 19,56699 | 19,075834 | 19,60062 | 19,588902 | -1,1903932 |
| 6,59580847 | 0,338174814 | 6,414844 | 6,6880418 | 6,367469 | 6,7759521 | 1,2664943 |
| 15,1445774 | 0,575136898 | 14,75374 | 15,455937 | 14,7121 | 15,362645 | 1,59811506 |
| 12,8016467 | 0,219873124 | 12,98994 | 12,716982 | 12,92041 | 12,671935 | -1,1980739 |
| 9,43172757 | 0,338402108 | 9,218613 | 9,5860381 | 9,245092 | 9,5396836 | 1,25789218 |
| 10,4614504 | 0,301453142 | 10,63854 | 10,370478 | 10,67667 | 10,290959 | -1,2543027 |
| 9,26452784 | 0,346495313 | 9,528366 | 9,040775 | 9,487473 | 9,1750015 | -1,3195367 |
| 10,6722278 | 0,353811939 | 10,95073 | 10,438744 | 10,84642 | 10,611165 | -1,2956002 |
| 13,355997 | 0,710311144 | 13,82132 | 12,896776 | 13,78915 | 13,25165 | -1,6598119 |
| 15,403996 | 0,309945422 | 15,17315 | 15,540103 | 15,23765 | 15,536695 | 1,25962891 |
| 8,80354591 | 0,244712278 | 9,015828 | 8,6513114 | 8,896309 | 8,7501297 | -1,1936236 |
| 9,38136141 | 0,553601577 | 9,207843 | 9,7476125 | 8,812012 | 9,4309602 | 1,49418495 |
| 18,8675414 | 0,398230087 | 18,69972 | 18,978007 | 18,49349 | 19,102061 | 1,35983163 |
| 4,73457006 | 0,190406284 | 4,862611 | 4,5884767 | 4,867399 | 4,7213498 | -1,1567617 |
| 1,72639757 | 0,376643807 | 1,382017 | 1,7983516 | 1,831822 | 1,8695219 | 1,17041282 |
| 3,56476845 | 0,199093333 | 3,724862 | 3,44716 | 3,669773 | 3,506816 | -1,1649999 |
| 4,18123775 | 0,251971835 | 3,951079 | 4,2980719 | 4,112989 | 4,2776206 | 1,1940076 |
| 13,4373855 | 0,346470081 | 13,21566 | 13,54023 | 13,21942 | 13,633219 | 1,29162251 |
| 8,29254334 | 0,234962632 | 8,493205 | 8,1383022 | 8,450721 | 8,2121638 | -1,228357 |
| 5,49211074 | 0,463311662 | 5,156968 | 5,7024408 | 5,354873 | 5,6069281 | 1,31837751 |
| 3,87012018 | 0,23941005 | 4,084734 | 3,7673359 | 3,873512 | 3,8090039 | -1,1415179 |
| 5,74306768 | 0,257527879 | 5,49611 | 5,8165458 | 5,829019 | 5,8132371 | 1,11136057 |
| 8,39957679 | 0,325036981 | 8,607012 | 8,2675995 | 8,597526 | 8,2628597 | -1,2631619 |
| 10,8531542 | 0,280492272 | 10,64814 | 10,958446 | 10,66585 | 11,01345 | 1,25610271 |
| 13,0808916 | 0,237150003 | 13,27046 | 12,994535 | 13,16248 | 12,974428 | -1,174456 |
| 9,43646253 | 0,291200741 | 9,694606 | 9,2153692 | 9,65245 | 9,3527599 | -1,3099062 |
| 10,0614577 | 0,284222053 | 10,33712 | 9,8222969 | 10,25057 | 10,00067 | -1,3034746 |
| 12,761219 | 0,239063258 | 12,98836 | 12,555413 | 12,8875 | 12,73935 | -1,2231043 |
| 9,949335 | 0,287210417 | 10,24028 | 9,8599334 | 9,880766 | 9,850451 | -1,1529526 |
| 16,0044029 | 0,295196229 | 15,78475 | 16,098517 | 15,84468 | 16,174006 | 1,24966895 |
| 6,77777797 | 0,2712571 | 6,632416 | 6,9104274 | 6,54601 | 6,8812171 | 1,23679744 |

| | | | | | | |
|---|---|---|---|---|---|---|
| 3,82241533 | 0,262231307 | 3,57622 | 3,8481861 | 4,023751 | 3,879955 | 1,04542169 |
| 16,3462423 | 0,459033126 | 16,07277 | 16,458601 | 16,01269 | 16,643996 | 1,42263921 |
| 19,6849992 | 0,450023141 | 19,52175 | 19,848604 | 19,21173 | 19,9114 | 1,42727581 |
| 9,67230756 | 0,21625341 | 9,836908 | 9,5429496 | 9,854575 | 9,5782663 | -1,2185231 |
| 6,48063407 | 0,330911144 | 6,665344 | 6,3191365 | 6,760869 | 6,3492865 | -1,3003457 |
| 5,89743033 | 0,392937726 | 6,186975 | 5,8472011 | 6,036701 | 5,6280302 | -1,2961406 |
| 5,91328146 | 0,372713862 | 6,179227 | 5,8687998 | 6,036065 | 5,666936 | -1,2655618 |
| 14,1274707 | 0,282103399 | 14,38296 | 13,934196 | 14,22303 | 14,084893 | -1,2255709 |
| 9,78547651 | 0,26608373 | 9,947569 | 9,6464376 | 9,949603 | 9,7160689 | -1,2035804 |
| 7,94969455 | 0,274978166 | 7,779277 | 8,0864299 | 7,796757 | 8,0222299 | 1,20273026 |
| 9,12994542 | 0,450715168 | 9,532123 | 8,9874324 | 9,004752 | 9,0342259 | -1,1954953 |
| 4,94055174 | 0,271121969 | 5,182592 | 4,8411383 | 5,027843 | 4,8017388 | -1,2173796 |
| 10,5357788 | 0,33763519 | 10,36414 | 10,676147 | 10,29755 | 10,656162 | 1,26164449 |
| 6,03572834 | 0,256811849 | 6,194222 | 5,8992716 | 6,183095 | 5,9763369 | -1,1899114 |
| 4,62288236 | 0,237956053 | 4,823377 | 4,4724279 | 4,799705 | 4,5269136 | -1,2413159 |
| 5,72437543 | 0,244658182 | 5,930201 | 5,6044839 | 5,830626 | 5,629543 | -1,2003042 |
| 12,4798524 | 0,250892426 | 12,69374 | 12,359402 | 12,56615 | 12,390804 | -1,1932083 |
| 13,5674149 | 0,258935206 | 13,82047 | 13,510357 | 13,44089 | 13,496702 | -1,0921342 |
| 12,5232446 | 0,28244729 | 12,33728 | 12,653302 | 12,35382 | 12,626899 | 1,22650163 |
| 22,5606024 | 0,525761564 | 22,38023 | 22,818155 | 22,00678 | 22,737145 | 1,49914664 |
| 8,55341779 | 0,395139493 | 8,238425 | 8,7330968 | 8,429552 | 8,679866 | 1,29458767 |
| 6,28261743 | 0,264700243 | 6,478772 | 6,1239246 | 6,428144 | 6,2190009 | -1,2158756 |
| 16,8395819 | 0,319651463 | 17,17212 | 16,729171 | 16,71728 | 16,7628 | -1,1476753 |
| 17,788895 | 0,299135812 | 18,08738 | 17,583358 | 17,75131 | 17,804384 | -1,1691585 |
| 12,247288 | 0,244176377 | 12,484 | 12,095009 | 12,27461 | 12,212424 | -1,1692536 |
| 5,30879654 | 0,230276804 | 5,453397 | 5,1446509 | 5,469623 | 5,2863646 | -1,1859165 |
| -0,77023352 | 0,290442372 | -0,94704 | -0,5390284 | -0,92189 | -0,8064 | 1,19893308 |
| 15,6557188 | 0,611055685 | 15,93051 | 15,168543 | 16,05753 | 15,76567 | -1,4408446 |
| 8,94000582 | 0,243470522 | 9,178045 | 8,7988766 | 8,980754 | 8,8825788 | -1,1799059 |
| 19,9061906 | 0,366340144 | 19,69175 | 19,966718 | 19,60293 | 20,197788 | 1,35182914 |
| 7,48054235 | 0,39780617 | 7,863613 | 7,4132285 | 7,337744 | 7,3213874 | -1,1755786 |
| 9,03647721 | 0,263933171 | 8,873656 | 9,1577456 | 8,884803 | 9,120358 | 1,19733136 |
| 10,2161415 | 0,218206185 | 10,46272 | 10,110228 | 10,21735 | 10,13292 | -1,1634913 |
| 7,21118975 | 0,312778531 | 6,903278 | 7,26572 | 7,258397 | 7,3791544 | 1,18230307 |
| 5,30081819 | 0,26208397 | 5,105822 | 5,4462347 | 5,071971 | 5,4293733 | 1,27359577 |
| 12,2012005 | 0,269884225 | 12,38945 | 12,010546 | 12,43213 | 12,131418 | -1,2655877 |
| 15,9971606 | 0,361372289 | 15,79116 | 16,151505 | 15,6679 | 16,184608 | 1,35522269 |
| 11,57964 | 0,227619593 | 11,83618 | 11,441918 | 11,61629 | 11,504856 | -1,1915577 |
| 12,0462322 | 0,215841664 | 12,20692 | 11,930001 | 12,1347 | 11,996049 | -1,1549109 |
| 8,41114164 | 0,254640236 | 8,632823 | 8,2332027 | 8,590222 | 8,3287909 | -1,2574715 |
| 5,18026319 | 0,290401486 | 5,377459 | 5,0337576 | 5,345135 | 5,0895301 | -1,2308485 |
| 4,48275017 | 0,303330934 | 4,691334 | 4,3298204 | 4,666141 | 4,3789354 | -1,2521083 |
| 10,6047484 | 0,215812906 | 10,80974 | 10,527094 | 10,64838 | 10,497513 | -1,1621183 |
| 13,7881714 | 0,260697925 | 14,01221 | 13,586659 | 13,86477 | 13,79359 | -1,1878629 |
| 9,31412616 | 0,316160377 | 9,531887 | 9,1012806 | 9,468088 | 9,2920669 | -1,2339756 |
| 2,86818953 | 0,279283693 | 3,082315 | 2,7363038 | 2,979301 | 2,777779 | -1,2089599 |
| 6,56739801 | 0,420851184 | 6,862361 | 6,3591213 | 6,706974 | 6,4828657 | -1,2866986 |
| 6,45064476 | 0,205172132 | 6,318228 | 6,5850292 | 6,351201 | 6,4616041 | 1,13965897 |
| 12,6673659 | 0,32255233 | 12,93292 | 12,407759 | 12,87005 | 12,63156 | -1,3029913 |
| 1,11166164 | 0,26586916 | 0,851617 | 1,2035123 | 1,097287 | 1,2311036 | 1,18333292 |
| 6,46338857 | 0,245602472 | 6,653122 | 6,3462917 | 6,62797 | 6,3438826 | -1,2272752 |
| 16,9039231 | 0,337191474 | 16,73467 | 17,006204 | 16,55131 | 17,136621 | 1,3457622 |
| 3,77257535 | 0,30856575 | 3,997677 | 3,538649 | 4,009554 | 3,7197634 | -1,2963087 |

| | | | | | | |
|---|---|---|---|---|---|---|
| 8,28244255 | 0,240711725 | 8,46264 | 8,1669476 | 8,386356 | 8,2055208 | -1,1795721 |
| 3,4198454 | 0,363667393 | 3,182468 | 3,695652 | 3,405609 | 3,2991904 | 1,15139502 |
| 11,8133569 | 0,234636822 | 11,63967 | 11,912956 | 11,68858 | 11,916336 | 1,18963552 |
| 0,4137262 | 0,226611516 | 0,205144 | 0,4923094 | 0,47816 | 0,4580616 | 1,09697753 |
| 6,73324256 | 0,42173912 | 7,02538 | 6,6774822 | 6,959619 | 6,4160124 | -1,3620241 |
| 16,7325618 | 0,371336151 | 16,94442 | 16,462053 | 16,97063 | 16,73425 | -1,2828699 |
| 15,3090739 | 0,266295438 | 15,46497 | 15,172516 | 15,4921 | 15,230613 | -1,2116493 |
| 10,9338253 | 0,299028499 | 11,09589 | 10,711632 | 11,15226 | 10,931646 | -1,2332222 |
| 5,35042757 | 0,373090299 | 5,628477 | 5,2366673 | 5,470866 | 5,1783344 | -1,2676628 |
| 17,0612618 | 0,280669366 | 17,30987 | 16,89486 | 17,15431 | 16,993799 | -1,2207429 |
| 15,8961884 | 0,421380789 | 15,92701 | 15,56265 | 16,11391 | 16,139603 | -1,1245382 |
| 16,6593715 | 0,45927852 | 16,73584 | 16,302706 | 16,88064 | 16,889606 | -1,1583629 |
| 6,46109146 | 0,276758149 | 6,649308 | 6,3193015 | 6,618906 | 6,3765662 | -1,2194017 |
| 4,06083042 | 0,210713128 | 4,247842 | 3,9817953 | 4,146889 | 3,9450778 | -1,1760331 |
| 8,20561304 | 0,289976342 | 8,398191 | 8,0770127 | 8,395748 | 8,0821609 | -1,246068 |
| 14,2885898 | 0,345032801 | 14,56928 | 14,07549 | 14,44814 | 14,209997 | -1,2887415 |
| 12,5638762 | 0,255859034 | 12,76879 | 12,431331 | 12,75462 | 12,434307 | -1,2560435 |
| 9,48334263 | 0,297768802 | 9,756834 | 9,3913932 | 9,438243 | 9,3882739 | -1,1548497 |
| 21,3900124 | 0,501672849 | 21,07176 | 21,632703 | 20,86731 | 21,682922 | 1,61136043 |
| 5,29881929 | 0,348470753 | 5,561476 | 5,2521739 | 5,402308 | 5,0694424 | -1,2492687 |
| 14,0885363 | 0,313678569 | 14,37556 | 13,865457 | 14,20439 | 14,042752 | -1,2621373 |
| 9,93738328 | 0,247320874 | 9,731909 | 10,077574 | 9,802487 | 10,024746 | 1,2175337 |
| 1,98598761 | 0,247371361 | 1,751614 | 2,0940344 | 1,936101 | 2,0854813 | 1,18583284 |
| 17,0488913 | 0,266496899 | 16,86329 | 17,150057 | 16,8708 | 17,192104 | 1,23458988 |
| 7,01486565 | 0,379015993 | 7,379722 | 6,9308784 | 7,041133 | 6,7897625 | -1,2746549 |
| 5,23551503 | 0,236045923 | 5,032221 | 5,4053414 | 5,184076 | 5,2353867 | 1,15846584 |
| 11,7586509 | 0,276407936 | 12,03906 | 11,621752 | 11,78298 | 11,669977 | -1,2017693 |
| 12,1872663 | 0,285085175 | 11,97378 | 12,2609 | 12,06304 | 12,354903 | 1,22220775 |
| 17,7965237 | 0,396072662 | 17,65041 | 17,949493 | 17,36061 | 17,998706 | 1,38375676 |
| 11,9186328 | 0,232977497 | 12,14023 | 11,728305 | 11,98998 | 11,91586 | -1,1834694 |
| 13,8751483 | 0,231443916 | 14,08542 | 13,729184 | 13,92664 | 13,84068 | -1,1656209 |
| 4,27707811 | 0,461949886 | 4,610414 | 4,1934464 | 4,461295 | 3,9835703 | -1,3635299 |
| 8,27208481 | 0,22348103 | 8,427211 | 8,1479367 | 8,459193 | 8,1769402 | -1,2148377 |
| 12,1871311 | 0,400565847 | 11,88595 | 12,2931 | 12,04663 | 12,400272 | 1,30169815 |
| 5,24121699 | 0,269627419 | 5,535315 | 5,1225062 | 5,335737 | 5,0769748 | -1,2620643 |
| 5,25615597 | 0,330830143 | 4,963262 | 5,4590564 | 5,302725 | 5,2336938 | 1,15940244 |
| 15,0638263 | 0,300115874 | 14,74159 | 15,235997 | 14,90727 | 15,22506 | 1,32509972 |
| 12,5409846 | 0,36085203 | 12,16584 | 12,81409 | 12,37136 | 12,633903 | 1,37115723 |
| 12,2150596 | 0,338191181 | 11,85643 | 12,461071 | 12,12794 | 12,276955 | 1,29848508 |
| 10,8284394 | 0,265449514 | 10,57014 | 10,903708 | 10,7874 | 10,982294 | 1,20099424 |
| 3,84950184 | 0,233694907 | 4,015048 | 3,7439814 | 3,943724 | 3,7788785 | -1,1630848 |
| 2,69330464 | 0,268786219 | 2,869794 | 2,585589 | 2,809041 | 2,6031058 | -1,1851503 |
| 8,18672955 | 0,334363465 | 8,458982 | 8,0197676 | 8,28009 | 8,0996533 | -1,2395576 |
| 5,72767017 | 0,235243909 | 5,512658 | 5,8163033 | 5,785493 | 5,769377 | 1,10478422 |
| 11,511698 | 0,294803597 | 11,34581 | 11,650308 | 11,26249 | 11,635896 | 1,26483854 |
| 8,50440176 | 0,237700006 | 8,708929 | 8,3228758 | 8,615918 | 8,4814745 | -1,1976852 |
| 13,0012783 | 0,271051781 | 13,23817 | 12,832733 | 13,08284 | 12,953234 | -1,2037396 |
| 11,4601451 | 0,419016448 | 11,42987 | 11,827891 | 11,08637 | 11,26885 | 1,2228529 |
| 3,02699508 | 0,381223093 | 2,783876 | 3,1685481 | 2,863212 | 3,1620523 | 1,26729822 |
| 15,2984913 | 0,280065013 | 15,52024 | 15,057517 | 15,48512 | 15,287201 | -1,2572905 |
| 12,7124175 | 0,200404318 | 12,84697 | 12,527599 | 12,90097 | 12,706698 | -1,1948425 |
| 6,49074042 | 0,226071415 | 6,700206 | 6,4051438 | 6,563004 | 6,3720524 | -1,1834567 |
| 15,3232081 | 0,200259177 | 15,53199 | 15,177438 | 15,31556 | 15,325251 | -1,1269584 |

| | | | | | | |
|---|---|---|---|---|---|---|
| 8,46935113 | 0,427280847 | 8,26423 | 8,6531336 | 8,147443 | 8,6163098 | 1,34619287 |
| 17,9093698 | 0,44208451 | 17,61239 | 18,072762 | 17,63437 | 18,130731 | 1,39316525 |
| 16,6257949 | 0,267372792 | 16,84866 | 16,46935 | 16,71749 | 16,569077 | -1,2006876 |
| 11,6070469 | 0,265100706 | 11,42948 | 11,698925 | 11,45234 | 11,740543 | 1,2132022 |
| 10,7656632 | 0,283888374 | 10,56495 | 10,913047 | 10,63222 | 10,839478 | 1,21224387 |
| 10,3718982 | 0,226525441 | 10,50948 | 10,217318 | 10,51185 | 10,356482 | -1,1677755 |
| 14,0699063 | 0,432436194 | 14,35796 | 13,827735 | 14,40604 | 13,913993 | -1,4251679 |
| 7,21908316 | 0,423471954 | 7,635439 | 7,010314 | 7,380094 | 7,0190974 | -1,4074276 |
| 11,1787733 | 0,259931201 | 11,4061 | 11,078971 | 11,28242 | 11,043123 | -1,2169 |
| 7,4514183 | 0,429824874 | 7,253784 | 7,736963 | 7,105416 | 7,484355 | 1,34822277 |
| 4,26807024 | 0,311580848 | 3,990896 | 4,3269299 | 4,438966 | 4,3304997 | 1,08206267 |
| 13,4944294 | 0,353827682 | 13,24871 | 13,685157 | 13,27371 | 13,606852 | 1,30567541 |
| 17,7513495 | 0,393540731 | 17,52935 | 17,851559 | 17,48874 | 17,977358 | 1,32446475 |
| 16,0660995 | 0,326406738 | 15,87451 | 16,164654 | 15,82828 | 16,253373 | 1,28130648 |
| 13,3637539 | 0,342175965 | 13,08538 | 13,488499 | 13,24574 | 13,521485 | 1,26525892 |
| 5,9698127 | 0,316820638 | 6,242393 | 5,8469262 | 6,061353 | 5,8306595 | -1,242357 |
| 4,87958632 | 0,33561112 | 5,142114 | 4,8605704 | 5,009072 | 4,6015651 | -1,2697334 |
| 11,5932329 | 0,350789472 | 11,83416 | 11,335167 | 11,72909 | 11,616608 | -1,2360522 |
| 4,58153984 | 0,345634248 | 4,809506 | 4,4426932 | 4,799948 | 4,4233399 | -1,2938858 |
| 6,23527796 | 0,196143335 | 6,40524 | 6,1216926 | 6,366407 | 6,1484349 | -1,1898337 |
| 4,73068771 | 0,228655407 | 4,93313 | 4,5996394 | 4,810677 | 4,6678758 | -1,179476 |
| 5,92905591 | 0,309536672 | 6,18373 | 5,7764551 | 6,011107 | 5,8464731 | -1,2192167 |
| 8,94933854 | 0,202297639 | 9,119267 | 8,8544455 | 9,037691 | 8,8657599 | -1,1634233 |
| 1,43976386 | 0,27190563 | 1,22804 | 1,571697 | 1,361901 | 1,508127 | 1,18504474 |
| 7,62239023 | 0,293429514 | 7,435686 | 7,7988737 | 7,416832 | 7,6926432 | 1,24789726 |
| 5,61611298 | 0,379946805 | 5,254793 | 5,8413752 | 5,597853 | 5,6638764 | 1,25379616 |
| 8,18523361 | 0,264565328 | 8,464761 | 8,0066008 | 8,219073 | 8,1416912 | -1,2039463 |
| 6,3188472 | 0,193946882 | 6,514153 | 6,1857479 | 6,347859 | 6,2951492 | -1,1412046 |
| 7,41732511 | 0,270893189 | 7,225068 | 7,5676367 | 7,266826 | 7,4906695 | 1,21689643 |
| 3,73084097 | 0,251877244 | 3,580689 | 3,938656 | 3,672354 | 3,6441844 | 1,12108738 |
| 3,13056011 | 0,173654795 | 2,960903 | 3,2711741 | 3,080444 | 3,1361013 | 1,13521386 |
| 14,7201788 | 0,32549991 | 14,54399 | 14,86922 | 14,44816 | 14,854302 | 1,2884948 |
| 14,9426805 | 0,429210465 | 14,59037 | 15,179248 | 14,77568 | 15,058466 | 1,35269093 |
| 15,4711321 | 0,321435026 | 15,2415 | 15,599872 | 15,22724 | 15,658172 | 1,31462855 |
| 9,23912871 | 0,213352042 | 9,420334 | 9,137024 | 9,310638 | 9,1647242 | -1,1603918 |
| 13,1785833 | 0,257117174 | 12,95258 | 13,226107 | 13,10327 | 13,358846 | 1,20126292 |
| 11,2557808 | 0,281836947 | 11,51965 | 11,064193 | 11,30618 | 11,231158 | -1,2018351 |
| 10,4414755 | 0,269009065 | 10,72683 | 10,252954 | 10,55972 | 10,354207 | -1,2654865 |
| 8,65006968 | 0,319736726 | 8,898998 | 8,5350152 | 8,802567 | 8,4850474 | -1,2664158 |
| 10,9370363 | 0,306502898 | 10,70894 | 11,117538 | 10,74738 | 11,02811 | 1,2698547 |
| 1,65851886 | 0,322853446 | 1,892301 | 1,5733956 | 1,948339 | 1,3880597 | -1,3562211 |
| 8,27072767 | 0,276012824 | 8,47666 | 8,1439747 | 8,368748 | 8,188977 | -1,1943518 |
| 17,0458001 | 0,342261752 | 16,8202 | 17,163925 | 16,86291 | 17,205544 | 1,26854912 |
| 14,6071149 | 0,373135597 | 14,36534 | 14,785925 | 14,25629 | 14,808543 | 1,40096046 |
| 8,83334276 | 0,439338745 | 9,080686 | 8,6330079 | 9,101927 | 8,7023523 | -1,3412948 |
| 13,4652986 | 0,253104519 | 13,3074 | 13,578827 | 13,29434 | 13,565849 | 1,20703387 |
| 14,2200011 | 0,380188715 | 13,97076 | 14,382053 | 13,87843 | 14,442339 | 1,40211307 |
| 9,13444219 | 0,265476721 | 9,295163 | 8,9741742 | 9,280802 | 9,102335 | -1,1889829 |
| 20,5506441 | 0,323705137 | 20,82831 | 20,314384 | 20,64946 | 20,538851 | -1,2416576 |
| 7,55901125 | 0,303818504 | 7,752798 | 7,4170665 | 7,72318 | 7,4661248 | -1,2280705 |
| 12,9249037 | 0,261536677 | 13,12366 | 12,699159 | 13,08087 | 12,933268 | -1,2193025 |
| 4,39604391 | 0,232611931 | 4,626961 | 4,3304832 | 4,481843 | 4,2269583 | -1,2105653 |
| 5,14199087 | 0,328456833 | 5,541663 | 4,9861833 | 5,245147 | 4,9273449 | -1,3534495 |

| | | | | | | |
|---|---|---|---|---|---|---|
| 7,50276554 | 0,27729421 | 7,791361 | 7,3000971 | 7,609613 | 7,4365527 | -1,2588987 |
| 11,0287767 | 0,227014155 | 11,2267 | 10,870916 | 11,11248 | 10,999755 | -1,1762975 |
| 16,5761355 | 0,322019309 | 16,39902 | 16,692613 | 16,32243 | 16,739133 | 1,27911613 |
| 8,56151829 | 0,449003009 | 8,077839 | 8,788755 | 8,53115 | 8,7181826 | 1,36506935 |
| 9,14899831 | 0,274502478 | 8,919384 | 9,2670991 | 8,983886 | 9,3015173 | 1,25934497 |
| 13,4456941 | 0,331876889 | 13,24036 | 13,591076 | 13,25934 | 13,557578 | 1,25220872 |
| 16,5577379 | 0,340098394 | 16,34182 | 16,668631 | 16,31151 | 16,755932 | 1,30641786 |
| 8,49512414 | 0,287538723 | 8,279975 | 8,6753965 | 8,290185 | 8,584637 | 1,27009493 |
| 10,2039627 | 0,380224855 | 9,950467 | 10,416892 | 9,998349 | 10,287287 | 1,29925239 |
| 15,2072292 | 0,299653168 | 14,99219 | 15,310975 | 14,96605 | 15,410229 | 1,30268214 |
| 11,6956135 | 0,218956364 | 11,95218 | 11,601151 | 11,5949 | 11,651322 | -1,1074945 |
| 13,9799193 | 0,29329249 | 14,18277 | 13,780457 | 14,12194 | 13,961639 | -1,2152954 |
| 5,54442882 | 0,458853314 | 5,198359 | 5,6261614 | 5,403905 | 5,8248234 | 1,34197746 |
| 10,274393 | 0,275173617 | 10,0419 | 10,392921 | 10,16494 | 10,39545 | 1,22328956 |
| 13,5840331 | 0,242107921 | 13,81934 | 13,447566 | 13,68182 | 13,489112 | -1,2160821 |
| 6,51894933 | 0,331561896 | 6,684 | 6,4022235 | 6,750998 | 6,3794979 | -1,2540878 |
| 8,79424558 | 0,225570405 | 8,614491 | 8,9132801 | 8,667195 | 8,8804256 | 1,19417117 |
| 14,1766913 | 0,290413359 | 14,42827 | 14,033191 | 14,35311 | 14,029199 | -1,2829769 |
| 8,019632 | 0,215371575 | 7,83426 | 8,1535925 | 7,950444 | 8,057958 | 1,15943583 |
| 3,81404397 | 0,230137012 | 3,988422 | 3,6783831 | 3,946377 | 3,7492159 | -1,1921782 |
| 3,21432395 | 0,349111248 | 2,855254 | 3,3351791 | 3,248346 | 3,3540939 | 1,22504648 |
| 8,36720224 | 0,245859133 | 8,216763 | 8,4906092 | 8,152986 | 8,4755172 | 1,22959976 |
| 2,48549482 | 0,284513056 | 2,664652 | 2,3452754 | 2,715159 | 2,3636758 | -1,2617535 |
| 2,51790452 | 0,231188158 | 2,711596 | 2,4437623 | 2,622077 | 2,3797334 | -1,1934094 |
| 2,24876682 | 0,202140341 | 2,063653 | 2,3307792 | 2,3391 | 2,2534989 | 1,06493303 |
| 11,1970538 | 0,292332927 | 11,03885 | 11,321923 | 10,96826 | 11,318963 | 1,24564251 |
| 6,81466261 | 0,213827382 | 6,620226 | 6,9423271 | 6,782353 | 6,8461222 | 1,14308701 |
| 9,06594463 | 0,218315862 | 8,9185 | 9,2077034 | 8,931357 | 9,1019142 | 1,17273748 |
| 1,8733996 | 0,226181306 | 2,048907 | 1,7780265 | 1,949714 | 1,7928775 | -1,1597859 |
| 1,14767206 | 0,243949682 | 1,334445 | 1,053857 | 1,248425 | 1,0410408 | -1,1842605 |
| 0,86125307 | 0,394458467 | 0,59738 | 1,1577083 | 0,791119 | 0,7718793 | 1,20626281 |
| 6,40861554 | 0,73764738 | 7,07617 | 6,4456278 | 6,262892 | 5,8842133 | -1,4187404 |
| 14,8680471 | 0,224304116 | 15,10447 | 14,782367 | 14,84871 | 14,781502 | -1,1444528 |
| 12,5438312 | 0,405635962 | 12,24922 | 12,711296 | 12,38598 | 12,687999 | 1,3031871 |
| 13,1096168 | 0,277061211 | 12,89843 | 13,220284 | 12,90964 | 13,276307 | 1,26949784 |
| 12,1780471 | 0,308459135 | 12,33913 | 11,975468 | 12,46598 | 12,111461 | -1,2826173 |
| 7,11233515 | 0,244583637 | 7,292859 | 6,9535018 | 7,303007 | 7,0350863 | -1,234254 |
| 8,19641183 | 0,248879587 | 7,950972 | 8,3082153 | 8,068148 | 8,3478298 | 1,24700092 |
| 2,74818646 | 0,392113412 | 2,510403 | 2,9917952 | 2,548008 | 2,7780791 | 1,27963438 |
| 10,6546592 | 0,25402098 | 10,95394 | 10,517006 | 10,65794 | 10,563482 | -1,2022177 |
| 13,3061557 | 0,285272381 | 13,11905 | 13,446664 | 13,13709 | 13,398029 | 1,22627158 |
| 15,7491199 | 0,311987015 | 15,46851 | 15,906204 | 15,64413 | 15,862134 | 1,25514246 |
| 10,39577 | 0,355328261 | 10,43155 | 10,096972 | 10,67349 | 10,557279 | -1,1690987 |
| 2,46452992 | 0,253481204 | 2,607016 | 2,3366729 | 2,620232 | 2,403424 | -1,1839232 |
| 13,3510739 | 0,357271834 | 13,17397 | 13,470388 | 13,01599 | 13,559486 | 1,3378881 |
| 12,3009339 | 0,344536284 | 12,09652 | 12,433963 | 12,04136 | 12,470802 | 1,30445499 |
| 18,9163216 | 0,287638965 | 19,0311 | 18,75535 | 19,20056 | 18,841377 | -1,2461442 |
| 11,068659 | 0,41590054 | 11,40973 | 10,853444 | 11,28819 | 10,905288 | -1,3847202 |
| 13,5641672 | 0,275132055 | 13,80521 | 13,403465 | 13,71256 | 13,463087 | -1,2531943 |
| 5,05568175 | 0,236352042 | 5,286657 | 4,9080346 | 5,150413 | 4,979691 | -1,209719 |
| 13,8205588 | 0,236849999 | 14,00743 | 13,696334 | 13,92075 | 13,750669 | -1,1814774 |
| 9,57763376 | 0,268129773 | 9,788336 | 9,4696585 | 9,706565 | 9,4507486 | -1,2203092 |
| -0,46228681 | 0,302794186 | -0,110427 | -0,5309617 | -0,53423 | -0,635789 | -1,1983464 |

| | | | | | | |
|---|---|---|---|---|---|---|
| 4,62724162 | 0,321553764 | 4,85274 | 4,4690957 | 4,830404 | 4,5034458 | -1,2792526 |
| 4,70999634 | 0,262476362 | 4,856299 | 4,5263112 | 4,954354 | 4,6594469 | -1,2418125 |
| 13,8597628 | 0,240128593 | 14,08006 | 13,759431 | 13,95177 | 13,737701 | -1,2035963 |
| 15,4917333 | 0,36824638 | 15,34052 | 15,646495 | 15,11201 | 15,662388 | 1,34553479 |
| 23,5980448 | 0,462312592 | 23,43765 | 23,762139 | 23,10391 | 23,833947 | 1,44119056 |
| 9,72031809 | 0,261474697 | 9,905948 | 9,5879052 | 9,827059 | 9,6573835 | -1,184156 |
| 6,67948544 | 0,32831211 | 6,938058 | 6,5480517 | 6,757397 | 6,5706728 | -1,2212555 |
| 8,11921826 | 0,265339809 | 8,326805 | 7,983678 | 8,218665 | 8,04575 | -1,1958371 |
| 9,00224909 | 0,235462537 | 9,147837 | 8,8274859 | 9,269663 | 8,9277669 | -1,2579927 |
| 18,9377089 | 0,31179574 | 19,25231 | 18,734805 | 18,90094 | 18,935843 | -1,1820581 |
| 10,2650483 | 0,239239759 | 10,44787 | 10,143826 | 10,38416 | 10,183648 | -1,1910864 |
| 6,37096946 | 0,236474539 | 6,556576 | 6,2371106 | 6,434946 | 6,335449 | -1,1562721 |
| 5,38048577 | 0,510200783 | 5,78607 | 5,1705911 | 5,544161 | 5,1894083 | -1,3996977 |
| 6,13713293 | 0,473867411 | 6,487474 | 5,9374855 | 6,285394 | 5,9899636 | -1,3404425 |
| 7,96292177 | 0,266043877 | 8,174618 | 7,8733696 | 8,078539 | 7,8210727 | -1,213654 |
| 8,0673864 | 0,393658882 | 8,39041 | 7,8990789 | 8,273044 | 7,8713907 | -1,3627229 |
| 9,67087414 | 0,26260715 | 9,836534 | 9,5291813 | 9,817118 | 9,612348 | -1,1942142 |
| 8,46272236 | 0,531015677 | 8,521843 | 8,0669786 | 8,765002 | 8,7059944 | -1,1949384 |
| 7,03785602 | 0,287163489 | 7,198786 | 6,8597342 | 7,334863 | 6,9366076 | -1,2911473 |
| 16,6491165 | 0,552085729 | 17,16601 | 16,32092 | 16,86385 | 16,474752 | -1,533783 |
| 13,7097083 | 0,225269615 | 13,88418 | 13,58922 | 13,85214 | 13,620532 | -1,2002082 |
| 14,9768864 | 0,30084218 | 14,78314 | 15,083802 | 14,78222 | 15,13007 | 1,25201765 |
| 10,476984 | 0,20068731 | 10,68549 | 10,393497 | 10,53209 | 10,366876 | -1,1716991 |
| 7,77238346 | 0,382556666 | 7,421839 | 7,7636055 | 7,952319 | 7,9729177 | 1,13381255 |
| 6,47637137 | 0,242731756 | 6,668606 | 6,355004 | 6,560058 | 6,4084011 | -1,1749744 |
| 7,31859808 | 0,464617563 | 7,710733 | 7,1797681 | 7,387709 | 7,110413 | -1,323291 |
| 6,8642049 | 0,198514051 | 7,02486 | 6,7565835 | 6,917175 | 6,8250116 | -1,1330566 |
| 9,45874309 | 0,302099695 | 9,708263 | 9,3458643 | 9,564816 | 9,3184622 | -1,2348845 |
| 8,97566912 | 0,264417174 | 8,735851 | 9,1167587 | 8,902979 | 9,0538209 | 1,20236474 |
| 2,99656888 | 0,512021711 | 3,320383 | 2,7948932 | 3,221179 | 2,8285717 | -1,3746349 |
| 11,4130524 | 0,299397005 | 11,18305 | 11,498219 | 11,31229 | 11,566811 | 1,21827898 |
| 14,9797439 | 0,305040757 | 14,78892 | 15,110838 | 14,77094 | 15,109915 | 1,25740323 |
| 15,5089474 | 0,248969515 | 15,76635 | 15,356044 | 15,48976 | 15,485149 | -1,154653 |
| 9,37649457 | 0,432012942 | 8,980177 | 9,5204409 | 9,303634 | 9,5843448 | 1,32913448 |
| 5,79807864 | 0,270607675 | 6,020209 | 5,679054 | 5,896053 | 5,6933124 | -1,2074371 |
| 1,90880589 | 0,288171234 | 1,644883 | 2,1096715 | 1,888872 | 1,9040623 | 1,18098424 |
| 7,54525798 | 0,225101617 | 7,757571 | 7,4915364 | 7,549956 | 7,4264386 | -1,1445467 |
| 11,0121619 | 0,361135474 | 11,35808 | 10,919381 | 10,81298 | 10,948977 | -1,1106099 |
| 3,48094076 | 0,310589666 | 3,16991 | 3,6040335 | 3,494272 | 3,5896067 | 1,20141039 |
| 12,354167 | 0,341338271 | 12,0996 | 12,55963 | 12,10977 | 12,470629 | 1,32909354 |
| 8,07865131 | 0,427471523 | 7,862716 | 8,2525127 | 7,743589 | 8,2545998 | 1,36642245 |
| 13,2535535 | 0,319757244 | 13,03258 | 13,376291 | 12,98654 | 13,454304 | 1,32476598 |
| 6,38728897 | 0,246597263 | 6,544928 | 6,2593691 | 6,513639 | 6,3309893 | -1,1761764 |
| 8,04013299 | 0,268565817 | 8,259502 | 7,8756899 | 8,106231 | 8,0113429 | -1,1804604 |
| 9,8940081 | 0,258710788 | 10,16478 | 9,7999993 | 9,828574 | 9,8159198 | -1,1397516 |
| 13,6354222 | 0,285955787 | 13,89949 | 13,46007 | 13,69437 | 13,586018 | -1,2090603 |
| 12,8007442 | 0,205807802 | 12,6563 | 12,925102 | 12,69887 | 12,835418 | 1,15083072 |
| 7,25784915 | 0,351143469 | 6,879206 | 7,4824541 | 7,128416 | 7,3878298 | 1,34847699 |
| 13,1665318 | 0,326961112 | 13,45095 | 13,051757 | 13,24633 | 13,014622 | -1,2444031 |
| 13,8992181 | 0,340250648 | 13,75615 | 14,020355 | 13,58698 | 14,062808 | 1,29236974 |
| 12,781182 | 0,282517496 | 12,99115 | 12,623576 | 12,92215 | 12,707257 | -1,2236854 |
| 14,0934698 | 0,201785512 | 14,27906 | 13,985646 | 14,13238 | 14,041764 | -1,1423557 |
| 14,1824236 | 0,294566079 | 14,48227 | 13,975989 | 14,32193 | 14,09157 | -1,2908498 |

| | | | | | | |
|---|---|---|---|---|---|---|
| 5,84599097 | 0,216895101 | 6,041826 | 5,7675546 | 5,95808 | 5,7064014 | -1,1999505 |
| 22,3449927 | 0,494344068 | 22,89139 | 22,128631 | 22,26183 | 22,190086 | -1,335381 |
| 8,26573263 | 0,511423012 | 8,556123 | 8,1762325 | 8,564166 | 7,9472407 | -1,4126539 |
| 13,0421177 | 0,336434225 | 13,29519 | 12,796009 | 13,24854 | 12,99848 | -1,2965007 |
| 8,09876473 | 0,364766709 | 8,302387 | 7,8634352 | 8,366292 | 8,0475652 | -1,3002951 |
| 2,4479003 | 0,506227857 | 1,966171 | 2,6739788 | 2,497605 | 2,5562525 | 1,30425616 |
| 10,7338157 | 0,206654541 | 10,89703 | 10,625822 | 10,81428 | 10,676395 | -1,1523253 |
| 10,4458682 | 0,224779744 | 10,56991 | 10,280926 | 10,58893 | 10,45252 | -1,1588559 |
| 10,9520628 | 0,274516842 | 11,24759 | 10,827163 | 11,02705 | 10,805752 | -1,2490772 |
| 16,7005554 | 0,3217045 | 16,40017 | 16,839519 | 16,56096 | 16,872884 | 1,29741195 |
| 8,52197683 | 0,542247592 | 9,066717 | 8,37483 | 8,611435 | 8,1818495 | -1,4750215 |
| 5,37141932 | 0,411823587 | 5,107696 | 5,6252723 | 5,193945 | 5,3974457 | 1,28390531 |
| 9,47838934 | 0,328227579 | 9,264587 | 9,6304789 | 9,23901 | 9,6212414 | 1,29599626 |
| 10,4816269 | 0,280620826 | 10,28128 | 10,608784 | 10,29674 | 10,610267 | 1,24877768 |
| 18,4367842 | 0,286864773 | 18,57186 | 18,256453 | 18,71107 | 18,373798 | -1,2538273 |
| 8,00434616 | 0,33818454 | 7,756775 | 8,1187432 | 7,891463 | 8,1452357 | 1,23787947 |
| 3,9503262 | 0,282279794 | 4,204376 | 3,8083406 | 4,093582 | 3,8188128 | -1,2617294 |
| 15,4556439 | 0,447936556 | 15,20582 | 15,606093 | 15,14165 | 15,675848 | 1,38245549 |
| 14,172302 | 0,320091367 | 14,36702 | 13,985704 | 14,38618 | 14,102388 | -1,2592372 |
| 15,0787026 | 0,208065042 | 15,27113 | 14,941343 | 15,13089 | 15,048658 | -1,1534928 |
| 4,60593283 | 0,365703443 | 4,89554 | 4,6071017 | 4,711602 | 4,2949628 | -1,2768054 |
| 8,69788671 | 0,442371745 | 8,464441 | 8,9545188 | 8,430475 | 8,7488041 | 1,32335807 |
| 10,9995161 | 0,227794911 | 11,18581 | 10,880386 | 11,09412 | 10,927357 | -1,1778003 |
| 11,6857341 | 0,232202597 | 11,87465 | 11,500496 | 11,82899 | 11,661486 | -1,2065019 |
| 14,1274119 | 0,344397028 | 13,8271 | 14,276465 | 13,93097 | 14,321681 | 1,3379644 |
| 7,37462155 | 0,21021569 | 7,572966 | 7,2781286 | 7,382704 | 7,3169709 | -1,1331079 |
| 15,8377291 | 0,286722339 | 16,12829 | 15,686912 | 15,9004 | 15,734128 | -1,2344133 |
| 10,5112565 | 0,282906141 | 10,71937 | 10,303166 | 10,70109 | 10,470172 | -1,2514139 |
| 16,6655324 | 0,17682949 | 16,86218 | 16,568974 | 16,67691 | 16,607422 | -1,1339445 |
| 19,4352499 | 0,457936383 | 19,70652 | 19,233047 | 19,79016 | 19,234366 | -1,4286325 |
| 14,1179917 | 0,200003836 | 14,30332 | 14,015383 | 14,17578 | 14,048922 | -1,1546024 |
| 19,3223162 | 0,426314905 | 19,06267 | 19,444814 | 19,03519 | 19,568296 | 1,37328004 |
| 14,4223506 | 0,263436776 | 14,65425 | 14,22505 | 14,55749 | 14,380914 | -1,2336103 |
| 13,3404413 | 0,246688031 | 13,18171 | 13,450238 | 13,17883 | 13,440575 | 1,20175014 |
| 9,35771313 | 0,306486483 | 9,564451 | 9,1715664 | 9,538254 | 9,2970374 | -1,2457812 |
| 18,6482159 | 0,460069975 | 18,98893 | 18,35687 | 18,85466 | 18,584356 | -1,3671607 |
| 10,9664823 | 0,262253081 | 11,15579 | 10,818042 | 11,08336 | 10,913573 | -1,1923159 |
| 14,5981538 | 0,365627636 | 14,35193 | 14,642918 | 14,4007 | 14,8722 | 1,30246509 |
| 0,87756525 | 0,245004489 | 0,698615 | 1,0039846 | 0,76619 | 0,9447946 | 1,18262025 |
| 12,2422425 | 0,389340472 | 12,02954 | 12,358952 | 11,95066 | 12,45794 | 1,33639626 |
| 18,0026259 | 0,456880818 | 17,918 | 18,418044 | 17,50086 | 17,877119 | 1,35486936 |
| 8,43965022 | 0,401653742 | 8,044928 | 8,4830476 | 8,488469 | 8,6937956 | 1,24982224 |
| 5,57541746 | 0,40326357 | 5,870608 | 5,4136032 | 5,744127 | 5,4174573 | -1,312063 |
| 8,42557856 | 0,296962135 | 8,271193 | 8,6144468 | 8,191106 | 8,4708478 | 1,24099536 |
| 6,9235844 | 0,222048666 | 6,774243 | 7,0469193 | 6,781771 | 6,9876112 | 1,18038586 |
| 14,9291909 | 0,277110134 | 15,21178 | 14,753962 | 14,86984 | 14,934872 | -1,1458315 |
| 20,0405575 | 0,371108911 | 19,83916 | 20,11041 | 19,72276 | 20,318597 | 1,35054381 |
| 19,8939322 | 0,466131499 | 20,37367 | 19,517817 | 19,93753 | 19,911332 | -1,3575715 |
| 13,0185088 | 0,220007088 | 13,29306 | 12,947517 | 12,87157 | 12,958492 | -1,0937715 |
| 6,44394458 | 0,300695921 | 6,58969 | 6,2923394 | 6,665001 | 6,3693539 | -1,22816 |
| 15,0122203 | 0,260019886 | 15,28888 | 14,897663 | 15,14864 | 14,832679 | -1,277734 |
| 8,88533752 | 0,230580547 | 9,103917 | 8,7448596 | 8,939434 | 8,8356606 | -1,1739861 |
| 8,16433259 | 0,352768334 | 8,484633 | 8,0249649 | 8,078457 | 8,1108442 | -1,1596104 |

| | | | | | | |
|---|---|---|---|---|---|---|
| 14,8110834 | 0,492907482 | 14,44379 | 15,173561 | 14,51239 | 14,867529 | 1,45645189 |
| 12,1758684 | 0,316265425 | 11,8685 | 12,307686 | 12,1435 | 12,298375 | 1,22861465 |
| 14,3195435 | 0,46062317 | 14,74741 | 14,367748 | 14,10971 | 14,023918 | -1,17505 |
| 1,49123063 | 0,490782035 | 1,058917 | 1,7293022 | 1,546259 | 1,5399941 | 1,25880957 |
| 7,21209096 | 0,317448093 | 7,425518 | 7,0340373 | 7,398442 | 7,1325318 | -1,2558772 |
| 16,1603879 | 0,361514086 | 15,88441 | 16,209838 | 15,98956 | 16,438121 | 1,30766386 |
| 12,150469 | 0,238367629 | 12,32659 | 11,956567 | 12,17712 | 12,217455 | -1,1210466 |
| 12,2560378 | 0,230592219 | 12,43414 | 12,074662 | 12,28248 | 12,306438 | -1,1233127 |
| 1,79689603 | 0,201713228 | 1,989596 | 1,7258302 | 1,816055 | 1,7068847 | -1,1379743 |
| 18,8573091 | 0,303548927 | 18,65624 | 19,0076 | 18,67789 | 18,955518 | 1,24357368 |
| 6,04776439 | 0,399059463 | 6,360775 | 5,8456623 | 6,201385 | 5,9320554 | -1,3124125 |
| 8,54143949 | 0,36295748 | 8,838133 | 8,3646567 | 8,691238 | 8,4115104 | -1,2982803 |
| 7,07067853 | 0,367758422 | 6,90306 | 7,2390306 | 6,786621 | 7,1815658 | 1,28829 |
| 10,3187057 | 0,227390881 | 10,44959 | 10,153166 | 10,47971 | 10,309495 | -1,1755387 |
| 19,4751916 | 0,556962326 | 19,14076 | 19,720817 | 19,04978 | 19,719957 | 1,54233827 |
| 0,89085803 | 0,193010002 | 0,704839 | 0,986834 | 0,949049 | 0,8988884 | 1,08366384 |
| 9,57887897 | 0,425395191 | 9,297324 | 9,7373102 | 9,332533 | 9,7758905 | 1,3581772 |
| 3,7437648 | 0,247733912 | 3,504763 | 3,8462287 | 3,78435 | 3,7996084 | 1,13159832 |
| 5,76018314 | 0,918938924 | 4,900545 | 5,976001 | 5,917267 | 6,1376442 | 1,5669039 |
| 11,8605733 | 0,31391517 | 11,64506 | 12,033232 | 11,70228 | 11,931547 | 1,23860801 |
| 7,0996037 | 0,240947411 | 7,357236 | 6,9319666 | 7,175067 | 7,036503 | -1,2158089 |
| 2,54687833 | 0,242383596 | 2,40614 | 2,6846417 | 2,408878 | 2,5839904 | 1,17024233 |
| 7,22428748 | 0,401650049 | 6,946937 | 7,3748889 | 7,024394 | 7,3992499 | 1,32079271 |
| 8,70083857 | 0,456946727 | 8,295232 | 8,6799124 | 8,696635 | 9,0732378 | 1,30192076 |
| 6,17137535 | 0,267225028 | 6,368452 | 6,0521753 | 6,283864 | 6,0794067 | -1,1977836 |
| 9,04093684 | 0,313150005 | 9,329429 | 8,9748709 | 8,98415 | 8,9090695 | -1,1605587 |
| 17,7594761 | 0,447666156 | 17,49737 | 17,94918 | 17,45365 | 17,938117 | 1,38332304 |
| 20,1888103 | 0,365815328 | 20,53192 | 19,882343 | 20,24058 | 20,233869 | -1,2553962 |
| 14,1133078 | 0,490197587 | 13,7387 | 14,278297 | 13,93824 | 14,338777 | 1,38517272 |
| 19,1551891 | 0,460141116 | 18,88823 | 19,272171 | 18,85237 | 19,423423 | 1,39232898 |
| 15,8869177 | 0,391597177 | 15,63667 | 16,047914 | 15,64414 | 16,0521 | 1,32831979 |
| 6,39426902 | 0,302782718 | 6,170536 | 6,567916 | 6,262304 | 6,4552449 | 1,22702144 |
| 6,68771322 | 0,260538067 | 6,443839 | 6,8830274 | 6,613176 | 6,705352 | 1,20220462 |
| 6,56402485 | 0,315799483 | 6,928497 | 6,37017 | 6,701698 | 6,4042449 | -1,3452649 |
| 17,6288032 | 0,335122371 | 17,49585 | 17,805045 | 17,31235 | 17,720199 | 1,28211379 |
| 7,88590122 | 0,301765739 | 7,69197 | 8,0660025 | 7,721956 | 7,9329883 | 1,22478824 |
| 1,76554481 | 0,165784353 | 1,588917 | 1,8535802 | 1,692129 | 1,8540855 | 1,15934494 |
| 5,96548111 | 0,668198718 | 5,597355 | 6,4505242 | 5,547438 | 5,9471619 | 1,54375757 |
| 5,84596447 | 0,247559021 | 5,679229 | 5,9507714 | 5,702789 | 5,9478267 | 1,19606035 |
| 8,07320464 | 0,896065829 | 7,652075 | 8,6775676 | 7,514513 | 8,0411449 | 1,71245086 |
| 9,76962984 | 0,583114148 | 9,619727 | 10,094801 | 9,172111 | 9,8653539 | 1,49916437 |
| 2,13309728 | 0,23534695 | 1,905481 | 2,2577415 | 2,164075 | 2,1584114 | 1,1276337 |
| 4,02220407 | 0,283393736 | 3,753155 | 4,1236718 | 4,117734 | 4,0718167 | 1,11906974 |
| 1,31859261 | 0,357206992 | 1,045066 | 1,5406937 | 1,21627 | 1,3459628 | 1,24199589 |
| 10,0422252 | 0,363533631 | 9,749659 | 10,346234 | 9,802313 | 10,070043 | 1,34924548 |
| 14,8583731 | 0,335024875 | 14,66785 | 15,03466 | 14,58484 | 14,972891 | 1,29902448 |
| 0,39743864 | 0,278201999 | 0,255531 | 0,6208554 | 0,355598 | 0,2750647 | 1,10373651 |
| 10,6251541 | 0,355269828 | 10,43764 | 10,790409 | 10,40087 | 10,720806 | 1,26256124 |
| 10,6111955 | 0,256998013 | 10,45369 | 10,747607 | 10,45255 | 10,676568 | 1,19662194 |
| 16,6897072 | 0,458869229 | 16,36683 | 16,915814 | 16,41247 | 16,859165 | 1,41209773 |
| 5,1664164 | 0,345510152 | 4,860774 | 5,4339783 | 4,963078 | 5,2271413 | 1,3366613 |
| 10,1646413 | 0,393974495 | 9,874822 | 10,335396 | 9,958269 | 10,329905 | 1,33431996 |
| 4,49054777 | 0,213472691 | 4,290566 | 4,5567821 | 4,620789 | 4,5029064 | 1,05275285 |

| | | | | | | |
|---|---|---|---|---|---|---|
| 11,1598924 | 0,324594035 | 10,98033 | 11,255293 | 10,94621 | 11,32625 | 1,25483843 |
| 15,0816889 | 0,394388984 | 14,91194 | 15,256169 | 14,73349 | 15,225523 | 1,33619813 |
| 17,1877491 | 0,405027819 | 16,96222 | 17,361778 | 16,87174 | 17,360218 | 1,36038809 |
| 6,97832454 | 0,244379629 | 6,821927 | 7,1147781 | 6,724071 | 7,1000706 | 1,26087531 |
| 10,5181158 | 0,372233833 | 10,291 | 10,701379 | 10,26068 | 10,645704 | 1,31740327 |
| 12,4760613 | 0,370241932 | 12,36877 | 12,649961 | 12,10647 | 12,580339 | 1,29911886 |
| 7,26992381 | 0,339540734 | 7,038981 | 7,4883104 | 7,050208 | 7,3359912 | 1,29016589 |
| 4,68442523 | 0,315273214 | 4,409394 | 4,8670966 | 4,547396 | 4,781214 | 1,27082077 |
| 8,47853526 | 0,317995754 | 8,2386 | 8,640533 | 8,357455 | 8,560731 | 1,23336903 |
| 2,44355488 | 0,156877491 | 2,279645 | 2,5447535 | 2,394562 | 2,4908359 | 1,13342688 |
| 0,99278624 | 0,226346392 | 0,773826 | 1,0914032 | 1,071136 | 1,0135517 | 1,09429078 |
| 6,97705123 | 0,270914939 | 6,701832 | 7,145675 | 6,780069 | 7,1268289 | 1,31521784 |
| 5,20384722 | 0,237606916 | 5,013422 | 5,3593795 | 5,054577 | 5,268632 | 1,2142001 |
| 6,37148721 | 0,234013678 | 6,235814 | 6,5038347 | 6,154254 | 6,4583319 | 1,21929649 |
| 2,31351524 | 0,274511229 | 2,078393 | 2,3902292 | 2,279977 | 2,4414358 | 1,17825182 |
| 7,74728992 | 0,34579032 | 7,558387 | 7,9425658 | 7,542883 | 7,7961701 | 1,24723449 |
| 4,76072838 | 0,274449709 | 4,499651 | 4,9043668 | 4,670426 | 4,86446 | 1,23061138 |
| 14,0057021 | 0,292487577 | 13,80628 | 14,181031 | 13,72416 | 14,133739 | 1,31236022 |
| 17,5345919 | 0,413682743 | 17,30669 | 17,61637 | 17,25106 | 17,800289 | 1,34672151 |
| 9,28365348 | 0,37289537 | 9,047164 | 9,4658732 | 9,093624 | 9,3800241 | 1,27681988 |
| 3,67277744 | 0,450852849 | 3,39936 | 3,7879065 | 3,431781 | 3,9116253 | 1,3511571 |
| 8,54160831 | 0,271421036 | 8,297027 | 8,7343379 | 8,377224 | 8,6168575 | 1,264417 |
| 5,80226677 | 0,275710083 | 5,637759 | 5,9784929 | 5,651986 | 5,8207952 | 1,19314664 |
| 2,71602736 | 0,21597352 | 2,584222 | 2,8491658 | 2,548246 | 2,7689646 | 1,18331265 |
| 18,3642085 | 0,404427667 | 18,11202 | 18,519581 | 18,08636 | 18,558829 | 1,35661701 |
| 1,50675188 | 0,313296534 | 1,21242 | 1,6570091 | 1,390122 | 1,6466036 | 1,27503392 |
| 15,4966164 | 0,572188312 | 15,15626 | 15,656714 | 15,12992 | 15,81382 | 1,50752037 |
| 6,08758042 | 0,376303944 | 5,752004 | 6,3027643 | 5,978119 | 6,1802763 | 1,29815141 |
| 10,6135493 | 0,3785894 | 10,36099 | 10,799986 | 10,32793 | 10,775875 | 1,35987306 |
| 7,11841266 | 0,468202672 | 6,732669 | 7,3300691 | 6,894071 | 7,3269115 | 1,42911286 |
| 4,53392058 | 0,31055851 | 4,712545 | 4,4103176 | 4,794551 | 4,3740352 | -1,2846466 |
| 6,20622436 | 0,269319323 | 5,957781 | 6,3669661 | 6,228358 | 6,2112309 | 1,14554093 |
| 9,05219116 | 0,470004165 | 8,674771 | 9,3374237 | 8,820946 | 9,1694667 | 1,41970071 |
| 6,04907566 | 0,308950332 | 5,762543 | 6,1578462 | 6,169015 | 6,0901408 | 1,11590553 |
| 11,3689303 | 0,38626955 | 11,15531 | 11,515008 | 11,12915 | 11,519081 | 1,29667212 |
| 1,68729735 | 0,234690864 | 1,485002 | 1,756921 | 1,656945 | 1,7939113 | 1,15224102 |
| 3,93456999 | 0,197104686 | 4,079887 | 3,7945112 | 4,063582 | 3,9017143 | -1,167661 |
| 6,82686494 | 0,317706301 | 7,077003 | 6,6357945 | 6,998913 | 6,7403035 | -1,2744801 |
| 8,77712833 | 0,445691762 | 8,439407 | 8,9355821 | 8,592528 | 8,9848068 | 1,36058462 |
| 11,9023519 | 0,269936418 | 11,75108 | 12,042046 | 11,72911 | 11,967248 | 1,20126419 |
| 3,39509094 | 0,242950502 | 3,161645 | 3,503891 | 3,335878 | 3,4984872 | 1,19120967 |
| 14,0475719 | 0,512570557 | 13,78812 | 14,277986 | 13,59607 | 14,262512 | 1,49294022 |
| 11,6902287 | 0,393575462 | 11,87172 | 11,479768 | 11,98386 | 11,612337 | -1,3029083 |
| 6,41327959 | 0,318471344 | 6,121303 | 6,5511948 | 6,433621 | 6,4837566 | 1,18100386 |
| 12,038158 | 0,474730937 | 11,70553 | 12,22698 | 11,82069 | 12,224793 | 1,37819544 |
| 7,42948712 | 0,241036232 | 7,22621 | 7,497545 | 7,350674 | 7,5678913 | 1,18449844 |
| 7,33826734 | 0,200334445 | 7,538388 | 7,2563881 | 7,313346 | 7,2813723 | -1,1149562 |
| 11,3078731 | 0,394032579 | 11,64745 | 11,046147 | 11,44113 | 11,253346 | -1,3145295 |
| 9,24824473 | 0,239732274 | 9,059648 | 9,393396 | 9,149852 | 9,2934062 | 1,17988895 |
| 8,75978771 | 0,339613544 | 8,982618 | 8,5594664 | 8,944407 | 8,699996 | -1,2603123 |
| 8,71750098 | 0,250856901 | 8,50043 | 8,8577138 | 8,670346 | 8,762049 | 1,168367 |
| 10,7530488 | 0,212188394 | 10,91437 | 10,637833 | 10,90706 | 10,661772 | -1,1982397 |
| 8,52486888 | 0,230775116 | 8,344164 | 8,6453939 | 8,400338 | 8,6085571 | 1,19310815 |

| | | | | | | |
|---|---|---|---|---|---|---|
| 5,17870517 | 0,539230902 | 4,730508 | 5,4762146 | 5,110878 | 5,243358 | 1,35575219 |
| 8,28525113 | 0,289628393 | 8,451045 | 8,1190611 | 8,518031 | 8,2040865 | -1,2508982 |
| 11,5944808 | 0,264380123 | 11,75349 | 11,482245 | 11,76445 | 11,492022 | -1,2073451 |
| 14,3623518 | 0,252520617 | 14,53311 | 14,245793 | 14,50061 | 14,274121 | -1,1949114 |
| 6,1115762 | 0,33857338 | 5,925368 | 6,2502567 | 5,844854 | 6,2634698 | 1,29392347 |
| 1,97596833 | 0,312559165 | 1,779996 | 2,1122083 | 1,715986 | 2,1350469 | 1,29741227 |
| 2,7589025 | 0,332102855 | 2,576679 | 2,9017663 | 2,475065 | 2,9126586 | 1,30255165 |
| 1,33216544 | 0,370859629 | 1,514515 | 1,2171854 | 1,677586 | 1,1078915 | -1,3505176 |
| 0,99759315 | 0,282041411 | 0,762799 | 1,1513159 | 0,887751 | 1,0786065 | 1,22237456 |
| 3,94862902 | 0,38628127 | 3,737299 | 4,1470784 | 3,691142 | 4,0446126 | 1,30280854 |
| 17,8858925 | 0,479551908 | 17,63168 | 18,184094 | 17,51968 | 17,963861 | 1,41254681 |
| 7,06531177 | 0,264510307 | 6,90376 | 7,1936202 | 6,883348 | 7,1578388 | 1,21602718 |
| 13,7351653 | 0,295148856 | 13,95032 | 13,572008 | 13,88772 | 13,656538 | -1,2352026 |
| 6,19162344 | 0,268613472 | 6,38528 | 6,058346 | 6,30476 | 6,1190661 | -1,1944232 |
| 13,3672683 | 0,27657266 | 13,58006 | 13,22882 | 13,50598 | 13,269312 | -1,2259915 |
| 11,0185085 | 0,304138514 | 11,27318 | 10,818489 | 11,18157 | 10,944221 | -1,2710507 |
| 18,2990022 | 0,404269705 | 18,0363 | 18,406481 | 17,99763 | 18,574142 | 1,38832321 |
| 3,28254351 | 0,184798789 | 3,333838 | 3,2130959 | 3,501858 | 3,1906913 | -1,1614722 |
| 14,2085635 | 0,220516688 | 14,12115 | 14,292238 | 14,00086 | 14,30708 | 1,17989302 |
| 6,96864506 | 0,218985875 | 7,050214 | 6,8898267 | 7,175433 | 6,8698206 | -1,1752762 |
| 4,44794658 | 0,280545476 | 4,614603 | 4,4033088 | 4,618251 | 4,2576717 | -1,2192014 |
| 6,5531218 | 0,225335829 | 6,669434 | 6,4937462 | 6,702622 | 6,4358071 | -1,1657443 |
| 8,86370333 | 0,431994683 | 9,183331 | 8,9111632 | 8,942985 | 8,487499 | -1,2868348 |
| 15,1482101 | 0,199592991 | 15,00908 | 15,180582 | 15,01223 | 15,309209 | 1,17628557 |
| 2,99588949 | 0,438769605 | 3,126463 | 3,172008 | 3,094783 | 2,6142233 | -1,1627232 |
| 3,82233453 | 0,235667806 | 3,918618 | 3,7795481 | 4,045718 | 3,6578075 | -1,200379 |
| 12,0656425 | 0,223651291 | 12,16489 | 11,963099 | 12,25523 | 11,990576 | -1,1754608 |
| 15,5517709 | 0,218937663 | 15,41619 | 15,607033 | 15,40679 | 15,687685 | 1,17761348 |
| 11,3854849 | 0,216621689 | 11,23457 | 11,46151 | 11,24192 | 11,508672 | 1,18661061 |
| 17,7367742 | 0,314053734 | 17,55138 | 17,811126 | 17,51214 | 17,939923 | 1,26906705 |
| 14,9221002 | 0,293048111 | 14,813 | 15,009359 | 14,64638 | 15,075555 | 1,24208736 |
| 0,10053067 | 0,198674869 | 0,243024 | 0,0565929 | 0,227807 | -0,044229 | -1,1722119 |
| 16,2646623 | 0,359171543 | 16,10386 | 16,35467 | 15,95005 | 16,482098 | 1,31168992 |
| 0,20795115 | 0,176416132 | 0,270172 | 0,164584 | 0,379232 | 0,1043352 | -1,1409556 |
| 15,5498296 | 0,265351656 | 15,33638 | 15,588946 | 15,45207 | 15,742976 | 1,20725844 |
| 4,47552922 | 0,228261539 | 4,651748 | 4,3902999 | 4,641955 | 4,3281633 | -1,2206248 |
| 0,31617619 | 0,251984307 | 0,443352 | 0,2666142 | 0,492076 | 0,1620112 | -1,1920141 |
| -1,57640529 | 0,172892446 | -1,489979 | -1,6233657 | -1,41684 | -1,689252 | -1,1510075 |
| 15,7528515 | 0,326660395 | 15,62233 | 15,820214 | 15,4641 | 15,956214 | 1,27015199 |
| 15,7966455 | 0,267893556 | 15,67243 | 15,899542 | 15,53715 | 15,934446 | 1,24160092 |
| 2,88970521 | 0,25235324 | 3,001054 | 2,812585 | 3,075878 | 2,7758991 | -1,1844556 |
| 17,3177252 | 0,284031674 | 17,14297 | 17,282744 | 17,15579 | 17,605404 | 1,22662412 |
| 6,22412058 | 0,23795133 | 6,33775 | 6,1789522 | 6,404238 | 6,0736668 | -1,1848339 |
| 11,3132859 | 0,197236477 | 11,18127 | 11,368246 | 11,14108 | 11,462872 | 1,19282676 |
| 8,36806954 | 0,225311138 | 8,481106 | 8,2834916 | 8,572668 | 8,2507229 | -1,1972959 |
| 7,22877872 | 0,19501336 | 7,327505 | 7,1850947 | 7,383321 | 7,1045567 | -1,1571592 |
| 1,264104 | 0,196088447 | 1,337559 | 1,2262698 | 1,440884 | 1,1410001 | -1,1531553 |
| 4,72324921 | 0,248729676 | 4,867522 | 4,6751851 | 4,879959 | 4,5642679 | -1,1925207 |
| 11,9208828 | 0,231989303 | 11,79079 | 12,016968 | 11,71171 | 12,041657 | 1,21256238 |
| -1,71465328 | 0,184367007 | -1,600625 | -1,7964544 | -1,5355 | -1,820906 | -1,1814968 |
| 1,35614152 | 0,210848242 | 1,45888 | 1,2768973 | 1,568417 | 1,236542 | -1,1949321 |
| 8,05406984 | 0,370308181 | 8,256371 | 8,0037405 | 8,311887 | 7,7878187 | -1,3088951 |
| 2,6719496 | 0,29366037 | 2,825304 | 2,5940378 | 2,868099 | 2,5174029 | -1,2234721 |

| | | | | | | |
|---|---|---|---|---|---|---|
| 16,0547076 | 0,247295899 | 15,94467 | 16,128361 | 15,83843 | 16,189621 | 1,20367036 |
| 11,1567447 | 0,315841073 | 10,98738 | 11,182767 | 10,95409 | 11,391067 | 1,24503025 |
| 2,33713417 | 0,21360455 | 2,427606 | 2,2262839 | 2,543541 | 2,2694093 | -1,1791333 |
| 11,21235 | 0,251482839 | 11,07229 | 11,319659 | 11,03606 | 11,308403 | 1,19735899 |
| 4,00602894 | 0,161534613 | 4,080487 | 3,9556578 | 4,171804 | 3,9037197 | -1,1458807 |
| 8,20021349 | 0,285265791 | 8,049409 | 8,2973469 | 7,9959 | 8,334425 | 1,22538169 |
| 5,6786377 | 0,217636705 | 5,830825 | 5,574871 | 5,910649 | 5,5345916 | -1,2448793 |
| 9,36202452 | 0,265983898 | 9,530126 | 9,2746963 | 9,539193 | 9,2176307 | -1,2213665 |
| 11,7935431 | 0,310954909 | 11,6132 | 11,848241 | 11,58802 | 12,004508 | 1,2533262 |
| 4,46877392 | 0,337326432 | 4,725192 | 4,4903438 | 4,545283 | 4,1790295 | -1,2316144 |
| 16,0391491 | 0,259827456 | 15,92048 | 16,169769 | 15,74883 | 16,157467 | 1,25611076 |
| 17,1263722 | 0,301720489 | 17,01039 | 17,252262 | 16,83951 | 17,246006 | 1,25195624 |
| 6,45670685 | 0,437867211 | 6,326789 | 6,2677381 | 7,018738 | 6,456681 | -1,2401837 |
| 8,80128635 | 0,243364542 | 8,648595 | 8,8499317 | 8,660577 | 8,9571253 | 1,18833576 |
| 2,32985983 | 0,279247261 | 2,527418 | 2,2744115 | 2,433315 | 2,1664004 | -1,1974459 |
| 7,81390964 | 0,233937114 | 7,670061 | 7,9132162 | 7,622975 | 7,931574 | 1,21072992 |
| 18,9814502 | 0,376618615 | 18,82402 | 19,074721 | 18,60855 | 19,227078 | 1,35154874 |
| -1,07621426 | 0,176871508 | -0,992768 | -1,1351389 | -0,90552 | -1,178853 | -1,1549684 |
| 13,2906784 | 0,310713787 | 13,12126 | 13,316346 | 13,06955 | 13,536565 | 1,25793162 |
| 10,8961827 | 0,211450268 | 10,74796 | 10,911591 | 10,79757 | 11,062849 | 1,1602639 |
| 3,43857751 | 0,191658446 | 3,476321 | 3,3721149 | 3,666932 | 3,349238 | -1,1574502 |
| 3,54532777 | 0,248742764 | 3,719524 | 3,5131515 | 3,6429 | 3,3773288 | -1,1777 |
| 9,45123997 | 0,228921529 | 9,572982 | 9,3758175 | 9,630329 | 9,3308129 | -1,1878398 |
| 5,77631948 | 0,224900109 | 5,8848 | 5,7477283 | 5,986568 | 5,592271 | -1,2022065 |
| 13,6601999 | 0,291571393 | 13,53204 | 13,777146 | 13,37549 | 13,799625 | 1,26104515 |
| 2,13170577 | 0,250315307 | 1,979503 | 2,2046562 | 1,971057 | 2,2699273 | 1,19914989 |
| 16,4686549 | 0,268694823 | 16,29507 | 16,548137 | 16,24873 | 16,652784 | 1,25576343 |
| 12,0707465 | 0,266456851 | 11,95893 | 12,154744 | 11,83813 | 12,204558 | 1,21513568 |
| 5,50621619 | 0,333589286 | 5,697592 | 5,5203624 | 5,687673 | 5,217697 | -1,2514519 |
| 5,04270491 | 0,25898163 | 4,890324 | 5,134936 | 4,874011 | 5,1627682 | 1,20304026 |
| 6,55070018 | 0,257191453 | 6,685178 | 6,4895245 | 6,732509 | 6,4007192 | -1,200572 |
| 9,27368973 | 0,2603498 | 9,144846 | 9,3142931 | 9,080356 | 9,4504829 | 1,2056297 |
| 5,69675799 | 0,217234079 | 5,569376 | 5,7616584 | 5,51953 | 5,8334888 | 1,19178207 |
| 9,9181321 | 0,216803942 | 9,82184 | 10,010298 | 9,73422 | 9,9997287 | 1,1703851 |
| 14,3489183 | 0,21164471 | 14,22725 | 14,382447 | 14,19722 | 14,503123 | 1,17328403 |
| 13,0449379 | 0,248934548 | 12,93299 | 13,171792 | 12,83661 | 13,11287 | 1,19543333 |
| 10,6882404 | 0,254284235 | 10,53946 | 10,801281 | 10,45837 | 10,816979 | 1,23989353 |
| 6,3745756 | 0,239752313 | 6,48968 | 6,3182809 | 6,544125 | 6,2425607 | -1,1781161 |
| 7,79287193 | 0,232396019 | 7,689576 | 7,8243678 | 7,589615 | 7,9648325 | 1,19333958 |
| 9,52734238 | 0,251295239 | 9,639251 | 9,4690689 | 9,734731 | 9,3777147 | -1,20047 |
| -1,39203113 | 0,221040602 | -1,429094 | -1,5086292 | -1,07818 | -1,408919 | -1,1527951 |
| 3,60643814 | 0,207542214 | 3,720758 | 3,4840066 | 3,81751 | 3,5295411 | -1,1994392 |
| 2,18129517 | 0,194602694 | 2,241225 | 2,116722 | 2,388822 | 2,0833263 | -1,1607035 |
| 6,4626189 | 0,296988064 | 6,283355 | 6,5426551 | 6,302574 | 6,6149763 | 1,21912906 |
| 5,21766289 | 0,185640643 | 5,363798 | 5,1217758 | 5,383159 | 5,1092151 | -1,1958056 |
| 3,27264752 | 0,374183695 | 3,372948 | 3,1241559 | 3,63133 | 3,1503725 | -1,2877699 |
| 5,08906877 | 0,299087266 | 5,255687 | 5,0883557 | 5,285879 | 4,830213 | -1,2409961 |
| 6,64471639 | 0,256089089 | 6,498985 | 6,7349609 | 6,489205 | 6,7536015 | 1,18936062 |
| 9,23334457 | 0,289036704 | 9,382813 | 9,1363254 | 9,424233 | 9,1081863 | -1,2152617 |
| 13,8029117 | 0,278380196 | 13,68872 | 13,926296 | 13,55203 | 13,902442 | 1,22602959 |
| 20,1964011 | 0,30378002 | 20,11464 | 20,291744 | 19,83782 | 20,366635 | 1,27717687 |
| 11,4929597 | 0,354059385 | 11,34752 | 11,585579 | 11,19992 | 11,681264 | 1,28315926 |
| 9,99710332 | 0,253921652 | 9,829151 | 10,059335 | 9,870076 | 10,1414 | 1,18982883 |

| | | | | | | |
|---|---|---|---|---|---|---|
| 5,57473124 | 0,219969109 | 5,695145 | 5,4876453 | 5,780523 | 5,4534077 | -1,2035595 |
| 3,61858144 | 0,219017162 | 3,81378 | 3,5553117 | 3,729114 | 3,4622668 | -1,1996867 |
| 7,19140256 | 0,213916403 | 7,348175 | 7,1196501 | 7,331902 | 7,0599492 | -1,1894041 |
| 2,59224806 | 0,18974914 | 2,687274 | 2,5533164 | 2,768706 | 2,4523194 | -1,1689166 |
| 4,34765521 | 0,328515636 | 4,550433 | 4,326277 | 4,567261 | 4,0691845 | -1,2844194 |
| 12,9730396 | 0,258995065 | 12,80537 | 13,063445 | 12,82913 | 13,093418 | 1,19846005 |
| 9,25414569 | 0,249241482 | 9,377625 | 9,1571035 | 9,472813 | 9,1344381 | -1,2137308 |
| 10,9786368 | 0,274213205 | 11,0773 | 10,841108 | 11,2269 | 10,910855 | -1,2109292 |
| 12,4383827 | 0,286678126 | 12,31552 | 12,541063 | 12,1475 | 12,594129 | 1,2623281 |
| 4,65731316 | 0,226601345 | 4,814589 | 4,5938516 | 4,789191 | 4,5206558 | -1,1847941 |
| 7,67554935 | 0,162694823 | 7,786343 | 7,5773536 | 7,878293 | 7,5775637 | -1,1932193 |
| 8,19213828 | 0,338515401 | 8,414523 | 8,19099 | 8,337524 | 7,9172581 | -1,249975 |
| 6,41694647 | 0,745056666 | 5,924272 | 6,2597535 | 6,333706 | 7,0742958 | 1,45199422 |
| 14,0005155 | 0,257915569 | 13,85648 | 14,106664 | 13,82759 | 14,099323 | 1,19827431 |
| 9,02782118 | 0,277381812 | 9,124987 | 8,9339604 | 9,294264 | 8,897997 | -1,225735 |
| 7,79411938 | 0,251222055 | 7,880787 | 7,7234825 | 8,046456 | 7,6538141 | -1,2099717 |
| 6,03354659 | 0,313780065 | 6,246767 | 5,9962265 | 6,21367 | 5,7890192 | -1,263649 |
| 2,64398495 | 0,196629073 | 2,749985 | 2,5538129 | 2,849306 | 2,5388987 | -1,1919218 |
| 4,32323153 | 0,242553519 | 4,463372 | 4,2344697 | 4,577554 | 4,158033 | -1,25198 |
| 6,47112323 | 0,331440454 | 6,298761 | 6,5245793 | 6,262022 | 6,6789448 | 1,24951704 |
| 7,89471149 | 0,356781175 | 8,079735 | 8,0201465 | 7,91952 | 7,5720342 | -1,1515183 |
| 3,85453347 | 0,199068199 | 3,99904 | 3,7820174 | 4,004222 | 3,7289087 | -1,1860528 |
| 0,84784759 | 0,158696348 | 0,912759 | 0,8031539 | 1,014746 | 0,7461668 | -1,1400458 |
| 8,46306073 | 0,265595187 | 8,548463 | 8,3840174 | 8,715087 | 8,3341046 | -1,2080786 |
| 0,15432692 | 0,225057444 | 0,243639 | 0,1181597 | 0,358231 | -0,00053 | -1,1827298 |
| 17,527285 | 0,311851173 | 17,42362 | 17,64692 | 17,20325 | 17,666257 | 1,26852515 |
| 16,1528418 | 0,29533654 | 16,01792 | 16,232657 | 15,90544 | 16,32019 | 1,24379169 |
| 19,6901064 | 0,307712945 | 19,52359 | 19,769364 | 19,41037 | 19,904377 | 1,2922532 |
| 11,7733432 | 0,2373165 | 11,65517 | 11,890316 | 11,55226 | 11,866076 | 1,20956046 |
| 11,64701 | 0,241383011 | 11,52888 | 11,727324 | 11,44532 | 11,77206 | 1,1996332 |
| 11,1232535 | 0,325825489 | 10,96014 | 11,183732 | 10,88055 | 11,334943 | 1,2648699 |
| 12,8026878 | 0,274488947 | 12,69885 | 12,897638 | 12,52269 | 12,945006 | 1,24018084 |
| 10,6304544 | 0,298494235 | 10,47358 | 10,732818 | 10,42641 | 10,763393 | 1,2295359 |
| -0,76115097 | 0,180238322 | -0,646369 | -0,780989 | -0,63758 | -0,909054 | -1,1511274 |
| 15,3378815 | 0,349140545 | 15,18227 | 15,434087 | 15,0478 | 15,528756 | 1,28912165 |
| 10,9574192 | 0,202902594 | 11,01619 | 10,836288 | 11,19991 | 10,907332 | -1,1779158 |
| 5,81792996 | 0,201790698 | 5,908993 | 5,813471 | 5,988385 | 5,6436047 | -1,1648554 |
| 8,97738694 | 0,247786631 | 8,80778 | 9,0518015 | 8,760687 | 9,1622756 | 1,25076024 |
| 11,7634321 | 0,286662354 | 11,85051 | 11,612196 | 12,04962 | 11,699186 | -1,226353 |
| 4,69330476 | 0,194301734 | 4,780975 | 4,5903989 | 4,891004 | 4,6236529 | -1,1719925 |
| 15,8613943 | 0,36915934 | 15,72884 | 15,954436 | 15,46502 | 16,100238 | 1,34761216 |
| 3,91252178 | 0,338374966 | 4,072923 | 4,0397808 | 3,926637 | 3,6150007 | -1,1269232 |
| 6,50321363 | 0,2516831 | 6,624568 | 6,4500076 | 6,683538 | 6,355715 | -1,1901898 |
| 20,317134 | 0,34108236 | 20,20165 | 20,399337 | 19,95298 | 20,535142 | 1,31032306 |
| 4,11548393 | 0,315489926 | 4,311912 | 4,1093729 | 4,271363 | 3,8623256 | -1,236094 |
| 6,39636956 | 0,325334142 | 6,492724 | 6,2407848 | 6,746007 | 6,2913875 | -1,2774611 |
| 13,0490504 | 0,313128591 | 12,86384 | 13,105558 | 12,81246 | 13,280628 | 1,27893716 |
| 9,54247671 | 0,280619584 | 9,687208 | 9,4388261 | 9,745514 | 9,4220132 | -1,2192056 |
| 4,73427796 | 0,655341225 | 4,321994 | 4,4997775 | 4,7773 | 5,3403068 | 1,29270698 |
| 8,26282196 | 0,172527018 | 8,328138 | 8,2042356 | 8,43829 | 8,1723261 | -1,1446711 |
| 15,606166 | 0,266848916 | 15,47514 | 15,712232 | 15,36093 | 15,7374 | 1,236944 |
| 4,62488913 | 0,333098662 | 4,811612 | 4,5914838 | 4,809278 | 4,3956279 | -1,2456416 |
| 6,02445791 | 0,242673603 | 6,197371 | 5,9733532 | 6,16941 | 5,8518357 | -1,2064736 |

| | | | | | | |
|---|---|---|---|---|---|---|
| 15,2225724 | 0,291148445 | 15,02371 | 15,253558 | 15,07753 | 15,441447 | 1,22848597 |
| 1,91497093 | 0,228944638 | 2,019794 | 1,7945666 | 2,142753 | 1,8336874 | -1,2034251 |
| 7,27950495 | 0,753272362 | 6,801368 | 7,1217403 | 7,16958 | 7,9411937 | 1,46002473 |
| 4,899154 | 0,631763164 | 5,080561 | 5,1601066 | 5,019619 | 4,3595356 | -1,2228684 |
| 4,19530594 | 0,204430624 | 4,094049 | 4,3146432 | 3,958668 | 4,2801528 | 1,20667718 |
| 1,82574943 | 0,272025873 | 1,96124 | 1,7176727 | 2,006608 | 1,7317595 | -1,1968214 |
| 7,21822367 | 0,304635772 | 7,358319 | 7,114576 | 7,446731 | 7,086416 | -1,2328768 |
| 17,5292359 | 0,250717512 | 17,32337 | 17,56814 | 17,41597 | 17,7255 | 1,21180088 |
| 4,66097746 | 0,21887036 | 4,748031 | 4,625032 | 4,888732 | 4,4934632 | -1,1967602 |
| 15,1076555 | 0,277800969 | 15,02248 | 15,261828 | 14,81156 | 15,172705 | 1,23135524 |
| 2,98287675 | 0,246768498 | 2,841252 | 3,0151541 | 2,826427 | 3,1583945 | 1,19162854 |
| 3,12259883 | 0,214420618 | 3,070697 | 3,1925141 | 2,892608 | 3,2208113 | 1,16878538 |
| 8,13808889 | 0,253516095 | 8,017919 | 8,2464127 | 7,89161 | 8,2581322 | 1,2290196 |
| -0,83695234 | 0,226400916 | -0,697299 | -0,9337884 | -0,67133 | -0,938831 | -1,190855 |
| 4,46828161 | 0,215663922 | 4,559333 | 4,4350298 | 4,653671 | 4,3195565 | -1,1721919 |
| 3,35250002 | 0,250069036 | 3,402121 | 3,4200064 | 3,51998 | 3,1288262 | -1,1381056 |
| 6,37864522 | 0,269879221 | 6,240109 | 6,4668464 | 6,19052 | 6,5034352 | 1,205663 |
| 3,86720546 | 0,226636046 | 3,692142 | 3,9307239 | 3,691976 | 4,0449249 | 1,22753611 |

| T2.vs.C-Old.FC | T2.vs.C-Young.FC | diff.Old | diff.Young | diff.Both |
|---|---|---|---|---|
| 1,337696154 | 1,451964055 | Yes | Yes | Yes |
| -1,4425677 | -1,744513604 | Yes | Yes | Yes |
| 1,211382655 | 1,177390815 | Yes | No | No |
| -1,264812096 | -1,095997763 | Yes | No | No |
| -1,301424339 | -1,086153011 | Yes | No | No |
| -1,212975413 | -1,272140285 | Yes | Yes | Yes |
| -1,236385646 | -1,479556144 | Yes | Yes | Yes |
| 1,561563802 | 1,115719923 | Yes | No | No |
| -1,213140622 | -1,310988146 | Yes | Yes | Yes |
| -1,269454295 | -1,051015728 | Yes | No | No |
| -1,598656119 | -1,089140708 | Yes | No | No |
| -1,225896736 | -1,071799839 | Yes | No | No |
| -1,274128413 | -1,119512218 | Yes | No | No |
| -1,221755562 | -1,172965076 | Yes | No | No |
| 1,329848547 | 1,230487537 | Yes | Yes | Yes |
| -1,458779943 | -1,255238093 | Yes | Yes | Yes |
| -1,220145603 | -1,122214667 | Yes | No | No |
| -1,212655513 | -1,143858286 | Yes | No | No |
| 1,52853025 | 1,43859192 | Yes | Yes | Yes |
| 1,38717325 | 1,214556152 | Yes | Yes | Yes |
| 1,334186087 | 1,161557175 | Yes | No | No |
| 1,582811499 | 1,393297133 | Yes | Yes | Yes |
| 1,275822004 | 1,154984199 | Yes | No | No |
| 1,326039832 | -1,011752519 | Yes | No | No |
| -1,323989698 | -1,354473511 | Yes | Yes | Yes |
| -1,211011249 | -1,116561298 | Yes | No | No |
| -1,472723863 | -1,057391018 | Yes | No | No |
| 1,245655696 | 1,122463373 | Yes | No | No |
| 1,264987036 | 1,218945223 | Yes | Yes | Yes |
| 1,337027555 | 1,254650227 | Yes | Yes | Yes |
| -1,413966852 | 1,003282305 | Yes | No | No |
| -1,278947788 | -1,070815264 | Yes | No | No |
| -1,204421096 | -1,045356527 | Yes | No | No |
| -1,214364 | -1,162106308 | Yes | No | No |
| -1,230070611 | -1,162394984 | Yes | No | No |
| -1,330577475 | -1,120931551 | Yes | No | No |
| 1,34955768 | 1,222954557 | Yes | Yes | Yes |
| -1,514324268 | -1,036373007 | Yes | No | No |
| -1,242705613 | -1,103490687 | Yes | No | No |
| -1,254553418 | -1,142170476 | Yes | No | No |
| -1,209996237 | -1,195136417 | Yes | No | No |
| -1,229319622 | -1,2296439 | Yes | Yes | Yes |
| -1,25701283 | -1,252907758 | Yes | Yes | Yes |
| -1,248673626 | -1,069361329 | Yes | No | No |
| -1,324804096 | -1,08730239 | Yes | No | No |
| -1,391760053 | -1,499043174 | Yes | Yes | Yes |
| -1,249525854 | -1,113306208 | Yes | No | No |
| -1,250381923 | -1,192824501 | Yes | No | No |
| 1,298645624 | 1,318360043 | Yes | Yes | Yes |
| 1,279028178 | 1,122523781 | Yes | No | No |
| 1,306140701 | 1,107148704 | Yes | No | No |
| -1,291353949 | -1,200913057 | Yes | Yes | Yes |

| | | | | |
|---|---|---|---|---|
| -1,34387056 | -1,072251482 | Yes | No | No |
| -1,245269507 | -1,12278792 | Yes | No | No |
| 1,206706913 | 1,352204727 | Yes | Yes | Yes |
| -1,237754359 | -1,052258062 | Yes | No | No |
| -1,232756871 | -1,19404055 | Yes | No | No |
| -1,465041927 | -1,094401956 | Yes | No | No |
| 1,225737597 | 1,060027333 | Yes | No | No |
| -1,202222601 | -1,095528096 | Yes | No | No |
| -1,207388502 | -1,122732271 | Yes | No | No |
| -1,256826347 | -1,058655139 | Yes | No | No |
| 1,327015598 | 1,115720003 | Yes | No | No |
| -1,276578581 | -1,194607319 | Yes | No | No |
| -1,206401867 | -1,152436507 | Yes | No | No |
| -1,239314442 | -1,18860735 | Yes | No | No |
| -1,237039893 | -1,122002791 | Yes | No | No |
| 1,219380759 | 1,315193606 | Yes | Yes | Yes |
| -1,311197222 | -1,116977199 | Yes | No | No |
| -1,36533509 | -1,129379275 | Yes | No | No |
| -1,23016747 | -1,075445679 | Yes | No | No |
| -1,333778678 | 1,112486463 | Yes | No | No |
| -1,369630693 | -1,105398739 | Yes | No | No |
| 1,223830946 | 1,098056315 | Yes | No | No |
| 1,248021198 | 1,232768674 | Yes | Yes | Yes |
| -1,323745109 | -1,12525821 | Yes | No | No |
| 1,34699165 | 1,232487367 | Yes | Yes | Yes |
| -1,224109714 | -1,112723878 | Yes | No | No |
| 1,422978888 | 1,022669828 | Yes | No | No |
| -1,237218785 | -1,195989721 | Yes | No | No |
| -1,223352844 | -1,157581631 | Yes | No | No |
| -1,526691285 | -1,096153016 | Yes | No | No |
| -1,206202975 | -1,061718387 | Yes | No | No |
| -1,357194805 | -1,101420917 | Yes | No | No |
| -1,285686923 | -1,163277227 | Yes | No | No |
| -4,141451337 | -1,895324398 | Yes | Yes | Yes |
| 1,209366918 | 1,159549898 | Yes | No | No |
| -1,268361743 | -1,095554095 | Yes | No | No |
| -1,271633601 | 1,078873919 | Yes | No | No |
| 1,226438065 | 1,324110311 | Yes | Yes | Yes |
| -1,304261669 | -1,14123681 | Yes | No | No |
| -1,336174165 | -1,040617959 | Yes | No | No |
| -1,3887337 | -1,003355642 | Yes | No | No |
| -1,241790194 | -1,109538503 | Yes | No | No |
| -1,358350603 | -1,167707275 | Yes | No | No |
| -1,329633201 | -1,254424966 | Yes | Yes | Yes |
| -1,244421286 | -1,044448725 | Yes | No | No |
| -1,21250048 | -1,149603455 | Yes | No | No |
| 1,408701352 | 1,367641066 | Yes | Yes | Yes |
| 1,262775824 | 1,217460783 | Yes | Yes | Yes |
| -1,231471882 | -1,028253349 | Yes | No | No |
| -1,200175169 | -1,116809356 | Yes | No | No |
| 1,295965465 | 1,124418655 | Yes | No | No |
| -1,3425663 | -1,062928562 | Yes | No | No |
| -1,229301498 | -1,09418954 | Yes | No | No |

| | | | | |
|---|---|---|---|---|
| 1,211174651 | 1,184944428 | Yes | No | No |
| -1,281236016 | -1,010025313 | Yes | No | No |
| -1,275935583 | -1,099536613 | Yes | No | No |
| -1,239670774 | -1,197768151 | Yes | No | No |
| 1,443784422 | 1,489426925 | Yes | Yes | Yes |
| -1,627672129 | -1,330594992 | Yes | Yes | Yes |
| -1,320344695 | -1,091823895 | Yes | No | No |
| -1,246060349 | -1,248147632 | Yes | Yes | Yes |
| -1,289076223 | -1,185149192 | Yes | No | No |
| -1,421126483 | -1,153792327 | Yes | No | No |
| -1,349819733 | -1,156774639 | Yes | No | No |
| -1,343435697 | -1,127750911 | Yes | No | No |
| -1,520739398 | -1,2433429 | Yes | Yes | Yes |
| -1,213513194 | -1,153686238 | Yes | No | No |
| -1,446483871 | -1,426956691 | Yes | Yes | Yes |
| 1,414827865 | 1,361860413 | Yes | Yes | Yes |
| -1,328916371 | 1,050460726 | Yes | No | No |
| -1,300712129 | -1,208286575 | Yes | Yes | Yes |
| -1,234972797 | -1,061003087 | Yes | No | No |
| -1,2461733 | -1,02823438 | Yes | No | No |
| -1,246553756 | -1,154960072 | Yes | No | No |
| -1,35795379 | 1,022013414 | Yes | No | No |
| -1,384047594 | -1,108270829 | Yes | No | No |
| 1,222825815 | 1,263498229 | Yes | Yes | Yes |
| 1,292254012 | 1,217055324 | Yes | Yes | Yes |
| -1,215651795 | -1,207663671 | Yes | Yes | Yes |
| -1,284573962 | -1,187450236 | Yes | No | No |
| 1,299569887 | 1,280899987 | Yes | Yes | Yes |
| -1,315618534 | -1,015905295 | Yes | No | No |
| 1,234619627 | 1,134191014 | Yes | No | No |
| 1,42029437 | 1,215195041 | Yes | Yes | Yes |
| 1,235403168 | 1,239042928 | Yes | Yes | Yes |
| -1,446522657 | -1,255168478 | Yes | Yes | Yes |
| 1,281665383 | 1,361896082 | Yes | Yes | Yes |
| -1,218745956 | -1,095698751 | Yes | No | No |
| -1,462496244 | -1,003655812 | Yes | No | No |
| 1,288818796 | 1,074304855 | Yes | No | No |
| -1,445056841 | -1,13548538 | Yes | No | No |
| 1,297086183 | 1,298983376 | Yes | Yes | Yes |
| 1,256317445 | 1,244348539 | Yes | Yes | Yes |
| 1,247204861 | 1,221430192 | Yes | Yes | Yes |
| 1,408116342 | 1,373960529 | Yes | Yes | Yes |
| 1,218444701 | 1,247049278 | Yes | Yes | Yes |
| -1,242505867 | -1,038703868 | Yes | No | No |
| -1,29292414 | -1,210261289 | Yes | Yes | Yes |
| 1,246486066 | 1,209078592 | Yes | Yes | Yes |
| 1,245375682 | 1,327723433 | Yes | Yes | Yes |
| 1,244211811 | 1,364848418 | Yes | Yes | Yes |
| 1,277356328 | 1,230637774 | Yes | Yes | Yes |
| -1,520506882 | -1,130738699 | Yes | No | No |
| -1,414460133 | -1,151627666 | Yes | No | No |
| -1,215240183 | -1,173700247 | Yes | No | No |
| 1,24135562 | 1,297805511 | Yes | Yes | Yes |

| | | | | |
|---|---|---|---|---|
| 1,218232411 | 1,110640225 | Yes | No | No |
| 1,257990208 | 1,123613338 | Yes | No | No |
| 1,541568152 | 1,090139613 | Yes | No | No |
| -1,328370406 | -1,146707355 | Yes | No | No |
| -1,392719636 | -1,097393673 | Yes | No | No |
| 1,296514513 | 1,046653597 | Yes | No | No |
| -1,23169571 | -1,273197009 | Yes | Yes | Yes |
| -1,340683068 | 1,004683468 | Yes | No | No |
| 1,294326926 | 1,604367182 | Yes | Yes | Yes |
| 1,274382851 | 1,162671819 | Yes | No | No |
| 1,300625594 | 1,408162583 | Yes | Yes | Yes |
| 1,33649964 | 1,300015052 | Yes | Yes | Yes |
| 1,348981195 | 1,272722498 | Yes | Yes | Yes |
| -1,366953646 | -1,418761304 | Yes | Yes | Yes |
| 1,238110911 | 1,219924165 | Yes | Yes | Yes |
| 1,201841619 | 1,177255909 | Yes | No | No |
| 1,209704637 | 1,350279515 | Yes | Yes | Yes |
| 1,287838748 | 1,096574097 | Yes | No | No |
| 1,300063159 | 1,446878921 | Yes | Yes | Yes |
| -1,23101714 | -1,134604952 | Yes | No | No |
| -1,544963841 | -1,08357387 | Yes | No | No |
| -1,298417195 | -1,118849412 | Yes | No | No |
| -1,317068497 | -1,202917154 | Yes | Yes | Yes |
| -1,247619294 | -1,142436154 | Yes | No | No |
| -1,231044818 | -1,100723117 | Yes | No | No |
| -1,422194273 | -1,05293638 | Yes | No | No |
| 1,338411231 | 1,370836057 | Yes | Yes | Yes |
| -1,229925438 | -1,148623226 | Yes | No | No |
| -1,433293628 | -1,268818791 | Yes | Yes | Yes |
| -1,465588328 | -1,158209122 | Yes | No | No |
| -1,311705648 | -1,208231054 | Yes | Yes | Yes |
| 1,22343314 | -1,04049735 | Yes | No | No |
| 1,329844032 | 1,028833465 | Yes | No | No |
| 1,403570851 | -1,068607369 | Yes | No | No |
| -1,214690241 | -1,154593105 | Yes | No | No |
| -1,216774325 | -1,178718033 | Yes | No | No |
| 1,277023051 | 1,274814886 | Yes | Yes | Yes |
| -1,264769825 | -1,186586392 | Yes | No | No |
| -1,277355967 | -1,20331187 | Yes | Yes | Yes |
| -1,748992866 | -1,764146001 | Yes | Yes | Yes |
| -1,541847071 | -1,037941003 | Yes | No | No |
| -1,320605514 | -1,181330247 | Yes | No | No |
| -1,324504586 | -1,164288759 | Yes | No | No |
| -1,301807535 | -1,218256815 | Yes | Yes | Yes |
| -1,20118608 | -1,134129065 | Yes | No | No |
| -1,513163767 | -1,093211642 | Yes | No | No |
| -1,608644498 | -1,102743193 | Yes | No | No |
| -1,254044996 | -1,118948419 | Yes | No | No |
| -1,277907993 | -1,047891912 | Yes | No | No |
| 1,30216272 | 1,165262204 | Yes | No | No |
| 1,276056935 | 1,112043732 | Yes | No | No |
| -1,378033075 | -1,237103549 | Yes | Yes | Yes |
| -1,716265635 | -1,143160029 | Yes | No | No |

| | | | | |
|---|---|---|---|---|
| 1,446897477 | 1,327449939 | Yes | Yes | Yes |
| 1,633462994 | 1,628992806 | Yes | Yes | Yes |
| -1,218236449 | -1,075660116 | Yes | No | No |
| -1,301361463 | -1,155639633 | Yes | No | No |
| -1,338071509 | -1,222653576 | Yes | Yes | Yes |
| -1,302728319 | -1,272255227 | Yes | Yes | Yes |
| 1,301723321 | 1,114763766 | Yes | No | No |
| -1,258950224 | -1,110682436 | Yes | No | No |
| -1,371576666 | 1,035391462 | Yes | No | No |
| -1,546850429 | -1,240709178 | Yes | No | No |
| -1,327609943 | -1,292221019 | Yes | Yes | Yes |
| -1,299131595 | -1,098023752 | Yes | No | No |
| 1,224310957 | 1,246813117 | Yes | Yes | Yes |
| 1,349244366 | 1,159761534 | Yes | No | No |
| 1,436443232 | 1,164124946 | Yes | No | No |
| -1,641705934 | 1,092380539 | Yes | No | No |
| 1,276055278 | 1,186639417 | Yes | No | No |
| -1,240026356 | -1,242754448 | Yes | Yes | Yes |
| 1,266198883 | 1,187474988 | Yes | No | No |
| -1,414341553 | -1,238451987 | Yes | Yes | Yes |
| 1,249139241 | -1,110248753 | Yes | No | No |
| -1,298014848 | -1,004112183 | Yes | No | No |
| -1,286252411 | -1,029224442 | Yes | No | No |
| -1,293641032 | -1,158296137 | Yes | No | No |
| -1,218117232 | -1,275340404 | Yes | Yes | Yes |
| -1,271889339 | -1,464817348 | Yes | Yes | Yes |
| -1,203176867 | -1,165005547 | Yes | No | No |
| -1,443547613 | -1,122098758 | Yes | No | No |
| -1,267563281 | -1,225840481 | Yes | Yes | Yes |
| -1,200993565 | -1,099034526 | Yes | No | No |
| -1,30931982 | -1,145101725 | Yes | No | No |
| 1,277597554 | 1,042584053 | Yes | No | No |
| 1,224323182 | 1,307153083 | Yes | Yes | Yes |
| 1,496894767 | -1,056909618 | Yes | No | No |
| -1,329786186 | -1,202701793 | Yes | Yes | Yes |
| -1,218515268 | -1,169959346 | Yes | No | No |
| 1,246921424 | 1,291930532 | Yes | Yes | Yes |
| -1,434512067 | -1,118209095 | Yes | No | No |
| -1,281048601 | -1,154374874 | Yes | No | No |
| -1,203812664 | -1,167803211 | Yes | No | No |
| -1,366016147 | -1,39079782 | Yes | Yes | Yes |
| -1,212813416 | -1,107673613 | Yes | No | No |
| 1,244752734 | 1,319961606 | Yes | Yes | Yes |
| 1,223643127 | 1,401110963 | Yes | Yes | Yes |
| -1,232556605 | -1,199469306 | Yes | No | No |
| 1,290352755 | 1,263177696 | Yes | Yes | Yes |
| 1,213368491 | -1,002566953 | Yes | No | No |
| 1,489381871 | -1,038403459 | Yes | No | No |
| -1,322383479 | -1,136836384 | Yes | No | No |
| 1,21915057 | 1,035146547 | Yes | No | No |
| 1,244809485 | 1,291854788 | Yes | Yes | Yes |
| -1,446557373 | -1,243376153 | Yes | Yes | Yes |
| 1,473399521 | 1,673639418 | Yes | Yes | Yes |

| | | | | |
|---|---|---|---|---|
| -1,573703931 | -1,227000405 | Yes | Yes | Yes |
| -1,260426723 | -1,185014649 | Yes | No | No |
| -1,262645007 | -1,163949917 | Yes | No | No |
| -1,293036995 | -1,117616647 | Yes | No | No |
| 1,379110396 | 1,204770646 | Yes | Yes | Yes |
| -1,554206885 | -1,118782776 | Yes | No | No |
| -1,595456748 | -1,10964575 | Yes | No | No |
| 1,323549153 | 1,353877891 | Yes | Yes | Yes |
| -1,275349043 | -1,186245581 | Yes | No | No |
| -1,21198354 | -1,128308397 | Yes | No | No |
| -1,409405067 | -1,271532769 | Yes | Yes | Yes |
| -1,326368186 | -1,142575053 | Yes | No | No |
| 1,231366199 | 1,208369547 | Yes | Yes | Yes |
| -1,416755572 | -1,123646026 | Yes | No | No |
| 1,283735448 | 1,028996295 | Yes | No | No |
| 1,229199455 | 1,279275398 | Yes | Yes | Yes |
| -1,21006158 | -1,317099805 | Yes | Yes | Yes |
| -1,789737886 | 1,029538536 | Yes | No | No |
| 1,205057796 | 1,246782042 | Yes | Yes | Yes |
| 1,206590152 | 1,436491575 | Yes | Yes | Yes |
| -1,239752976 | -1,492910058 | Yes | Yes | Yes |
| 1,575432486 | -1,027606577 | Yes | No | No |
| 1,207742568 | 1,253389065 | Yes | Yes | Yes |
| 1,323041461 | 1,302617676 | Yes | Yes | Yes |
| 1,286908451 | 1,130354598 | Yes | No | No |
| -1,327651394 | -1,174213804 | Yes | No | No |
| -1,337907754 | -1,101613561 | Yes | No | No |
| -1,284784113 | -1,261189187 | Yes | Yes | Yes |
| -1,35043014 | -1,136926078 | Yes | No | No |
| -1,36312296 | 1,055400055 | Yes | No | No |
| 1,678761747 | 1,108989692 | Yes | No | No |
| 1,2515102 | 1,380855593 | Yes | Yes | Yes |
| 1,419432688 | 1,268981007 | Yes | Yes | Yes |
| -1,206815788 | -1,221028922 | Yes | Yes | Yes |
| 1,470721707 | 1,531353964 | Yes | Yes | Yes |
| 1,350833811 | 1,368512601 | Yes | Yes | Yes |
| -1,203275508 | -1,095790081 | Yes | No | No |
| 1,226549296 | 1,291963604 | Yes | Yes | Yes |
| 1,293858007 | 1,366432369 | Yes | Yes | Yes |
| -1,597662963 | -1,187443118 | Yes | No | No |
| 1,253523215 | 1,340862962 | Yes | Yes | Yes |
| -1,500785754 | -1,242818117 | Yes | Yes | Yes |
| 1,304073979 | 1,248986426 | Yes | Yes | Yes |
| 1,394788641 | 1,099880751 | Yes | No | No |
| -1,345663758 | -1,312525558 | Yes | Yes | Yes |
| 1,25069493 | 1,023419569 | Yes | No | No |
| 1,887032565 | 1,363777701 | Yes | Yes | Yes |
| -2,209075096 | -1,35216148 | Yes | No | No |
| -1,375833819 | -1,17337042 | Yes | No | No |
| -1,620035676 | -1,093134064 | Yes | No | No |
| 1,30554549 | 1,093266482 | Yes | No | No |
| 1,29467222 | -1,006279739 | Yes | No | No |
| -1,30552667 | -1,301634489 | Yes | Yes | Yes |

| | | | | |
|---|---|---|---|---|
| 1,273123917 | 1,259595131 | Yes | Yes | Yes |
| -1,295635583 | 1,017042762 | Yes | No | No |
| 1,283251099 | 1,356934668 | Yes | Yes | Yes |
| -1,327059169 | -1,025233711 | Yes | No | No |
| -1,589188193 | -1,063314172 | Yes | No | No |
| -1,51374701 | 1,093460448 | Yes | No | No |
| -1,247786208 | -1,167112435 | Yes | No | No |
| 1,346823664 | 1,29491367 | Yes | Yes | Yes |
| -1,202556729 | -1,10261283 | Yes | No | No |
| -1,375754702 | -1,389984704 | Yes | Yes | Yes |
| -1,221066697 | -1,131796404 | Yes | No | No |
| -1,325821202 | -1,12432025 | Yes | No | No |
| 1,236702773 | 1,216039281 | Yes | Yes | Yes |
| -1,201733108 | 1,011341127 | Yes | No | No |
| -1,4967602 | -1,220042872 | Yes | Yes | Yes |
| -1,280254704 | -1,024048964 | Yes | No | No |
| -1,351664257 | -1,113565692 | Yes | No | No |
| -1,31909547 | -1,760204124 | Yes | Yes | Yes |
| -1,310949808 | -1,632134299 | Yes | Yes | Yes |
| 1,224027872 | 1,119977568 | Yes | No | No |
| 1,368196808 | 1,208897286 | Yes | Yes | Yes |
| -1,208974094 | -1,195145081 | Yes | No | No |
| -1,368768072 | -1,012234521 | Yes | No | No |
| -1,282581892 | -1,083936178 | Yes | No | No |
| -1,319919612 | -1,06986634 | Yes | No | No |
| -1,345682715 | -1,003029013 | Yes | No | No |
| 1,36215214 | 1,434947884 | Yes | Yes | Yes |
| 1,347487961 | 1,443572423 | Yes | Yes | Yes |
| 1,248533558 | 1,131896094 | Yes | No | No |
| -1,227070769 | -1,139069719 | Yes | No | No |
| 1,288627046 | 1,240502557 | Yes | Yes | Yes |
| -1,522779707 | -1,007913219 | Yes | No | No |
| -1,297166077 | -1,41453202 | Yes | Yes | Yes |
| -1,485531908 | -1,267371498 | Yes | Yes | Yes |
| -1,434797369 | -1,273007101 | Yes | Yes | Yes |
| -1,215122815 | -1,107366593 | Yes | No | No |
| -1,259644399 | -1,069164255 | Yes | No | No |
| 1,29838247 | 1,200564921 | Yes | Yes | Yes |
| 1,380532846 | 1,304903887 | Yes | Yes | Yes |
| -1,315616598 | -1,485534069 | Yes | Yes | Yes |
| 1,270801946 | 1,394287596 | Yes | Yes | Yes |
| 1,380534525 | 1,395774579 | Yes | Yes | Yes |
| 1,22656705 | 1,160786595 | Yes | No | No |
| -1,285996731 | -1,047683414 | Yes | No | No |
| -1,254215355 | -1,160700304 | Yes | No | No |
| -1,464880103 | -1,035569283 | Yes | No | No |
| 1,36693248 | 1,06383143 | Yes | No | No |
| 1,65421648 | -1,056929531 | Yes | No | No |
| 1,275459081 | 1,046264397 | Yes | No | No |
| 1,289109632 | 1,457292638 | Yes | Yes | Yes |
| 1,205236335 | 1,386866197 | Yes | Yes | Yes |
| -1,258390344 | -1,14221046 | Yes | No | No |
| -1,231294654 | -1,154776179 | Yes | No | No |

| | | | | |
|---|---|---|---|---|
| 1,283411568 | 1,341110039 | Yes | Yes | Yes |
| 1,320495866 | 1,33340827 | Yes | Yes | Yes |
| 1,444134025 | 1,107895001 | Yes | No | No |
| 1,240236444 | 1,039648284 | Yes | No | No |
| 1,299409678 | 1,176517688 | Yes | No | No |
| 1,25141081 | 1,205564626 | Yes | Yes | Yes |
| 1,520942351 | 1,496864094 | Yes | Yes | Yes |
| -1,307228158 | -1,45450097 | Yes | Yes | Yes |
| 2,128479702 | 1,168749181 | Yes | No | No |
| 1,330650715 | 1,278833001 | Yes | Yes | Yes |
| -1,263714372 | -1,07266061 | Yes | No | No |
| -1,386876493 | -1,150359224 | Yes | No | No |
| -1,303829786 | -1,117024201 | Yes | No | No |
| -1,318618292 | -1,14071282 | Yes | No | No |
| 1,398595069 | 1,436737416 | Yes | Yes | Yes |
| -1,273383391 | -1,125219604 | Yes | No | No |
| -1,229937261 | -1,082874771 | Yes | No | No |
| 1,347413636 | 1,270438191 | Yes | Yes | Yes |
| 1,360351571 | 1,360831173 | Yes | Yes | Yes |
| 1,432135259 | 1,437745741 | Yes | Yes | Yes |
| 1,201780197 | 1,175395573 | Yes | No | No |
| -1,507768485 | -1,287403085 | Yes | Yes | Yes |
| -1,320207553 | -1,358963371 | Yes | Yes | Yes |
| -1,223797986 | -1,120185482 | Yes | No | No |
| 1,246434716 | 1,226598262 | Yes | Yes | Yes |
| 1,249946926 | -1,042313033 | Yes | No | No |
| 1,358559373 | 1,125242324 | Yes | No | No |
| 1,453569776 | 1,075019222 | Yes | No | No |
| -1,300540321 | -1,22110887 | Yes | Yes | Yes |
| -1,26267575 | -1,197822746 | Yes | No | No |
| -1,251657769 | -1,372750622 | Yes | Yes | Yes |
| -1,3165306 | -1,223746959 | Yes | Yes | Yes |
| 1,216249787 | 1,2103134 | Yes | Yes | Yes |
| -1,260390664 | -1,249592003 | Yes | Yes | Yes |
| -1,238202098 | -1,290885731 | Yes | Yes | Yes |
| 1,274361393 | 1,166311222 | Yes | No | No |
| 1,416009287 | 1,32664464 | Yes | Yes | Yes |
| -1,355833377 | -1,078276319 | Yes | No | No |
| -1,34171506 | -1,156683478 | Yes | No | No |
| 1,392772133 | 1,450317145 | Yes | Yes | Yes |
| -1,225619485 | -1,149005795 | Yes | No | No |
| -1,436809704 | -1,113439604 | Yes | No | No |
| -1,210035408 | -1,199586928 | Yes | No | No |
| 1,202539427 | -1,022093648 | Yes | No | No |
| 1,433640742 | 1,936970475 | Yes | Yes | Yes |
| 1,450475901 | 1,923417137 | Yes | Yes | Yes |
| -1,225167168 | 1,003491593 | Yes | No | No |
| -1,366239121 | 1,110157928 | Yes | No | No |
| -1,254598632 | -1,193259113 | Yes | No | No |
| 1,208559011 | 1,050729039 | Yes | No | No |
| -1,326100005 | -1,313550388 | Yes | Yes | Yes |
| -1,303445131 | -1,206965683 | Yes | Yes | Yes |
| 1,460741596 | 1,3066446 | Yes | Yes | Yes |

| | | | | |
|---|---|---|---|---|
| -1,252804813 | -1,114623861 | Yes | No | No |
| -1,521791444 | -1,180283529 | Yes | No | No |
| 1,266046664 | 1,43468212 | Yes | Yes | Yes |
| 1,215689611 | 1,037727777 | Yes | No | No |
| 2,096084635 | 1,261629515 | Yes | No | No |
| -1,379760754 | -1,211055753 | Yes | No | No |
| -1,33603683 | -1,088535832 | Yes | No | No |
| -1,246137583 | 1,064745151 | Yes | No | No |
| -1,651276927 | -1,336996875 | Yes | Yes | Yes |
| -1,243379347 | -1,181493083 | Yes | No | No |
| 1,285978152 | 1,280481576 | Yes | Yes | Yes |
| 1,225930657 | 1,399121844 | Yes | Yes | Yes |
| 1,232523581 | 1,195139789 | Yes | No | No |
| 1,322967098 | 1,242218431 | Yes | Yes | Yes |
| 1,29534789 | 1,275034021 | Yes | Yes | Yes |
| -1,441315987 | -1,173747738 | Yes | No | No |
| -1,276770571 | -1,148494759 | Yes | No | No |
| 1,291016446 | -1,021175417 | Yes | No | No |
| 1,476681203 | 1,608457543 | Yes | Yes | Yes |
| 1,401695785 | 1,565519459 | Yes | Yes | Yes |
| -1,351758433 | -1,325578324 | Yes | Yes | Yes |
| -1,547686222 | -1,179013496 | Yes | No | No |
| 1,29300738 | 1,120354538 | Yes | No | No |
| -1,201616122 | -1,05557159 | Yes | No | No |
| -1,646605404 | 1,039280995 | Yes | No | No |
| 1,257911255 | 1,113632141 | Yes | No | No |
| -1,246928395 | -1,224608542 | Yes | Yes | Yes |
| 1,268518868 | -1,115888441 | Yes | No | No |
| 1,487744549 | 1,139126822 | Yes | No | No |
| 1,472563807 | 1,154039287 | Yes | No | No |
| 1,212049426 | 1,189794611 | Yes | No | No |
| 1,284223568 | 1,236709952 | Yes | Yes | Yes |
| 1,266881615 | 1,120662367 | Yes | No | No |
| 1,207722381 | 1,213524002 | Yes | Yes | Yes |
| 1,314214472 | 1,217339147 | Yes | Yes | Yes |
| 1,253317821 | 1,090785631 | Yes | No | No |
| 1,374938131 | 1,146165954 | Yes | No | No |
| 1,34645644 | 1,093717956 | Yes | No | No |
| 1,318832954 | 1,466119689 | Yes | Yes | Yes |
| 1,34485032 | 1,188390954 | Yes | No | No |
| 1,55028398 | 1,099781764 | Yes | No | No |
| 1,332708939 | 1,233931731 | Yes | Yes | Yes |
| 1,304235095 | 1,087928503 | Yes | No | No |
| 1,234542195 | 1,04272024 | Yes | No | No |
| 1,266546358 | 1,155163398 | Yes | No | No |
| -1,338995603 | -1,178688751 | Yes | No | No |
| 1,493614192 | 1,60337308 | Yes | Yes | Yes |
| 1,379177583 | 1,424091218 | Yes | Yes | Yes |
| 1,310442373 | 1,344633039 | Yes | Yes | Yes |
| 1,254670692 | 1,42313758 | Yes | Yes | Yes |
| 1,327734845 | 1,298433384 | Yes | Yes | Yes |
| -1,353961408 | -1,187438186 | Yes | No | No |
| 1,282978698 | 1,073004145 | Yes | No | No |

| | | | | |
|---|---|---|---|---|
| -1,33602229 | -1,176622542 | Yes | No | No |
| 1,244300482 | 1,215443869 | Yes | Yes | Yes |
| -1,223205306 | -1,135451435 | Yes | No | No |
| 1,4284105 | 1,242909516 | Yes | Yes | Yes |
| 1,451620574 | 1,164942699 | Yes | No | No |
| -1,204745566 | -1,082246511 | Yes | No | No |
| -1,253654066 | -1,187457485 | Yes | No | No |
| 1,348807646 | 1,162523135 | Yes | No | No |
| -1,557307011 | -1,138789535 | Yes | No | No |
| 1,402144936 | 1,064138165 | Yes | No | No |
| 1,200162234 | 1,041759841 | Yes | No | No |
| 1,365985278 | 1,33013046 | Yes | Yes | Yes |
| 1,278839321 | 1,027007742 | Yes | No | No |
| -1,233872222 | -1,209492642 | Yes | Yes | Yes |
| 1,378645538 | 1,20829208 | Yes | Yes | Yes |
| 1,42902625 | 1,179122937 | Yes | No | No |
| -1,226603624 | -1,13612717 | Yes | No | No |
| -1,275115183 | -1,115936198 | Yes | No | No |
| 1,215418449 | 1,669747421 | Yes | Yes | Yes |
| 1,220484998 | 1,365055633 | Yes | Yes | Yes |
| -1,246573656 | -1,16745195 | Yes | No | No |
| 1,334341413 | 1,440568542 | Yes | Yes | Yes |
| 1,31079291 | 1,238371998 | Yes | Yes | Yes |
| 1,232946246 | 1,122983286 | Yes | No | No |
| 1,248343367 | 1,370551621 | Yes | Yes | Yes |
| 1,522885752 | 1,216982283 | Yes | Yes | Yes |
| -1,23079089 | -1,087788403 | Yes | No | No |
| -1,306828183 | -1,149621428 | Yes | No | No |
| -1,562170537 | -1,260018587 | Yes | Yes | Yes |
| -1,748992866 | -1,764146001 | Yes | Yes | Yes |
| -1,31068263 | -1,130210813 | Yes | No | No |
| -1,437349924 | -1,141946144 | Yes | No | No |
| 1,405250539 | 1,271306229 | Yes | Yes | Yes |
| -1,274079383 | -1,112892289 | Yes | No | No |
| 1,204275017 | 1,324817927 | Yes | Yes | Yes |
| 1,241172702 | 1,728437297 | Yes | Yes | Yes |
| -1,506345784 | 1,013034266 | Yes | No | No |
| 1,232195893 | 1,122083513 | Yes | No | No |
| -1,235843562 | -1,158267576 | Yes | No | No |
| 1,220550059 | -1,124677079 | Yes | No | No |
| 1,200364197 | 1,344802736 | Yes | Yes | Yes |
| 1,217540031 | 1,416956681 | Yes | Yes | Yes |
| -1,228843733 | -1,403361825 | Yes | Yes | Yes |
| 1,261379413 | 1,383016709 | Yes | Yes | Yes |
| -1,248878597 | -1,185908291 | Yes | No | No |
| 1,366487379 | 1,454955326 | Yes | Yes | Yes |
| 1,211631187 | 1,155421921 | Yes | No | No |
| 1,34517445 | 1,45227187 | Yes | Yes | Yes |
| 1,212495817 | 1,120180775 | Yes | No | No |
| -1,211714265 | -1,25302101 | Yes | Yes | Yes |
| 1,327471117 | 1,018229727 | Yes | No | No |
| 1,259864057 | 1,148123251 | Yes | No | No |
| -1,211781782 | -1,161273545 | Yes | No | No |

| | | | | |
|---|---|---|---|---|
| 1,300777329 | 1,165365981 | Yes | No | No |
| -1,463323998 | -1,211599673 | Yes | Yes | Yes |
| 1,25123514 | 1,184916918 | Yes | No | No |
| 1,455143956 | 1,038858846 | Yes | No | No |
| -1,353191181 | -1,200007195 | Yes | Yes | Yes |
| -1,204879427 | -1,139900103 | Yes | No | No |
| -1,409734424 | -1,161896896 | Yes | No | No |
| 1,39609584 | 1,310940269 | Yes | Yes | Yes |
| 1,201318569 | 1,444386409 | Yes | Yes | Yes |
| 1,267758425 | 1,2177306 | Yes | Yes | Yes |
| -1,318212655 | -1,053753783 | Yes | No | No |
| -1,313181543 | -1,09265736 | Yes | No | No |
| -1,2907877 | -1,353372823 | Yes | Yes | Yes |
| 1,252294297 | 1,332185818 | Yes | Yes | Yes |
| -1,200772465 | -1,106736959 | Yes | No | No |
| 1,242069678 | 1,388722312 | Yes | Yes | Yes |
| -1,256929777 | -1,205750623 | Yes | Yes | Yes |
| -1,480540367 | -1,492474465 | Yes | Yes | Yes |
| -1,28262444 | -1,182924679 | Yes | No | No |
| 1,637111756 | 1,509938538 | Yes | Yes | Yes |
| 1,234407452 | 1,298843856 | Yes | Yes | Yes |
| 1,224104675 | 1,509020605 | Yes | Yes | Yes |
| -1,267921878 | -1,097129408 | Yes | No | No |
| 1,282596483 | 1,20323487 | Yes | Yes | Yes |
| 1,399531184 | 1,24495152 | Yes | Yes | Yes |
| -1,409550271 | -1,035062987 | Yes | No | No |
| -1,437471879 | -1,123681128 | Yes | No | No |
| -1,20084451 | -1,192248995 | Yes | No | No |
| 1,419651124 | 1,71297452 | Yes | Yes | Yes |
| -1,235134976 | -1,189896138 | Yes | No | No |
| -1,202581761 | -1,066548405 | Yes | No | No |
| -1,279891897 | -1,191469204 | Yes | No | No |
| -1,325737492 | -1,096450661 | Yes | No | No |
| -1,253743028 | -1,203218483 | Yes | Yes | Yes |
| -1,290299925 | -1,200860037 | Yes | Yes | Yes |
| 1,235641511 | 1,185461298 | Yes | No | No |
| 1,257208223 | 1,078533372 | Yes | No | No |
| 1,435268729 | 1,010162822 | Yes | No | No |
| -1,338776526 | -1,131519729 | Yes | No | No |
| -1,401685075 | -1,174050039 | Yes | No | No |
| 1,220353374 | 1,317419368 | Yes | Yes | Yes |
| -1,420719958 | 1,088249128 | Yes | No | No |
| 1,235840727 | 1,206670349 | Yes | Yes | Yes |
| 1,237514857 | 1,299847409 | Yes | Yes | Yes |
| 1,396160292 | 1,054368728 | Yes | No | No |
| 1,454278378 | 1,038369294 | Yes | No | No |
| 1,217568978 | 1,383927546 | Yes | Yes | Yes |
| 1,213324078 | 1,178502736 | Yes | No | No |
| -1,286632483 | -1,083632734 | Yes | No | No |
| 1,377755978 | 1,015828038 | Yes | No | No |
| 1,244234479 | 1,27589598 | Yes | Yes | Yes |
| 1,223686463 | 1,442558646 | Yes | Yes | Yes |
| -1,253802564 | -1,314768699 | Yes | Yes | Yes |

| | | | | |
|---|---|---|---|---|
| -1,209367276 | -1,137202242 | Yes | No | No |
| -1,327951017 | -1,204815384 | Yes | Yes | Yes |
| -1,37872529 | -1,127489577 | Yes | No | No |
| -1,585139535 | -1,096813641 | Yes | No | No |
| -1,651887787 | -1,149789428 | Yes | No | No |
| -1,221056878 | -1,198008633 | Yes | No | No |
| -1,352984632 | -1,20787489 | Yes | Yes | Yes |
| -1,234388899 | -1,103146391 | Yes | No | No |
| -1,293068479 | -1,192080628 | Yes | No | No |
| -1,213321385 | -1,275661125 | Yes | Yes | Yes |
| -1,22329971 | -1,130277419 | Yes | No | No |
| -1,209976785 | -1,079870924 | Yes | No | No |
| -1,204031052 | -1,17360475 | Yes | No | No |
| -1,31405185 | -1,237047117 | Yes | Yes | Yes |
| -1,278556116 | -1,069685213 | Yes | No | No |
| -1,279092508 | -1,091074208 | Yes | No | No |
| -1,254878076 | -1,157703613 | Yes | No | No |
| -1,277462533 | -1,221544106 | Yes | Yes | Yes |
| 1,309595078 | 1,07821411 | Yes | No | No |
| -1,285560486 | -1,221303106 | Yes | Yes | Yes |
| -1,941644458 | -1,129484197 | Yes | No | No |
| -1,282223033 | -1,167065477 | Yes | No | No |
| -1,405569467 | -1,008157875 | Yes | No | No |
| 1,208483522 | 1,327289774 | Yes | Yes | Yes |
| 1,626981932 | 1,569760368 | Yes | Yes | Yes |
| -1,208286496 | -1,187947626 | Yes | No | No |
| 1,29004825 | 1,226537639 | Yes | Yes | Yes |
| -1,20419062 | -1,306500158 | Yes | Yes | Yes |
| -1,40210214 | -1,241833213 | Yes | Yes | Yes |
| -1,426015207 | -1,177112187 | Yes | No | No |
| -1,898083782 | -1,451451036 | Yes | Yes | Yes |
| 1,289629304 | 1,230326405 | Yes | Yes | Yes |
| -1,287449854 | -1,106635112 | Yes | No | No |
| 1,453739991 | 1,535755136 | Yes | Yes | Yes |
| 1,212754966 | 1,524745002 | Yes | Yes | Yes |
| -1,209268555 | -1,106534798 | Yes | No | No |
| 1,334532879 | 1,026476148 | Yes | No | No |
| -1,212262694 | -1,119579719 | Yes | No | No |
| 1,271906588 | 1,120879606 | Yes | No | No |
| 1,252294297 | 1,332185818 | Yes | Yes | Yes |
| -1,278899194 | -1,179812185 | Yes | No | No |
| 1,459498156 | 1,190901996 | Yes | No | No |
| -1,2460816 | -1,045728509 | Yes | No | No |
| 1,24870739 | -1,010998975 | Yes | No | No |
| -1,265241567 | -1,261085699 | Yes | Yes | Yes |
| 1,239968892 | 1,27244646 | Yes | Yes | Yes |
| -1,21077323 | -1,13922803 | Yes | No | No |
| -1,394006433 | -1,230879784 | Yes | Yes | Yes |
| -1,428818462 | -1,189126657 | Yes | No | No |
| -1,349986022 | -1,108147845 | Yes | No | No |
| -1,301658751 | -1,021235245 | Yes | No | No |
| 1,242951117 | 1,256423092 | Yes | Yes | Yes |
| 1,212522661 | 1,261558195 | Yes | Yes | Yes |

| | | | | |
|---|---|---|---|---|
| 1,207451843 | -1,104807993 | Yes | No | No |
| 1,306612575 | 1,548968966 | Yes | Yes | Yes |
| 1,254275825 | 1,624137372 | Yes | Yes | Yes |
| -1,225999587 | -1,211092179 | Yes | Yes | Yes |
| -1,271214545 | -1,330144322 | Yes | Yes | Yes |
| -1,265558328 | -1,327461914 | Yes | Yes | Yes |
| -1,240074734 | -1,291572621 | Yes | Yes | Yes |
| -1,364872737 | -1,10048654 | Yes | No | No |
| -1,232110027 | -1,175711468 | Yes | No | No |
| 1,237263677 | 1,16916071 | Yes | No | No |
| -1,458707909 | 1,020640067 | Yes | No | No |
| -1,267032704 | -1,169672308 | Yes | No | No |
| 1,241429128 | 1,282189036 | Yes | Yes | Yes |
| -1,226842938 | -1,154091688 | Yes | No | No |
| -1,275399539 | -1,208143059 | Yes | Yes | Yes |
| -1,253287661 | -1,149560609 | Yes | No | No |
| -1,260802466 | -1,129237921 | Yes | No | No |
| -1,239805335 | 1,03944488 | Yes | No | No |
| 1,244890288 | 1,208384596 | Yes | Yes | Yes |
| 1,35465114 | 1,659054929 | Yes | Yes | Yes |
| 1,408999951 | 1,189465785 | Yes | No | No |
| -1,27885051 | -1,156001719 | Yes | No | No |
| -1,359379565 | 1,032054589 | Yes | No | No |
| -1,418159102 | 1,037476327 | Yes | No | No |
| -1,309480236 | -1,044043221 | Yes | No | No |
| -1,2386308 | -1,135445721 | Yes | No | No |
| 1,326856159 | 1,083343153 | Yes | No | No |
| -1,695798581 | -1,224221607 | Yes | No | No |
| -1,300592017 | -1,070418632 | Yes | No | No |
| 1,209963789 | 1,510327873 | Yes | Yes | Yes |
| -1,366404837 | -1,011402391 | Yes | No | No |
| 1,217641859 | 1,177359648 | Yes | No | No |
| -1,276767528 | -1,060265091 | Yes | No | No |
| 1,285600457 | 1,087305581 | Yes | No | No |
| 1,266118629 | 1,281117062 | Yes | Yes | Yes |
| -1,300352809 | -1,231752037 | Yes | Yes | Yes |
| 1,28373415 | 1,430692274 | Yes | Yes | Yes |
| -1,314271803 | -1,080301442 | Yes | No | No |
| -1,211602626 | -1,100871789 | Yes | No | No |
| -1,31916054 | -1,198667249 | Yes | No | No |
| -1,269008508 | -1,193836012 | Yes | No | No |
| -1,284772909 | -1,220274209 | Yes | Yes | Yes |
| -1,216421263 | -1,110239458 | Yes | No | No |
| -1,343086808 | -1,050578657 | Yes | No | No |
| -1,347799898 | -1,129763939 | Yes | No | No |
| -1,271041605 | -1,14991054 | Yes | No | No |
| -1,417393028 | -1,168055127 | Yes | No | No |
| 1,20313752 | 1,07952961 | Yes | No | No |
| -1,439098117 | -1,179757137 | Yes | No | No |
| 1,276235822 | 1,097192834 | Yes | No | No |
| -1,236987172 | -1,217639446 | Yes | Yes | Yes |
| 1,207092659 | 1,500361964 | Yes | Yes | Yes |
| -1,37461498 | -1,222463247 | Yes | Yes | Yes |

| | | | | |
|---|---|---|---|---|
| -1,227473569 | -1,133540041 | Yes | No | No |
| 1,427197025 | -1,076552553 | Yes | No | No |
| 1,208557897 | 1,171009401 | Yes | No | No |
| 1,220240825 | -1,014028334 | Yes | No | No |
| -1,272704712 | -1,457611934 | Yes | Yes | Yes |
| -1,39703404 | -1,178035185 | Yes | No | No |
| -1,224725897 | -1,198712334 | Yes | No | No |
| -1,305185935 | -1,165226376 | Yes | No | No |
| -1,31203839 | -1,224788014 | Yes | Yes | Yes |
| -1,333305515 | -1,117683191 | Yes | No | No |
| -1,287313592 | 1,017972257 | Yes | No | No |
| -1,350165882 | 1,006231448 | Yes | No | No |
| -1,257019474 | -1,182909704 | Yes | No | No |
| -1,202508118 | -1,150141059 | Yes | No | No |
| -1,249350876 | -1,242793834 | Yes | Yes | Yes |
| -1,408134516 | -1,179471585 | Yes | No | No |
| -1,263527767 | -1,248603547 | Yes | Yes | Yes |
| -1,288275051 | -1,035243063 | Yes | No | No |
| 1,475232304 | 1,760049873 | Yes | Yes | Yes |
| -1,239108115 | -1,259512638 | Yes | Yes | Yes |
| -1,42415063 | -1,118554745 | Yes | No | No |
| 1,270736155 | 1,1665587 | Yes | No | No |
| 1,267882087 | 1,109093298 | Yes | No | No |
| 1,219900899 | 1,249455726 | Yes | Yes | Yes |
| -1,364945406 | -1,190337075 | Yes | No | No |
| 1,295150911 | 1,036205963 | Yes | No | No |
| -1,335437558 | -1,08148024 | Yes | No | No |
| 1,220199491 | 1,224219316 | Yes | Yes | Yes |
| 1,230360873 | 1,556277352 | Yes | Yes | Yes |
| -1,330462831 | -1,052716176 | Yes | No | No |
| -1,28007938 | -1,061396801 | Yes | No | No |
| -1,33511854 | -1,392545858 | Yes | Yes | Yes |
| -1,21358454 | -1,216092191 | Yes | Yes | Yes |
| 1,326064875 | 1,277779161 | Yes | Yes | Yes |
| -1,331275329 | -1,196451367 | Yes | No | No |
| 1,410096789 | -1,049012105 | Yes | No | No |
| 1,408743709 | 1,246422086 | Yes | Yes | Yes |
| 1,567265505 | 1,199587521 | Yes | No | No |
| 1,520605142 | 1,108810866 | Yes | No | No |
| 1,260125692 | 1,144637537 | Yes | No | No |
| -1,206699893 | -1,121046063 | Yes | No | No |
| -1,217738695 | -1,153433933 | Yes | No | No |
| -1,355865445 | -1,133226851 | Yes | No | No |
| 1,234258956 | -1,011233297 | Yes | No | No |
| 1,234986765 | 1,295411876 | Yes | Yes | Yes |
| -1,306813818 | -1,09766969 | Yes | No | No |
| -1,324490214 | -1,093997501 | Yes | No | No |
| 1,317700571 | 1,134832327 | Yes | No | No |
| 1,305562643 | 1,230155284 | Yes | Yes | Yes |
| -1,378138441 | -1,147039711 | Yes | No | No |
| -1,247788418 | -1,144143217 | Yes | No | No |
| -1,226938023 | -1,141516213 | Yes | No | No |
| -1,27859116 | 1,006736834 | Yes | No | No |

| | | | | |
|---|---|---|---|---|
| 1,309397669 | 1,384022046 | Yes | Yes | Yes |
| 1,37589718 | 1,410650036 | Yes | Yes | Yes |
| -1,30071901 | -1,108349138 | Yes | No | No |
| 1,205340815 | 1,221114851 | Yes | Yes | Yes |
| 1,272882535 | 1,154493959 | Yes | No | No |
| -1,22447125 | -1,113704809 | Yes | No | No |
| -1,444150781 | -1,406434448 | Yes | Yes | Yes |
| -1,54234422 | -1,28431284 | Yes | Yes | Yes |
| -1,254513478 | -1,180414211 | Yes | No | No |
| 1,397820261 | 1,300385093 | Yes | Yes | Yes |
| 1,262282049 | -1,078081453 | Yes | No | No |
| 1,353267574 | 1,25975698 | Yes | Yes | Yes |
| 1,250239629 | 1,403096526 | Yes | Yes | Yes |
| 1,222758705 | 1,342657627 | Yes | Yes | Yes |
| 1,322360649 | 1,210622948 | Yes | Yes | Yes |
| -1,315367941 | -1,173398667 | Yes | No | No |
| -1,215495054 | -1,326391915 | Yes | Yes | Yes |
| -1,41322764 | -1,081089109 | Yes | No | No |
| -1,289500923 | -1,298285533 | Yes | Yes | Yes |
| -1,217184319 | -1,163097582 | Yes | No | No |
| -1,260058545 | -1,104046884 | Yes | No | No |
| -1,326178236 | -1,120881974 | Yes | No | No |
| -1,201487154 | -1,126565293 | Yes | No | No |
| 1,26896938 | 1,10667054 | Yes | No | No |
| 1,286264517 | 1,210674431 | Yes | Yes | Yes |
| 1,501685244 | 1,046827101 | Yes | No | No |
| -1,373788814 | -1,05510146 | Yes | No | No |
| -1,255624792 | -1,037211143 | Yes | No | No |
| 1,268012485 | 1,167840959 | Yes | No | No |
| 1,281618662 | -1,019717553 | Yes | No | No |
| 1,239940639 | 1,039332415 | Yes | No | No |
| 1,252866294 | 1,32513649 | Yes | Yes | Yes |
| 1,504080611 | 1,216539018 | Yes | Yes | Yes |
| 1,281982421 | 1,348106017 | Yes | Yes | Yes |
| -1,216983827 | -1,106431373 | Yes | No | No |
| 1,208763522 | 1,193808868 | Yes | No | No |
| -1,371218231 | -1,053375494 | Yes | No | No |
| -1,388831983 | -1,153095706 | Yes | No | No |
| -1,286974244 | -1,246185768 | Yes | Yes | Yes |
| 1,327393721 | 1,214809839 | Yes | Yes | Yes |
| -1,247384104 | -1,474554256 | Yes | Yes | Yes |
| -1,259355068 | -1,132703822 | Yes | No | No |
| 1,269027294 | 1,268071119 | Yes | Yes | Yes |
| 1,338466906 | 1,466371869 | Yes | Yes | Yes |
| -1,363843471 | -1,319118958 | Yes | Yes | Yes |
| 1,206998534 | 1,207069208 | Yes | Yes | Yes |
| 1,32987947 | 1,4782701 | Yes | Yes | Yes |
| -1,249186717 | -1,131680609 | Yes | No | No |
| -1,42793431 | -1,079681072 | Yes | No | No |
| -1,262017027 | -1,195037112 | Yes | No | No |
| -1,34211243 | -1,107730292 | Yes | No | No |
| -1,228142032 | -1,193240177 | Yes | No | No |
| -1,469657465 | -1,246430195 | Yes | Yes | Yes |

| | | | | |
|---|---|---|---|---|
| -1,405676141 | -1,127447504 | Yes | No | No |
| -1,279682309 | -1,081265013 | Yes | No | No |
| 1,225690299 | 1,334870707 | Yes | Yes | Yes |
| 1,636843335 | 1,138419473 | Yes | No | No |
| 1,272543762 | 1,246283071 | Yes | Yes | Yes |
| 1,275191607 | 1,229640052 | Yes | Yes | Yes |
| 1,254236099 | 1,360770618 | Yes | Yes | Yes |
| 1,315326635 | 1,226418667 | Yes | Yes | Yes |
| 1,381681209 | 1,221741131 | Yes | Yes | Yes |
| 1,247281312 | 1,36054372 | Yes | Yes | Yes |
| -1,275466601 | 1,039886414 | Yes | No | No |
| -1,3216209 | -1,11752392 | Yes | No | No |
| 1,345182852 | 1,338779708 | Yes | Yes | Yes |
| 1,275462029 | 1,1732512 | Yes | No | No |
| -1,293943041 | -1,142906285 | Yes | No | No |
| -1,215690769 | -1,293697682 | Yes | Yes | Yes |
| 1,230111333 | 1,159281073 | Yes | No | No |
| -1,315011752 | -1,251722432 | Yes | Yes | Yes |
| 1,247752739 | 1,077370064 | Yes | No | No |
| -1,239740782 | -1,146440394 | Yes | No | No |
| 1,394671513 | 1,076051856 | Yes | No | No |
| 1,209027174 | 1,250522401 | Yes | Yes | Yes |
| -1,247791105 | -1,275872096 | Yes | Yes | Yes |
| -1,203998909 | -1,182913022 | Yes | No | No |
| 1,203408223 | -1,061129479 | Yes | No | No |
| 1,216783478 | 1,275186009 | Yes | Yes | Yes |
| 1,250150301 | 1,045192664 | Yes | No | No |
| 1,221965033 | 1,125493088 | Yes | No | No |
| -1,206543905 | -1,11483986 | Yes | No | No |
| -1,214690241 | -1,154593105 | Yes | No | No |
| 1,474605051 | -1,013425534 | Yes | No | No |
| -1,548146981 | -1,300150692 | Yes | No | No |
| -1,250155394 | -1,047687554 | Yes | No | No |
| 1,377517684 | 1,232867375 | Yes | Yes | Yes |
| 1,249932138 | 1,289369808 | Yes | Yes | Yes |
| -1,28668889 | -1,278558679 | Yes | Yes | Yes |
| -1,265192999 | -1,204071535 | Yes | Yes | Yes |
| 1,280976112 | 1,21392685 | Yes | Yes | Yes |
| 1,396090491 | 1,172892551 | Yes | No | No |
| -1,353727143 | -1,067665226 | Yes | No | No |
| 1,254938455 | 1,198259557 | Yes | No | No |
| 1,354441459 | 1,163123437 | Yes | No | No |
| -1,261008902 | -1,083887503 | Yes | No | No |
| -1,206094546 | -1,162159352 | Yes | No | No |
| 1,228088759 | 1,457504229 | Yes | Yes | Yes |
| 1,263518908 | 1,346717338 | Yes | Yes | Yes |
| -1,210626966 | -1,282703445 | Yes | Yes | Yes |
| -1,470476592 | -1,303964979 | Yes | Yes | Yes |
| -1,321103956 | -1,188775483 | Yes | No | No |
| -1,300099436 | -1,125621549 | Yes | No | No |
| -1,24065235 | -1,1251249 | Yes | No | No |
| -1,247186867 | -1,194010791 | Yes | No | No |
| -1,338423159 | -1,072929768 | Yes | No | No |

| | | | | |
|---|---|---|---|---|
| -1,304633378 | -1,254365526 | Yes | Yes | Yes |
| -1,257002558 | -1,226805975 | Yes | Yes | Yes |
| -1,248877249 | -1,159957137 | Yes | No | No |
| 1,236254991 | 1,464474466 | Yes | Yes | Yes |
| 1,252221419 | 1,658676501 | Yes | Yes | Yes |
| -1,24663814 | -1,124805552 | Yes | No | No |
| -1,310398822 | -1,138176335 | Yes | No | No |
| -1,268503167 | -1,127333612 | Yes | No | No |
| -1,248634335 | -1,267421232 | Yes | Yes | Yes |
| -1,431478883 | 1,024489043 | Yes | No | No |
| -1,234603648 | -1,149103081 | Yes | No | No |
| -1,247867641 | -1,071399745 | Yes | No | No |
| -1,532065948 | -1,278765907 | Yes | Yes | Yes |
| -1,464073632 | -1,227251201 | Yes | Yes | Yes |
| -1,23220984 | -1,195377505 | Yes | No | No |
| -1,405741243 | -1,321020941 | Yes | Yes | Yes |
| -1,237435289 | -1,152502671 | Yes | No | No |
| -1,370653938 | -1,041749213 | Yes | No | No |
| -1,264924805 | -1,317913355 | Yes | Yes | Yes |
| -1,796372864 | -1,309577947 | Yes | Yes | Yes |
| -1,226854255 | -1,174140928 | Yes | No | No |
| 1,231709898 | 1,272660223 | Yes | Yes | Yes |
| -1,22433123 | -1,121329566 | Yes | No | No |
| 1,267306978 | 1,014380044 | Yes | No | No |
| -1,242806576 | -1,110844419 | Yes | No | No |
| -1,444895425 | -1,21192091 | Yes | Yes | Yes |
| -1,20436854 | -1,065967191 | Yes | No | No |
| -1,285561644 | -1,18620502 | Yes | No | No |
| 1,302160947 | 1,110216809 | Yes | No | No |
| -1,439421939 | -1,312763844 | Yes | Yes | Yes |
| 1,244160424 | 1,192935922 | Yes | No | No |
| 1,249990957 | 1,264859453 | Yes | Yes | Yes |
| -1,328969546 | -1,00320092 | Yes | No | No |
| 1,454238119 | 1,214793121 | Yes | Yes | Yes |
| -1,266770553 | -1,150882708 | Yes | No | No |
| 1,380115364 | 1,010584928 | Yes | No | No |
| -1,20249823 | -1,089387951 | Yes | No | No |
| -1,355380183 | 1,098849126 | Yes | No | No |
| 1,351089579 | 1,068313266 | Yes | No | No |
| 1,375571544 | 1,284185948 | Yes | Yes | Yes |
| 1,310208595 | 1,425048142 | Yes | Yes | Yes |
| 1,269015653 | 1,382965537 | Yes | Yes | Yes |
| -1,21888268 | -1,134966411 | Yes | No | No |
| -1,304784556 | -1,067982228 | Yes | No | No |
| -1,287689067 | -1,008810004 | Yes | No | No |
| -1,356057596 | -1,077997601 | Yes | No | No |
| 1,204808693 | 1,099271066 | Yes | No | No |
| 1,519132651 | 1,196992375 | Yes | No | No |
| -1,318772674 | -1,174227499 | Yes | No | No |
| 1,200977591 | 1,390716661 | Yes | Yes | Yes |
| -1,290181454 | -1,160616578 | Yes | No | No |
| -1,225535831 | -1,064821187 | Yes | No | No |
| -1,420386542 | -1,173126524 | Yes | No | No |

| | | | | |
|---|---|---|---|---|
| -1,209383301 | -1,190591226 | Yes | No | No |
| -1,696733716 | -1,050985508 | Yes | No | No |
| -1,301243229 | -1,533603425 | Yes | Yes | Yes |
| -1,413415898 | -1,189256471 | Yes | No | No |
| -1,355618869 | -1,247229148 | Yes | Yes | Yes |
| 1,633319869 | 1,041488661 | Yes | No | No |
| -1,206819663 | -1,100291631 | Yes | No | No |
| -1,22178294 | -1,099169917 | Yes | No | No |
| -1,338324664 | -1,16578137 | Yes | No | No |
| 1,355989179 | 1,241365196 | Yes | Yes | Yes |
| -1,615394587 | -1,346846361 | Yes | Yes | Yes |
| 1,431548603 | 1,15148926 | Yes | No | No |
| 1,28867776 | 1,30335633 | Yes | Yes | Yes |
| 1,254840207 | 1,242744449 | Yes | Yes | Yes |
| -1,244359051 | -1,26336755 | Yes | Yes | Yes |
| 1,285178353 | 1,192321343 | Yes | No | No |
| -1,315887256 | -1,209800452 | Yes | Yes | Yes |
| 1,319753568 | 1,448136406 | Yes | Yes | Yes |
| -1,302526257 | -1,217386775 | Yes | Yes | Yes |
| -1,256826347 | -1,058655139 | Yes | No | No |
| -1,221317279 | -1,334814533 | Yes | Yes | Yes |
| 1,404520621 | 1,246885623 | Yes | Yes | Yes |
| -1,235785051 | -1,122536265 | Yes | No | No |
| -1,296082522 | -1,123112699 | Yes | No | No |
| 1,36544085 | 1,311040854 | Yes | Yes | Yes |
| -1,22674685 | -1,046616457 | Yes | No | No |
| -1,357904372 | -1,122152865 | Yes | No | No |
| -1,334410998 | -1,173579032 | Yes | No | No |
| -1,225361967 | -1,049347153 | Yes | No | No |
| -1,388446484 | -1,469981673 | Yes | Yes | Yes |
| -1,220890587 | -1,091913237 | Yes | No | No |
| 1,303277976 | 1,447042083 | Yes | Yes | Yes |
| -1,346485916 | -1,130197092 | Yes | No | No |
| 1,204580728 | 1,198926208 | Yes | No | No |
| -1,313016248 | -1,181989096 | Yes | No | No |
| -1,549774066 | -1,206065038 | Yes | Yes | Yes |
| -1,263780888 | -1,124892092 | Yes | No | No |
| 1,223478107 | 1,386551421 | Yes | Yes | Yes |
| 1,235735102 | 1,131788402 | Yes | No | No |
| 1,256501934 | 1,421370638 | Yes | Yes | Yes |
| 1,414259392 | 1,29797333 | Yes | No | No |
| 1,354836932 | 1,152947343 | Yes | No | No |
| -1,372688815 | -1,254114828 | Yes | Yes | Yes |
| 1,26861428 | 1,213977727 | Yes | Yes | Yes |
| 1,208047041 | 1,153358044 | Yes | No | No |
| -1,373463896 | 1,046106181 | Yes | No | No |
| 1,206851204 | 1,511345049 | Yes | Yes | Yes |
| -1,809829516 | -1,018328142 | Yes | No | No |
| -1,270632549 | 1,062103401 | Yes | No | No |
| -1,228885236 | -1,22743511 | Yes | Yes | Yes |
| -1,311498087 | -1,244839178 | Yes | Yes | Yes |
| -1,282587699 | -1,074580263 | Yes | No | No |
| -1,375224985 | 1,022702985 | Yes | No | No |

| | | | | |
|---|---|---|---|---|
| 1,658379634 | 1,279111285 | Yes | Yes | Yes |
| 1,355841842 | 1,11332598 | Yes | No | No |
| -1,301032533 | -1,061266645 | Yes | No | No |
| 1,591497301 | -1,004351736 | Yes | No | No |
| -1,311739103 | -1,202394193 | Yes | Yes | Yes |
| 1,253031898 | 1,364677764 | Yes | Yes | Yes |
| -1,292374208 | 1,028349952 | Yes | No | No |
| -1,282957174 | 1,016742047 | Yes | No | No |
| -1,200608714 | -1,078607439 | Yes | No | No |
| 1,275763341 | 1,212196214 | Yes | Yes | Yes |
| -1,429105575 | -1,205248009 | Yes | Yes | Yes |
| -1,388451231 | -1,213965296 | Yes | Yes | Yes |
| 1,262225939 | 1,314892271 | Yes | Yes | Yes |
| -1,228096894 | -1,125229874 | Yes | No | No |
| 1,494907851 | 1,591273561 | Yes | Yes | Yes |
| 1,215874803 | -1,035379814 | Yes | No | No |
| 1,356591122 | 1,359765138 | Yes | Yes | Yes |
| 1,267043281 | 1,010632218 | Yes | No | No |
| 2,107388578 | 1,165038026 | Yes | No | No |
| 1,308738212 | 1,172235805 | Yes | No | No |
| -1,342823233 | -1,100808602 | Yes | No | No |
| 1,212934599 | 1,12905272 | Yes | No | No |
| 1,345322595 | 1,296710089 | Yes | Yes | Yes |
| 1,30557075 | 1,298280969 | Yes | No | No |
| -1,245113387 | -1,152252847 | Yes | No | No |
| -1,278593945 | -1,053420107 | Yes | No | No |
| 1,367758316 | 1,39906489 | Yes | Yes | Yes |
| -1,568706482 | -1,004661898 | Yes | No | No |
| 1,453563776 | 1,319999513 | Yes | Yes | Yes |
| 1,304905551 | 1,48560943 | Yes | Yes | Yes |
| 1,329834817 | 1,326806495 | Yes | Yes | Yes |
| 1,317113964 | 1,143091377 | Yes | No | No |
| 1,355841686 | 1,065976928 | Yes | No | No |
| -1,472561033 | -1,228972906 | Yes | Yes | Yes |
| 1,239017906 | 1,32670865 | Yes | Yes | Yes |
| 1,295970174 | 1,157516013 | Yes | No | No |
| 1,201355574 | 1,118803395 | Yes | No | No |
| 1,806464262 | 1,319255227 | Yes | Yes | Yes |
| 1,207097611 | 1,185124008 | Yes | No | No |
| 2,035654812 | 1,440562474 | Yes | Yes | Yes |
| 1,389989422 | 1,616914326 | Yes | Yes | Yes |
| 1,276558996 | -1,003933157 | Yes | No | No |
| 1,292815801 | -1,032339034 | Yes | No | No |
| 1,409934235 | 1,094060816 | Yes | No | No |
| 1,512122962 | 1,203912251 | Yes | Yes | Yes |
| 1,289495104 | 1,308624271 | Yes | Yes | Yes |
| 1,288171601 | -1,057408761 | Yes | No | No |
| 1,277008586 | 1,248277347 | Yes | Yes | Yes |
| 1,225960189 | 1,167985776 | Yes | No | No |
| 1,463050861 | 1,362919117 | Yes | Yes | Yes |
| 1,487824488 | 1,200856314 | Yes | Yes | Yes |
| 1,376089073 | 1,293818681 | Yes | Yes | Yes |
| 1,202649568 | -1,085141189 | Yes | No | No |

| | | | | |
|---|---|---|---|---|
| 1,209967099 | 1,301373806 | Yes | Yes | Yes |
| 1,269474121 | 1,406429186 | Yes | Yes | Yes |
| 1,319099929 | 1,40296858 | Yes | Yes | Yes |
| 1,225058956 | 1,297738799 | Yes | Yes | Yes |
| 1,329030035 | 1,305878229 | Yes | Yes | Yes |
| 1,21520144 | 1,388831309 | Yes | Yes | Yes |
| 1,365405777 | 1,219072051 | Yes | Yes | Yes |
| 1,37335318 | 1,175943262 | Yes | No | No |
| 1,321276801 | 1,151309972 | Yes | No | No |
| 1,201726641 | 1,069008913 | Yes | No | No |
| 1,246235561 | -1,040721824 | Yes | No | No |
| 1,36022323 | 1,271701542 | Yes | Yes | Yes |
| 1,270994023 | 1,159943986 | Yes | No | No |
| 1,204154457 | 1,234628935 | Yes | Yes | Yes |
| 1,241286909 | 1,118417777 | Yes | No | No |
| 1,305116508 | 1,191919551 | Yes | No | No |
| 1,323828278 | 1,143958325 | Yes | No | No |
| 1,296615368 | 1,328296265 | Yes | Yes | Yes |
| 1,239431281 | 1,463299214 | Yes | Yes | Yes |
| 1,336731331 | 1,219593617 | Yes | Yes | Yes |
| 1,30907432 | 1,394592712 | Yes | Yes | Yes |
| 1,354078017 | 1,180692945 | Yes | No | No |
| 1,266400451 | 1,124130126 | Yes | No | No |
| 1,201589102 | 1,165314184 | Yes | No | No |
| 1,326443071 | 1,387477342 | Yes | Yes | Yes |
| 1,360926708 | 1,194562117 | Yes | No | No |
| 1,414657782 | 1,606478754 | Yes | Yes | Yes |
| 1,464857206 | 1,150417304 | Yes | No | No |
| 1,355660934 | 1,364098277 | Yes | Yes | Yes |
| 1,512987011 | 1,349888369 | Yes | Yes | Yes |
| -1,233047005 | -1,338405577 | Yes | Yes | Yes |
| 1,327935397 | -1,011942245 | Yes | No | No |
| 1,582990861 | 1,273254424 | Yes | Yes | Yes |
| 1,315219461 | -1,056193202 | Yes | No | No |
| 1,283159561 | 1,310326974 | Yes | Yes | Yes |
| 1,207413008 | 1,099590085 | Yes | No | No |
| -1,218727501 | -1,118734177 | Yes | No | No |
| -1,357741092 | -1,196324999 | Yes | No | No |
| 1,410468661 | 1,312464835 | Yes | Yes | Yes |
| 1,223459888 | 1,179471166 | Yes | No | No |
| 1,2677284 | 1,11930952 | Yes | No | No |
| 1,404314812 | 1,587158713 | Yes | Yes | Yes |
| -1,312166023 | -1,293715834 | Yes | Yes | Yes |
| 1,347132415 | 1,0353623 | Yes | No | No |
| 1,435400529 | 1,323270146 | Yes | Yes | Yes |
| 1,206924056 | 1,162489514 | Yes | No | No |
| -1,215879589 | -1,022409871 | Yes | No | No |
| -1,517090881 | -1,139013954 | Yes | No | No |
| 1,260283225 | 1,104623082 | Yes | No | No |
| -1,340853615 | -1,184608956 | Yes | No | No |
| 1,281012113 | 1,065627283 | Yes | No | No |
| -1,211287596 | -1,185332373 | Yes | No | No |
| 1,232194775 | 1,155261394 | Yes | No | No |

| | | | | |
|---|---|---|---|---|
| 1,676795638 | 1,096176513 | Yes | No | No |
| -1,258742918 | -1,24310229 | Yes | Yes | Yes |
| -1,206853079 | -1,207837393 | Yes | Yes | Yes |
| -1,220367105 | -1,169986754 | Yes | No | No |
| 1,252567724 | 1,336644654 | Yes | Yes | Yes |
| 1,258942817 | 1,337057242 | Yes | Yes | Yes |
| 1,252740139 | 1,354343775 | Yes | Yes | Yes |
| -1,22886806 | -1,48420959 | Yes | Yes | Yes |
| 1,309047311 | 1,141440454 | Yes | No | No |
| 1,328482921 | 1,27763034 | Yes | Yes | Yes |
| 1,466536371 | 1,360544833 | Yes | Yes | Yes |
| 1,222521568 | 1,209567283 | Yes | Yes | Yes |
| -1,299822431 | -1,173795246 | Yes | No | No |
| -1,254345241 | -1,137363722 | Yes | No | No |
| -1,275652099 | -1,178264122 | Yes | No | No |
| -1,370488338 | -1,178827955 | Yes | No | No |
| 1,292512036 | 1,49123666 | Yes | Yes | Yes |
| -1,087294332 | -1,2407106 | No | Yes | No |
| 1,12590933 | 1,236465044 | No | Yes | No |
| -1,117587289 | -1,235942881 | No | Yes | No |
| -1,157726104 | -1,283941097 | No | Yes | No |
| -1,129502587 | -1,203148882 | No | Yes | No |
| -1,207621224 | -1,37124446 | No | Yes | No |
| 1,126229606 | 1,22856631 | No | Yes | No |
| 1,032072857 | -1,395285289 | No | Yes | No |
| -1,101194944 | -1,308496515 | No | Yes | No |
| -1,150129305 | -1,201350303 | No | Yes | No |
| 1,141429177 | 1,214944857 | No | Yes | No |
| 1,170352232 | 1,203094845 | No | Yes | No |
| 1,197270765 | 1,345168723 | No | Yes | No |
| 1,145800089 | 1,346466124 | No | Yes | No |
| -1,137945257 | -1,207510439 | No | Yes | No |
| 1,189872793 | 1,445978481 | No | Yes | No |
| -1,075932954 | -1,209907716 | No | Yes | No |
| 1,191321826 | 1,223408251 | No | Yes | No |
| -1,198680896 | -1,242970365 | No | Yes | No |
| -1,130324846 | -1,257070168 | No | Yes | No |
| -1,096865196 | -1,207822243 | No | Yes | No |
| 1,147018673 | 1,40650377 | No | Yes | No |
| 1,170486751 | 1,317035713 | No | Yes | No |
| -1,13955407 | -1,231126329 | No | Yes | No |
| 1,101734573 | 1,365670799 | No | Yes | No |
| -1,116356851 | -1,257511232 | No | Yes | No |
| 1,138375683 | 1,249882349 | No | Yes | No |
| -1,146800471 | -1,250014763 | No | Yes | No |
| -1,103747431 | -1,213155706 | No | Yes | No |
| -1,08019316 | -1,231045629 | No | Yes | No |
| -1,14261316 | -1,244608061 | No | Yes | No |
| 1,16972985 | 1,256963329 | No | Yes | No |
| -1,145382199 | -1,218750161 | No | Yes | No |
| -1,134441523 | -1,258648173 | No | Yes | No |
| -1,191377245 | -1,438004872 | No | Yes | No |
| -1,173864572 | -1,27517607 | No | Yes | No |

| | | | | |
|---|---|---|---|---|
| 1,135785293 | 1,275612865 | No | Yes | No |
| 1,145028815 | 1,353765342 | No | Yes | No |
| -1,14975135 | -1,209266186 | No | Yes | No |
| 1,18704082 | 1,207766847 | No | Yes | No |
| -1,090378783 | -1,204207793 | No | Yes | No |
| 1,187508266 | 1,26446302 | No | Yes | No |
| -1,194125245 | -1,297790466 | No | Yes | No |
| -1,193691651 | -1,24968298 | No | Yes | No |
| 1,176936355 | 1,334674171 | No | Yes | No |
| -1,176782788 | -1,289000808 | No | Yes | No |
| 1,188621373 | 1,327432162 | No | Yes | No |
| 1,182525017 | 1,325464081 | No | Yes | No |
| -1,041780138 | -1,476372616 | No | Yes | No |
| 1,149763092 | 1,228202468 | No | Yes | No |
| -1,191687666 | -1,20323202 | No | Yes | No |
| 1,183578483 | 1,238504221 | No | Yes | No |
| 1,189783793 | 1,535307514 | No | Yes | No |
| -1,103717467 | -1,208599101 | No | Yes | No |
| 1,14479521 | 1,38224894 | No | Yes | No |
| 1,120103645 | 1,201864054 | No | Yes | No |
| -1,074902671 | -1,246336952 | No | Yes | No |
| -1,153783696 | -1,202112107 | No | Yes | No |
| -1,146442686 | -1,230731775 | No | Yes | No |
| -1,099671112 | -1,314302401 | No | Yes | No |
| 1,185175916 | 1,341771176 | No | Yes | No |
| 1,168901561 | 1,230180981 | No | Yes | No |
| 1,191741122 | 1,323225124 | No | Yes | No |
| 1,145366306 | 1,289155029 | No | Yes | No |
| -1,130710842 | -1,38508602 | No | Yes | No |
| 1,184774313 | 1,221587824 | No | Yes | No |
| -1,145242951 | -1,258574055 | No | Yes | No |
| 1,124627508 | 1,29246613 | No | Yes | No |
| 1,142569926 | 1,243113847 | No | Yes | No |
| 1,139545004 | 1,202059828 | No | Yes | No |
| 1,113575032 | 1,236194581 | No | Yes | No |
| 1,180015672 | 1,211052435 | No | Yes | No |
| 1,198993178 | 1,282189084 | No | Yes | No |
| -1,126150452 | -1,232479726 | No | Yes | No |
| 1,097934125 | 1,297035332 | No | Yes | No |
| -1,125200795 | -1,280774334 | No | Yes | No |
| -1,056677385 | -1,25765592 | No | Yes | No |
| -1,178336294 | -1,22092013 | No | Yes | No |
| -1,090132209 | -1,235843339 | No | Yes | No |
| 1,196897636 | 1,241773413 | No | Yes | No |
| -1,182649093 | -1,209108442 | No | Yes | No |
| -1,188211627 | -1,39567005 | No | Yes | No |
| -1,122978955 | -1,371416068 | No | Yes | No |
| 1,177703048 | 1,201133582 | No | Yes | No |
| -1,186315594 | -1,244914158 | No | Yes | No |
| 1,179008073 | 1,274926424 | No | Yes | No |
| 1,130613443 | 1,442739585 | No | Yes | No |
| 1,179407008 | 1,396038573 | No | Yes | No |
| 1,17298459 | 1,206914956 | No | Yes | No |

| | | | | |
|---|---|---|---|---|
| -1,154684986 | -1,254502768 | No | Yes | No |
| -1,196207961 | -1,203175572 | No | Yes | No |
| -1,171636563 | -1,207441044 | No | Yes | No |
| -1,09729943 | -1,245207916 | No | Yes | No |
| -1,168093817 | -1,412329255 | No | Yes | No |
| 1,195884399 | 1,201041256 | No | Yes | No |
| -1,165154812 | -1,26433199 | No | Yes | No |
| -1,177877183 | -1,244908749 | No | Yes | No |
| 1,169215644 | 1,362855724 | No | Yes | No |
| -1,165329087 | -1,204584225 | No | Yes | No |
| -1,155878016 | -1,231766919 | No | Yes | No |
| -1,167589046 | -1,338174083 | No | Yes | No |
| 1,261798428 | 1,670858959 | No | Yes | No |
| 1,18935791 | 1,207257561 | No | Yes | No |
| -1,141575821 | -1,316098458 | No | Yes | No |
| -1,115201419 | -1,312795698 | No | Yes | No |
| -1,189653075 | -1,342247458 | No | Yes | No |
| -1,145654763 | -1,240057399 | No | Yes | No |
| -1,171942965 | -1,337483129 | No | Yes | No |
| 1,169440695 | 1,335076547 | No | Yes | No |
| -1,042168434 | -1,272341749 | No | Yes | No |
| -1,162332671 | -1,210257086 | No | Yes | No |
| -1,078932575 | -1,204620514 | No | Yes | No |
| -1,120735523 | -1,302228597 | No | Yes | No |
| -1,090870288 | -1,282324487 | No | Yes | No |
| 1,167397733 | 1,378412868 | No | Yes | No |
| 1,160494949 | 1,333067219 | No | Yes | No |
| 1,185730122 | 1,408346052 | No | Yes | No |
| 1,17702995 | 1,24299004 | No | Yes | No |
| 1,147463581 | 1,254174714 | No | Yes | No |
| 1,167633917 | 1,370203312 | No | Yes | No |
| 1,147734211 | 1,340073783 | No | Yes | No |
| 1,196847911 | 1,263116658 | No | Yes | No |
| -1,097803763 | -1,207041178 | No | Yes | No |
| 1,190704893 | 1,395672955 | No | Yes | No |
| -1,132804399 | -1,224823731 | No | Yes | No |
| -1,068451635 | -1,269957391 | No | Yes | No |
| 1,184289648 | 1,320961634 | No | Yes | No |
| -1,179610195 | -1,27494808 | No | Yes | No |
| -1,141219187 | -1,203595626 | No | Yes | No |
| 1,169255961 | 1,553174492 | No | Yes | No |
| -1,023238653 | -1,241114174 | No | Yes | No |
| -1,128620538 | -1,25511791 | No | Yes | No |
| 1,146857128 | 1,497088423 | No | Yes | No |
| -1,150721849 | -1,32780002 | No | Yes | No |
| -1,190806831 | -1,370421075 | No | Yes | No |
| 1,182401413 | 1,383354454 | No | Yes | No |
| -1,187874171 | -1,251363402 | No | Yes | No |
| 1,131144659 | 1,47734538 | No | Yes | No |
| -1,089678483 | -1,202439101 | No | Yes | No |
| 1,178614761 | 1,298159934 | No | Yes | No |
| -1,164837096 | -1,332051413 | No | Yes | No |
| -1,167982184 | -1,246233588 | No | Yes | No |

| | | | | |
|---|---|---|---|---|
| 1,172710103 | 1,286914628 | No | Yes | No |
| -1,168961638 | -1,238904665 | No | Yes | No |
| 1,248652272 | 1,707178422 | No | Yes | No |
| 1,056684797 | -1,580173964 | No | Yes | No |
| 1,165213698 | 1,249616108 | No | Yes | No |
| -1,183916589 | -1,209866895 | No | Yes | No |
| -1,184060426 | -1,28370571 | No | Yes | No |
| 1,184904951 | 1,239307308 | No | Yes | No |
| -1,088996604 | -1,315187845 | No | Yes | No |
| 1,180460722 | 1,284444028 | No | Yes | No |
| 1,128105585 | 1,258728427 | No | Yes | No |
| 1,088104106 | 1,255449056 | No | Yes | No |
| 1,171611233 | 1,289240959 | No | Yes | No |
| -1,178122786 | -1,203724904 | No | Yes | No |
| -1,089981182 | -1,260603384 | No | Yes | No |
| 1,012474253 | -1,311442168 | No | Yes | No |
| 1,170185905 | 1,242215666 | No | Yes | No |
| 1,179832425 | 1,277168576 | No | Yes | No |
