## Supplemental Table 2 and 3 for "Decreased expression of mitochondrial aminoacyl-tRNA synthetases causes downregulation of mitochondrial OXPHOS subunits in type 2 diabetic skeletal muscle"

**Supplementary Table 2. Total Amino Acid composition of the 13 well-validated proteins that are encoded and translated in human and mouse mitochondria.**

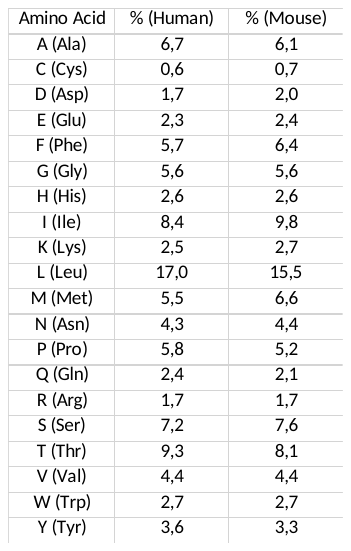

**Supplementary Table 3. Amino acid composition of each of the 13 proteins encoded in mouse mitochondria**

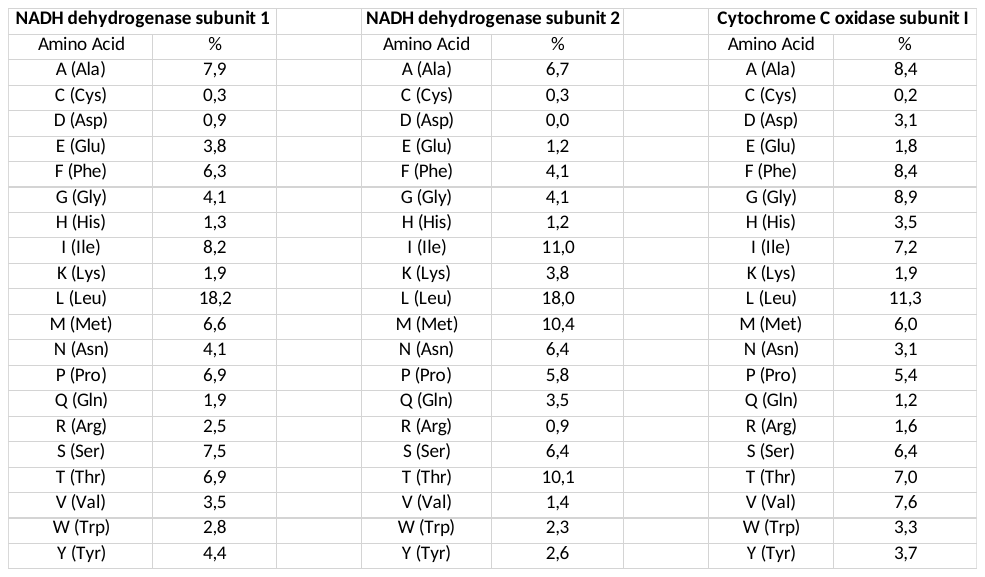

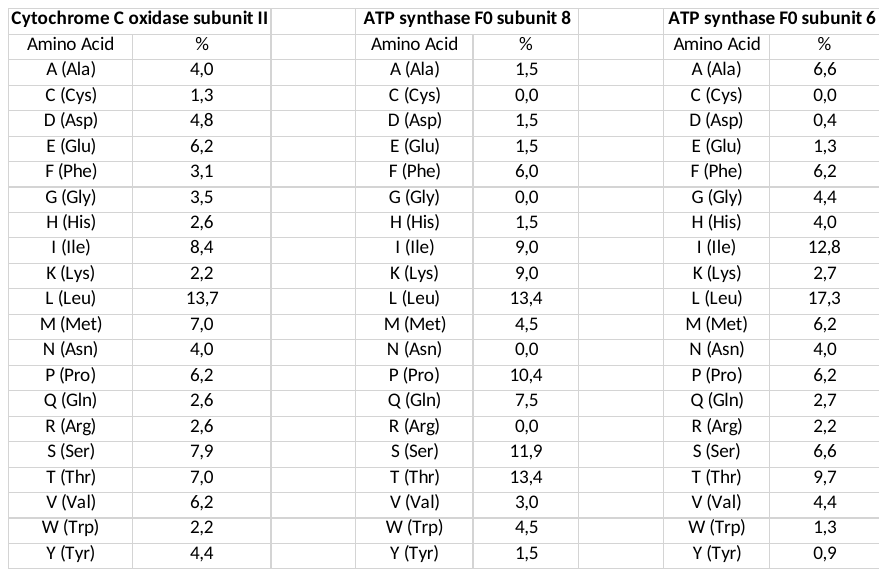

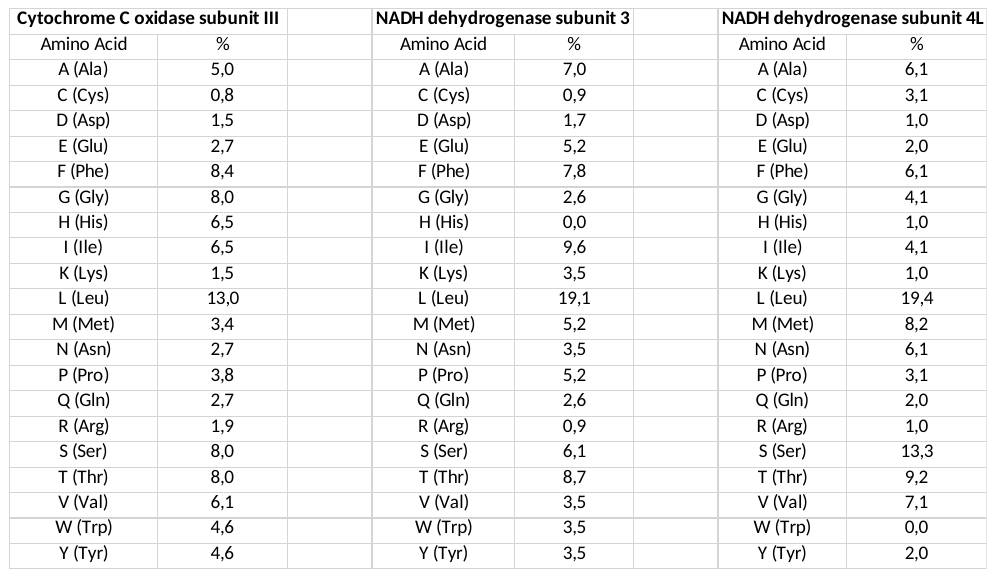

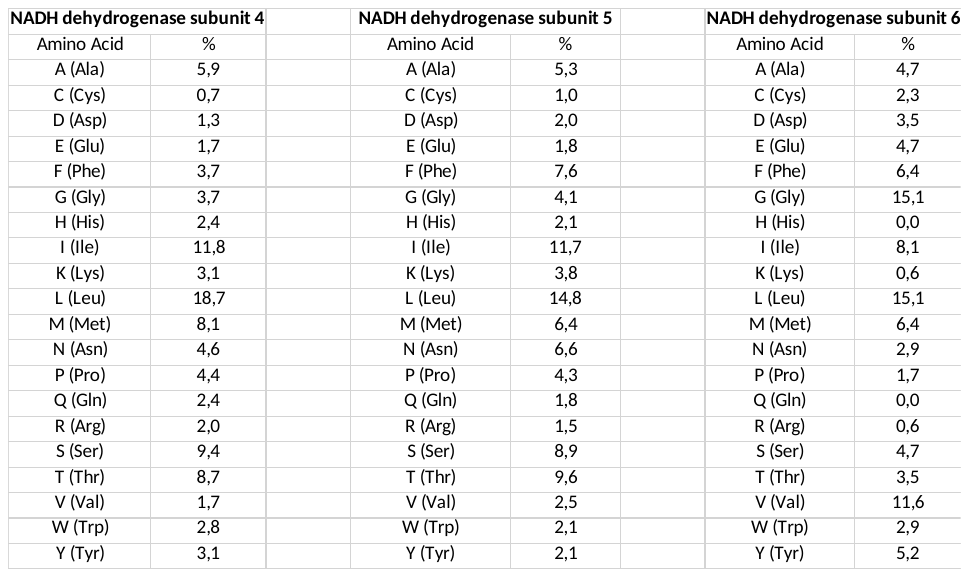

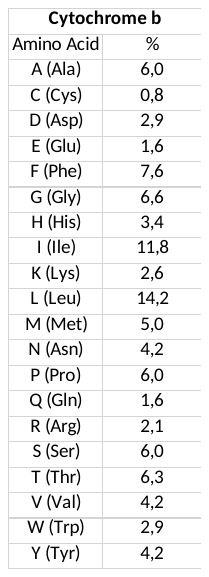
